# In Search of Ethical Procedures for LLM-Assisted Systematic Review Production: A Proof-of-Concept Evaluation of Selected Review Components

**DOI:** 10.64898/2026.02.18.26346559

**Authors:** Liam McLaughlin, Michael S. Walz, Cade Arries

## Abstract

Large language models (LLMs) are increasingly used in scientific writing, but the conditions under which they can be applied responsibly to evidence synthesis remain poorly defined. We conducted a proof-of-concept study examining selected components of the systematic review process rather than the systematic review as a whole, with the aim of identifying where LLM assistance is defensible and where human judgment remains necessary. First, using a recently published umbrella review as a benchmark, we compared LLM screening decisions against those of the original human authors. Applied to abstracts, the model adhered to its stated inclusion criteria in all cases examined; applied to full texts, agreement with the human authors reached 86.2% (Cohen’s κ = 0.38), but only after the published criteria were iteratively reworded. Applied verbatim, the published criteria led the model to reject every article the original authors had included, indicating that criteria wording, rather than model capability alone, governed performance. Second, in an exploratory blinded evaluation, six board-certified hematopathologists rated two LLM-produced reviews and one published human-authored review on the same topic. Ratings favored the LLM-produced reviews (means 3.4–3.66 versus 2.6 of 5), and raters did not identify AI involvement above chance (27.8% accuracy; permutation p = 0.84), with the human-authored review most frequently attributed to AI. Given the sample size, these observations are hypothesis-generating rather than definitive. Internal auditing of the LLM-produced reviews found citation misattribution in 4.13–7.06% of cited claims, which text-restriction strategies reduced but did not eliminate. Taken together, these findings suggest that LLMs can perform bounded, well-specified subtasks of review production under supervision, while verification, disclosure, and final adjudication remain human responsibilities. Informed by these observations, we developed ReviewFoundry, an open-source platform built to support human-supervised, AI-assisted review workflows.

## INTRODUCTION

Since the public popularization of large language model (LLM) tools [1], researchers have sought ways to incorporate them into the paper-writing process [2]. LLMs such as OpenAI’s ChatGPT and Anthropic’s Claude offer two capabilities relevant to manuscript production: **1.** the ability to summarize large volumes of text [3], which offers a route to partial automation of literature review, and **2.** the ability to synthesize or revise writing into a format that reads as professional and scientifically sound [4]. Both functions can be performed under user-specified instructions [5], which opens the possibility of drafting steered by a human editor rather than executed by one.

These developments raise two questions for the academic community. The first is how LLM use can be integrated into the research process in an ethical manner [6]. This question is pressing given that vocabulary-based analysis indicates a measurable increase in AI-associated language in published material [7], while researchers remain divided over how, and whether, LLM use should be reported [8].

The second question concerns the quality and accuracy of LLM-assisted publications. A panel of 20 neurosurgeons was unable to detect a significant difference in writing quality between human- and AI-written publications drawing on the same base datasets [4]. The AI-generated articles in that study were nonetheless not free of error, which illustrates a tension inherent to publishing AI-assisted work: text that has not been verified claim by claim may still read as authoritative, and the verification burden erodes the efficiency that motivated the automation in the first place.

Systematic reviews are one area of medical publication where AI assistance has been proposed. A systematic review evaluates the body of literature on a topic in a procedural manner [9] and reports findings in a form that serves as a reference for other professionals [10]. Because systematic reviews inform both future research and clinical practice, the information they contain must be presented accurately, as reported in the source papers, and transparently, with a correct citation for every claim. Several recent articles describe how AI can assist systematic review synthesis [11] [12], although these focus largely on human-directed use of the web-based chat interfaces offered by LLM vendors. By working instead through an Application Programming Interface (API), a software interface allowing two programs to communicate in a predefined manner [13], several components of the review process can be queried iteratively and at scale.

In this study we used such an API-driven pipeline as an investigational tool. Querying an LLM programmatically allows the same operation to be applied to hundreds of articles within a single script execution [13], which makes it possible to characterize model behavior across a corpus rather than anecdotally. We used this capacity to examine two components of the review process in isolation: article screening against explicit criteria, and the drafting of review prose from a fixed set of sources. We also examined citation fidelity, since prior work has reported citation error rates as high as 70% in AI-assisted review writing [12].

We emphasize at the outset that this work is a proof of concept. We do not present evidence that systematic reviews can or should be produced without human involvement. Our aims were: **1.** to assess whether an LLM applies explicit screening criteria in a manner comparable to human reviewers, using a published review as a benchmark; **2.** to explore, in a small expert panel, how LLM-produced review text is perceived relative to published human-authored text, and whether domain experts identify AI involvement; and **3.** to use these observations to define what a responsible, human-supervised LLM-assisted review workflow would need to provide. The third aim led to the development of ReviewFoundry, an open-source application described near the end of this paper.

## RESULTS

### Part 1 — Proof-of-Concept Comparison of LLM Screening Against a Published Benchmark

To examine whether an LLM can apply explicit inclusion and exclusion criteria, we compared the screening decisions reported in a recently published umbrella review by Yabinze et al. [14] with LLM screening of the same article set. This design uses a single published review as a benchmark and is intended to characterize model behavior rather than to estimate screening performance across reviews in general.

For the full-text comparison, we used 87 articles that reached the full-text stage in the original umbrella review, of which 9 were accepted and 78 rejected by the original authors. When the Claude Sonnet 4.6 API was provided with the same articles and the inclusion criteria as written verbatim by Yabinze et al. [14], all 9 originally included articles were rejected. On review, this was not attributable to the model disregarding instructions. Rather, the published criteria, read literally, were more stringent than the decision process the human authors appeared to have applied. For example, the original authors included studies in which the outcome of interest was assessed secondarily, or in which the population of interest was pooled with other populations; read literally, both scenarios fell outside the stated criteria and prompted rejection. Accommodating these cases required stating explicitly that secondary assessment of the outcome was permissible.

We therefore revised the criteria iteratively, loosening them until the model consistently accepted all articles included in the original study. We refer to this set as the ‘Most Lenient Criteria’. Rescreening the 87 articles under these criteria yielded 27 acceptances (31%), comprising the original 9 and 18 articles that the umbrella review had rejected, for 79.3% agreement with the human authors. Classifying the 18 discordant decisions, 4 (4.6% of 87) were attributable to ‘Improper AI Criteria’, in which the model followed its criteria as a human reasonably might but produced a false positive because some aspect of the criteria remained undertuned; 8 (9.2%) to an ‘AI False Positive’, in which the model accepted an article despite criteria that would reasonably have led a human to reject it; and 6 (6.9%) to a ‘Human False Negative / Improper Criteria’, in which the original authors either appeared to have rejected an article inconsistently with their accepted set, or had not specified their criteria sufficiently for a reader to reconstruct the distinction. Cohen’s kappa [15] was 0.41, indicating moderate agreement, and McNemar’s test [16] yielded p = 7.63e-6, indicating that the model under these criteria was significantly more inclusive than the human reviewers.

To reduce the false positive rate, we further specified the criteria so that incidental mention of the target population was insufficient for inclusion and some meaningful analysis of that population was required, yielding what we term the ‘Lenient Criteria’. Rescreening under these criteria yielded 13 acceptances, comprising 5 of the original 9 and 8 articles rejected by the umbrella review, for 86.2% agreement. Of the 12 discordant decisions, 2 (2.3% of 87) were attributable to ‘Improper AI Criteria’, 3 (3.4%) to an ‘AI False Positive’, 4 (4.6%) to a ‘Human False Negative / Improper Criteria’, and 3 (3.4%) to a ‘Human False Positive / Improper Criteria’. The four originally included articles that the model now rejected were, on our reading, rejected consistently with the ‘Lenient Criteria’, as each addressed a pooled or non-target population. This does not indicate that their inclusion in the umbrella review was unwarranted. It indicates instead that the original authors appeared to reject comparable articles on the same population grounds, so the model’s rejections could not be classified as aberrant. Across both full-text evaluations, AI false positives arose from two recurring behaviors: **1.** failure to recognize that subtypes of systematic review, such as a systematic scoping review, did not meet a stated study design requirement of ‘systematic review or meta-analysis’, a distinction a human reviewer would likely not have needed stated; and **2.** occasional conflation of the intervention and the outcome of interest when the outcome appeared in the position of the intervention, which represented a more clearly aberrant behavior. Cohen’s kappa [15] under the ‘Lenient Criteria’ was 0.38, indicating fair agreement, and McNemar’s test [16] yielded p = 0.39, indicating no significant difference in leniency between the model and the human reviewers.

We next used the ‘Most Lenient Criteria’ to evaluate abstract screening. This assessment included the abstracts of the 9 studies included by Yabinze et al. [14] alongside 82 additional articles comprising a mix of topically similar studies and studies constructed to violate one inclusion criterion outright. Seven of the nine originally included articles were accepted. On review of the model’s stated rationales, we found no case in which it failed to follow its abstract screening instructions; the two non-acceptances followed from the criteria as written. This assessment therefore addresses instruction adherence rather than whether the accepted articles belonged in the umbrella review.

Taken together, these results indicate that an LLM can apply explicit screening criteria in a manner that is reproducible and, in most cases, defensible on the criteria’s own terms. They also indicate that agreement with human screeners is contingent on criteria wording rather than achieved by default, and that the residual disagreements include errors on both sides. We interpret these findings as supporting a supervised screening role in which criteria are deliberately biased toward false positives, with human adjudication of the remaining set. Results of this portion of the study are summarized in **Table 1**, **Table 2**, and **Figure 1**. Included and excluded studies at each stage, the model’s stated rationales, and our classification of each decision are provided in **Supplemental Tables 1–7**.

**Table 1.**
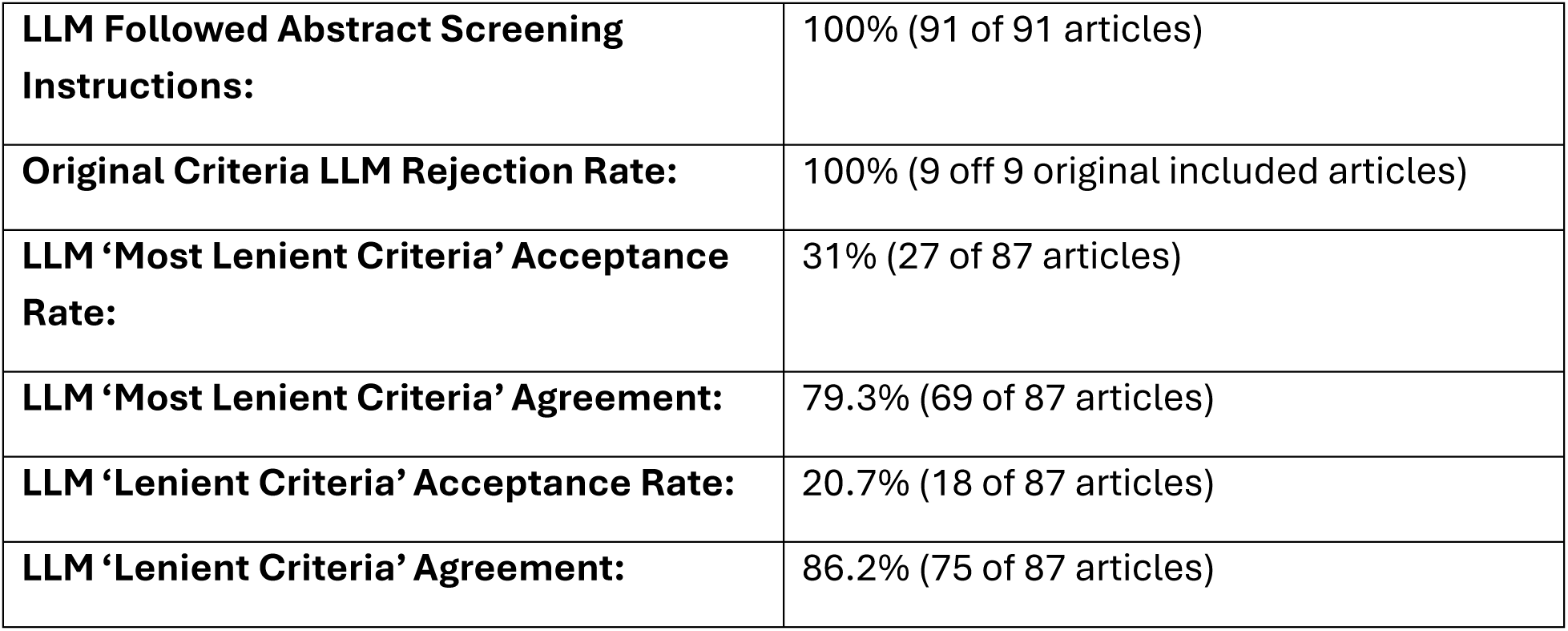
Overall Results from the Screening Comparison Study.

|  |  |
| --- | --- |
| **LLM Followed Abstract Screening Instructions:** | 100% (91 of 91 articles) |
| **Original Criteria LLM Rejection Rate:** | 100% (9 off 9 original included articles) |
| **LLM ‘Most Lenient Criteria’ Acceptance Rate:** | 31% (27 of 87 articles) |
| **LLM ‘Most Lenient Criteria’ Agreement:** | 79.3% (69 of 87 articles) |
| **LLM ‘Lenient Criteria’ Acceptance Rate:** | 20.7% (18 of 87 articles) |
| **LLM ‘Lenient Criteria’ Agreement:** | 86.2% (75 of 87 articles) |

**Table 2.** ‘Most Lenient Criteria’ vs ‘Lenient Criteria’ Analysis.

|  | **Most Lenient Criteria** | **Lenient Criteria** |
| --- | --- | --- |
| **Disagreement due to ‘Improper AI Criteria’** | 4.6% (4/87 articles) | 2.3% (2/87 articles) |
| **Disagreement due to ‘Human False Negative’/ ‘Improper Human Criteria’** | 6.9% (6/87 articles) | 4.6% (4/87 articles) |
| **Disagreement due to ‘Human False Positive’/ ‘Improper Human Criteria’** | 0% | 3.4% (3/87 articles) |
| **Disagreement due to ‘AI False Positive’** | 9.2% (8/87 articles) | 3.4% (3/87 articles) |
| **Cohen’s $\kappa$ (-1 = Complete Disagreement, 1 = Complete Agreement)** | 0.41 | 0.38 |
| **McNemar’s Test p-val** | 7.63e-6 | 0.39 |

**Figure 1.**
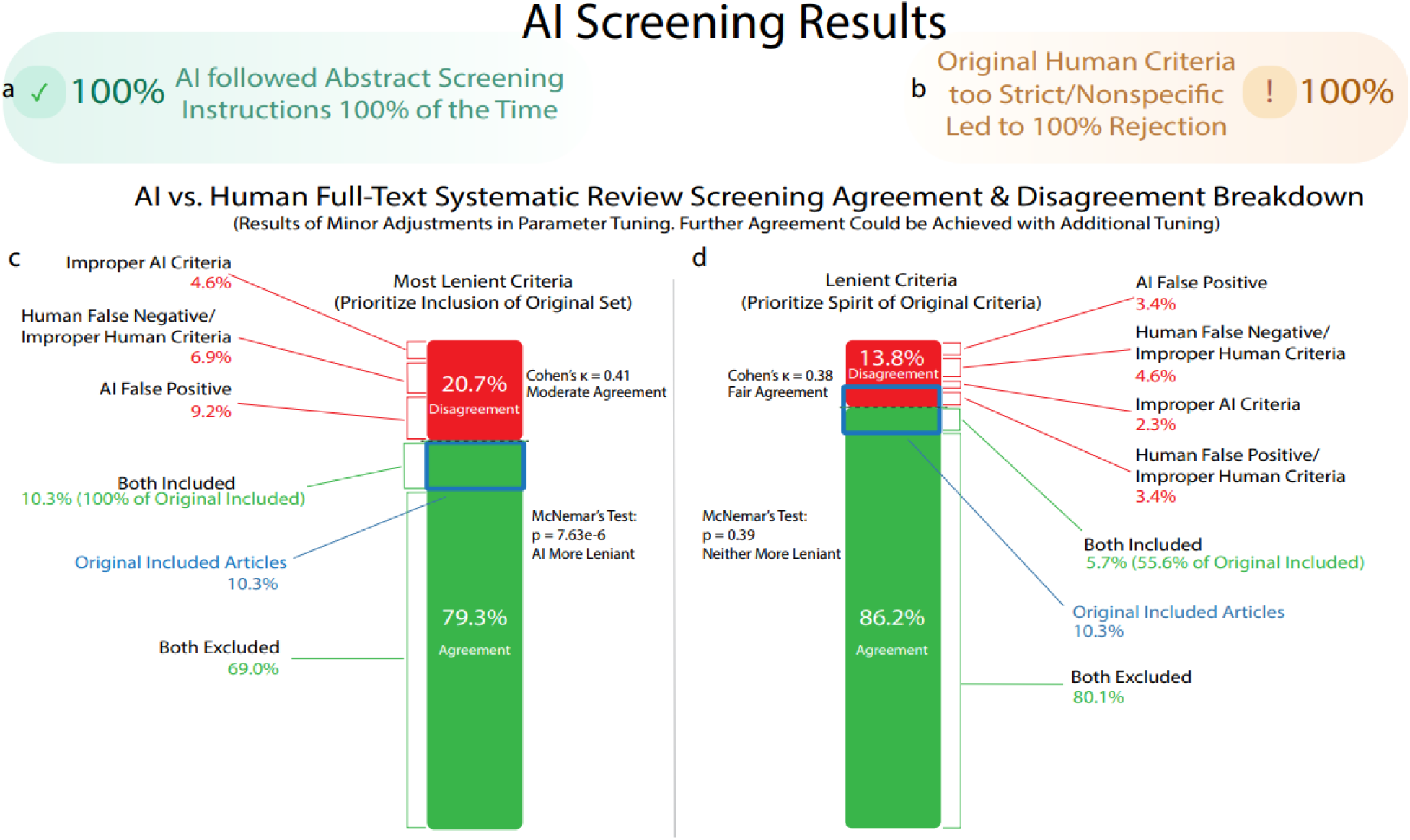
Evaluation of LLM Screening Capabilities Compared to Human Authors. **a.** Evaluation of LLM Abstract Screening capabilities showed no evidence of instruction disobedience. The AI followed the inclusion/exclusion criteria accurately for all abstracts. **b.** Full-text screening with the criteria provided by the authors of Yabinze et al. [14] led to rejection of all the human-included sources. This was determined to be due to the criteria being too strict when taken literally rather than the LLM ignoring instructions. **c.** Criteria tuned to accept all the articles also included as Yabinze et al. [14] (‘Most Lenient Criteria’) led to overall moderate agreement between the LLM and human reviewers (Cohen’s κ = 0.41) and a statistically significant increase in leniency by the LLM compared to the humans (McNemar’s test p = 7.63e-6). When comparing the human and LLM, 79.3% of the articles were agreed upon, while 20.7% additional articles were accepted by the LLM, including: 4.6% due to the criteria still being undertuned, 6.9% due to possible human false negative or human criteria being under-descriptive, and 9.2% due to the LLM not obeying instructions in the same manner a human reasonably may. **d.** Criteria tuned to make the LLM accept/reject articles in line to the author’s original presumed intent, elaborating on specific inclusion/exclusion cases beyond the original criteria but not overly forcing leniency (‘Lenient Criteria’) led to overall fair agreement between the LLM and human reviewers (Cohen’s κ = 0.38) and no significant difference in leniency/stringency between the LLM and the humans (McNemar’s test p = 0.39). When comparing the human and LLM, 86.2% of the articles were agreed upon, with 13.8% disagreement, including: 2.3% due to the criteria still being undertuned, 4.6% due to possible human false negative or human criteria being under-descriptive, 3.4% due to possible human false positive or human criteria being under-descriptive, and 3.4% due to the LLM not obeying instructions in the same manner a human reasonably may.

### Part 2 — Exploratory Expert Evaluation of LLM-Produced Review Text

We produced paired sets of ‘Automated’ reviews, drafted end-to-end through the API, and ‘Semi-Automated’ reviews, drafted through the web chat interface from a human-assembled source set. The first set matched the topic of van den Ende et al.’s “Triple-Negative Breast Cancer and Predictive Markers of Response to Neoadjuvant Chemotherapy: A Systematic Review” [17] using the Claude API ‘Sonnet 3.5’ model and served as a development run. The second set, used in the evaluation reported here, matched the topic of Santisteban-Espejo et al.’s “The Need for Standardization in Next-Generation Sequencing Studies for Classic Hodgkin Lymphoma: A Systematic Review” [18] using the Claude API ‘Sonnet 4.0’ model.

Both review types were drafted in a piecemeal fashion. For the automated review, the model was first given a topic and returned boolean search terms, which were used to query PubMed through the NCBI API. The model then assessed each returned source against predefined criteria. For both review types, the model next summarized the retained source papers and generated a set of results subsections to serve as anchor points, then drafted the introduction, results, discussion, conclusion, and abstract with the summaries and the accumulating manuscript available for reference. The pipeline is depicted in **Figure 2**.

**Figure 2:**
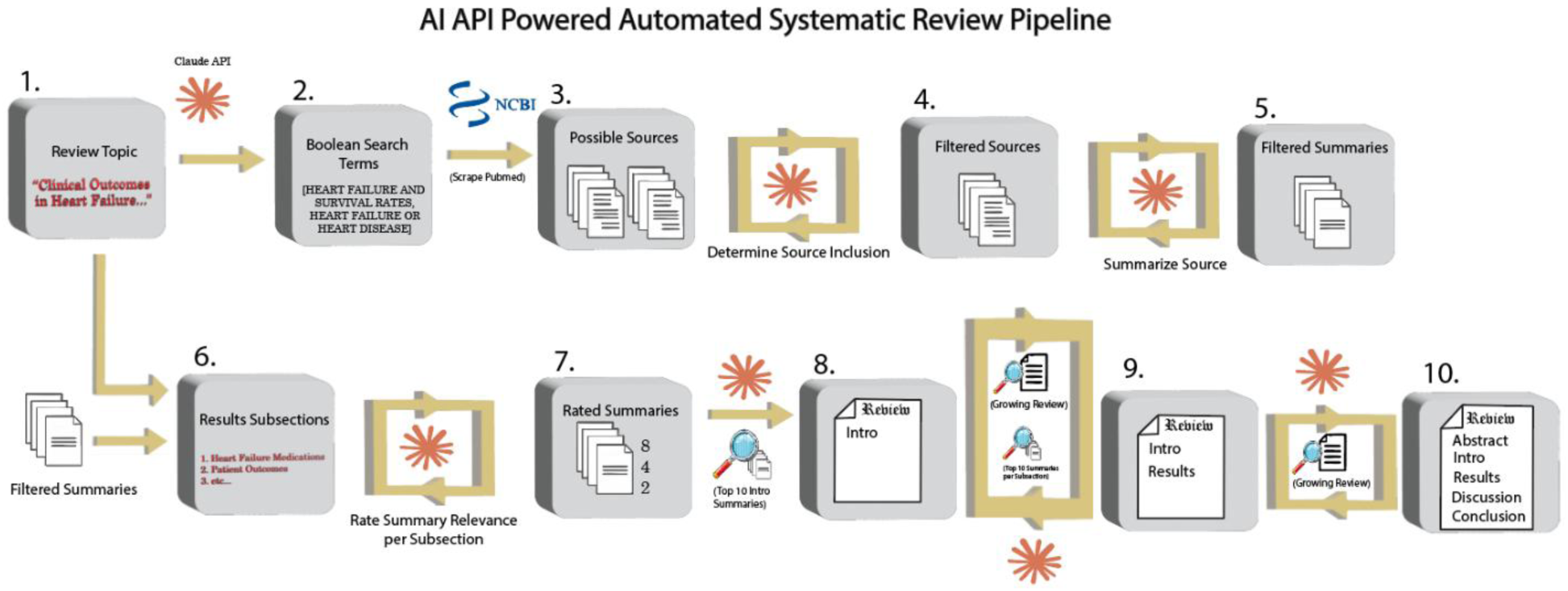
Complete Pipeline for Automated Systematic Review Creation. We utilized the following steps to automate systematic review creation. 1. Any review topic was provided by the user. 2. The Claude API was provided the topic and generated boolean terms to search online databases. 3. The NCBI API was used to search pubmed (or any API associated database) for papers associated with the boolean search terms. 4. The Claude API was used to evaluate all sources for inclusion. 5. The Claude API was used to summarize all papers that were not excluded. 6. The summaries and topics were used to create ‘results subsections’ to act as anchor points for review creation. 7. To mitigate text glut and thus drastically reduce the Claude API’s citation misattribution rate, all source summaries were ranked by how relevant they were to each section of the review. 8. The top 10 rated summaries for the intro were used to write the intro. 9. The top 10 rated summaries for each results subsection were used to write that subsection. 10. The intro and results were used to write the discussion, conclusion, and abstract. 11. After the AI assisted and AI automated systematic reviews were generated, they were submitted for review by physicians with familiarity with the subject matter.

The three deidentified reviews were presented to six board-certified hematopathologists at the University of Minnesota, followed by a short survey grading each review. Survey questions were oriented toward apparent quality on an initial read rather than toward verification of sourcing. Results are shown in **Table 3** and **Figure 3**. Given the panel size and the single topic evaluated, these results should be read as exploratory.

**Table 3.**
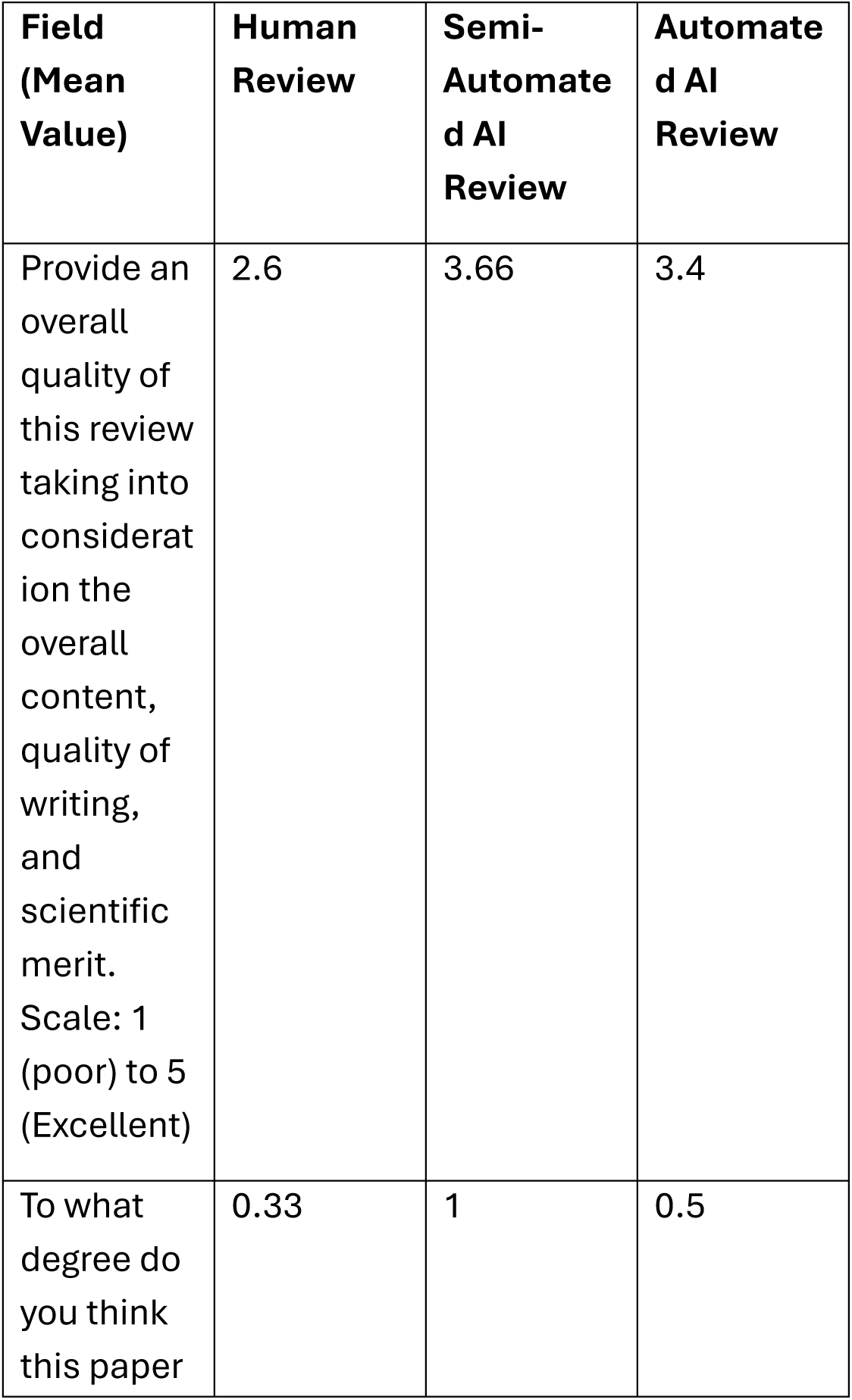
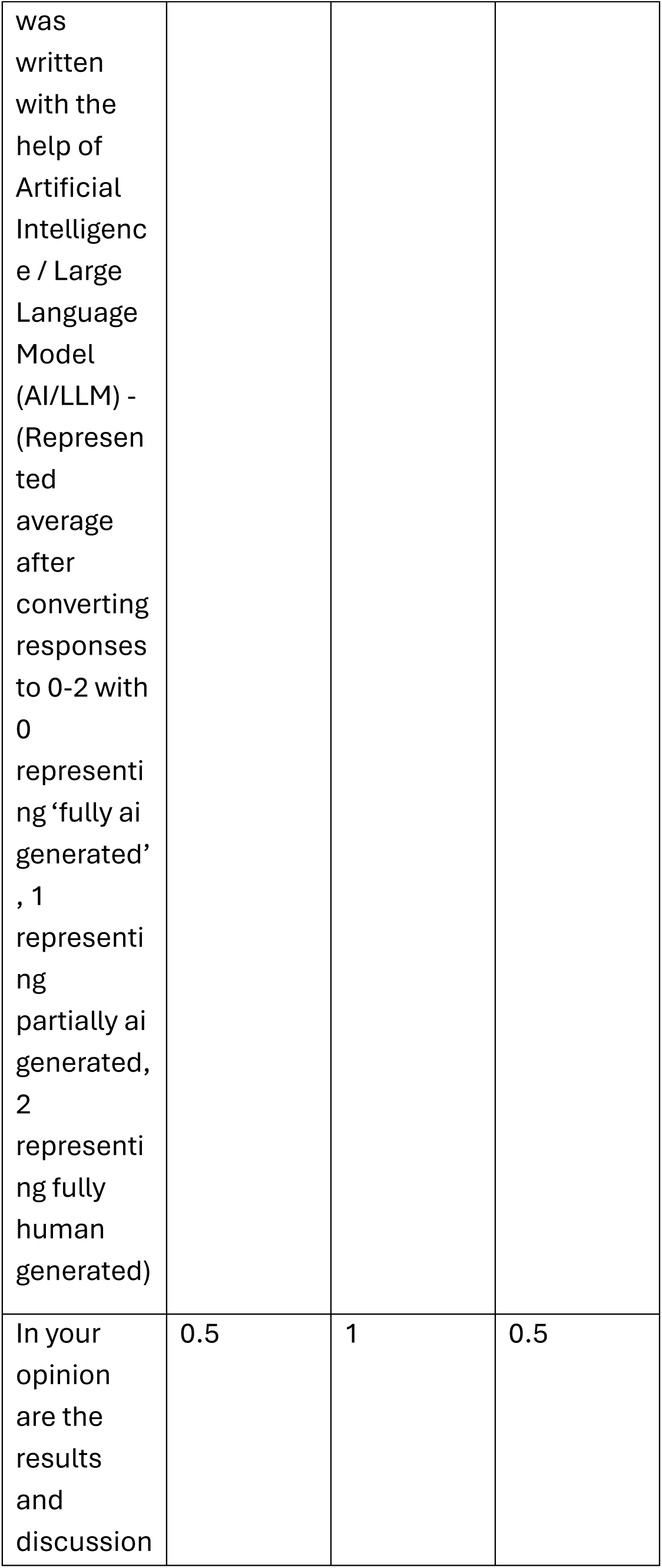
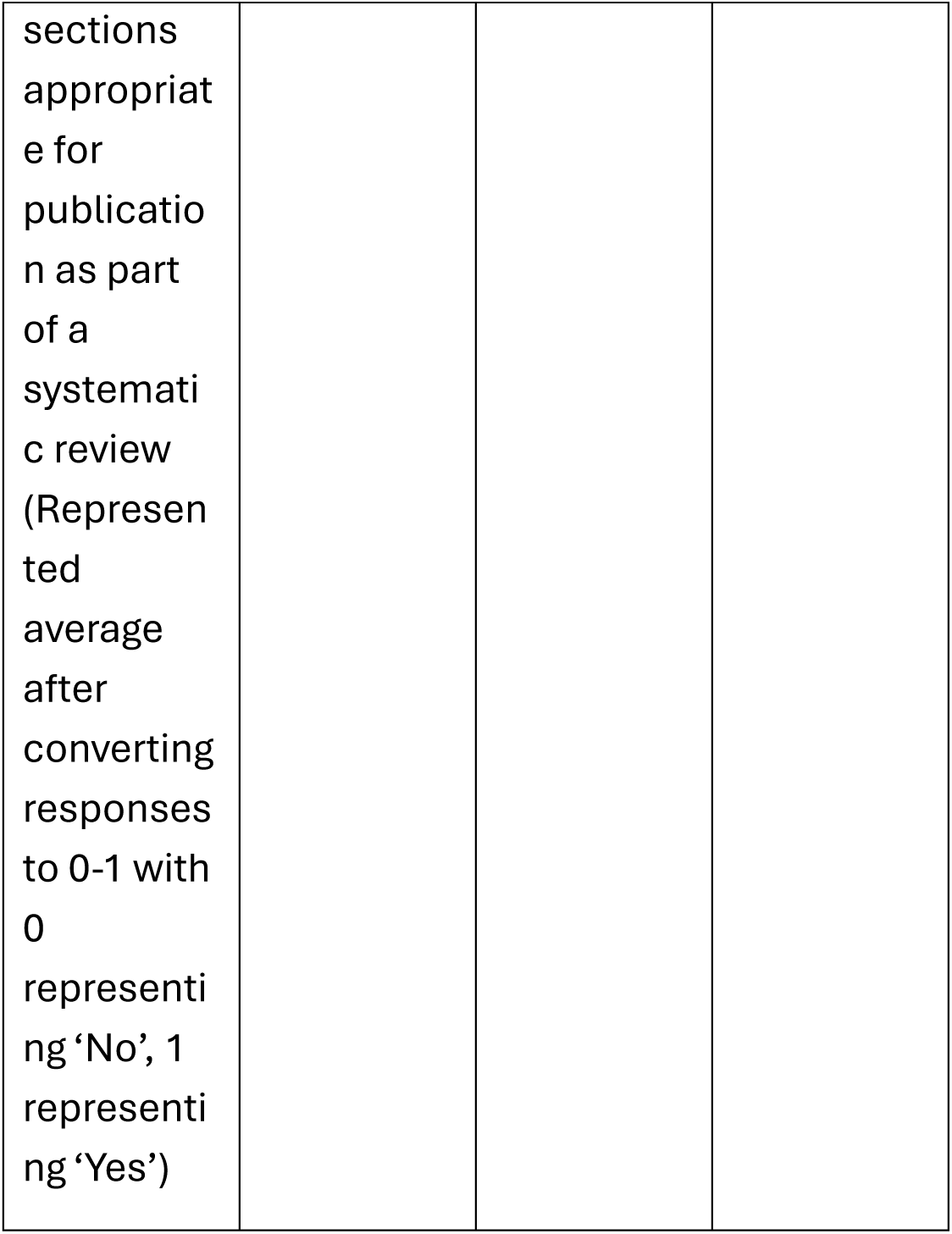
Survey Results.

**Figure 3.**
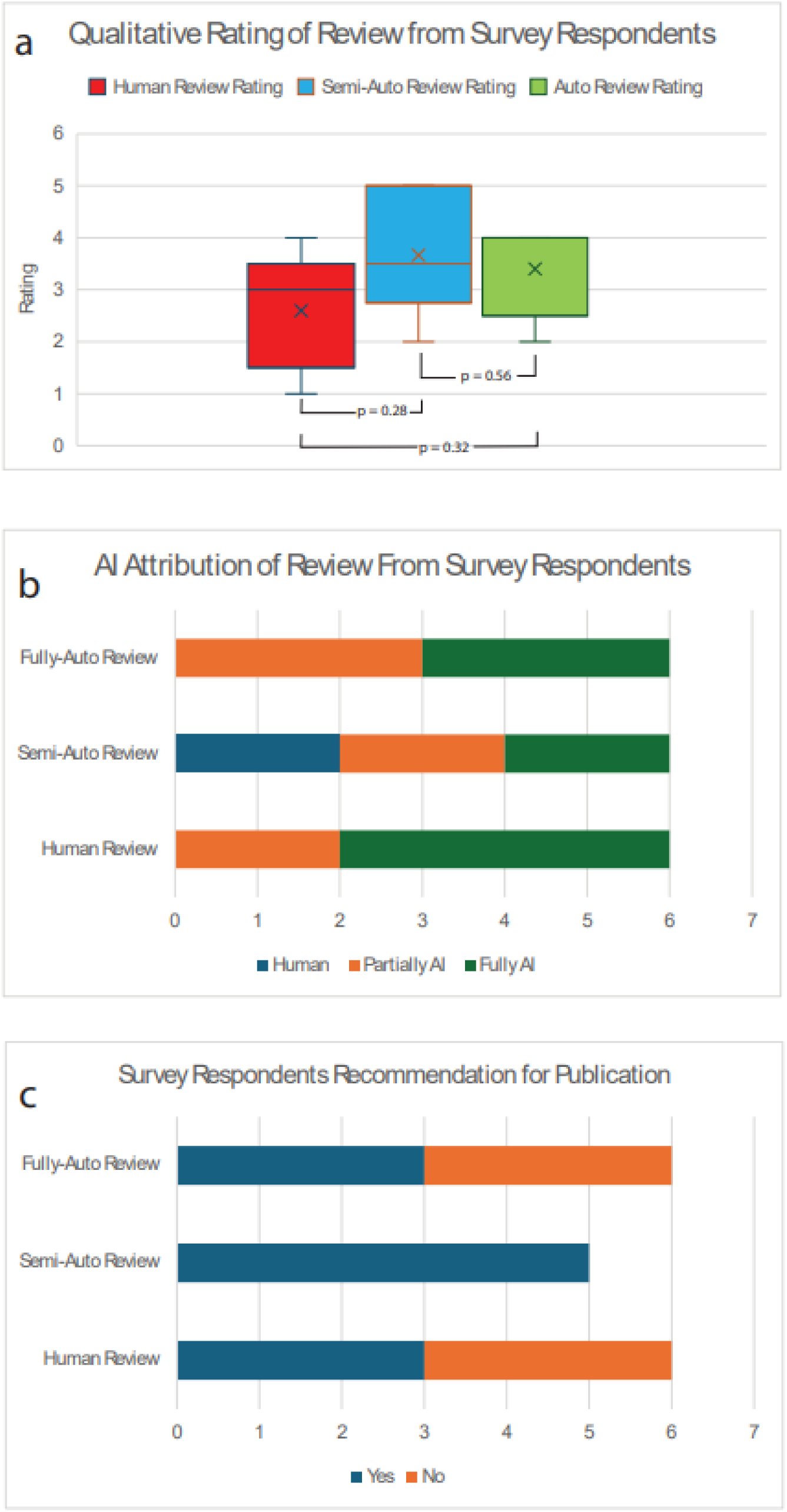
Survey Results. **a.** Qualitative scores responding to the question ‘Provide an overall quality of this review taking into consideration the overall content, quality of writing, and scientific merit. Scale: 1 (poor) to 5 (Excellent)’. The ‘Semi-Automated’ review (blue) scored the highest, followed by the ‘Fully-Automated’ review (green), followed by the ‘Human’ review (red). **b.** Frequency of AI-attribution for each paper. The ‘Semi-Automated’ review was identified as ‘human’ the most, while the ‘Fully-Automated’ and ‘Human’ reviews were never identified as ‘human’. **c.** Recommendation for publication for each review. The ‘Semi-Automated’ review was recommended for publication the most (5 yes, with 1 non-respondent), while the ‘Human’ and ‘Fully-Automated’ reviews were recommended for publication 3 times, and not recommended 3 times.

Asked to rate the reviews from 1 to 5 on overall content, quality of writing, and scientific merit, reviewers gave the highest mean rating to the ‘Semi-Automated’ review (μ = 3.66), followed by the ‘Fully-Automated’ review (μ = 3.4) and the human review (μ = 2.6). Wilcoxon signed-rank tests [19] found no significant differences among these ratings, which is expected at this sample size and does not establish equivalence. In an optional feedback section, reviewers commented favorably on the readability of the AI-produced reviews, particularly the ‘Semi-Automated’ review, which one reviewer described as having ‘flowed nicely’. One reviewer described the ‘Semi-Automated’ review as the most ‘trustworthy’, and another noted it best addressed the question posed in its title (”The Need for Standardization in Next-Generation Sequencing Studies for Classic Hodgkin Lymphoma”); the latter reviewer also suspected AI involvement in that review on the basis of its use of mini-headers. Criticisms of both AI-produced reviews included repetition, more pronounced in the ‘Fully-Automated’ review, which may reflect the piecemeal drafting process. The human review was criticized for addressing the stated question less directly and for inconsistency.

Reviewers most often judged the ‘Semi-Automated’ review to merit publication, with five endorsing publication and one not responding. The ‘Human’ and ‘Fully-Automated’ reviews were each endorsed by three reviewers and not endorsed by three.

Asked to what degree each review was written with AI, with options ‘fully human’, ‘partially AI’, and ‘fully AI’, the human review was described as ‘fully AI’ by four reviewers and ‘partially AI’ by two, and was never described as ‘fully human’. The ‘Semi-Automated’ review was described as ‘fully AI’ by two reviewers, ‘partially AI’ by two, and ‘fully human’ by two. The ‘Fully-Automated’ review was described as ‘fully AI’ by three reviewers and ‘partially AI’ by three, and was never described as ‘fully human’. In this panel, therefore, the review actually written by humans was attributed to AI most often, while the review drafted through the Claude Sonnet 4.0 chat interface was judged the most human. Overall, raters classified 27.8% of observations correctly, which did not exceed the 33.3% chance rate (binomial test [20], p = 0.77). Mean Cohen’s kappa [15] was −0.08, indicating no agreement with ground truth beyond chance. The mean signed difference between responses and ground truth was −0.39, consistent with a tendency to overestimate AI involvement. A cluster-aware permutation test yielded p = 0.84. These tests are summarized in **Table 4**. We interpret them as showing that this panel did not identify AI involvement above chance on this material, not as a general estimate of expert detection ability.

**Table 4.** Statistical Tests of Survey Results.

|  |  |
| --- | --- |
| **p-value (Wilcoxon Signed-Rank Test) of Human vs Semi-Automated** | 0.276303 |
| **p-value (Wilcoxon Signed-Rank Test) of Human vs Fully-Automated** | 0.317311 |
| **p-value (Wilcoxon Signed-Rank Test) of Semi Automated vs Fully-Automated** | 0.563703 |
| **p-value (Binomial Test) of AI Contribution Assessment Results** | 0.7689 |
| **Cohen’s Kappa of AI Contribution Assessment Results** | -0.083 |
| **Mean signed difference between Rater Responses and Ground Truth for AI Contribution Assessment** | -0.389 |
| **Permutation Test (10000 Randomized Iterations) for AI Contribution Assessment** | 0.8407 |

A complete set of survey results is provided in **Supplemental Table 8**.

### Internal Analysis of Citation Fidelity

For the lymphoma set, the ‘Fully-Automated’ review ultimately drew on 14 papers. Four of these overlapped with the sources used in the human review, and by extension the ‘Semi-Automated’ review, which used the human-assembled reference set; five were published after the human review; and five were published before it but not cited by it, equating to 44.4% concordance in citation with the human source set once later publications are excluded. All four overlapping papers were also cited in the ‘Semi-Automated’ review, indicating consistent source preference across both approaches. Five additional papers among the thirty cited in the human review were retrieved in the initial NCBI API batch but discarded for lacking retrievable full text; three of these were cited in the ‘Semi-Automated’ review. The ‘Semi-Automated’ review cited 18 of the 51 sources it was provided, a 35% concordance. For comparison, the ‘Semi-Automated’ triple-negative breast cancer review, which was not included in the survey, cited 54 of 149 provided sources.

Regarding fidelity, our methods yielded a citation misattribution rate of 7.06% in the ‘Semi-Automated’ review and 4.13% in the ‘Fully-Automated’ review. Hallucinations, defined as fabricated content not traceable to any source [21], were rare, with one instance identified in the ‘Semi-Automated’ review. The ‘Semi-Automated’ review also showed a tendency toward over-citation, in which a claim carried the correct citation alongside additional citations that did not support it, affecting 8.24% of claims. Additional results are given in **Table 5**, with the complete internal analysis in **Supplemental Tables 9, 10, 11, and 12.**

**Table 5.** Pertinent Internal Analysis Results.

| **Field** | **Semi-Auto Review**<br>**(Lymphoma); 51**<br>**Sources Seen at a**<br>**Time** | **Fully-Auto Review**<br>**(Lymphoma); 10**<br>**Sources Seen at a**<br>**Time** |
| --- | --- | --- |
| Total Claims (Cited) | 85 | 121 |
| Citation<br>Misattribution Rate<br>(%) | 7.06 | 4.13 |
| Citation<br>Overattribution Rate<br>(%) | 8.24 | 0* |
| Hallucinations<br>Observed | 1 | 0 |
| Used Same Sources as Human Review Rate (%) | 35** | 44.4*** |
| Sources Pulled from Pubmed | N/A | 493 |
| Pubmed Returned Empty Text | N/A | 307 |
| Number of Reviews Discarded By Claude Personally | N/A | 166 |
| Percentage of Available Sources Ultimately Used (%) | 35 | 70 |

\*There were debatable instances of over-attribution, but never blatant ones like in the Semi-Auto review
\*\*The Semi-Auto review could only see the sources in the human review; the percentage is what it opted to use.
\*\*\*Of papers that contained full texts. Five additional papers from the human review were discarded due to being textless in the pubmed automated search. Claude did not actually personally discard any reviews that appeared in the 'human review', tentatively demonstrating reliable inclusion standards for evaluating text inclusion.

## DISCUSSION

Our principal observations were as follows. **1.** The LLM applied explicit screening criteria consistently, with no instruction-following failures identified at the abstract stage and infrequent errors at the full-text stage, but agreement with human screeners depended on how the criteria were worded rather than arising by default. **2.** LLMs offer practical time savings on repetitive, text-bounded operations such as screening, summarization, and targeted extraction. **3.** In a small expert panel, LLM-produced review text was rated more favorably than a published human-authored review on the same topic for readability and topical focus, while showing more repetition and narrower informational breadth. **4.** That panel did not identify AI involvement above chance, and most often attributed the human-authored review to AI. **5.** Citation misattribution persisted at 4–7% of cited claims even under text-restriction strategies designed to minimize it.

We first consider what the screening comparison does and does not establish, then place the exploratory expert evaluation in the context of prior work, then discuss the constraints we encountered and the ethical implications that follow. We close with the platform we built in response to these findings and with directions for further study.

### 1. Interpreting the Screening Comparison

Our findings indicate that the model followed its provided inclusion and exclusion criteria consistently, with no observable errors at the abstract stage and infrequent errors during full-text review, the latter possibly reflecting the greater volume of text under consideration. The more informative finding concerns how the model interpreted those criteria relative to the human authors. The model applied criteria close to literally, whereas the human authors appeared to exercise discretion about when a literal reading should be relaxed. Neither behavior is more correct in itself, but the difference has a practical consequence: criteria written for a human audience may under-specify the decision rule an LLM will execute.

This suggests that criteria intended for LLM screening warrant more explicit drafting than criteria intended for human screeners, including statements about the edge cases a human would resolve by inference. It also suggests a defensible workflow. We would propose an initial abstract or full-text screening pass using deliberately lenient criteria that favor false positives over false negatives, on the reasoning that a human adjudicator can remove false positives efficiently while false negatives are, by construction, invisible. Where the resulting set remains too large for manual review, a human-verified subset of included and excluded papers can be used to tune the criteria until model behavior matches reviewer expectation, after which the tuned criteria can narrow the residual set to a size amenable to human adjudication. This workflow is summarized in **Figure 4**.

**Figure 4.**
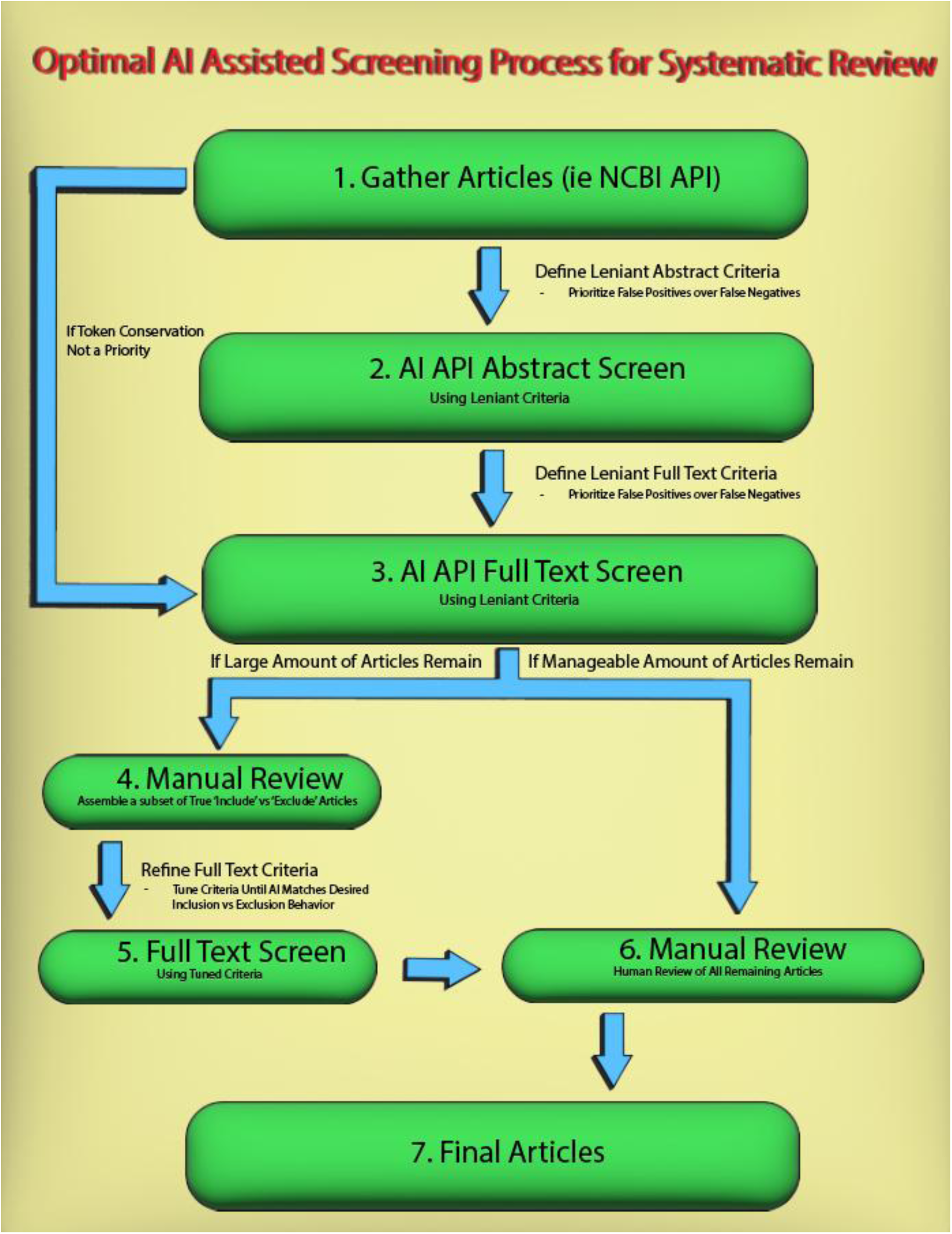
Recommended Pipeline for LLM-Assisted Systematic Review Screening. 1. Begin by gathering the desired set of articles, including their abstracts and full texts. 2. Next, define a set of detailed, overly lenient criteria to prioritize false positives over false negatives. Use these lenient criteria to screen the abstracts with the desired LLM, ideally with the API to allow for iterative screening. 3. Modify the lenient criteria to pertain to a full text screen, before using the AI API once more to screen the set of remaining full text articles. If token conservation is not an issue (i.e., a small number of articles in the first place), it is possible to skip from step 1 to step 3. 4. If the number of remaining articles remains very large, it may be a sign the criteria were overly lenient. At this point, the user is advised to manually review several of the articles, assembling a set of true positives and negatives based on their vision for the review. This small subset can be used to further tune the AI criteria until the LLM’s criteria cause it to accept the true positives and reject the true negatives in the subset. 5. The tuned criteria may then be applied to the remaining full-texts for a more tailored LLM screening process. 6. The user should then manually review any remaining articles for an ultimate inclusion decision. If the number of remaining articles from step 3 were already manageable, it is possible to skip from step 3 to step 6, foregoing criteria tuning. 7. The user has obtained a final set of articles in line with their vision for the systematic review.

**Figure 5.**
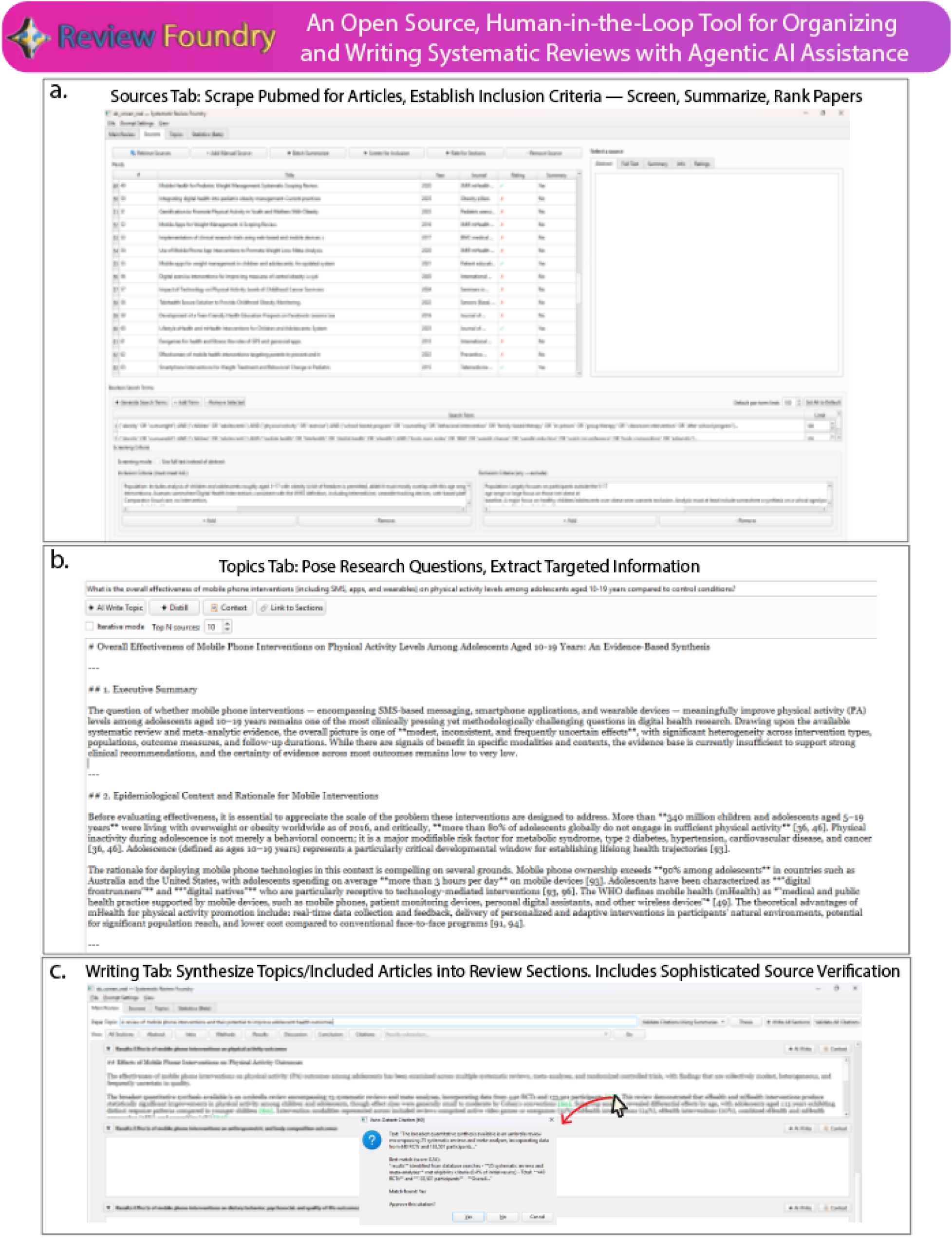
ReviewFoundry Interface. **a.** The sources tab can be used to organize sources pulled from the pubmed API or imported by the user. It can then summarize sources, screen them for inclusion, rate the sources for relevance to various review sections, and assess the bias for each source. **b.** The topics tab allows the user to pose targeted research questions, from which targeted information can be extracted from the set of included sources. **c.** The writing tab allows the user and an LLM assistant to work together in the drafting process. In this example, automated text-matching is being utilized to evaluate the accuracy of one of the citations the AI provided.

We stress that this is a proposal derived from a single benchmark comparison against one published review, using one model. The kappa values we obtained, 0.41 and 0.38, correspond to moderate and fair agreement respectively and are not high enough to support unsupervised screening. What the comparison does support is narrower: that the model can apply explicit criteria in a reproducible and potentially useful way, and that criteria wording, human oversight, and final human adjudication remain necessary components of the process.=

### 2. Interpreting the Exploratory Expert Evaluation

None of the quantified survey fields reached significance, which is unsurprising at n = 6 and should not be read as evidence of equivalence. Within those constraints, our reviewers showed a directional preference for the ‘Semi-Automated’ review, produced through the chat interface from human-gathered sources, over both the ‘Fully-Automated’ review and the published human review, and did not identify AI involvement above chance. We note that our panel did not consist of frequent AI users: asked to rate their own AI usage, three reported using AI less than once per month and one more than once a week, with two not responding. Familiarity with LLM output may plausibly affect detection, and our panel was not positioned to speak to that.

Although our sample was too small to support inference on its own, the direction of our findings is consistent with a body of published work on the perception of AI-generated scientific text. Gao et al. [22] reported that blinded human reviewers found it difficult to distinguish ChatGPT-generated abstracts from published ones, and that the abstracts reviewers suspected of being generated were characterized as vaguer and more formulaic. Casal and Kessler [23] found that 72 linguists and reviewers drawn from leading journals identified authorship correctly only 38.9% of the time. In a medical setting, Ren et al. [24] reported that healthcare professionals identified the origin of research abstracts correctly 43% of the time, with individual accuracy ranging from 20% to 57%. Our own accuracy figure of 27.8% falls within this range, and our observation that experience did not appear to confer advantage parallels these reports.

Our second observation, that the human-authored review was attributed to AI most often while an AI-produced review was judged the most human, also has precedent. A comparable pattern has been reported in the medical literature by Hakam et al. [25], who found that neither researchers nor AI-detection software identified AI-generated orthopaedic texts reliably, and that previously proposed textual criteria did not correlate with reviewers’ judgments. Work on labeling effects further suggests that these judgments are not driven by textual features alone: Zhu et al. [26] reported that raters who could not distinguish AI from human text in blind conditions nonetheless preferred text labeled as human-generated by a margin exceeding 30%, a preference that persisted when the labels were deliberately swapped.

Read against this literature, our survey results are best understood as one further instance of a consistently reported pattern rather than as an independent demonstration of it. The convergence is nonetheless relevant to the present study, since it suggests that the perceptual limitations we observed in a hematopathology panel are unlikely to be specific to this panel, this specialty, or this material.

### 3. Constraints Observed in LLM-Assisted Review Production 3a. Automated source retrieval introduces coverage gaps

Five papers cited in the human review were retrieved by the automated pipeline but discarded for lacking retrievable full text. Because many paywalled publishers do not support open API access, automated source gathering carries a systematic risk of coverage gaps that are invisible from within the pipeline. Manual verification of the retrieved corpus, including manual addition of missing full texts, is therefore necessary rather than optional.

### 3b. Model performance degrades with text volume, more so through the API than the chat interface

When presented with summaries from many sources at once, the API Sonnet models lost the ability to cite accurately. The failure mode was not primarily fabrication of factual claims; claims could usually be located somewhere in the provided sources, but were attributed to the wrong one, as though the model retained the content of the corpus but not the mapping between content and source. The chat interface versions did not show this weakness as prominently and maintained largely accurate citation across a greater volume of text, which is a notable difference between two deployments of the same underlying model. A more pronounced failure occurred with the Sonnet 3.5 API, which stopped following instructions when asked to generate results subsections from the full set of 189 source summaries selected for the triple-negative breast cancer review, instead summarizing what it had been shown. Behavior returned to expectation when the input was capped at 150 summaries. This was not retested on Sonnet 4.0, and we cannot distinguish between gradual degradation and a threshold effect.

### 3c. Text restriction reduces citation error at a cost to breadth

We addressed citation misattribution by having the API rate each source for its relevance to each results subsection and to the introduction, then exposing only the ten highest-rated sources when drafting each section. This reduced misattribution by the Sonnet 4.0 API to 4.13% of cited claims in the automated lymphoma review, compared with an earlier unranked attempt in which Sonnet 3.5 misattributed the large majority of its citations in the triple-negative breast cancer review. For contrast, the ‘Semi-Automated’ review drafted through the chat interface, which saw all 51 summaries simultaneously, showed a 7.06% misattribution rate, an 8.24% over-citation rate, and one hallucination [21].

Text restriction carried a cost. In the ‘Fully-Automated’ lymphoma review, restricting each section to its top-rated sources produced overlap between sections and, consequently, repetition of the same content across them. This was less pronounced in the more source-rich triple-negative breast cancer review but represents a structural trade-off: reducing the text available at any one point reduces error and simultaneously reduces the breadth of material any section can draw on. Repetition was also observed in the ‘Semi-Automated’ review, indicating that providing the accumulating manuscript for reference did not by itself prevent it.

We also encountered inconsistency between model versions. Sonnet 4.0 was more verbose than Sonnet 3.5 and required prompt revision to constrain word counts, and both models occasionally left excessive headers in their output. Prompt configurations tuned for one model should therefore not be assumed to transfer to another.

### 4. Ethical Implications

The findings above bear on a question that is, in our view, prior to any question of capability: under what conditions is LLM assistance in evidence synthesis defensible, and what obligations attach to its use?

The first obligation concerns verification. Our internal analysis found citation misattribution in 4.13% of claims in the best-performing automated configuration. A systematic review is consulted precisely because its claims are traceable to sources; a review in which roughly one claim in twenty-five points to the wrong source is not merely imperfect but misleading in a way that compounds when it is itself cited. The efficiency argument for automation is therefore conditional. Time saved on screening and summarization is genuine, whereas time saved by not verifying claims is borrowed against the reader. This distinction is not always made explicit in discussions of AI-assisted review, and we think it should be.

The second obligation concerns disclosure, and our exploratory findings speak directly to why it matters. If domain experts cannot identify AI involvement from the text, then readers cannot calibrate their scrutiny on the basis of the text alone, and disclosure becomes the only mechanism by which that calibration is possible. This matters more given evidence that AI use in published material is increasing [7], is not consistently reported [27], and carries stigma that may itself discourage reporting [28]. A norm in which AI assistance is common but undisclosed is worse than either consistent disclosure or consistent non-use, because it degrades the reliability of the inference readers draw from the absence of a disclosure statement.

Our panel’s attribution pattern raises a further concern. Reviewers rated the AI-produced text highest and judged it the most human, while rating the human-authored text lowest and judging it the most machine-like. Taken with prior reports that neither domain experts nor automated detectors reliably identify AI-generated academic text [25], this suggests that expectations about what AI writing looks like may be inaccurate in a specific direction: experts unfamiliar with current model output appear to anticipate writing that is less fluent than human writing, when the opposite may hold. Two consequences follow. First, informal AI detection during peer review is unreliable, and confident attribution based on prose style risks misjudging human authors, a risk likely to fall unevenly on authors whose writing conventions differ from a reviewer’s expectations. Second, if fluency is read as a marker of human authorship and of quality, then authors face an incentive to adopt AI-polished prose regardless of whether AI was used substantively, which would tend to increase undisclosed use in a publication culture already described as ‘publish or perish’ [29].

There is also a question of scale. The pipeline described here required a single script execution to move from topic to draft manuscript. We report this as a proof of concept rather than a recommendation, but the same capability is available to anyone with API access, and the low marginal cost of producing additional reviews is a structural feature rather than an artifact of our implementation. Reviews produced this way can be fluent, plausibly sourced, and still contain misattributions, omissions arising from retrieval gaps, and the narrowed breadth that text restriction imposes. The relevant safeguard is not detection after publication but verification before it, together with editorial norms that make the verification process itself reportable.

Finally, transparency obligations extend to the tools themselves. Proprietary AI research tools now in academic and clinical use, including platforms such as OpenEvidence [30] and ambient clinical documentation systems [31], generally do not expose the prompts, parameters, or intermediate decisions that produce their output. A reviewer or clinician relying on such a system cannot audit how a conclusion was reached. Where these tools inform evidence synthesis, we would argue that the same standards of traceability applied to the review itself should apply to the process that produced it, and that recommendations for ethical AI-assisted academic writing [6] would benefit from addressing tool opacity explicitly alongside author disclosure.

Taken together, our results support a limited and conditional role for LLM assistance in systematic review production. Bounded, text-restricted operations such as screening against explicit criteria, summarization of individual papers, and targeted extraction of specified information were performed reliably enough to be useful under supervision. Unsupervised synthesis was not, and our evidence does not support drafting whole review sections without claim-level human verification, given documented error and bias in AI-assisted scientific writing [32]. Where a fully automated draft is generated, we would regard it as a rapid personal literature orientation rather than a publishable artifact, and only where the reader independently verifies any claim before relying on it.

### 5. ReviewFoundry: A Platform Developed in Response to These Findings

The constraints described above are, for the most part, not addressable by prompt engineering alone. They are properties of the interface through which the model is used: chat interfaces do not restrict context systematically, do not expose their intermediate decisions, and do not provide a mechanism for verifying citations at the claim level. In response, we developed ReviewFoundry, an open-source Python application intended to support human-supervised, AI-assisted review workflows. It is available for Windows installation at https://github.com/mclaughlinliam3/AI-Systematic-Review-Foundry/releases.

ReviewFoundry decomposes the review process into discrete steps within a graphical interface, with the human user retained as the decision-maker at each step. We describe it here as an outcome of this study rather than as a validated instrument; it has not been evaluated against the workflows it is intended to replace, and such an evaluation would be necessary before any claim about its effect on review quality could be made. Its components are summarized in **Figure 4** and correspond to the specific problems set out in **Table 6**.

1. Sources Tab — **a.** A window to establish boolean search terms (with or without LLM assistance); **b.** organize articles by manual import or retrieval from PubMed via its API, including abstract, full text, and associated metadata; **c.** establish inclusion/exclusion criteria followed by either batch abstract or batch full-text screening with or without LLM assistance, including an LLM explanation of each decision; **d.** batch summarize articles with AI assistance; **e.** rank the relevance of remaining articles for incorporation into specific segments of the review, or for targeted information extraction within the topics tab.
2. Topics Tab — **a.** A window to establish and organize data from targeted research questions; **b.** create topics or research questions manually or with LLM assistance; **c.** configure which articles and components of articles (summaries versus full text, for example) are evaluated for targeted data extraction for each topic, with optional LLM inclusion assistance based on relevance ranking; **d.** extract all data pertaining to a topic from the included article set, either as one batch query or iteratively per article; **e.** further summarize topics using the LLM; **f.** link topic results to the corresponding sections of the review for reference during drafting.
3. Stats Tab — **a.** A window to establish and organize data from targeted statistical questions; **b.** similar to the topics tab but directed specifically at numerical and statistical data; **c.** established as a framework for potential meta-analysis features (beta).
4. Main/Writing Tab — **a.** A window to write and organize the components of the review in progress; **b.** establish a review topic and thesis to guide LLM behavior, and generate results subsections manually or with LLM assistance; **c.** configure which articles, article components, and topics are available for each section, with optional relevance-based inclusion assistance; **d.** draft review sections with or without LLM assistance; **e.** auto-organize all citations, and apply LLM assistance or automated text-matching to verify that cited sources support their claims; f. export the review text in a formatted document.
5. Additional Features — **a.** Select the LLM used, including current Anthropic Claude models or locally hosted models via Ollama [33]; **b.** view and configure all default prompts and parameters; **c.** import and export data from the sources, topics, and statistics tabs.

**Table 6.**
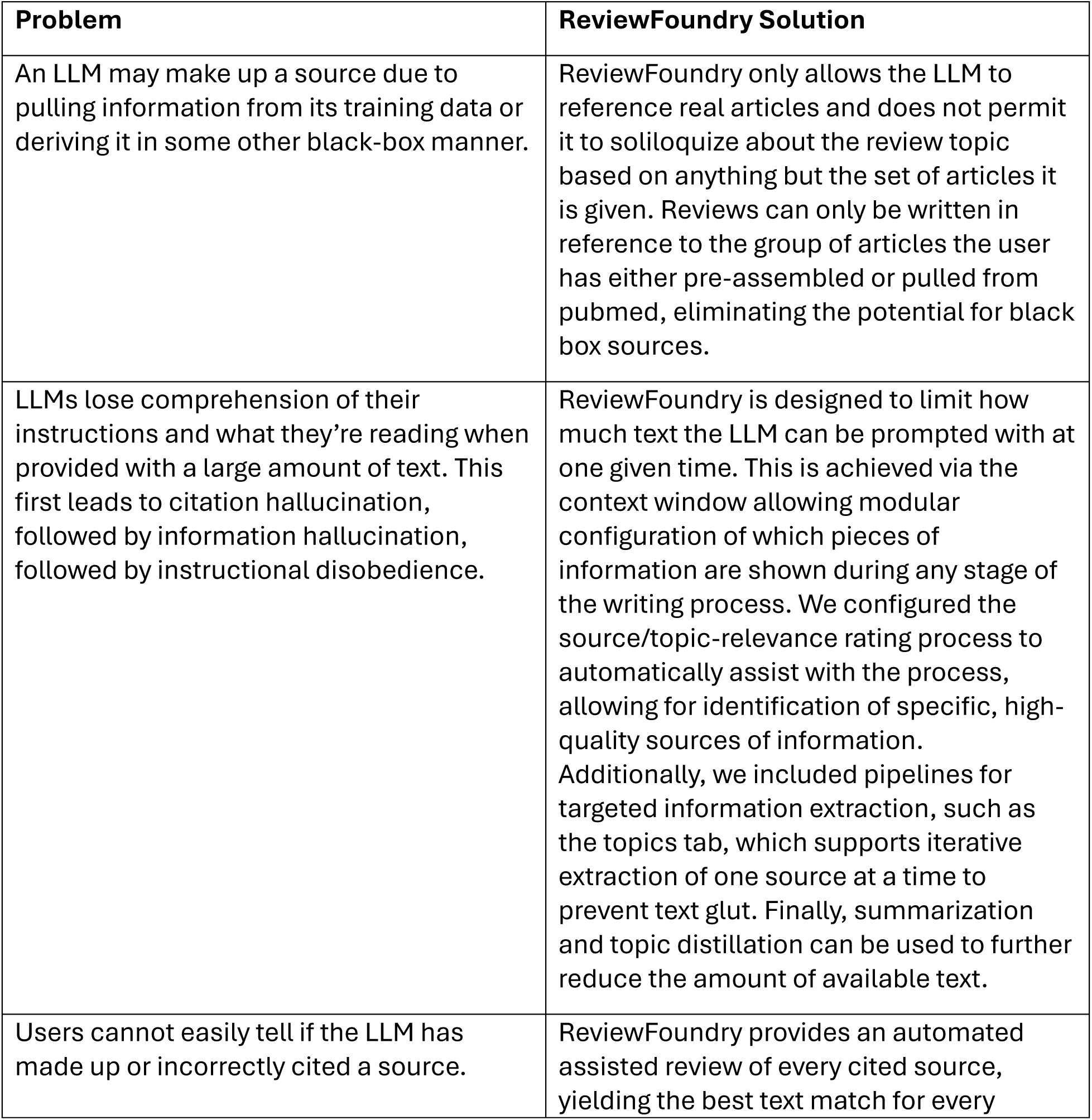
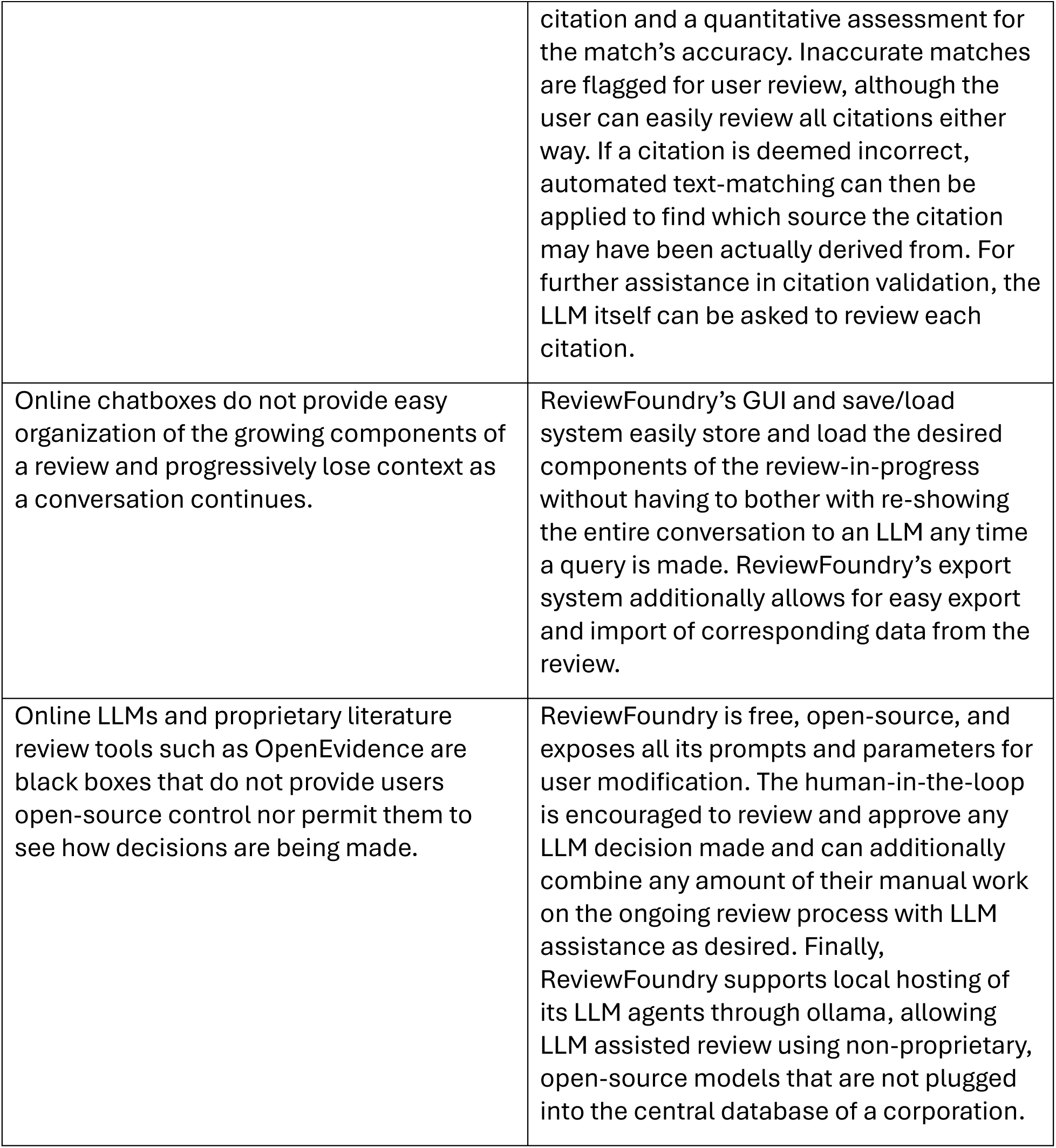
Issues with LLM-assisted Systematic Reviews addressed by ReviewFoundry.

## 6. Future Directions

Several questions raised by this work remain open, and we would identify the following as priorities.

First, the screening comparison should be extended across multiple published reviews, multiple clinical domains, and multiple model families before any general statement about LLM screening agreement is warranted. A single benchmark cannot distinguish model behavior from the idiosyncrasies of one author group’s criteria. A useful design would apply the same criteria-tuning procedure across a set of reviews with published screening decisions, which would allow the contribution of criteria wording to be separated from the contribution of model capability. Related to this, the criteria-tuning process itself deserves formalization: our procedure was iterative and investigator-driven, and a reproducible protocol, including a defined stopping rule and a pre-specified tuning subset, would make LLM-assisted screening auditable in a way that ours was not.

Second, the perceptual findings warrant a properly powered study. A larger panel, multiple topics, and reviewers stratified by self-reported AI familiarity would allow the question we could only raise, whether familiarity with LLM output improves detection, to be answered. A design in which the same reviewers assess both disclosed and undisclosed AI-assisted text would additionally allow the labeling effects reported elsewhere to be separated from judgments based on the text itself.

Third, citation fidelity requires a measurement standard. Our internal analysis was performed manually and is unlikely to have captured every error, and rates reported across studies are not currently comparable because misattribution, over-citation, and hallucination are not consistently distinguished. A shared taxonomy and an automated or semi-automated auditing procedure would allow error rates to be compared across pipelines, models, and interfaces. The difference we observed between API and chat-interface behavior on the same underlying model suggests that deployment configuration, not only model version, should be a reported variable in such comparisons.

Fourth, the trade-off between context restriction and informational breadth deserves direct study. We addressed misattribution by restricting each section to its ten highest-rated sources and observed increased repetition as a result. Whether retrieval strategies that select sources by content rather than by ranked relevance can reduce this trade-off, and how model context handling has changed across generations, are empirical questions that our design was not built to answer.

Fifth, ReviewFoundry itself requires evaluation. The appropriate comparison would assess review quality, error rates, and time to completion for reviews produced under human-supervised AI assistance against those produced by conventional methods, with claim-level verification performed by assessors blinded to production method. Until such a comparison exists, the platform should be understood as an implementation of the safeguards our findings suggested are necessary, rather than as a demonstration that those safeguards are sufficient.

Finally, we would note that the normative questions here are not resolvable by further measurement alone. What constitutes adequate verification, what level of AI involvement requires disclosure, and who bears responsibility for an error introduced by a model but published under a human author’s name are questions for journals, institutions, and professional bodies. Empirical work of the kind reported here can inform those decisions by establishing where the failure modes lie, but cannot substitute for them.

## Limitations

### Several limitations should be noted

Regarding the screening comparison, our benchmark consisted of a single published umbrella review screened by one model, and our results should not be taken as representative of LLM screening performance in general. Our recommendation of false-positive-biased criteria follows from this limitation rather than despite it: false positives are correctable by a human reviewer, whereas false negatives are not visible to one. Characterizing the difference between LLM and human screening would require a substantially wider set of reviews.

Regarding the survey, our sample of six reviewers assessed a single set of reviews on a single topic. The results are exploratory and describe trends that warrant further investigation rather than findings about the relative quality of AI and human writing. Formatting the human review to match the AI reviews for blinding purposes, including the removal of figures, may have introduced bias. Our decision to alter as little as possible of the AI-generated output, made to limit confounding, does not represent how many authors would use these tools in practice, such as revising prose they had already written. Our internal error analysis was performed manually and is unlikely to have identified every error present.

Regarding ReviewFoundry, we make no claim that a review produced with it will be free of error. It has not been evaluated against conventional review production, and its features address the failure modes we observed rather than a validated set of requirements. We stress human verification of all claims arising from any prompt, and transparency in any publication that uses AI assistance in its review process.

Finally, our evaluation covered the Claude Sonnet 3.5, 4.0, and 4.6 models, and our observations may not extend to other models or to later versions of these.

## Conclusion

This proof-of-concept study examined selected components of the systematic review process under LLM assistance. An LLM applied explicit screening criteria reproducibly, but reached only fair to moderate agreement with human screeners and did so only after its criteria were iteratively reworded, indicating that criteria specification and human adjudication remain necessary. In a small expert panel, LLM-produced review text was rated favorably and was not identified as AI-produced above chance, consistent with prior reports, though our sample does not support inference beyond hypothesis generation. Citation misattribution persisted at rates incompatible with unsupervised publication. These findings support a bounded role for LLM assistance in review production, conditioned on claim-level verification, transparent disclosure, and human responsibility for final decisions. ReviewFoundry was developed to make those conditions practicable within a single workflow, and its evaluation remains future work.

## METHODS

### Screening Analysis

All AI screening was performed with Claude Sonnet 4.6, the most recent Sonnet model available when this portion of the study was conducted.

### Full Text Screen

We assembled the available full texts for the articles that reached full-text screening in the umbrella review by Yabinze et al. [14]. Of the 91 articles, full texts were located for 86. Articles without full texts were discarded, except for one textless article that had been included in the original review and removed during its screening process. We assembled the inclusion and exclusion criteria as written by Yabinze et al. [14] into a prompt for the Claude Sonnet 4.6 API and used it to evaluate the nine articles included in the original umbrella review. On finding that all nine were rejected for what we judged to be valid reasons under a literal reading of the criteria, we systematically loosened the criteria on the basis of the model’s stated exclusion rationales until all nine were included; these were designated the ‘Most Lenient Criteria’. Applying these criteria to articles excluded by the umbrella review, we observed frequent inclusion of studies that addressed the target population only incidentally. To better match the apparent intent of the original criteria, we modestly reduced the leniency of the population criterion, yielding the ‘Lenient Criteria’. Both criteria sets were used to screen all 86 articles, with the model returning an inclusion decision and an explanation for each. Further agreement with the human authors might have been achievable through additional tuning, as the ‘Lenient Criteria’ were finalized before the full set had been screened, and we did not provide additional context such as the distinction between systematic review subtypes or an instruction against conflating intervention and outcome.

The AI screening results were compared with the original human screen. For each disagreement, we compared the umbrella review authors’ and the model’s stated rationales, then read the relevant portions of the article in question to assess the validity of each decision. Disagreements were classified as: 1. ‘Improper AI Criteria’, where the model followed its criteria as a human reasonably might but produced a false positive because the criteria remained undertuned; 2. ‘AI False Positive’, where the model accepted an article despite criteria that would reasonably have led a human to reject it; 3. ‘Human False Negative / Improper Criteria’, where the original authors appeared to have rejected an article inconsistently with their accepted set, or had not specified their criteria sufficiently for a reader to reconstruct the distinction; and 4. ‘Human False Positive / Improper Criteria’, the corresponding case for inclusion.

### Abstract Screen

As the umbrella review by Yabinze et al. [14] did not appear to report the set of studies assessed at abstract review, we assembled a synthetic dataset. We first used the boolean search terms provided by Yabinze et al. [14] to retrieve papers and abstracts from PubMed, then used additional boolean terms each designed to violate one inclusion criterion outright, such as restricting to an adult population. These sets were combined with the abstracts of the nine articles included in the umbrella review, with duplicates removed. The combined set was submitted to Claude Sonnet 4.6 alongside our ‘Most Lenient Criteria’, the same set used in the full-text review, with the model returning an inclusion decision and an explanation for each abstract. We assessed these decisions against what we judged a human screener would reasonably conclude from the same abstract, and against whether the model’s stated rationale was internally coherent. This assessment addressed instruction adherence rather than whether the accepted papers belonged in such an umbrella review.

The full criteria used across the screening evaluation, the model’s assessments, and our evaluation of those results are provided in Supplementary Tables 1–7.

### Automated Review Creation

Automated review creation was performed through controlled calls to the Anthropic Claude API within a single execution of a Python script, beginning with a user-supplied topic. During pipeline development we matched the topic of van den Ende et al.’s “Triple-Negative Breast Cancer and Predictive Markers of Response to Neoadjuvant Chemotherapy: A Systematic Review” [17] using the Claude API ‘Sonnet 3.5’ model. For the reviews presented to the hematopathologists we matched the topic of Santisteban-Espejo et al.’s “The Need for Standardization in Next-Generation Sequencing Studies for Classic Hodgkin Lymphoma: A Systematic Review” [18] using the Claude API ‘Sonnet 4.0’ model. Human reviews were selected as systematic reviews published within the previous three years in a freely available peer-reviewed journal with at least ten citations, with additional consideration given to whether their structure could match the subsection style of the automated reviews.

The AI-generated reviews were constructed in a piecemeal fashion using repeated prompts, the contents of which are given in **Supplemental Table 13.** For the ‘Fully-Automated’ review, prompts were issued with user-specified parameters including temperature, which governs the likelihood of a less conventional response, with lower values producing more conventional but less error-prone output [34], and a token limit governing response length. Parameter settings for each prompt are given in **Supplemental Table 14.**

With the topic set, the model generated boolean search terms for retrieval from PubMed via the NCBI API, using an API key, a parameter controlling the number of articles returned per term, and a relevance-based search method. Retrieved articles were stored as a Source class in Python, organizing title, abstract, text, and DOI. For the triple-negative breast cancer review, additional sources were retrieved via the Scopus API using the same relevance method; these proved largely redundant with the PubMed results, and many unique returns lacked full texts, so this step was not repeated for the lymphoma review.

Papers returning no full text were excluded from consideration. For all articles returning full text, we iteratively queried the model for an inclusion decision, returned as a boolean usable directly in code alongside a written explanation, both of which were saved to a CSV file. We also tested abstract screening prior to full-text screening but found it largely redundant within this pipeline.

We next iteratively queried the model to summarize the full texts of all papers passing the inclusion screen. Once summaries were obtained, the first 150 were presented to the model with a request to generate a series of results subsection headers to serve as anchor points for construction of the review. The 150-summary cap was introduced because larger inputs disrupted Sonnet 3.5’s instruction-following. For Sonnet 4.0, an additional instruction to produce only five subsections was added, as this model tended toward verbosity.

With the subsections defined, each source summary was again presented iteratively and rated for its usefulness to the introduction and to each results subsection. Each section was then drafted with only the ten highest-rated sources for that section visible to the model. This limited the number of papers available for any one subsection but was necessary for accurate citation, as citation accuracy degraded when the model was shown too many sources at once.

The introduction was drafted from its ten highest-rated summaries. Each results subsection was then drafted from its ten highest-rated summaries together with the accumulating manuscript. With the introduction and results complete, the full current manuscript was presented and the discussion drafted from it; source summaries were not provided at this stage, so that the discussion interpreted the material already presented in the results. The conclusion and then the abstract were drafted in reference to the accumulating manuscript. No methods section was generated, as the intended comparison was blinded.

Once the text was complete, Python was used to construct the reference section. Source numbers were reordered to match their order of appearance, and citations were assembled from PubMed metadata. The review was then reformatted and converted to a .tex file and compiled to PDF using TeXstudio.

### ‘Semi-Automated’ Review Creation

The ‘Semi-Automated’ review was generated through the Anthropic Claude web chat interface, drafted in a comparable piecemeal fashion but based on the same source set as the corresponding human-authored review, approximating how an author might use these tools in practice. To limit the text shown at once, the papers were first compiled manually into a spreadsheet containing their full texts and associated information. A Python script then converted the full texts to AI-generated summaries using the same summarization strategy as the automated pipeline.

All steps referencing summaries in the ‘Semi-Automated’ process were able to see the full set of summaries rather than a ranked selection, as the chat interface showed less tendency to misattribute sources when presented with abundant text. Summaries were organized by numerical header and saved as a JSON file, then uploaded to the chat interface, using Sonnet 3.5 for the triple-negative breast cancer review and Sonnet 4.0 for the lymphoma review. Exact prompts are given in **Supplemental Table 15**. The model was first prompted to write the introduction. It was then given the summaries and the accumulating manuscript and prompted to generate results subsections, then to draft each subsection. The discussion, conclusion, and abstract were drafted in the same manner. Reference assembly, reformatting, and PDF compilation followed the procedure described above.

### Survey Methods

Survey respondents were recruited by email from board-certified faculty of the University of Minnesota Department of Laboratory Medicine and Pathology hematopathology service, selected for expertise in the lymphoma subtype addressed by the reviews. The three reviews were assigned code names (‘Maroon and Gold’ for the human review, ‘North Star’ for the ‘Semi-Automated’ review, and ‘Paul Bunyan’ for the ‘Fully-Automated’ review) and truncated to the results and discussion sections. Reviews were provided in a blinded survey through Qualtrics.

Participant consent was obtained prior to participation. Respondents reviewed the three reviews at their own pace, each accompanied by a short set of questions. This included a screening question on prior familiarity with the human review, in which case the response would be excluded; no such case occurred.

### Internal Analysis

Citation misattribution was assessed by evaluating each cited claim made by the model. The corresponding papers and their summaries were checked for concordance with the claim, primarily by keyword matching. Statistical testing was performed in Python using the scipy stats module. Graphing was performed in Microsoft Excel.

### Statistical Testing

Statistical testing was performed in Python using the scipy and sklearn libraries.

For the screening comparisons, Cohen’s kappa [15] was computed on a contingency table of the results to quantify overall agreement. The same table was used to compute McNemar’s test [16] to assess differences in leniency.

For the qualitative ratings assigned by survey participants, the Wilcoxon signed-rank test [19] was used to test for differences, with the caveat that power was low at n = 6.

For assessment of reviewers’ ability to detect AI writing, we first conducted a one-sided exact binomial test [20] against a null probability of 1/3, reflecting equiprobable selection among three response categories; this treats each observation as independent. Second, Cohen’s kappa [15] was computed for each rater individually against ground truth to quantify agreement beyond chance, then averaged across the six raters. A kappa of 0 indicates chance-level agreement, positive values indicate agreement above chance, and negative values indicate systematic disagreement with ground truth. Third, the mean signed difference between rater responses and ground truth was computed across all 18 observations (response − truth). Given the ordinal coding (0 = fully AI-generated, 2 = human-written), a negative mean difference indicates a tendency to overestimate AI involvement. Finally, a non-parametric permutation test was conducted respecting the clustered data structure: in each of 10,000 iterations, ground truth labels were shuffled across articles while preserving the clustering of six ratings per article, and overall accuracy was recomputed, yielding a null distribution of accuracy under the hypothesis that responses are unrelated to true labels. The p-value was calculated as the proportion of permuted accuracies equal to or exceeding the observed accuracy.

### ReviewFoundry

ReviewFoundry was implemented in Python, primarily using PyQt6, a package for constructing graphical user interfaces, on the basis of the findings from the screening comparison and survey described above.

## Supplementary Data

**Supplementary Table 1 - Initial Criteria.** The set of criteria we initially used that were more or less taken verbatim from the authors of Yabinze et al. [14]

**Supplementary Table 2 - Initial Criteria Screen.** LLM Screening results using the initial criteria.

**Supplementary Table 3 - Most Lenient Criteria.** The set of screening criteria for the LLM screening study after the criteria were altered to accept all the articles also included by Yabinze et al. [14]

**Supplementary Table 4 - Most Lenient Criteria Screen.** LLM Screening results using the most lenient criteria.

**Supplementary Table 5 - Lenient Criteria.** The set of screening criteria for the LLM screening study after the criteria were altered to match the spirit of the original criteria of ‘Yabinze et al’ [14]

**Supplementary Table 6 - Lenient Criteria Screen.** LLM Screening results using the lenient criteria.

**Supplementary Table 7 - Abstract Screen.** LLM Screening results of abstracts using the most lenient criteria.

**Supplementary Table 8 - Survey Results.** Contains the Pertinent Survey Results from the 6 Hematopathologist Respondents to the Papers used in the Survey. Note that ‘North Star’ paper was the blinded name for the ‘Semi-Automated’ Lymphoma paper, the ‘Paul Bunyan’ paper was the blinded name for the ‘Human’ Lymphoma paper, and the ‘Maroon and Gold’ paper was the blinded name for the ‘Fully-Automated’ paper.

**Supplementary Table 9 - Internal Analysis Fully Auto Lymphoma.** Source Data Results of Internal Analysis of the ‘Fully Automated Lymphoma Paper’

**Supplementary Table 10 - Internal Analysis Semi Auto Lymphoma.** Source Data Results of Internal Analysis of the ‘Semi-Automated Lymphoma Paper’

**Supplementary Table 11 - Internal Analysis Fully Auto Triple Negative.** Source Data Results of Internal Analysis of the ‘Fully Automated Triple Negative Breast Cancer Paper’

**Supplementary Table 12 - Internal Analysis Correlation.** Source Data Results of Internal Analysis of Correlation of Cited Papers between the ‘Human’, ‘Semi-Automated’, and ‘Automated’ Lymphoma Papers; as well as between the ‘Semi-Automated’ and ‘Human’ Triple Negative Breast Cancer Papers

**Supplementary Table 13 - Fully Automated Prompts.** Source Data Prompts used in Automated Review Creation.

**Supplementary Table 14 - API Parameters.** Source Data Parameters Used for Automated Review Creation. Contains information detailing relevant Claude querying parameters for each step of the automated review creation process. The ‘Token Limit’ parameter controlled how verbose Claude’s response can be, while the ‘Temperature’ parameter controlled how ‘Creative’ Claude’s response was allowed to be.

**Supplementary Table 15 - Semi Automated Prompts.** Source Data Prompts used in Semi-Automated Review Creation

## Author Contributions

**L.M.** conceived the idea; **L.M.** wrote the first draft, generated the figures, performed the analysis, and generated the systematic reviews; **L.M.** wrote the code; **L.M.** and **M.W.** compiled source information necessary for creating the systematic reviews. **L.M. and C.A.**helped coordinate survey participants; L**.M., M.W. and C.A.** designed the survey; **L.M., M.W. and C.A.** came up with the study design for the blinded review; All authors edited the manuscript; **L.M.** and **C.A.** finished the final draft.

## Acknowledgements

We would like to thank the Hematopathologists at the University of Minnesota who agreed to participate in our survey.

## Ethics Declaration

This study was reviewed and approved by the Institutional Review Board of the University of Minnesota in accordance with the standards set by the Declaration of Helsinki. (IRB STUDY00024452).

All survey participants were provided with a consent form that gave a broad description of the study, what their participation entailed, and their rights as a participant. Consent to have participant results published was obtained via electronic signature prior to administration of the survey.

## Data Availability

All data reported on can be found in the Supplementary_Tables file. Additional copies of the surveys and the AI generated reviews will be available in the source data. On MedRxiv, these files are accessible by clicking ‘Data/Code’ on the upper-right side of the webpage. The human review used for comparison in the survey can be found in the article by Santisteban-Espejo et al. [18], although the version used for the survey included only the results, discussion, and references sections, with figures and tables removed. The human review used for the screening comparison can be found at Yabinze et al [14].

## Code Availability

The code, including a readme with instructions on usage, can be found in source data. A permanent repo for ReviewFoundry v1.0.5 is available here: 10.5281/zenodo.22232930 The main github page for ReviewFoundry is available here, including some user instructions: https://github.com/mclaughlinliam3/AI-Systematic-Review-Foundry

## Information for Correspondence

Cade Arries

