## Supplementary material for "In Search of Ethical Procedures for LLM-Assisted Systematic Review Production: A Proof-of-Concept Evaluation of Selected Review Components": source_data: fully_auto_lymphoma.pdf

### The Need for Standardization in Next-Generation Sequencing Studies for Classic Hodgkin Lymphoma: A Systematic Review

#### 1 Abstract

Classic Hodgkin lymphoma (cHL) presents unique molecular diagnostic challenges due to sparse neoplastic Hodgkin and Reed-Sternberg cells comprising only 0.1-10% of tumor cellularity. While next-generation sequencing has revolutionized understanding of cHL pathogenesis, significant methodological heterogeneity exists across studies, limiting clinical implementation and reproducibility.

**Purpose:** To evaluate the current landscape of next-generation sequencing methodologies in classic Hodgkin lymphoma studies and identify critical areas requiring standardization to optimize clinical implementation.

**Results:** Substantial heterogeneity was identified across all analytical domains. NGS platforms varied from Ion Torrent semiconductor systems to FoundationOne targeted sequencing, with gene panels ranging from focused 9-gene approaches to comprehensive 522-gene analyses. Sample preparation strategies demonstrated dramatic variability, from sophisticated laser microdissection requiring 15 hours per case to whole-tissue analysis without enrichment. Detection sensitivities varied markedly, with fresh frozen specimens achieving 88% clonality detection compared to 56% for FFPE materials. Variant calling thresholds ranged from 0.1% to >40% allele frequency, while coverage requirements varied from 100× to 600× median coverage. Quality control metrics and validation approaches showed substantial deficiencies, with inconsistent reference materials and cross-platform validation protocols.

**Conclusions:** The profound methodological heterogeneity across pre-analytical, analytical, and post-analytical domains fundamentally compromises clinical utility and reproducibility of NGS in cHL. Given that detection sensitivity variations from 25% to 88% directly impact diagnostic accuracy, and 29% of late recurrences represent clonally unrelated disease requiring different therapeutic approaches, comprehensive standardization represents the most critical priority for advancing precision medicine applications in classic Hodgkin lymphoma.

#### 2 Introduction

Classic Hodgkin lymphoma (cHL) represents a unique B-cell lymphoproliferative malignancy characterized by sparse neoplastic Hodgkin and Reed-Sternberg (HRS) cells comprising only 0.1-10% of tumor cellularity, surrounded by an abundant reactive inflammatory microenvironment [1,2]. This distinctive cellular composition has historically posed significant challenges for molecular characterization, as conventional clonality assays and mutation analyses are hampered by the low frequency of malignant cells among reactive B cells and plasma cells [1,3]. The advent of next-generation sequencing (NGS) technologies has revolutionized our understanding of cHL pathogenesis, revealing recurrent alterations in key signaling pathways including NF-$\kappa$B, JAK/STAT, and chromatin remodeling complexes [1,4].

Despite these advances, significant heterogeneity exists in NGS methodologies applied to cHL research. Studies have employed diverse approaches ranging from targeted gene panels covering 9-522 genes [4,5] to comprehensive genomic profiling platforms [2], with varying sample preparation techniques including HRS cell enrichment strategies [3,4] and whole-tissue analysis [6]. Detection sensitivities differ markedly between platforms, with NGS-based immunoglobulin clonality testing demonstrating 2.4-fold higher detection rates compared to conventional capillary electrophoresis methods [3,6]. Furthermore, circulating tumor DNA analysis has emerged as a promising liquid biopsy approach, though mutation detection rates vary from 70% using targeted panels [5] to higher rates with comprehensive sequencing approaches [2].

The clinical applications of NGS in cHL span multiple domains, including differential diagnosis of atypical presentations, establishing clonal relationships in recurrent disease, and identifying therapeutically targetable alterations [3,7]. However, methodological inconsistencies have led to variable results across studies. For instance, clonality detection rates range from 25% with conventional methods to 88% with optimized NGS protocols depending on sample type and technical approach [3,6]. Similarly, the identification of second primary lymphomas versus true relapses has been facilitated by molecular analysis, revealing that approximately 29% of late recurrences represent clonally unrelated disease [7].

The integration of germline predisposition analysis has further expanded the clinical utility of NGS, with novel variants such as those affecting PD-L2 identified in familial lymphoma cases [8]. Additionally, composite lymphoma presentations, though rare, benefit from comprehensive molecular characterization to distinguish between independent origins and clonal evolution [9]. The therapeutic implications of these findings are substantial, as targeted approaches based on pathway-specific alterations show promise in preclinical models [4].

Given the demonstrated clinical utility of NGS in cHL diagnosis, prognosis, and treatment selection, there is an urgent need to establish standardized protocols that ensure reproducible and clinically actionable results across different laboratories and healthcare systems. Therefore, the purpose of this systematic review is to evaluate the current landscape of next-generation sequencing methodologies in classic Hodgkin lymphoma studies and identify critical areas requiring standardization to optimize clinical implementation and improve patient outcomes.

### 3 Results

#### 3.1 Variability in NGS Platform Selection and Technical Methodologies Across Studies

The reviewed studies demonstrated substantial heterogeneity in NGS platform selection and technical methodologies employed for cHL molecular characterization. Platform diversity was evident across multiple sequencing technologies, with studies utilizing Ion Torrent semiconductor-based systems [4], ABI Ion GeneStudio S5 Plus sequencing systems [6], and FoundationOne targeted sequencing platforms [2]. This technological diversity extended to sequencing depth approaches, ranging from shallow whole-genome sequencing at ˜0.25x coverage for copy number alteration detection [10] to high-coverage targeted panels achieving 600× average median exon coverage [7].

Gene panel design varied dramatically across investigations, reflecting the lack of standardized target selection criteria. Studies employed panels ranging from focused 9-gene targeted approaches using AmpliSeq technology [5] to comprehensive 35-gene custom panels covering 353 amplicons [4], and extensive 405-gene cancer panels [2]. The EuroClonality-NGS Working Group's standardized protocols for immunoglobulin clonality testing represented a notable exception, providing consistent methodology across multiple studies [3,7]. However, even within clonality assessment, technical variations persisted, with some investigations utilizing LymphoTrack clonality assay systems [6] while others employed ARResT/Interrogate platforms [3].

Sample preparation methodologies exhibited significant variability, particularly regarding HRS cell enrichment strategies. Several studies implemented targeted enrichment approaches, including 1-mm-diameter puncher targeting of regions containing >10% tumor cells [4] and laser microdissection of CD30+ HRS cells combined with DEPArray™ digital sorting for single-cell analysis [3]. Conversely, other investigations analyzed whole-tissue specimens without enrichment [5,6], potentially impacting detection sensitivity given the sparse nature of malignant cells in cHL. Sample type preferences also varied, with some studies focusing exclusively on formalin-fixed paraffin-embedded specimens [2,4] while others compared fresh frozen and FFPE materials [3].

Detection thresholds and analytical parameters demonstrated considerable inconsistency across studies. Minimum coverage requirements ranged from 100× total coverage with 10× variant coverage [4] to more stringent criteria, while variant allele frequency thresholds varied from 0.1% near NGS noise thresholds [5] to >40% allele frequency filters designed to minimize germline variants [4]. Quality control measures also differed substantially, with some studies implementing duplicate sequencing procedures [4] and others relying on single-pass analysis. The establishment of clonality thresholds showed particular variation, with some investigations using normal distribution analysis from reactive lymphoid tissues [3] while others employed different statistical approaches for defining clonal populations [10,11].

#### 3.2 Inconsistent Sample Preparation and DNA Extraction Protocols for HRS Cell Analysis

The reviewed studies revealed substantial heterogeneity in sample preparation methodologies and DNA extraction protocols specifically designed for HRS cell analysis, reflecting the absence of standardized approaches for handling the unique cellular composition of cHL. Sample preparation strategies varied dramatically across investigations, with some studies implementing sophisticated HRS cell enrichment techniques while others analyzed whole-tissue specimens without targeted isolation. Several studies employed laser-capture microdissection (LCM) of CD30+ HRS cells combined with advanced sorting technologies, including DEPArray™ digital sorting for single-cell analysis [3] and PALM MicroBeam IV systems requiring approximately 15 hours per case to isolate 50-1000 HRS cells [12]. In contrast, other investigations utilized 1-mm-diameter puncher targeting of regions containing >10% tumor cells as a less labor-intensive enrichment approach [4], while additional studies analyzed whole-tissue specimens without any enrichment strategy [5,6].

DNA extraction protocols demonstrated considerable variability in both methodology and starting material requirements. Studies utilizing FFPE specimens employed diverse extraction approaches, including the Xiamen AmoyDx FFPE DNA Extraction Kit with DNA concentrations ranging from 46.2-147.2 ng/$\mu$L and OD 260/280 ratios of 1.7-2.0 [6], while other investigations used standard FFPE DNA extraction procedures with starting materials as low as 10 ng genomic DNA [4]. For microdissected samples, specialized extraction protocols were necessary, with some studies employing PicoPure kits designed for small cell populations obtained through laser-capture microdissection [12]. The QIAamp Circulating Nucleic Acid kit was utilized for cell-free DNA extraction from plasma samples, with volumes ranging from 0.38-2 mL plasma [13], while shallow whole-genome sequencing approaches required cfDNA extraction using standard protocols adapted from noninvasive prenatal testing workflows [10].

Sample type preferences and processing protocols varied significantly across studies, with important implications for detection sensitivity. Fresh frozen specimens consistently demonstrated superior performance compared to FFPE materials, with clonality detection rates of 88% versus 56% respectively [3], though most clinical samples are FFPE, making improved detection protocols crucial for routine implementation. Some investigations compared paired fresh frozen and FFPE specimens to validate consistency of results [3], while others focused exclusively on archival FFPE specimens reflecting real-world clinical scenarios [2,4]. The establishment of quality control measures also differed substantially, with some studies implementing duplicate DNA extraction and sequencing procedures to minimize false-positive rates [4], while others relied on single-pass analysis with different validation approaches [5,6].

The lack of standardized protocols for HRS cell enrichment and DNA extraction has resulted in variable detection sensitivities and inconsistent results across studies. While sophisticated microdissection approaches achieved high specificity by isolating pure HRS cell populations [3,12], these methods are technically demanding and time-intensive, limiting their clinical applicability. Conversely, whole-tissue analysis approaches are more practical for routine implementation but may suffer from reduced sensitivity due to dilution effects from the abundant reactive microenvironment [5,6]. These methodological inconsistencies underscore the critical need for standardized sample preparation protocols that balance analytical sensitivity with clinical feasibility for widespread adoption in cHL molecular diagnostics.

#### 3.3 Heterogeneous Gene Panel Design and Target Selection Strategies

The reviewed studies revealed substantial heterogeneity in gene panel design and target selection strategies employed across cHL investigations, reflecting the absence of standardized approaches for molecular characterization. Gene panel sizes varied dramatically, ranging from focused 9-gene targeted approaches using AmpliSeq technology [5] to comprehensive panels covering 35 genes across 353 amplicons [4], and extensive 405-gene cancer panels [2]. This variability extended to target selection methodologies, with some studies employing discovery-driven approaches where initial analysis of 522 lymphomagenesis-related genes informed subsequent custom panel design of 35 selected targets [4], while others utilized established commercial platforms such as FoundationOne targeted sequencing covering 405 cancer-related genes [2].

Target gene selection criteria demonstrated considerable inconsistency across investigations, with studies prioritizing different biological pathways and molecular mechanisms. Several studies focused on established cHL-associated pathways, including NF-$\kappa$B, JAK/STAT, and chromatin remodeling complexes, while others incorporated broader cancer gene panels without cHL-specific optimization [2,4]. The 9-gene targeted panel specifically selected commonly mutated genes in cHL, achieving 70% mutation detection rates with genes including SOCS1 (50% frequency), B2M (33.3%), TNFAIP3 (31.7%), STAT6 (23.3%), and ITPKB (23.3%) [5]. In contrast, comprehensive approaches identified alterations across 44 cancer-related genes, though specific frequencies varied substantially between targeted and broad-spectrum methodologies [2].

Platform-specific design considerations further contributed to methodological heterogeneity, with Ion Torrent semiconductor-based systems requiring different optimization strategies compared to other NGS technologies. Studies utilizing Ion Torrent platforms implemented custom amplicon designs with specific coverage requirements, including minimum 100× total coverage with 10× variant coverage thresholds [4], while FoundationOne approaches employed different analytical parameters suited to their proprietary methodology [2]. The establishment of variant detection thresholds also varied significantly, ranging from 0.1% variant allele frequency near NGS noise thresholds [5] to more stringent >40% allele frequency filters designed to minimize germline variant detection [4].

Quality control measures and validation strategies for gene panel performance showed substantial variation across studies. Some investigations implemented duplicate sequencing procedures with identical gene panels to minimize false-positive rates [4], while others relied on single-pass analysis with different validation approaches including Sanger sequencing confirmation of high-frequency mutations [4,5]. The integration of functional validation studies was inconsistent, with some studies incorporating cell line testing and protein expression analysis to confirm biological relevance of identified variants [4], while others focused primarily on detection rates without extensive functional characterization [2,5]. These methodological inconsistencies in gene panel design and target selection strategies underscore the critical need for standardized approaches that balance comprehensive coverage with clinical feasibility and analytical reliability across different laboratory settings and patient populations.

#### 3.4 Disparate Data Analysis Pipelines and Variant Calling Thresholds

The reviewed studies revealed substantial heterogeneity in data analysis pipelines and variant calling thresholds employed across cHL investigations, reflecting the absence of standardized bioinformatics approaches for molecular characterization. Variant calling methodologies demonstrated considerable inconsistency, with studies implementing dramatically different threshold criteria for mutation detection. Variant allele frequency (VAF) thresholds ranged from 0.1% near NGS noise thresholds for circulating tumor DNA analysis [5] to stringent >40% allele frequency filters specifically designed to minimize germline variant detection in tissue-based studies [4]. These disparate thresholds resulted in markedly different detection sensitivities, with some investigations prioritizing high sensitivity for liquid biopsy applications while others emphasized specificity for somatic mutation identification.

Coverage requirements and quality control parameters varied substantially across studies, contributing to inconsistent analytical reliability. Minimum coverage thresholds ranged from 100× total coverage with 10× variant coverage requirements [4] to more stringent criteria achieving 600× average median exon coverage for comprehensive mutation analysis [7]. Some investigations implemented duplicate sequencing procedures with identical analytical parameters to minimize false-positive rates [4], while others relied on single-pass analysis with different validation approaches including Sanger sequencing confirmation of high-frequency mutations [5]. The establishment of statistical thresholds for clonality determination showed particular variation, with studies using normal distribution analysis derived from reactive lymphoid tissues [3] compared to alternative statistical modeling approaches for defining clonal populations [10,11].

Bioinformatics software selection and analytical workflows demonstrated significant diversity across investigations, with studies utilizing platform-specific analysis tools including Torrent Suite programs with variant caller plug-ins for Ion Torrent systems [4], LymphoTrack Dx PGM software for clonality assessment [6], and ARResT/Interrogate platforms following EuroClonality-NGS Working Group protocols [3]. Copy number alteration detection employed shallow whole-genome sequencing approaches at ~0.25× coverage with specialized computational algorithms adapted from noninvasive prenatal testing workflows [10], while comprehensive genomic profiling utilized FoundationOne proprietary analytical pipelines with different parameter optimization strategies [2]. The lack of standardized reference materials and control samples further complicated cross-study comparisons, with some investigations establishing thresholds from reactive lymphoid tissue specimens [3] while others employed germline DNA control pools from limited case numbers [4]. These methodological inconsistencies in data analysis pipelines and variant calling thresholds underscore the critical need for standardized bioinformatics protocols that ensure reproducible and clinically actionable results across different laboratory settings and patient populations.

#### 3.5 Lack of Standardized Quality Control Metrics and Validation Approaches

The reviewed studies revealed substantial deficiencies in standardized quality control metrics and validation approaches across cHL NGS investigations, reflecting the absence of consensus guidelines for ensuring analytical reliability and clinical validity. Quality control thresholds demonstrated considerable inconsistency, with studies implementing dramatically different acceptance criteria for sample adequacy and sequencing performance. DNA quality requirements varied substantially, ranging from basic concentration measurements of 46.2-147.2 ng/$\mu$L with OD 260/280 ratios of 1.7-2.0 [6] to more stringent starting material requirements of 10 ng genomic DNA with specialized extraction protocols [4]. Sample success rates also differed markedly, with some investigations achieving nearly universal sample adequacy with only single failures [14], while others experienced variable success rates depending on specimen type and processing methodology.

Validation strategies showed significant heterogeneity across studies, with some investigations implementing comprehensive multi-layered approaches while others relied on minimal confirmation procedures. Several studies employed duplicate sequencing procedures with identical analytical parameters to minimize false-positive rates [4], while others utilized single-pass analysis with Sanger sequencing confirmation limited to high-frequency mutations [4,5]. The establishment of analytical thresholds varied substantially, with some studies using normal distribution analysis derived from reactive lymphoid tissue specimens to define clonality cutoffs [3], while others employed different statistical modeling approaches including random forest algorithms with cross-validation for diagnostic performance assessment [10]. Functional validation studies were inconsistently applied, with some investigations incorporating cell line testing and protein expression analysis to confirm biological relevance of identified variants [4], while others focused primarily on detection rates without extensive functional characterization.

Cross-platform validation and standardization efforts were notably limited, with most studies utilizing platform-specific analysis tools including Torrent Suite programs for Ion Torrent systems [4], LymphoTrack Dx PGM software for clonality assessment [6], and FoundationOne proprietary analytical pipelines [2]. The EuroClonality-NGS Working Group's standardized protocols represented a notable exception, providing consistent methodology across multiple studies for immunoglobulin clonality testing [3,7], though even within this framework, technical variations persisted in threshold establishment and quality control implementation. Reference material utilization showed considerable variation, with some studies establishing control pools from limited germline DNA samples [4], while others employed reactive lymphoid tissues for threshold determination [3] or utilized specialized reference samples including DNA oligonucleotides and clinical specimens for batch effect correction [14]. These methodological inconsistencies in quality control metrics and validation approaches underscore the critical need for standardized protocols that ensure reproducible, clinically actionable results across different laboratory settings and patient populations.

### 4 Discussion

Classic Hodgkin lymphoma (cHL) represents a unique B-cell lymphoproliferative malignancy characterized by sparse neoplastic Hodgkin and Reed-Sternberg (HRS) cells comprising only 0.1-10% of tumor cellularity, surrounded by an abundant reactive inflammatory microenvironment [1,2]. This distinctive cellular composition has historically posed significant challenges for molecular characterization, as conventional clonality assays and mutation analyses are hampered by the low frequency of malignant cells among reactive B cells and plasma cells [1,3]. The advent of next-generation sequencing (NGS) technologies has revolutionized our understanding of cHL pathogenesis, revealing recurrent alterations in key signaling pathways including NF-$\kappa$B, JAK/STAT, and chromatin remodeling complexes [1,4].

Despite these advances, significant heterogeneity exists in NGS methodologies applied to cHL research. Studies have employed diverse approaches ranging from targeted gene panels covering 9-522 genes [4,5] to comprehensive genomic profiling platforms [2], with varying sample preparation techniques including HRS cell enrichment strategies [3,4] and whole-tissue analysis [6]. Detection sensitivities differ markedly between platforms, with NGS-based immunoglobulin clonality testing demonstrating 2.4-fold higher detection rates compared to conventional capillary electrophoresis methods [3,6]. Furthermore, circulating tumor DNA analysis has emerged as a promising liquid biopsy approach, though mutation detection rates vary from 70% using targeted panels [5] to higher rates with comprehensive sequencing approaches [2].

The clinical applications of NGS in cHL span multiple domains, including differential diagnosis of atypical presentations, establishing clonal relationships in recurrent disease, and identifying therapeutically targetable alterations [3,7]. However, methodological inconsistencies have led to variable results across studies. For instance, clonality detection rates range from 25% with conventional methods to 88% with optimized NGS protocols depending on sample type and technical approach [3,6]. Similarly, the identification of second primary lymphomas versus true relapses has been facilitated by molecular analysis, revealing that approximately 29% of late recurrences represent clonally unrelated disease [7].

The integration of germline predisposition analysis has further expanded the clinical utility of NGS, with novel variants such as those affecting PD-L2 identified in familial lymphoma cases [8]. Additionally, composite lymphoma presentations, though rare, benefit from comprehensive molecular characterization to distinguish between independent origins and clonal evolution [9]. The therapeutic implications of these findings are substantial, as targeted approaches based on pathway-specific alterations show promise in preclinical models [4].

Given the demonstrated clinical utility of NGS in cHL diagnosis, prognosis, and treatment selection, there is an urgent need to establish standardized protocols that ensure reproducible and clinically actionable results across different laboratories and healthcare systems. Therefore, the purpose of this systematic review is to evaluate the current landscape of next-generation sequencing methodologies in classic Hodgkin lymphoma studies and identify critical areas requiring standardization to optimize clinical implementation and improve patient outcomes.

### 5 Conclusion

This systematic review demonstrates that the current landscape of next-generation sequencing applications in classic Hodgkin lymphoma is characterized by profound methodological heterogeneity that fundamentally compromises clinical utility and reproducibility. The documented variability across all analytical domains—from HRS cell enrichment strategies requiring 15 hours per case to whole-tissue analysis without enrichment, gene panels ranging from 9 to 522 targets, and variant calling thresholds spanning 0.1% to >40% allele frequency—renders cross-study comparisons virtually meaningless and prevents evidence-based clinical implementation.

The clinical implications are immediate and substantial. Detection sensitivity variations from 25% to 88% across different protocols directly impact diagnostic accuracy, while the identification that 29% of late recurrences represent clonally unrelated second primaries requiring different therapeutic approaches underscores the critical need for standardized molecular characterization. The superior performance of fresh frozen specimens (88% detection) compared to FFPE materials (56% detection) highlights the urgent need for optimized protocols suitable for routine clinical specimens.

Comprehensive standardization across pre-analytical, analytical, and post-analytical domains represents the most critical priority for advancing NGS applications in cHL. The successful implementation of EuroClonality-NGS Working Group protocols for immunoglobulin clonality testing provides a valuable framework, though broader consensus guidelines encompassing sample preparation, gene panel design, variant calling thresholds, and quality control metrics remain urgently needed. Without such standardization, the promising therapeutic implications of pathway-specific alterations and liquid biopsy applications cannot be reliably translated into clinical practice, ultimately limiting precision medicine advances for cHL patients.

13. Veltmaat N, Tan GW, Zhong Y, Teesink S, Terpstra M, Bult J, et al.. Molecu-

lar profiling of cell-free DNA from classic Hodgkin lymphoma patients identifies potential prognostic clusters and corresponds with disease dynamics.. Annals of hematology. 2025;104(3):1789-1800. doi: 10.1007/s00277-025-06328-8.

14. Talhouk A, Kommoss S, Mackenzie R, Cheung M, Leung S, Chiu DS, et al.. Single-Patient Molecular Testing with NanoString nCounter Data Using a Reference-Based Strategy for Batch Effect Correction.. PloS one. 2016;11(4):e0153844. doi: 10.1371/journal.pone.0153844.
