## Supplementary material for "In Search of Ethical Procedures for LLM-Assisted Systematic Review Production: A Proof-of-Concept Evaluation of Selected Review Components": source_data: fully_auto_triple_negative.pdf

### Triple-Negative Breast Cancer and Predictive Markers of Response to Neoadjuvant Chemotherapy: A Systematic Review

Systematic Review Team

January 11, 2025

#### 1 Abstract

Background: Triple-negative breast cancer (TNBC) is an aggressive subtype with limited therapeutic options. Neoadjuvant chemotherapy (NAC) has emerged as a standard approach, but response rates vary significantly. This systematic review synthesizes current evidence on predictive markers of response to NAC in TNBC.

Results: Tumor-infiltrating lymphocytes (TILs) consistently emerged as robust predictors of pathological complete response (pCR) and improved outcomes. Molecular profiling revealed distinct TNBC subtypes with differential NAC responses, with basal-like tumors showing higher pCR rates. Gene expression signatures, including Notch 5-TSPs and TNBC-RPS, demonstrated promise in predicting NAC response. Germline BRCA mutations were associated with improved response to platinum-based NAC. Imaging biomarkers, particularly from DCE-MRI and PET/CT, showed potential for non-invasive response prediction. Circulating tumor DNA dynamics strongly correlated with treatment response and outcomes. While pCR remained a strong predictor of long-term outcomes, its relationship with survival was complex. Toxicity profiles of novel NAC regimens, including immunotherapy combinations, highlighted the need for careful patient selection and management.

Conclusion: This review reveals a shift towards personalized NAC approaches in TNBC, integrating multiple biomarkers to predict response and guide treatment. Future research should focus on validating promising biomarkers, developing integrated prediction models, and exploring adaptive treatment strategies. Addressing these areas can lead to improved outcomes and reduced toxicity in TNBC patients undergoing NAC.

#### 2 Introduction

Triple-negative breast cancer (TNBC) is an aggressive subtype of breast cancer characterized by the lack of expression of estrogen receptor (ER), progesterone receptor (PR), and human epidermal growth factor receptor 2 (HER2) [1,2]. Representing 10-20% of all breast cancers, TNBC is associated with poor prognosis and limited therapeutic options [1,3]. Neoadjuvant chemotherapy (NAC) has emerged as a standard treatment approach for locally advanced and early-stage TNBC, with pathological complete response (pCR) being the optimal outcome [2,4].

The response to NAC in TNBC patients is heterogeneous, with only about 20% achieving pCR after standard chemotherapy regimens [1]. This variability in treatment response highlights the critical need for predictive biomarkers to guide treatment decisions and optimize patient outcomes [4]. Currently, there are no validated biomarkers in clinical use to predict NAC response in TNBC patients [2,4].

Recent studies have explored various molecular and genetic markers as potential predictors of NAC response in TNBC. These include tumor-infiltrating lymphocytes, gene expression signatures, and specific genetic alterations [1,4]. For instance, BRCA1/2 mutations have been associated with improved response to platinum-based chemotherapy and poly (ADP-ribose) polymerase inhibitors (PARPi) in TNBC patients [3].

Other promising biomarkers under investigation include matrix metalloproteinase-9 (MMP-9) levels in serum and tumor tissue [5], androgen receptor (AR) expression [2], and various gene expression-based signatures [4,6]. Additionally, the integration of clinical factors with molecular biomarkers has shown potential in improving prediction accuracy [6].

Despite these advances, the complex and dynamic nature of TNBC presents challenges in developing robust predictive biomarkers [7]. Tumor evolution during treatment and the heterogeneity of residual disease further complicate the landscape of biomarker development [7].

The purpose of this systematic review is to comprehensively evaluate the current state of research on predictive markers of response to neoadjuvant chemotherapy in triple-negative breast cancer. By synthesizing the available evidence, we aim to identify the most promising biomarkers and highlight areas for future research to improve treatment strategies and outcomes for TNBC patients.

### 3 Results

#### 3.1 Pathological complete response (pCR) rates

Pathological complete response (pCR) is a critical endpoint in evaluating the efficacy of neoadjuvant chemotherapy (NAC) for triple-negative breast cancer (TNBC). Several studies have reported varying pCR rates depending on the treatment regimens used and patient characteristics.

In a phase II randomized clinical trial (NACATRINE) comparing standard NAC with or without carboplatin, the addition of carboplatin resulted in a non-statistically significant increase in pCR rate from 27.4% to 41.1% (p=0.085) [8]. This trend towards improved pCR rates with carboplatin addition is consistent with other studies. For instance, a prospective, open-label, randomized phase II study comparing carboplatin plus paclitaxel (PC) with epirubicin plus paclitaxel (EP) found a significantly higher pCR rate in the PC arm (38.6% vs. 14.0%, p=0.014) [9].

The KEYNOTE-522 study, which evaluated the addition of pembrolizumab to NAC in early TNBC, reported pCR rates of 58.7% in the pembrolizumab group compared to 40.0% in the placebo group, with a treatment difference of 18.7% [10]. This significant improvement in pCR rates highlights the potential benefit of incorporating immunotherapy into neoadjuvant regimens for TNBC.

A real-world analysis of NAC plus pembrolizumab in early TNBC reported an overall pCR rate of 63.6% [11]. This study found that completion of $\geq$8 cycles of neoadjuvant pembrolizumab and younger age (<55 years) were independently associated with achieving pCR.

The impact of androgen receptor (AR) expression on pCR rates has also been investigated. A multi-center retrospective study reported that 60% of AR-negative (quadruple-negative) breast cancers achieved pCR compared to only 24% of AR-positive TNBC [2]. This suggests that AR status may be a potential predictive marker for NAC response in TNBC.

Tumor-infiltrating lymphocytes (TILs) have emerged as another potential predictor of pCR in TNBC. A retrospective analysis of 166 TNBC patients found that both stromal TILs (sTILs) and intratumoral TILs (iTILs) were significantly associated with pCR [12]. Every 10% increment in sTILs and iTILs increased the odds of achieving pCR.

In the CALGB 40603 trial, which tested the addition of carboplatin or bevacizumab to standard NAC, pCR rates were increased with both agents. However, despite the higher pCR rates, this did not translate into improved long-term outcomes [13].

Overall, these studies demonstrate that pCR rates in TNBC can be improved by incorporating platinum agents, immunotherapy, or both into standard NAC regimens. However, the relationship between increased pCR rates and long-term outcomes remains complex and requires further investigation.

#### 3.2 Factors associated with pCR

Several studies have identified various factors associated with pathological complete response (pCR) in triple-negative breast cancer (TNBC) patients undergoing neoadjuvant chemotherapy (NAC).

Tumor-infiltrating lymphocytes (TILs) have emerged as a significant predictor of pCR in TNBC. A retrospective analysis of 166 TNBC patients found that both stromal TILs (sTILs) and intratumoral TILs (iTILs) were significantly associated with pCR, with every 10% increment in sTILs and iTILs increasing the odds of achieving pCR [12]. Another study reported that high TILs levels (≥50%) were associated with better NAC efficacy in univariate analysis [14].

Androgen receptor (AR) expression has been identified as a potential predictive marker for NAC response in TNBC. A multi-center retrospective study of 95 stage II-III TNBC patients reported that 60% of AR-negative (quadruple-negative) breast cancers achieved pCR compared to only 24% of AR-positive TNBC [2]. This suggests that AR status may be a significant factor in predicting NAC response.

Germline BRCA (gBRCA) pathogenic variants have been associated with improved pCR rates in TNBC patients receiving platinum-based NAC. A retrospective study of 64 TNBC patients found that gBRCA carriers had significantly higher pCR rates (74.2%) compared to BRCA wild-type patients (48.5%, p=0.035) [15].

Several molecular and genetic markers have been identified as potential predictors of pCR. A study integrating tumor genome and microenvironment analysis found that Tumor Mutational Burden (TMB) and Tumor Neoantigen Burden (TNB) were higher in responders to NAC [16]. The ratio of CD8 T cells to M2 macrophages was also found to be predictive of better outcomes [16].

Platelet-to-lymphocyte ratio (PLR) and neutrophil-to-lymphocyte ratio (NLR) have been identified as potential predictive markers for NAC response in TNBC. A post hoc analysis of a randomized controlled clinical study found that patients with lower PLR and NLR had significantly higher tpCR rates [17].

MORC2 expression has been reported as a significant biomarker for predicting pCR to standard NAC in TNBC patients. A study of 50 locally advanced TNBC patients found that high MORC2 expression was associated with worse pCR rates in both univariate and multivariate analyses [14].

A nomogram incorporating multiple factors has been developed to predict pCR probability in breast cancer patients after NAC. The model includes ER status, Ki67 status, HER2 status, pre-NAC tumor size, and NAC cycle number [18]. Specific nomograms were also created for HER2-positive and TNBC subgroups [18].

Early clinical response (ECR) during NAC has been identified as a valuable predictor of pCR and long-term outcomes. A retrospective study of 237 TNBC patients found a strong association between ECR and pCR, with 54% of responders achieving pCR compared to only 7% of non-responders [19].

In conclusion, various factors including TILs, AR expression, gBRCA status, molecular markers, PLR/NLR ratios, MORC2 expression, and early clinical response have been associated with pCR in TNBC patients undergoing NAC. These findings contribute to the ongoing efforts to identify reliable predictive biomarkers for NAC response in TNBC.

#### 3.3 Biomarkers predictive of pCR

Several studies have investigated various biomarkers for their potential to predict pathological complete response (pCR) in triple-negative breast cancer (TNBC) patients undergoing neoadjuvant chemotherapy (NAC).

Tumor Mutational Burden (TMB) and Tumor Neoantigen Burden (TNB) have been identified as potential predictors of NAC response. A study integrating tumor genome and microenvironment analysis found that both TMB and TNB were higher in responders to NAC compared to non-responders [16]. This suggests that these genomic features may serve as biomarkers for predicting treatment efficacy in TNBC patients.

The ratio of tumor-infiltrating lymphocytes (TILs) has emerged as another promising predictor of NAC response. The same study reported that the ratio of CD8 T cells to M2 macrophages was predictive of better outcomes [16]. This finding highlights the importance of the tumor immune microenvironment in determining treatment response.

Gene expression-based signatures have shown potential in predicting NAC response. A study developed a Notch 5-TSPs signature consisting of 5 gene pairs, which demonstrated good performance in predicting pCR in TNBC patients, with AUC values of 0.76 and 0.85 in two testing sets [4]. This signature outperformed other published signatures and showed specificity for TNBC.

Another study introduced a computational framework to calculate a response-probability score (RPS) based on patient transcriptomics [6]. The TNBC-specific RPS (TNBC-RPS) achieved the highest accuracy in predicting response compared to 143 other signatures. When combined with clinical factors, TNBC-RPS demonstrated high prediction accuracy comparable to Oncotype DX and MammaPrint in ER-positive patients.

Protein biomarkers have also been investigated for their predictive value. A study examining p53, Ki-67, and Bcl-2 expression in TNBC patients found that overexpression of p53 and Ki-67 ($\geq$10%) was associated with higher pCR rates [20]. Specifically, p53 expression was strongly associated with increased response to NAC, while positive Bcl-2 expression was associated with poor survival and reduced chemotherapy sensitivity.

Androgen receptor (AR) expression has been identified as a potential predictive marker for NAC response in TNBC. A multi-center retrospective study reported that 60% of

AR-negative (quadruple-negative) breast cancers achieved pCR compared to only 24% of AR-positive TNBC [2]. This suggests that AR status may be a significant factor in predicting NAC response.

Genetic alterations have also been explored as potential predictors of NAC response. A study found that germline BRCA (gBRCA) pathogenic variants were associated with improved pCR rates in TNBC patients receiving platinum-based NAC [21]. gBRCA carriers had significantly higher pCR rates (74.2%) compared to BRCA wild-type patients (48.5%, p=0.035).

In conclusion, various biomarkers including TMB, TNB, TIL ratios, gene expression signatures, protein markers (p53, Ki-67, Bcl-2), AR expression, and genetic alterations (gBRCA) have shown potential in predicting pCR in TNBC patients undergoing NAC. These findings contribute to the ongoing efforts to identify reliable predictive biomarkers for NAC response in TNBC, potentially allowing for more personalized treatment approaches.

#### 3.4 Tumor-infiltrating lymphocytes (TILs) and pCR

Multiple studies have demonstrated a significant association between tumor-infiltrating lymphocytes (TILs) and pathological complete response (pCR) in triple-negative breast cancer (TNBC) patients undergoing neoadjuvant chemotherapy (NAC).

A retrospective analysis of 166 TNBC patients found that both stromal TILs (sTILs) and intratumoral TILs (iTILs) were significantly associated with pCR [12]. Every 10% increment in sTILs and iTILs increased the odds of achieving pCR. The optimal threshold for predicting pCR was 20% for sTILs and 10% for iTILs based on ROC curve analysis [12].

Another study of 143 TNBC patients identified TILs as one of the key predictive factors for pCR, along with histological grade, degree of necrosis, and small-cell feature [22]. A combined predictive model incorporating these factors achieved an area under the curve (AUC) of 0.777 [22].

The prognostic value of TILs was further confirmed in a study of young (<40 years), node-negative, chemotherapy-naïve TNBC patients [23]. Patients with high sTILs (≥75%) had excellent 10-year overall survival and low incidence of distant metastasis without systemic therapy [23].

However, interobserver variability in TILs assessment remains a challenge. A study involving 40 pathologists evaluating sTILs in 41 TNBC cases found intraclass correlation coefficients ranging from -0.376 to 0.947 (mean: 0.659), indicating substantial variability [24]. Despite this, high sTILs scores were significantly associated with pCR for 90% of the participating pathologists [24].

The composition of TILs may also be important in predicting NAC response. A study using multiplex fluorescent immunohistochemistry found that patients achieving pCR showed higher densities of intratumoral and stromal CD4+ T cells [25]. Multivariate analysis identified intratumoral CD4+ T cell density as an independent predictor of pCR [25].

Furthermore, the dynamics of TILs during NAC may provide additional prognostic information. The WSG-ADAPT TN trial found that both baseline (sTIL-0) and 3-week (sTIL-3) TIL measurements were associated with higher pCR rates [26]. Interestingly, sTIL-3 had a significant independent impact on invasive disease-free survival beyond pCR [26].

In conclusion, while TILs have consistently shown strong associations with pCR in

TNBC patients undergoing NAC, challenges remain in standardizing assessment methods and determining optimal thresholds for clinical decision-making. Future research should focus on refining TILs evaluation techniques and exploring the prognostic value of specific TIL subsets.

#### 3.5 Molecular subtypes and response to neoadjuvant chemotherapy

Several studies have investigated the relationship between breast cancer molecular subtypes and response to neoadjuvant chemotherapy (NAC). The PAM50-defined molecular subtypes have emerged as valuable predictors of pathological complete response (pCR) rates across different breast cancer subtypes [7].

In HER2-positive breast cancer, the HER2-enriched subtype has been associated with higher pCR rates compared to other subtypes [7]. For triple-negative breast cancer (TNBC), the basal-like subtype predicts higher pCR rates [7]. In contrast, the luminal A subtype consistently demonstrates low pCR rates across studies [7,27].

A retrospective analysis of 240 breast cancer patients receiving NAC with taxanes and anthracyclines found significant differences in pCR rates among molecular subtypes: luminal A (1.6%), luminal B (13.4%), HER2 (22.6%), and TNBC (23.8%) [27]. This study also reported that high Ki67 expression (>40%) and negative estrogen receptor (ER) status correlated with higher pCR rates [27].

The I-SPY 2 trial, which analyzed pre-treatment data from 736 patients across 8 treatment arms, identified 11 distinct protein signaling-based clusters with varying pCR rates and long-term outcomes [28]. This study developed a HER2 Activation Response Predictive Signature (HARPS) that could potentially identify TNBC patients likely to respond to HER2-targeted therapy [28].

A systematic review and meta-analysis of 101 studies including 19,708 Asian breast cancer patients confirmed that TNBC and HER2-enriched subtypes had higher pCR rates than luminal subtypes when treated with taxane-anthracycline (TA) and taxane-platinum (TP) regimens [29]. The luminal A subtype consistently showed the lowest pCR rates across studies [27,29].

While higher pCR rates are generally associated with improved outcomes, this relationship may vary among molecular subtypes. For instance, one study found that despite having the lowest pCR rate, luminal A patients demonstrated the highest disease-free survival (DFS) rate [27]. This highlights the complex relationship between molecular subtypes, treatment response, and long-term outcomes.

The Integrative Cluster (IntClust) classification has been reported to be more informative than PAM50 for pCR prediction [7]. Additionally, the Trastuzumab Risk (TRAR) signature has shown promise in predicting response to anti-HER2 therapies [7].

These findings underscore the importance of considering molecular subtypes in predicting response to NAC and tailoring treatment strategies. However, the dynamic nature of breast cancer and its heterogeneity necessitate ongoing research to identify subtype-specific resistance determinants and optimize treatment approaches [7].

#### 3.6 Imaging markers of treatment response

Several studies have investigated the use of various imaging modalities and techniques to predict and assess response to neoadjuvant chemotherapy (NAC) in breast cancer, particularly triple-negative breast cancer (TNBC).

Dynamic contrast-enhanced magnetic resonance imaging (DCE-MRI) has emerged as a promising tool for predicting pathological complete response (pCR) to NAC. A study by Braman et al. explored the use of radiomic textural analysis of intratumoral and peritumoral regions on pretreatment DCE-MRI to predict pCR [30]. The study found that combining intratumoral and peritumoral radiomic features outperformed individual regions in predicting response. Receptor subtype-specific analysis further improved prediction accuracy, with AUC values of 0.93 for both hormone receptor-positive/HER2-negative and triple-negative/HER2-positive subtypes [30].

Positron emission tomography/computed tomography (PET/CT) has also shown potential in predicting and assessing NAC response. A study by Groheux et al. investigated the use of [18F]fluorodeoxyglucose (FDG) PET/CT in predicting axillary pCR in clinically node-positive breast cancer patients [31]. While the study found that baseline axillary disease extent on PET/CT was not a significant predictor of axillary pCR, molecular subtype was identified as an important factor in axillary response to NAC [31].

The combination of MRI and PET/CT has been explored to optimize response monitoring during NAC. Pengel et al. evaluated different imaging scenarios for various breast cancer subtypes and found that the optimal approach varied by subtype [32]. For HER2-positive and triple-negative tumors, MRI alone was found to be optimal (AUC 0.76 and 0.74, respectively), while for ER-positive tumors, combining MRI with PET/CT in incomplete responders showed the best performance (AUC 0.82) [32].

Novel imaging techniques have also been investigated for their potential in predicting NAC response. A study by Cochran et al. explored the use of near-infrared (NIR) optical tomography and spectroscopy to predict pCR based on pretreatment total hemoglobin (tHb) levels and early changes in tHb [33]. The study found that combining tumor biomarkers with pretreatment tHb measurements provided strong predictors of NAC response, with the optimal assessment window being after 1-2 treatment cycles, depending on tumor subtype [33].

Synthetic MRI (syMRI), a technique allowing quantitative mapping of T1, T2, and proton density (PD) relaxation times, has been investigated in conjunction with apparent diffusion coefficient (ADC) maps. A study by Xie et al. found that the correlation between ADC and relaxation maps varied across breast cancer subtypes and treatment response groups [34]. Notably, in TNBC patients achieving pCR, moderate to strong correlations were observed between ADC and PD map features [34].

In preclinical studies, the use of [18F]fluorothymidine (FLT) PET has shown promise for early prediction of TNBC response to paclitaxel therapy. A study by Whisenant et al. found that [18F]FLT PET demonstrated better performance than [18F]FDG PET in discriminating between responders and non-responders in a TNBC xenograft model [35].

These findings highlight the potential of various imaging modalities and techniques in predicting and assessing NAC response in breast cancer, particularly TNBC. However, further research and validation in larger clinical cohorts are needed before these approaches can be implemented in routine clinical practice.

#### 3.7 Circulating biomarkers and treatment response

Several studies have investigated the utility of circulating biomarkers for predicting and monitoring response to neoadjuvant chemotherapy (NAC) in triple-negative breast cancer (TNBC) patients.

Circulating tumor DNA (ctDNA) has emerged as a promising biomarker for treatment response and outcomes. One study of 37 TNBC patients found that ctDNA clearance at mid-NAC was significantly associated with pathological complete response (pCR), with 58% of patients achieving ctDNA clearance at mid-NAC ultimately achieving pCR [36]. Another study of 26 TNBC patients detected ctDNA in 96% of patients at baseline and found that ctDNA levels fell dramatically after one cycle of NAC, especially in patients who would achieve pCR [37]. Importantly, ctDNA detection at the end of NAC was strongly predictive of residual tumor at surgery and indicated significantly worse relapse-free and overall survival [37].

Circulating tumor cells (CTCs) have also been examined as potential biomarkers. One study found that baseline CTCs were detected in 32.4% of TNBC patients, though no significant association was observed between baseline CTC status and tumor response or survival outcomes [36]. However, analysis of CTCs at disease progression revealed mostly non-conventional CTCs lacking epithelial markers, highlighting potential limitations of epithelial marker-based CTC detection methods [38].

Exosomal biomarkers are an emerging area of research. One study found that exosomal miR-20a-5p levels were increased in TNBC patient plasma compared to healthy controls and correlated with poorer prognosis [39]. Functional studies suggested exosomal miR-20a-5p may inhibit CD8+ T cell function and contribute to anti-PD-1 therapy resistance [39]. Another study examined exosome-associated cytokines and found that high levels of EV_APRIL, EV_CXCL13, and EV_VEGF-A were associated with shorter overall survival in TNBC patients who underwent NAC [40].

Serum protein biomarkers have also shown potential. One study found that serum matrix metalloproteinase-9 (MMP-9) levels decreased during NAC and correlated with tumor regression [5]. Lower platelet-to-lymphocyte ratio (PLR) and neutrophil-to-lymphocyte ratio (NLR) at baseline were associated with higher pCR rates in TNBC patients receiving NAC [17].

Finally, soluble HLA-G (sHLA-G) levels were found to increase significantly after chemotherapy in TNBC patients, with high post-chemotherapy sHLA-G levels associated with positive lymph node status and reduced progression-free and overall survival [41].

These studies highlight the potential of various circulating biomarkers for predicting and monitoring treatment response in TNBC. However, further validation in larger prospective studies is needed before clinical implementation.

#### 3.8 Long-term outcomes based on pCR status

Pathological complete response (pCR) after neoadjuvant chemotherapy (NAC) has been consistently associated with improved long-term outcomes in triple-negative breast cancer (TNBC) patients. Several studies have demonstrated the prognostic value of pCR in predicting disease-free survival (DFS) and overall survival (OS).

In a large retrospective study of 638 TNBC patients treated with NAC, those achieving pCR had significantly better 5-year DFS and OS compared to non-pCR patients, regardless of HER2-low or HER2-zero status [42]. For HER2-low patients, 5-year DFS was 84.3% for pCR vs 62.6% for non-pCR ($p=0.002$), and 5-year OS was 93.1% vs 73.0% ($p=0.002$). Similarly, for HER2-zero patients, 5-year DFS was 86.4% vs 51.4% ($p<0.001$) and 5-year OS was 95.7% vs 63.0% ($p<0.001$) for pCR vs non-pCR, respectively [42].

The prognostic impact of pCR was further confirmed in a study of 254 breast cancer patients treated with NAC, where pCR (RCB-0) was associated with improved relapse-free survival compared to patients with residual disease [43]. Notably, even minimal residual disease after NAC was associated with worse outcomes compared to pCR [43].

However, the relationship between increased pCR rates and long-term outcomes is not always straightforward. In the CALGB 40603 trial, which tested the addition of carboplatin or bevacizumab to standard NAC, both agents increased pCR rates but this did not translate into improved long-term outcomes [13]. This highlights the complex relationship between pCR and survival outcomes.

The impact of pCR on long-term outcomes may vary based on breast cancer subtype. In a study comparing cabazitaxel to paclitaxel as NAC, landmark analyses stratified by pCR showed no significant differences in outcomes between the two treatment arms [44]. This suggests that factors beyond pCR may influence long-term prognosis.

Interestingly, a retrospective analysis of 561 breast cancer patients treated with NAC found that breast-conserving surgery (BCS) did not negatively affect survival compared to mastectomy, even after adjusting for confounders [45]. This indicates that the type of surgery after NAC may not significantly impact long-term outcomes when pCR is achieved.

In conclusion, while pCR is generally associated with improved long-term outcomes in TNBC patients treated with NAC, the relationship is complex and may be influenced by factors such as treatment regimen, breast cancer subtype, and surgical approach. Further research is needed to fully elucidate the prognostic value of pCR in different clinical contexts.

#### 3.9 Toxicity and adverse events

Several studies reported on the toxicity and adverse events associated with neoadjuvant chemotherapy regimens for triple-negative breast cancer (TNBC).

In a phase II study evaluating nab-paclitaxel followed by epirubicin/cyclophosphamide, the most common adverse events were alopecia, anemia, neutrophil count decrease, and white blood cell decrease [46]. Grade 3 or higher adverse events occurred in 39.7% of patients, with neutropenia (39.7%), leukopenia (22.5%), and peripheral sensory neuropathy (9.7%) being most frequent [47].

A study comparing carboplatin plus paclitaxel (PC) to epirubicin plus paclitaxel (EP) found that both regimens were generally well-tolerated, but with differing toxicity profiles [9]. The PC arm had higher rates of thrombocytopenia and anemia, while the EP arm had higher rates of neutropenia and nausea/vomiting [9].

In a trial of panitumumab with carboplatin and paclitaxel, the most common grade 3-4 adverse events were neutropenia, skin rash, and hypomagnesemia [48]. Only 58.1% of patients received all planned doses of panitumumab due to toxicity [48].

A phase II study of neoadjuvant talazoparib monotherapy reported that 95.1% of patients experienced treatment-related adverse events [49]. The most common were anemia, nausea, fatigue, neutropenia, and alopecia, with 39.3% experiencing grade 3 anemia [49]. This led to 26.2% of patients discontinuing treatment before completion [49].

Real-world data on the pembrolizumab plus chemotherapy regimen from KEYNOTE-522 found a higher immune-related adverse event (irAE) rate of 54% compared to the clinical trial, with 12% experiencing grade $\geq$3 irAEs [50]. Approximately 90% of irAEs occurred during the neoadjuvant phase [50]. Additionally, 21% of patients discontinued pembrolizumab due to toxicity and about 50% received reduced relative dose intensity of chemotherapy [50].

These findings highlight the significant toxicity associated with neoadjuvant regimens for TNBC. Careful monitoring and management of adverse events is crucial to optimize treatment delivery and outcomes. Further research is needed to identify strategies to mitigate toxicity while maintaining efficacy.

### 4 Discussion

Triple-negative breast cancer (TNBC) is an aggressive subtype of breast cancer characterized by the lack of expression of estrogen receptor (ER), progesterone receptor (PR), and human epidermal growth factor receptor 2 (HER2) [1,2]. Representing 10-20% of all breast cancers, TNBC is associated with poor prognosis and limited therapeutic options [1,3]. Neoadjuvant chemotherapy (NAC) has emerged as a standard treatment approach for locally advanced and early-stage TNBC, with pathological complete response (pCR) being the optimal outcome [2,4].

The response to NAC in TNBC patients is heterogeneous, with only about 20% achieving pCR after standard chemotherapy regimens [1]. This variability in treatment response highlights the critical need for predictive biomarkers to guide treatment decisions and optimize patient outcomes [4]. Currently, there are no validated biomarkers in clinical use to predict NAC response in TNBC patients [2,4].

Recent studies have explored various molecular and genetic markers as potential predictors of NAC response in TNBC. These include tumor-infiltrating lymphocytes, gene expression signatures, and specific genetic alterations [1,4]. For instance, BRCA1/2 mutations have been associated with improved response to platinum-based chemotherapy and poly (ADP-ribose) polymerase inhibitors (PARPi) in TNBC patients [3].

Other promising biomarkers under investigation include matrix metalloproteinase-9 (MMP-9) levels in serum and tumor tissue [5], androgen receptor (AR) expression [2], and various gene expression-based signatures [4,6]. Additionally, the integration of clinical factors with molecular biomarkers has shown potential in improving prediction accuracy [6].

Despite these advances, the complex and dynamic nature of TNBC presents challenges in developing robust predictive biomarkers [7]. Tumor evolution during treatment and the heterogeneity of residual disease further complicate the landscape of biomarker development [7].

The purpose of this systematic review is to comprehensively evaluate the current state of research on predictive markers of response to neoadjuvant chemotherapy in triple-negative breast cancer. By synthesizing the available evidence, we aim to identify the most promising biomarkers and highlight areas for future research to improve treatment strategies and outcomes for TNBC patients.

### 5 Conclusion

Conclusion

This systematic review highlights the significant progress made in identifying predictive markers of response to neoadjuvant chemotherapy (NAC) in triple-negative breast cancer (TNBC), while also underscoring the complexity and challenges that remain in this field. The evidence synthesized from multiple studies points towards a multi-faceted approach to predicting NAC response, incorporating clinical, pathological, molecular, and imaging biomarkers.

Tumor-infiltrating lymphocytes (TILs) have emerged as one of the most consistent and robust predictors of NAC response and long-term outcomes in TNBC. The strong association between higher TIL levels and improved pathological complete response (pCR) rates underscores the critical role of the immune microenvironment in TNBC biology and treatment response. This finding has important implications for the integration of immunotherapy into NAC regimens, as demonstrated by the promising results of the KEYNOTE-522 study.

Molecular profiling of TNBC has revealed distinct subtypes with differential response patterns to NAC. The consistently higher pCR rates observed in basal-like TNBC compared to other subtypes highlight the heterogeneity within TNBC and emphasize the importance of molecular classification in treatment planning. The development of gene expression signatures, such as the Notch 5-TSPs and TNBC-RPS, offers promising tools for predicting NAC response, although their clinical implementation remains challenging.

The identification of germline BRCA mutations as predictors of response to platinum-based NAC opens avenues for personalized treatment approaches based on genetic profiling. This finding aligns with the concept of synthetic lethality and suggests potential for other DNA repair deficiency markers as predictive biomarkers.

Imaging biomarkers, particularly those derived from dynamic contrast-enhanced magnetic resonance imaging (DCE-MRI) and positron emission tomography/computed tomography (PET/CT), show promise in non-invasively predicting and monitoring NAC response. The potential to combine imaging features with molecular and clinical data could significantly enhance prediction accuracy, although standardization of imaging protocols remains a challenge.

Circulating biomarkers, especially circulating tumor DNA (ctDNA), have emerged as powerful tools for real-time monitoring of treatment response. The strong association between ctDNA clearance and pCR, as well as its prognostic value, suggests potential for ctDNA as a surrogate endpoint in clinical trials and a guide for adaptive treatment strategies.

While pCR remains a strong predictor of long-term outcomes in TNBC, the complex relationship between increased pCR rates and survival benefits observed in some trials highlights the need for caution in using pCR as a surrogate endpoint. This complexity underscores the importance of identifying additional biomarkers that can predict long-term outcomes beyond pCR.

The toxicity profiles associated with various NAC regimens, particularly those incorporating novel agents like immunotherapy or PARP inhibitors, emphasize the need for careful patient selection and management strategies to optimize treatment delivery and outcomes.

In conclusion, this systematic review reveals a shift towards a more personalized approach to NAC in TNBC, integrating multiple biomarkers to predict response and guide treatment decisions. Future research should focus on validating and standardizing promising biomarkers, developing integrated prediction models, investigating adaptive treatment strategies based on early response biomarkers, exploring novel combinations of targeted therapies and immunotherapies with standard NAC, and addressing the challenges of intratumoral heterogeneity and clonal evolution during treatment.

By advancing these areas, we can move closer to truly personalized neoadjuvant approaches for TNBC, ultimately improving patient outcomes while minimizing unnecessary toxicity. The findings of this review have significant implications for clinical practice, trial design, and future research directions in the field of TNBC treatment.

12. Ruan M, Tian T, Rao J, Xu X, Yu B, Yang W, et al.. Predictive value of tumor-infiltrating lymphocytes to pathological complete response in neoadjuvant treated

triple-negative breast cancers.. Diagnostic pathology. 2018;13(1):66. doi: 10.1186/s13000-018-0743-7.

24. Van Bockstal MR, François A, Altinay S, Arnould L, Balkenhol M, Broeckx G, et al.. Interobserver variability in the assessment of stromal tumor-infiltrating lymphocytes

(sTILs) in triple-negative invasive breast carcinoma influences the association with pathological complete response: the IVITA study.. Modern pathology : an official journal of the United States and Canadian Academy of Pathology, Inc. 2021;34(12):2130-2140. doi: 10.1038/s41379-021-00865-z.

nal.pone.0197754.
36. Chen JH, Addanki S, Roy D, Bassett R, Kalashnikova E, Spickard E, et al.. Monitoring response to neoadjuvant chemotherapy in triple negative breast cancer using circulating tumor DNA.. BMC cancer. 2024;24(1):1016. doi: 10.1186/s12885-024-12689-6.
37. Cavallone L, Aguilar-Mahecha A, Lafleur J, Brousse S, Aldamry M, Roseshter T, et al.. Prognostic and predictive value of circulating tumor DNA during neoadjuvant chemotherapy for triple negative breast cancer.. Scientific reports. 2020;10(1):14704. doi: 10.1038/s41598-020-71236-y.
38. Ortolan E, Appierto V, Silvestri M, Miceli R, Veneroni S, Folli S, et al.. Blood-based genomics of triple-negative breast cancer progression in patients treated with neoadjuvant chemotherapy.. ESMO open. 2021;6(2):100086. doi: 10.1016/j.esmoop.2021.100086.
39. Li W, Han G, Li F, Bu P, Hao Y, Huang L, et al.. Cancer cell-derived exosomal miR-20a-5p inhibits CD8. Cancer science. 2024;115(2):347-356. doi: 10.1111/cas.16036.
40. Jung HH, Kim JY, Cho EY, Lee JE, Kim SW, Nam SJ, et al.. A Retrospective Exploratory Analysis for Serum Extracellular Vesicles Reveals APRIL (TNFSF13), CXCL13, and VEGF-A as Prognostic Biomarkers for Neoadjuvant Chemotherapy in Triple-Negative Breast Cancer.. International journal of molecular sciences. 2023;24(21). doi: 10.3390/ijms242115576.
41. Hoffmann O, Wormland S, Bittner AK, Hölzenbein J, Schwich E, Schramm S, et al.. Elevated sHLA-G plasma levels post chemotherapy combined with ILT-2 rs10416697C allele status of the sHLA-G-related receptor predict poorest disease outcome in early triple-negative breast cancer patients.. Frontiers in immunology. 2023;14:1188030. doi: 10.3389/fimmu.2023.1188030.
42. Shi Z, Liu Y, Fang X, Liu X, Meng J, Zhang J. Efficacy and prognosis of HER2-Low and HER2-Zero in triple-negative breast cancer after neoadjuvant chemotherapy.. Scientific reports. 2024;14(1):16899. doi: 10.1038/s41598-024-67795-z.
43. Xu X, Zhao W, Liu C, Gao Y, Chen D, Wu M, et al.. The residual cancer burden index as a valid prognostic indicator in breast cancer after neoadjuvant chemotherapy.. BMC cancer. 2024;24(1):13. doi: 10.1186/s12885-023-11719-z.
44. Meyer-Wilmes P, Huober J, Untch M, Blohmer JU, Janni W, Denkert C, et al.. Long-term outcomes of a randomized, open-label, phase II study comparing cabazitaxel versus paclitaxel as neoadjuvant treatment in patients with triple-negative or luminal B/HER2-negative breast cancer (GENEVIEVE).. ESMO open. 2024;9(5):103009. doi: 10.1016/j.esmoop.2024.103009.
45. Simons JM, Jacobs JG, Roijers JP, Beek MA, Boonman-de Winter LJM, Rijken AM, et al.. Disease-free and overall survival after neoadjuvant chemotherapy in breast cancer: breast-conserving surgery compared to mastectomy in a large single-centre cohort study.. Breast cancer research and treatment. 2021;185(2):441-451. doi: 10.1007/s10549-020-05966-y.
46. Liu Y, Fan L, Wang ZH, Shao ZM. Nab-paclitaxel Followed by Dose-dense Epirubicin/Cyclophosphamide in Neoadjuvant Chemotherapy for Triple-negative Breast Cancer: A Phase II Study.. The oncologist. 2023;28(1):86-e76. doi: 10.1093/oncolo/oyac223.
47. Futamura M, Oba M, Masuda N, Bando H, Okada M, Yamamoto Y, et al.. Meta-analysis of nanoparticle albumin-bound paclitaxel used as neoadjuvant chemotherapy for operable breast cancer based on individual patient data (JBCRG-S01 study).. Breast cancer (Tokyo, Japan). 2021;28(5):1023-1037. doi: 10.1007/s12282-021-01238-9.
48. Yam C, Patel M, Hill HA, Sun R, Bassett RL, Kong E, et al.. Targeting the Epidermal Growth Factor Receptor Pathway in Chemotherapy-Resistant Triple-Negative

Breast Cancer: A Phase II Study.. Cancer research communications. 2024;4(10):2823-2834. doi: 10.1158/2767-9764.CRC-24-0255.

49. Litton JK, Beck JT, Jones JM, Andersen J, Blum JL, Mina LA, et al.. Neoadjuvant Talazoparib in Patients With Germline BRCA1/2 Mutation-Positive, Early-Stage Triple-Negative Breast Cancer: Results of a Phase II Study.. The oncologist. 2023;28(10):845-855. doi: 10.1093/oncolo/oyad139.

50. . Detrimental Impact of Chemotherapy Dose Reduction or Discontinuation in Early Stage Triple-Negative Breast Cancer Treated With Pembrolizumab and Neoadjuvant Chemotherapy: A Multicenter Experience. Clinical Breast Cancer. 2024. doi: 10.1016/j.clbc.2024.08.005.
