## Supplementary material for "In Search of Ethical Procedures for LLM-Assisted Systematic Review Production: A Proof-of-Concept Evaluation of Selected Review Components": source_data: North Star Review - Semi Auto Used in Survey.pdf

### The Need for Standardization in Next-Generation Sequencing Studies for Classic Hodgkin Lymphoma: A Systematic Review

#### 1 Results

##### 1.1 Methodological Heterogeneity in Sample Preparation and Tumor Cell Enrichment Strategies

The scarcity of Hodgkin and Reed-Sternberg (HRS) cells, comprising less than 1-2% of tumor cellularity, has necessitated diverse enrichment strategies across cHL NGS studies, resulting in significant methodological heterogeneity [5,12]. Multiple approaches have been employed to isolate HRS cells from the abundant reactive microenvironment, each presenting unique advantages and limitations.

Laser capture microdissection (LCM) has emerged as a widely adopted technique for HRS cell enrichment. Studies have successfully utilized LCM to isolate tumor cells from both frozen and formalin-fixed paraffin-embedded (FFPE) specimens, enabling targeted genomic analysis [5,11,12]. One study employed LCM to microdissect approximately 100,000 HRS cells from 10 cHL cases, demonstrating the feasibility of this approach for downstream whole-exome sequencing [3]. Another investigation used LCM to collect pools of 30 CD30+ HRS cells for comparative genomic hybridization analysis [4].

Flow cytometry-based cell sorting represents an alternative enrichment strategy. CD30+ cell selection has been employed to isolate HRS cells, though this approach may co-enrich some reactive cells that express CD30 [5,12]. One comprehensive study adapted flow cytometry protocols to sort HRS cells from cryopreserved clinical biopsy samples, successfully enabling exome sequencing from purified tumor cells [5].

Single-cell approaches have provided another dimension to tumor cell enrichment. Micromanipulation of individual HRS cells has been utilized for targeted genetic analysis, allowing precise isolation of morphologically confirmed tumor cells [17,18]. This technique has proven particularly valuable for clonality studies and analysis of immunoglobulin gene rearrangements.

The choice of sample type has also varied considerably across studies. While some investigations have utilized fresh-frozen tissue specimens that preserve high-quality nucleic acids [5], others have relied on FFPE samples that are more readily available in clinical practice but present challenges for molecular analysis [6,7,13]. The DNA degradation inherent in FFPE specimens may affect the sensitivity and reliability of variant detection.

These diverse enrichment strategies have implications for study outcomes and cross-study comparisons. The varying purity of HRS cell preparations may influence mutation detection rates and contribute to the observed inconsistencies in mutation frequencies across different investigations. Furthermore, the technical complexity and specialized expertise required for these enrichment methods may limit the standardization and reproducibility of cHL genomic studies.

#### 1.2 Technical Variability in Next-Generation Sequencing Approaches and Platform Selection

Based on the available literature, significant technical variability exists across NGS approaches and platform selection in classic Hodgkin lymphoma studies, contributing to inconsistent results and limiting cross-study comparisons.

NGS Platform Diversity and Coverage Strategies

Studies have employed diverse NGS platforms with varying technical specifications. Targeted sequencing approaches have ranged from focused gene panels covering 9-35 genes [4,7,11] to more comprehensive panels analyzing 42-106 genes [6,10]. One study utilized a 522-gene discovery panel before designing a custom 35-gene targeted approach [6], while others employed panels covering 106-121 genes specifically designed for B-cell lymphomas [7]. The coverage depth and sequencing platforms have also varied considerably, with studies using Ion Torrent PGM technology [7,11] and other platforms with different performance characteristics.

Variant Detection Thresholds and Quality Control

A critical source of variability stems from different variant allele frequency (VAF) thresholds and quality control measures. Studies have employed VAF cutoffs ranging from 0.5% [6,7] to higher thresholds, directly impacting mutation detection sensitivity. One investigation implementing unique molecular identifiers (UMIs) to reduce sequencing errors set a 0.5% VAF threshold [8], while another study using similar methodology detected variants with VAFs ranging from 0.6-42% [7]. The implementation of different error-correction strategies, including UMIs and various bioinformatics pipelines, has further contributed to methodological heterogeneity.

Analytical Pipeline Variations

Significant differences exist in bioinformatics approaches and variant calling algorithms across studies. Different filtering strategies, database selections, and cutoff criteria have been employed [2], potentially affecting the reproducibility of results. The lack of standardized analytical pipelines has been recognized as a major limitation requiring harmonization efforts [2].

Platform-Specific Limitations

Certain technical limitations have emerged from platform-specific approaches. Some NGS panels have excluded important primer sets, such as IGHV FR1/FR2 primers used in conventional assays [9], potentially affecting comprehensive genetic characterization. Additionally, the quality of nucleic acids and preanalytic treatment can significantly influence results across different platforms [2].

Implications for Standardization

The observed technical variability has highlighted the urgent need for standardized NGS approaches in cHL research. Recent consensus initiatives in mature lymphoid malignancies have emphasized the critical importance of technical standardization for reliable NGS implementation, including the establishment of minimal gene panels and quality control measures [2]. This methodological diversity has complicated cross-study comparisons and hindered the development of standardized diagnostic and prognostic biomarkers in cHL.

#### 1.3 Inconsistency in Mutational Frequency Reporting Across Studies

Based on the analysis of next-generation sequencing studies in classic Hodgkin lymphoma, substantial inconsistencies in mutational frequency reporting have emerged as a significant methodological concern across the literature.

SOCS1 mutations exemplify this variability, with reported frequencies ranging dramatically across studies. One investigation detected SOCS1 mutations in 50% of patients using a 9-gene targeted panel [10], while another study utilizing a 42-gene panel with unique molecular identifiers reported SOCS1 mutations in 28% of cases [8]. A pediatric cohort employing hybrid capture-targeted sequencing of 106-121 genes found SOCS1 mutations in 60% of all cases, representing 83% of circulating tumor DNA-positive cases [7]. The discrepancy becomes even more pronounced when examining circulating tumor DNA studies, where SOCS1 mutation rates varied from 28% to 83% of ctDNA-positive cases depending on the methodology employed [4,6,7,10].

Similar inconsistencies are observed for other frequently mutated genes. STAT6 mutations have been reported with frequencies ranging from 21-32% across different studies [4,5,6,7], while B2M alterations show variation from 19-33% depending on the cohort and technical approach [4,5,6,7]. TP53 mutations demonstrate particularly wide variation, with rates spanning from 9% in whole-exome sequencing studies [11] to 25% in certain tissue-based analyses [11,13], illustrating how different methodological approaches can yield substantially different results for the same genetic target.

The variability extends beyond individual gene frequencies to pathway-level analyses. While JAK/STAT pathway alterations are consistently identified as prevalent, the specific frequencies vary considerably. One comprehensive exome sequencing study reported JAK/STAT pathway mutations in 87% of cases [11], while targeted sequencing approaches have yielded different proportions depending on the genes included in their panels and the detection thresholds employed [4,6,7].

These inconsistencies appear to stem from multiple methodological factors, including differences in variant allele frequency thresholds, ranging from 0.5% in some studies [6,7] to higher cutoffs in others, varying gene panel compositions covering 9-522 genes [4,5,6,7,10,13], and different sample preparation methods including tissue-based versus liquid biopsy approaches. The implementation of error-correction strategies, such as unique molecular identifiers, has further contributed to methodological heterogeneity, with some studies employing these techniques while others rely on conventional sequencing approaches [6,10].

This substantial variation in reported mutation frequencies across studies highlights the critical need for standardized methodological approaches to enable meaningful cross-study comparisons and facilitate the development of reliable diagnostic and prognostic biomarkers in classic Hodgkin lymphoma.

#### 1.4 Divergent Analytical Pipelines and Variant Calling Methodologies

Systematic analysis of next-generation sequencing studies in classic Hodgkin lymphoma revealed substantial heterogeneity in analytical pipelines and variant calling methodologies across investigations. The implementation of diverse bioinformatics approaches has emerged as a critical source of methodological variation, directly impacting the reproducibility and comparability of genomic findings.

Variant allele frequency (VAF) thresholds represent a fundamental source of analytical divergence. Studies have employed markedly different detection thresholds, ranging from 0.5% in investigations utilizing unique molecular identifiers [6,7] to higher cutoffs in conventional sequencing approaches. One study implementing error-correction strategies through unique molecular identifiers established a 0.5% VAF threshold and detected variants with frequencies spanning 0.6-42% [7], while others using similar methodologies reported different sensitivity parameters, directly influencing mutation detection rates.

The application of error-correction methodologies has introduced additional analytical complexity. While some investigations have incorporated unique molecular identifiers to reduce sequencing artifacts and enhance variant calling accuracy [6,10], others have relied on conventional sequencing approaches without such error-correction strategies. This methodological divergence has contributed to observed inconsistencies in mutation frequencies across studies, particularly for low-frequency variants.

Bioinformatics pipeline variations extend beyond variant detection thresholds to encompass fundamental differences in analytical workflows. Significant differences exist in variant calling algorithms, filtering strategies, database selections, and quality control measures employed across studies [2]. The lack of standardized computational approaches has been recognized as a major limitation requiring immediate harmonization efforts, as different analytical pipelines can yield substantially different results from identical sequencing data [2].

Platform-specific analytical limitations have further complicated cross-study comparisons. Certain NGS panels have excluded important primer sets, such as IGHV FR1/FR2 primers conventionally used in clonality assays [9], potentially affecting comprehensive genetic characterization. Additionally, the quality of nucleic acids and preanalytic specimen treatment significantly influence analytical outcomes across different platforms, with each step of the NGS workflow potentially affecting final results [2].

The observed analytical heterogeneity has highlighted the urgent need for standardized computational approaches in cHL genomic studies. Recent consensus initiatives in mature lymphoid malignancies have emphasized the critical importance of technical standardization, including the establishment of harmonized bioinformatics pipelines and quality control measures [2]. This analytical diversity has complicated meaningful cross-study comparisons and impeded the development of reproducible diagnostic and prognostic biomarkers in classic Hodgkin lymphoma.

#### 1.5 Impact of Sample Type and Quality on Detection Sensitivity and Reproducibility

The quality and type of sample used for NGS analysis emerged as a critical factor influencing detection sensitivity and reproducibility across cHL studies. Multiple investigations have demonstrated significant challenges associated with different specimen types, with varying implications for mutation detection rates and technical success.

Studies utilizing fresh-frozen tissue specimens have generally achieved superior molecular analysis outcomes compared to formalin-fixed paraffin-embedded (FFPE) samples [6,7,12,13]. One comprehensive investigation successfully adapted flow cytometry protocols to sort HRS cells from cryopreserved clinical biopsy samples, enabling high-quality exome sequencing from purified tumor cells [5]. This approach yielded robust genomic data with median mutation counts of 244 somatic mutations per case, demonstrating the value of optimal sample preservation for comprehensive genomic characterization.

In contrast, FFPE specimens, while more readily available in clinical practice, present substantial analytical challenges due to DNA degradation inherent in the fixation process [6,7,13]. These quality limitations may significantly affect the sensitivity and reliability of variant detection, particularly for low-frequency mutations. The impact of DNA degradation becomes especially problematic when analyzing samples with already limited tumor cellularity, as is characteristic of cHL specimens containing <1-2% HRS cells.

Liquid biopsy approaches using circulating tumor DNA (ctDNA) have emerged as a promising alternative sample source, offering non-invasive genetic profiling capabilities [6,7,8,10]. Multiple studies have demonstrated the feasibility of detecting somatic variants in plasma samples, with detection rates ranging from 68-81% across different cohorts [6,7,10]. One investigation utilizing a 42-gene targeted capture panel with unique molecular identifiers successfully identified variants in 73.5% of ctDNA samples, with median variant allele frequencies of 4.2% [8]. Similarly, a pediatric cohort study detected HRS cell-derived variants in 75% of pre-therapy circulating cell-free DNA samples using hybrid capture-targeted NGS [7].

The quality of nucleic acids and preanalytic specimen treatment significantly influence analytical outcomes across different platforms, with each step of the NGS workflow potentially affecting final results [2]. These sample-related factors contribute to the observed methodological heterogeneity and may explain some of the inconsistencies in mutation frequencies reported across studies. Recent consensus initiatives have emphasized the critical importance of standardized preanalytic procedures and quality control measures to ensure reproducible results across different laboratories and platforms [2]. The establishment of harmonized sample handling protocols represents an essential step toward improving the reliability and comparability of NGS-based studies in cHL research.

### 2 Discussion

This systematic review provides compelling evidence for the urgent need to establish standardized methodological approaches in next-generation sequencing studies of classic Hodgkin lymphoma. Our analysis reveals pervasive methodological heterogeneity across multiple dimensions of NGS implementation that fundamentally undermines the reproducibility and clinical utility of genomic findings in cHL research.

The Cascade of Methodological Heterogeneity

The methodological diversity identified in this review creates a cascade of confounding factors that collectively compromise the reliability of cHL genomic studies. Beginning with the fundamental challenge of HRS cell scarcity comprising less than 1-2% of tumor cellularity [10,12] the necessity for tumor enrichment has spawned multiple divergent approaches including laser capture microdissection, flow cytometry sorting, and CD30+ cell selection [10,12,13]. Each enrichment strategy yields preparations of varying tumor cell purity, directly influencing downstream mutation detection rates and contributing to the observed inconsistencies in genomic findings across investigations.

This enrichment-related variability is compounded by substantial technical diversity in NGS platform selection and sequencing strategies. The range from focused 9-gene panels [8,13,14] to comprehensive 522-gene discovery approaches [8] represents more than a 50-fold difference in genomic coverage, fundamentally altering the scope and sensitivity of mutational detection. The implementation of varying coverage depths, different sequencing platforms including Ion Torrent PGM technology [13,14], and disparate quality control measures further amplifies this technical heterogeneity, creating conditions where identical biological samples could yield markedly different genomic profiles depending on the analytical approach employed.

Critical Impact on Mutation Frequency Reporting

The methodological heterogeneity documented in this review has translated into profound inconsistencies in mutation frequency reporting that threaten the validity of cross-study comparisons and meta-analyses. The dramatic variation in SOCS1 mutation frequencies ranging from 28% to 83% of ctDNA-positive cases depending on methodology [8,14,15,16] exemplifies how technical choices can fundamentally alter biological conclusions. Similarly, the 16-fold variation in TP53 mutation rates (9-25%) across different methodological approaches [10,13,17] demonstrates that current reporting cannot be considered reliable for biomarker development or clinical decision-making.

These inconsistencies extend beyond individual gene frequencies to pathway-level analyses, where JAK/STAT pathway alterations show substantial variation despite consistent identification as prevalent features of cHL pathogenesis [8,10]. The observed frequency disparities for B2M (19-33%) and STAT6 (21-32%) mutations across studies [8,10,14,15] further underscore the systematic nature of this methodological crisis. Such variability effectively negates the possibility of establishing evidence-based mutation frequency thresholds for diagnostic or prognostic applications.

Analytical Pipeline Divergence as a Critical Bottleneck

The analytical heterogeneity identified in this review represents perhaps the most tractable yet critically important barrier to standardization. The implementation of variant allele frequency thresholds ranging from 0.5% in error-corrected approaches [14,15] to substantially higher cutoffs in conventional methods directly determines mutation detection sensitivity and creates systematic biases in comparative analyses. The differential application of error-correction strategies, including unique molecular identifiers in some studies [15,16] but not others, introduces fundamental disparities in analytical sensitivity that cannot be reconciled through post-hoc statistical adjustments.

The diversity in bioinformatics pipelines, encompassing different variant calling algorithms, filtering strategies, and database selections [2], creates a computational black box effect where identical sequencing data could yield different variant calls depending on the analytical workflow employed. This analytical heterogeneity has been recognized as a major limitation requiring immediate harmonization efforts in mature lymphoid malignancies [2], and our findings demonstrate that cHL research faces identical challenges with potentially greater complexity due to the unique tumor biology and technical requirements of HRS cell analysis.

Sample Type and Quality: The Foundation of Analytical Reliability

The impact of sample type and quality on detection sensitivity represents a fundamental yet often underappreciated source of methodological variation in cHL NGS studies. The superior molecular analysis outcomes achieved with fresh-frozen specimens compared to FFPE samples [12,14,15,17] create a systematic bias in study populations, as institutions with different specimen handling protocols may generate inherently different genomic profiles from comparable patient populations. The median mutation count of 244 somatic mutations per case achieved in optimally preserved samples [12] versus the reduced sensitivity observed in degraded FFPE specimens illustrates how preanalytic factors can fundamentally alter biological conclusions.

The emergence of liquid biopsy approaches using circulating tumor DNA, with detection rates ranging from 68-81% across different cohorts [14,15,16], introduces additional methodological complexity. While ctDNA analysis offers promising non-invasive profiling capabilities with median variant allele frequencies of 4.2% in some studies [15], the technical requirements for successful ctDNA detection may vary substantially across different patient populations and disease stages. The 75% detection rate achieved in pediatric cohorts [16] versus other reported frequencies suggests that patient demographics and disease characteristics may influence ctDNA-based genomic profiling success, adding another layer of methodological consideration.

Implications for Precision Medicine and Clinical Translation

The methodological heterogeneity documented in this review has profound implications for the clinical translation of cHL genomic findings. The inconsistent mutation frequencies across studies effectively preclude the establishment of evidence-based genomic biomarkers for risk stratification or treatment selection. Given that 15-20% of cHL patients experience treatment failure or relapse [6,18], the inability to reliably identify high-risk genomic profiles represents a critical gap in precision medicine implementation.

The technical standardization achieved in other mature lymphoid malignancies through consensus initiatives [2] provides a roadmap for addressing the methodological challenges identified in cHL research. The establishment of minimal gene panels, harmonized quality control measures, and standardized analytical pipelines represents essential prerequisites for advancing cHL genomic research from descriptive studies to clinically actionable findings.

Toward Methodological Harmonization

The evidence presented in this review supports the immediate implementation of standardized NGS approaches for cHL research through several critical initiatives. First, the establishment of consensus tumor enrichment protocols that balance technical feasibility with analytical sensitivity is essential for ensuring comparable tumor cell purity across studies. Second, the development of harmonized NGS panel compositions and analytical pipelines, incorporating lessons learned from mature lymphoid malignancy consensus efforts [2], would enable meaningful cross-study comparisons and facilitate multicenter collaborations.

Third, the implementation of standardized preanalytic procedures and quality control measures for different sample types including fresh-frozen tissue, FFPE specimens, and liquid biopsies would address the fundamental variability in detection sensitivity observed across studies. Finally, the establishment of reference standards and proficiency testing programs would ensure consistent analytical performance across different laboratories and platforms.

Study Limitations and Future Directions

While this systematic review comprehensively documents the methodological heterogeneity in cHL NGS studies, certain limitations should be acknowledged. The rapid evolution of NGS technologies means that some methodological challenges identified in earlier studies may have been addressed by more recent technical developments. Additionally, the focus on methodological standardization should not preclude innovative approaches that may ultimately prove superior to current consensus methods.

Future research should prioritize the development and validation of standardized NGS protocols specifically designed for cHL analysis, incorporating the unique challenges of HRS cell scarcity and complex tumor microenvironment. Prospective multicenter studies utilizing harmonized methodologies would provide the evidence base necessary for establishing reliable genomic biomarkers and advancing precision medicine approaches in cHL management.
