## Supplementary material for "In Search of Ethical Procedures for LLM-Assisted Systematic Review Production: A Proof-of-Concept Evaluation of Selected Review Components": source_data: Paul Bunyan Review - Fully Auto Used in Survey.pdf

2. Liang WS, Vergilio JA, Salhia B, Huang HJ, Oki Y, Garrido-Laguna I, et al.. Com-

prehensive Genomic Profiling of Hodgkin Lymphoma Reveals Recurrently Mutated Genes and Increased Mutation Burden.. The oncologist. 2019;24(2):219-228. doi: 10.1634/theoncologist.2018-0058.

nal.pone.0153844.
