## Supplementary material for "In Search of Ethical Procedures for LLM-Assisted Systematic Review Production: A Proof-of-Concept Evaluation of Selected Review Components": source_data: class_file.py

```
import main
import pandas as pd

class Source:

	def __init__(self, title = None, number = None, abstract = None, pmid = None, pmc_id = None, citation = None, doi = None, year = None, journal = None, full_text = None, cited_by = None, summary = None, rating = None, rate_explain = None, section_rate = None):
		self._title = title #A sources title
		self._number = number #A sources 'number' within the set being evaluated, just for organization.
		self._abstract = abstract #The abstract
		self._pmid = pmid
		self._pmc_id = pmc_id
		self._citation = citation #The citation of the source
		self._doi = doi
		self._year = year #Year of publication
		self._journal = journal #Journal of publication
		self._full_text = full_text #A sources complete text
		self._cited_by = cited_by
		self._summary = summary #A summary of the source, as generated by Claude
		self._rating = rating #A True/False rating of whether a source ought to be included at all for systematic review, as determined by Claude
		self._rate_explain = rate_explain #An explanation (reason of exclusion or inclusion) of the above, by Claude
		self._section_rate = section_rate #Pertains specifically to the Review object. A list where each index corresponds to a step in the review, and how useful Claude thinks this paper would be for writing that section. Needless to say, low scored elements are straight up ignored for writing a section.

	#getters/setters for attributes:
	@property
	def title(self):
		return self._title

	@title.setter
	def title(self, value):
		self._title = value

	@property
	def number(self):
		return self._number

	@number.setter
	def number(self, value):
		self._number = value

	@property
	def abstract(self):
		return self._abstract

	@abstract.setter
	def abstract(self, value):
		self._abstract = value

	@property
	def pmid(self):
		return self._pmid

	@pmid.setter
	def pmid(self, value):
		self._pmid = value

	@property
	def pmc_id(self):
		return self._pmc_id

	@pmc_id.setter
	def pmc_id(self, value):
		self._pmc_id = value

	@property
	def citation(self):
		return self._citation

	@citation.setter
	def citation(self, value):
		self._citation = value

	@property
	def doi(self):
		return self._doi

	@doi.setter
	def doi(self, value):
		self._doi = value

	@property
	def year(self):
		return self._year

	@year.setter
	def year(self, value):
		self._year = value

	@property
	def journal(self):
		return self._journal

	@journal.setter
	def journal(self, value):
		self._journal = value

	@property
	def full_text(self):
		return self._full_text

	@full_text.setter
	def full_text(self, value):
		self._full_text = value

	@property
	def cited_by(self):
		return self._cited_by

	@cited_by.setter
	def cited_by(self, value):
		self._cited_by = value

	@property
	def summary(self):
		return self._summary

	@summary.setter
	def summary(self, value):
		self._summary = value

	@property
	def rating(self):
		return self._rating

	@rating.setter
	def rating(self, value):
		self._rating = value

	@property
	def rate_explain(self):
		return self._rate_explain

	@rate_explain.setter
	def rate_explain(self, value):
		self._rate_explain = value

	@property
	def section_rate(self):
		return self._section_rate

	@section_rate.setter
	def section_rate(self, value):
		self._section_rate = value

	def build_abtext(self):

		if self._abstract is None and self._full_text is not None:
			return self._full_text
		elif self._abstract is not None and self._full_text is None:
			return self._abstract
		elif self._abstract is not None and self._full_text is not None:
			return f'Abstract: {self._abstract}\nFull Text: {self._full_text}'
		else:
			return None

	def build_abtitle(self):
		if self._abstract is None and self._title is not None:
			return f'Title: {self._title}'
		elif self._abstract is not None and self._title is None:
			return f'Abstract: {self._abstract}'
		elif self._abstract is not None and self._title is not None:
			return f'Title: {self._title}\nAbstract: {self._abstract}'
		else:
			return None

class Results_section:

	def __init__(self, section_number = None, section = None, text = None):

		self._section_number = section_number
		self._section = section
		self._text = text

	@property
	def section_number(self):
		return self._section_number

	@section_number.setter
	def section_number(self, value):
		self._section_number = value

	@property
	def section(self):
		return self._section

	@section.setter
	def section(self, value):
		self._section = value

	@property
	def text(self):
		return self._text

	@text.setter
	def text(self, value):
		self._text = value
	


class Review:

	def __init__(self, abstract = None, intro = None, methods = None, results = None, discussion = None, conclusion = None, citations = None, ordinal = []):

		self._abstract = abstract
		self._intro = intro
		self._methods = methods
		self._results = results
		self._discussion = discussion
		self._conclusion = conclusion
		self._citations = citations
		self._ordinal = ordinal

	@property
	def abstract(self):
		return self._abstract

	@abstract.setter
	def abstract(self, value):
		self._abstract = value

	@property
	def intro(self):
		return self._intro

	@intro.setter
	def intro(self, value):
		self._intro = value

	@property
	def methods(self):
		return self._methods

	@methods.setter
	def methods(self, value):
		self._methods = value
	
	@property
	def results(self):
		return self._results

	@results.setter
	def results(self, value):
		self._results = value


	@property
	def discussion(self):
		return self._discussion

	@discussion.setter
	def discussion(self, value):
		self._discussion = value

	@property
	def conclusion(self):
		return self._conclusion

	@conclusion.setter
	def conclusion(self, value):
		self._conclusion = value

	@property
	def citations(self):
		return self._citations

	@citations.setter
	def citations(self, value):
		self._citations = value

	@property
	def ordinal(self):
		return self._ordinal

	@ordinal.setter
	def ordinal(self, value):
		self._ordinal = value


	def format_results_for_save(self):

		output = {}

		for topic in self._results:

			output[topic.section_number] = [topic.section, topic.text]

		return output

	def save_review(self, filename):

		review = {}

		if self._abstract is not None:

			review['abstract'] = self._abstract

		if self._intro is not None:

			review['intro'] = self._intro

		if self._methods is not None:

			review['methods'] = self._methods

		if self._results is not None:

			if type(self._results) is str:

				review['results'] = self._results

			else:

				results = self.format_results_for_save()

				review['results'] = results

		if self._discussion is not None:

			review['discussion'] = self._discussion

		if self._conclusion is not None:

			review['conclusion'] = self._conclusion

		if self._citations is not None:

			review['citations'] = self._citations

		if self._ordinal is not []:

			review['ordinal'] = self._ordinal

		main.save_dict_to_json(review, filename)

	def review_to_tex(self, title = 'Title'):
		review = {}

		if self._abstract is not None:

			review['abstract'] = self._abstract

		if self._intro is not None:

			review['intro'] = self._intro

		if self._methods is not None:

			review['methods'] = self._methods

		if self._results is not None:

			if type(self._results) is str:

				review['results'] = self._results

			else:

				results = self.format_results_for_save()

				review['results'] = results

		if self._discussion is not None:
			review['discussion'] = self._discussion

		if self._conclusion is not None:

			review['conclusion'] = self._conclusion

		if self._citations is not None:

			review['citations'] = self._citations

		main.json_to_latex(review, title = title)


	def format_results_for_load(self, my_dict):

		results = []

		for i in range(len(my_dict)):

			section = Results_section()

			section.section_number = i

			try:
				contents = my_dict[i]
			except:
				contents = my_dict[f'{i}']

			section.section = contents[0]

			section.text = contents[1]

			results.append(section)

		return(results)


	def load_review(self, filename):

		my_dict = main.load_json_to_dict(filename)

		if 'abstract' in my_dict:

			self._abstract = my_dict['abstract']

		if 'intro' in my_dict:

			self._intro = my_dict['intro']

		if 'methods' in my_dict:

			self._methods = my_dict['methods']

		if 'results' in my_dict:

			self._results = my_dict['results']

			if type(self._results) is not str:

				self._results = self.format_results_for_load(self._results)

		if 'discussion' in my_dict:

			self._discussion = my_dict['discussion']

		if 'conclusion' in my_dict:

			self._conclusion = my_dict['conclusion']

		if 'citations' in my_dict:

			self._citations = my_dict['citations']

		if 'ordinal' in my_dict:

			self._ordinal = my_dict['ordinal']

	def build_review_string(self):

		review_string = ''

		if self._abstract is not None:

			review_string = review_string + f"ABSTRACT:\n" + self._abstract

		if self._intro is not None:

			review_string = review_string + f"\nINTRO:\n" + self._intro


		if self._methods is not None:

			review_string = review_string + f"\nMETHODS:\n" + self._methods

		if self._results is not None:

			if type(self._results) == str:

				review_string = review_string + f"\nresults:\n" + self._results

			else:

				for section in self._results:

					if section.text is not None:

						review_string = review_string + f"\nresults SECTION {section.section_number} - {section.section}:\n" + section.text

		if self._discussion is not None:

			review_string = review_string + f"\nDISCUSSION:\n" + self._discussion


		if self._conclusion is not None:

			review_string = review_string + f"\nCONCLUSION:\n" + self._conclusion

		return review_string

	def uncork(self):

		if self._intro is not None:

			self._intro = main.separate_bracketed_lists(str(self._intro))


		if self._results is not None:

			if type(self._results) is str:

				self._results = main.separate_bracketed_lists(str(self._results))

			else:
				for section in self._results:
					section.text = main.separate_bracketed_lists(str(section.text))

		if self._discussion is not None:

			self._discussion = main.separate_bracketed_lists(str(self._discussion))

		if self._conclusion is not None:

			self._conclusion = main.separate_bracketed_lists(str(self._conclusion))


	def replace_cits(self, order):

		def replace_targ(some_string, target1, replace2):

			return some_string.replace(target1, replace2)

		def adjust_for_sources(some_string, order):
			output = some_string
			for i in range(len(order)-1, -1, -1):
				num = order[i]
				output = replace_targ(output, f'[{num}]', f'[{i + 1},]')
			for i in range(len(order)-1, -1, -1):
				num = order[i]
				output = replace_targ(output, f'[{i + 1},]', f'[{i + 1}]')
			return output

		if self._intro is not None:


			self._intro = adjust_for_sources(str(self._intro), order)


		if self._results is not None:

			if type(self._results) is str:

				self._results = adjust_for_sources(str(self._results), order)

			else:
				for section in self._results:
					section.text = adjust_for_sources(str(section.text), order)

		if self._discussion is not None:

			self._discussion = adjust_for_sources(str(self._discussion), order)

		if self._conclusion is not None:

			self._conclusion = adjust_for_sources(str(self._conclusion), order)

	def recork(self):

		if self._intro is not None:


			self._intro = main.combine_adjacent_brackets(str(self._intro))


		if self._results is not None:

			if type(self._results) is str:

				self._results = main.combine_adjacent_brackets(str(self._results))

			else:
				for section in self._results:
					section.text = main.combine_adjacent_brackets(str(section.text))

		if self._discussion is not None:

			self._discussion = main.combine_adjacent_brackets(str(self._discussion))

		if self._conclusion is not None:

			self._conclusion = main.combine_adjacent_brackets(str(self._conclusion))

	def build_from_scratch(self):


		#Should be refactored:
		while True:
			print("Input intro (type 'N' to skip'): ")

			user_response = input("Intro?: ").replace('\\n', '\n')

			if user_response == 'N':
				break
			else:
				self._intro = user_response
				self.save_review('scratch_review.json')
				self.review_to_tex()
				break

		while True:
			print("Input methods (type 'N' to skip'): ")

			user_response = input("Methods?: ").replace('\\n', '\n')

			if user_response == 'N':
				break
			else:
				self._methods = user_response
				self.save_review('scratch_review.json')
				break


		self._results = []

		i = 0

		while True:

			results_section = Results_section()
			print("Input results section header (type 'N' to skip'): ")

			user_response = input("results section header?: ").replace('\\n', '\n')


			if user_response == 'N':
				break
			else:
				results_section.section = user_response
				self.save_review('scratch_review.json')
				results_section.section_number = i
				i += 1
				
				while True:
					print("Input results section body text? (Type 'N' to skip'): ")
					user_response = input("results section text?: ").replace('\\n', '\n')

					if user_response == 'N':
						self._results.append(results_section)
						break
					else:
						results_section.text = user_response
						self._results.append(results_section)
						self.save_review('scratch_review.json')
						break


		while True:
			print("Input discussion (type 'N' to skip'): ")

			user_response = input("Discussion?: ").replace('\\n', '\n')

			if user_response == 'N':
				break
			else:
				self._discussion = user_response
				self.save_review('scratch_review.json')
				break


		while True:
			print("Input conclusion (type 'N' to skip'): ")

			user_response = input("Conclusion?: ").replace('\\n', '\n')

			if user_response == 'N':
				break
			else:
				self._conclusion = user_response
				self.save_review('scratch_review.json')
				break

		while True:
			print("Input abstract (type 'N' to skip'): ")

			user_response = input("Abstract?: ").replace('\\n', '\n')

			if user_response == 'N':
				break
			else:
				self._abstract = user_response
				self.save_review('scratch_review.json')
				break


		while True:
			print("Input citation (type 'N' to skip'): ")
			print("Note, this refers to citations as a bulk string. If you need the citations to be rearranged by number, you will have to sort them prior to this into a spreadsheet and open them as a list of Source objects, then run the build_citations() method")

			user_response = input("Citations?: ").replace('\\n', '\n')

			if user_response == 'N':
				break
			else:
				self._citation = user_response
				self.save_review('scratch_review.json')
				break

		self.save_review('scratch_review.json')
```
