## Supplementary material for "In Search of Ethical Procedures for LLM-Assisted Systematic Review Production: A Proof-of-Concept Evaluation of Selected Review Components": source_data: main.py

```
import requests
import xml.etree.ElementTree as ET
import json
from urllib.parse import quote_plus
import pandas as pd
import time
import random
import re
import class_file as cf
import io
from collections import OrderedDict
import unicodedata
import sourcing
import ast


def sanitize_for_latex(text):
    """
    Helper function to make a string block compatible with latex.

    Args:
    text (string): A string block to be sanitized for latex.

    Returns: 
    string: A string with characters replaced with latex compatible versions.
    """

    # Define replacements for special LaTeX characters
    replacements = [
        ('\\', r'\textbackslash{}'),  # Backslash must be first
        ('&', r'\&'),
        ('%', r'\%'),
        ('$', r'\$'),
        ('#', r'\#'),
        ('_', r'\_'),
        ('{', r'\{'),
        ('}', r'\}'),
        ('~', r'\textasciitilde{}'),
        ('^', r'\textasciicircum{}'),
        ('≥', r'$\geq$'),  # Greater than or equal to
        ('≤', r'$\leq$'),  # Less than or equal to
        ('>', r'$>$'),     # Greater than
        ('<', r'$<$'),     # Less than
    ]

    # Function to replace a single character or string if it's not already escaped
    def replace_unescaped(text, char, replacement):
        result = ""
        i = 0
        while i < len(text):
            if text[i:i+len(char)] == char:
                if i == 0 or text[i-1] != '\\':
                    result += replacement
                    i += len(char)
                else:
                    result += char
                    i += len(char)
            else:
                result += text[i]
                i += 1
        return result

    # Function to replace Greek Unicode characters with LaTeX equivalents
    def replace_greek(text):
        result = ""
        for char in text:
            if unicodedata.category(char) == 'Ll' and unicodedata.name(char).startswith('GREEK SMALL LETTER'):
                latex_name = unicodedata.name(char).split()[-1].lower()
                result += f'$\\{latex_name}$'
            elif unicodedata.category(char) == 'Lu' and unicodedata.name(char).startswith('GREEK CAPITAL LETTER'):
                latex_name = unicodedata.name(char).split()[-1].lower()
                result += f'$\\{latex_name.capitalize()}$'
            else:
                result += char
        return result

    # Apply special character replacements
    for char, replacement in replacements:
        text = replace_unescaped(text, char, replacement)

    # Replace Greek characters
    text = replace_greek(text)

    return text


def json_to_latex(json_data, title = 'Title'):
    """
    Function that converts a Review object into a latex file for easy, good-looking formatting.

    Args:
    json_data (Review()): A Review() object.
    title (String): (Optional val = 'Title'): A string that will serve as the title of the Latex document.

    Returns: 
    Nothing, but will try to save a .tex file to the active directory.
    """

    latex_output = []
    
    # Add document class and packages
    latex_output.append("\\documentclass[12pt,a4paper]{article}")
    latex_output.append("\\usepackage[utf8]{inputenc}")
    latex_output.append("\\usepackage{hyperref}")
    latex_output.append("\\usepackage{geometry}")
    latex_output.append("\\geometry{margin=1in}")
    latex_output.append("\\usepackage{microtype}")  # For better text justification
    
    # Begin document
    latex_output.append("\\begin{document}")
    
    # Title and author
    latex_output.append("\\title{" + f"{title}" + "}")
    latex_output.append("\\author{Systematic Review Team}")  # Add an author
    latex_output.append("\\maketitle")
    
    # Abstract
    try:
        latex_output.append("\\section{Abstract}")
        latex_output.append(sanitize_for_latex(json_data["abstract"].replace("\n", "\n\n")))
    except:
        pass

    # Introduction
    try:
        latex_output.append("\\section{Introduction}")
        latex_output.append(sanitize_for_latex(json_data["intro"].replace("\n", "\n\n")))
    except:
        pass
    
    # results
    try:
        latex_output.append("\\section{Results}")
        for key, value in json_data["results"].items():
            subsection_title = value[0]
            subsection_content = value[1]
            
            # Remove numbering from subsection title
            subsection_title = re.sub(r'^\d+\.\s*', '', subsection_title)
            
            latex_output.append(f"\\subsection{{{sanitize_for_latex(subsection_title)}}}")
            latex_output.append(sanitize_for_latex(subsection_content.replace("\n", "\n\n")))
    except:
        pass

    # Discussion
    try:
        latex_output.append("\\section{Discussion}")
        latex_output.append(sanitize_for_latex(json_data["intro"].replace("\n", "\n\n")))
    except:
        pass
    
    # Conclusion
    try:
        latex_output.append("\\section{Conclusion}")
        latex_output.append(sanitize_for_latex(json_data["conclusion"].replace("\n", "\n\n")))
    except:
        pass

    # Citations
    try:
        latex_output.append("\\section{Citations}")
        latex_output.append(sanitize_for_latex(json_data["citations"].replace("\n", "\n\n")))
    except:
        pass
    
    # End document
    latex_output.append("\\end{document}")

    latex_output = "\n\n".join(latex_output)
    
    with io.open('output.tex', 'w', encoding='utf-8') as file:
        file.write(latex_output)

def save_dict_to_json(dictionary, filename):
    """
    Save a dictionary to a JSON file.
    
    Args:
    dictionary (dict): The dictionary to be saved
    filename (str): The name of the file to save the dictionary to (including .json extension)
    
    Returns:
    None
    """
    with open(filename, 'w') as json_file:
        json.dump(dictionary, json_file, indent=4)

def load_json_to_dict(filename):
    """
    Load a JSON file and return its contents as a dictionary.
    
    Args:
    filename (str): The name of the JSON file to load (including .json extension)
    
    Returns:
    dict: The dictionary containing the data from the JSON file
    """
    try:
        with open(filename, 'r') as json_file:
            return json.load(json_file)
    except FileNotFoundError:
        print(f"Error: The file {filename} was not found.")
        return None
    except json.JSONDecodeError:
        print(f"Error: The file {filename} is not a valid JSON file.")
        return None

def extract_bracketed_numbers(text):
    """Helper function that takes a string and returns a list of all integers that exist inside brackets, such as '[5] and [6]'' -> [5,6]"""
    pattern = r'\[(\d+)\]'
    matches = re.findall(pattern, text)
    return [int(match) for match in matches]

def separate_bracketed_lists(text):
    """Help function that takes a string containing bracketed integers seperated by commas and returns the string with those commas split up, for example 'a string with [5,6]' -> 'a string with [5][6]'"""
    def replace_bracket(match):
        content = match.group(1)
        numbers = [num.strip() for num in content.split(',')]
        return ''.join(f'[{num}]' for num in numbers)

    #pattern = r'\[([\d,\s]+)\]' #for brackets
    #pattern = r'\(([\d,\s]+)\)' #for parens
    pattern = r'[\[\(]([\d,\s]+)[\]\)]' #For both
    return re.sub(pattern, replace_bracket, text)

def combine_adjacent_brackets(text):
    """Helper function that reverses the above method, for example 'a string with [5][6]' -> 'a string with [5,6]'"""
    def replace_adjacent_brackets(match):
        # Get all numbers from the match and convert to integers
        numbers = [int(num) for num in match.group(0).replace('][', ' ').strip('[]').split()]
        # Sort the numbers
        numbers.sort()
        # Join them back together with commas
        return f'[{",".join(str(x) for x in numbers)}]'
    
    pattern = r'(?:\[(\d+)\])+' 
    return re.sub(pattern, replace_adjacent_brackets, text)


def query_claude(api_key, prompt, model="claude-sonnet-4-20250514", max_tokens=1000, temperature=0.3, top_p=None, max_retries=5, initial_delay=1):
    """
    Method that queries the Claude LLM via API

    Args:
    api_key (string): An string anthropic api key.
    prompt (string): A string prompt for Claude to respond to.
    model (string): (Optional value - 'claude-3-5-sonnet-20240620') A string model name for which Claude model to use. 
    max_tokens (int): (Optional value - 1000) A int value of the maximum number of tokens that Claude can return.
    temperature (float): (Optional value - 0.3) A 0-1 float vlaue that tells claude how 'creative' to be in its responses, with 1 representing maximum creativity
    top_p (float): (Optional value - None) A similar param to temperature, but typically I recommend keeping this as None because the temperature is more straighforward.
    max_retries (int): (Optional value - 5) An int for the maximum times that Claude can retry responding (with exponential increasing backoff pause time) if the API querying token limit was reached.
    initial_delay (int): (Optional value - 1) The number of seconds to delay the first query retry for Claude to respond if the API querying token limit was reached.

    Returns:
    string: A response from Claude.
    """

    url = "https://api.anthropic.com/v1/messages"
    
    headers = {
        "Content-Type": "application/json",
        "x-api-key": api_key,
        "anthropic-version": "2023-06-01"
    }
    
    data = {
        "model": model,
        "max_tokens": max_tokens,
        "messages": [{"role": "user", "content": prompt}]
    }
    
    if temperature is not None:
        data["temperature"] = temperature
    elif top_p is not None:
        data["top_p"] = top_p
    
    retries = 0
    while retries < max_retries:
        try:
            response = requests.post(url, headers=headers, json=data)
            response.raise_for_status()
            
            result = response.json()
            return result['content'][0]['text']
        
        except requests.exceptions.HTTPError as e:
            if e.response.status_code == 429 or e.response.status_code == 529:  # Too Many Requests
                if retries == max_retries - 1:
                    print("Could not generate (likely due to rate limit)")
                    return f"Max retries exceeded. Last error: {str(e)}"
                
                sleep_time = (2 ** retries) * initial_delay + random.uniform(0, 1)
                print(f"Rate limit exceeded. Retrying in {sleep_time:.2f} seconds...")
                time.sleep(sleep_time)
                retries += 1
            else:
                return f"An HTTP error occurred: {str(e)}"
        
        except requests.RequestException as e:
            return f"An error occurred while querying Claude: {str(e)}"
    
    return "Max retries exceeded without successful response"

def claude_narrow(api_key, prompt):

    """Deprecated"""

    reply = ast.literal_eval(query_claude(api_key, prompt, temperature = 1.0))

    for item in reply:
        item = int(item)

    return reply


def build_premade_sources(file_path):
    """
    Takes a path to an properly formatted excel file and opens its elements into a list of Source() objects.

    Args:
    file_path (string) - A string directory address to a compatible .xlsx file.

    Returns:
    list - A list of Source objects()
    """

    # Read the Excel file
    df = pd.read_excel(file_path)

    column_names = df.columns.tolist()

    sources = []

    x = 0
    for index, row in df.iterrows():
        # Extract values from each column using iloc
        
        new_source = cf.Source(number = row.iloc[0], citation = row.iloc[1], full_text = row.iloc[2], abstract = row.iloc[3], rating = row.iloc[4], rate_explain = row.iloc[5])

        if pd.isna(new_source.citation):
            print(f"Source {new_source.number} has no citation") 
            new_source.citation = None

        if pd.isna(new_source.full_text):
            print(f"Source {new_source.number} has no full text")
            new_source.full_text = None

        if pd.isna(new_source.abstract):
            new_source.abstract = None

        if pd.isna(new_source.rating):
            new_source.rating = None

        if pd.isna(new_source.rate_explain):
            new_source.rate_explain = None

        sources.append(new_source)

    return sources

def summarize_papers(sources, api_key, summary_dict = {}, targets = None):
    """
    Takes a list of Source() objects and summarizes their full texts (if available) using Claude.

    Args:
    sources (List) - A list of Source() objects that ideally have their full_text properties set.
    api_key (String) - A string anthropic api_key to query Claude with.
    summary_dict (Dict) - (Optional val - {}) A dictionary of Source number IDs as keys and string full text summaries as values, for adding more summaries to a partially summarized set.
    targets (List) - (Optional val - None) A list of Source number IDs to summarize, assuming not all of the Sources in the list want to be summarized.

    Returns:
    Dict: A dictionary of Source number IDs as keys and string full text summaries as values.
    """


    if targets is not None:

        for source in sources:
            prompt = f"I am doing a literature analysis. Please produce a detailed bullet point summary of all contents in this paper: {source.build_abtext()}"

            if int(source.number) in targets:
                try:

                    source.summary = query_claude(api_key, prompt, temperature = 0.3, max_tokens = 2000)

                    summary_dict[str(source.number)] = source.summary

                    print(f"Summarized paper {source.number}")

                except:

                    print(f"Summarization failed for some reason on paper {source.number}")

                    summary_dict[source.number] = None

                save_dict_to_json(summary_dict, 'summaries.json')

    else:

        for source in sources:
            prompt = f"I am doing a literature analysis. Please produce a detailed bullet point summary of all contents in this paper: {source.build_abtext()}"

            try:

                source.summary = query_claude(api_key, prompt, temperature = 0.3, max_tokens = 2000)

                print(f"Summarized source {source.number}")

                summary_dict[source.number] = source.summary

            except:

                print(f"Summarization failed for some reason on paper {source.number}")

                summary_dict[source.number] = None

            save_dict_to_json(summary_dict, 'summaries.json')

    return summary_dict

def build_intro(sources, topic, api_key, review):


    source_string = {}

    sources = choose_sources(review, sources, 'intro')
 
    for source in sources:

        source_string[source.number] = f"{source.summary}"

    prompt = f"""You are a physician scientist writing the intro for a systematic review on the topic: {topic}. You must write the intro based only on the information in the following sources. Each source is numerically categorized in a dictionary. 
    When you write the intro, all information you draw from the sources must be referenced by its numerical identifier in your text response. You must write an intro with citations in the style of a systematic review rather than a summary. 
    You must reference the source correctly when you obtain information. 
    All information you draw upon must be from the sources. You must try to cite each unique source at least once. Try to end your intro with a statement about the purpose of this systematic review on {topic}. If you write a good intro, you will receive a large amount of grant funding. Finally, please do not exceed 400 or so words in your response. Here are the sources: {source_string}."""

    intro = query_claude(api_key, prompt, max_tokens = 2000)

    return intro

def build_results_topics(sources, topic, review, api_key):

    source_string = {}

    for i, source in enumerate(sources):

        source_string[source.number] = f"{source.summary}"

        if i > 150:
            break

    prompt = f"You are a physician scientist coming up with subsections topics for the results section for a systematic review on the topic: {topic}. You must come up with the five most relevant subsection headers (to organize the results section) to argue your systematic review based only on the information in the following sources. In your response, begin and end each subsection header with a semicolon character (;), to make them seperable (Never placing a ; within a header). If you do a good job creating subsection headers for the results section, you will receive a large amount of grant funding. Here are the sources: {source_string} "

    results_topics = query_claude(api_key, prompt, max_tokens = 2000)

    return results_topics

def build_results_section(sources, topic, section, api_key, review):

    source_string = {}

    sources = choose_sources(review, sources, section.section)

    for source in sources:

        source_string[source.number] = f"{source.summary}"

    review_string = review.build_review_string()

    prompt = f"""You are a physician scientist writing a subsection for the results for a systematic review on the topic: {topic}. 
    The subsection you are writing is titled: {section.section}. You must write systematic review contents on this subsection of the results based only on the information in the following sources. Each source is numerically categorized in a dictionary. 
    When you write the subsection, all information you draw from the sources must be referenced by its numerical identifier in your text response. For reference, here is the current systematic review that this subsection will be added to: {review_string}. 
    \nEND OF SYSTEMATIC REVIEW DRAFT\nYou must write a results subsection in the style of a systematic review with citations rather than a summary. 
    You must reference the source correctly when you obtain information. All information you draw upon must be from the sources. You must try to reference each source at least one time, unless a source is very irrelevant.
    If you write a good results subsection on {section.section}, you will receive a large amount of grant funding. Finally, please do not exceed 400 or so words in your response. Here are the sources: {source_string}."""

    text = query_claude(api_key, prompt, max_tokens = 2000, initial_delay = 2)

    section.text = text

def rate_sources(sources, results, api_key, topic):

    topics = [f'An introduction about {topic}']

    for thing in results:

        topics.append(thing.section)

    for source in sources:
        summary = source.summary
        journal = source.journal

        prompt = f"""You are a physician scientist trying to rate the relevance of a source for different subsections of a systematic review. You must do as follows:
        The topics of the subsections will be presented to you in a list. (These are part of the results, unless specified as the intro). You must respond with a
        Python style list where you rate the source on a scale of 1-10 on its relevance to include as a source for each section shown in the list. For example, if presented ['Hip Replacement Techniques', 'Patient Outcomes', 'Future Directions'],
        your reply could be [5,9,2]. Relevance is the primary indicator for ranking, but the following qualities may indicate a higher ranking: 1. A robust study design (such as randomized trials and statistical transparency). 2. Comprehensive reporting of abstract, methods, results, conclusions, and supplementary info. 3. Novel contributions to the field. 4. The source is from a prestigious, high impact journal. You must reply with ONLY a Python list of equivalent indices as the presented list, where each the index of each number corresponds to the
        topic being rated in the original list. I require only a list because your reply must be immediately utilized in code. Here is the list of topics: {topics}. Here is the source, published in {journal} to rate for each topic: ({summary})"""
        response = query_claude(api_key, prompt)

        response = ast.literal_eval(response)

        for item in response:
            item = int(item)

        source.section_rate = response

        print(f'Source {source.number} has been rated')

        sourcing.save_relevant_parts(sources)

    return sources

def build_ordinal(review):

    ordinal = {}

    ordinal['intro'] = 0

    for section in review.results:
        ordinal[section.section] = section.section_number + 1 #section.section?

    review.ordinal = ordinal

    return ordinal


def build_results(results_topics):

    def replace_enters(s):
        return s.replace('\n', '').replace('\r', '')

    def split_string(s):
        if ';' in s:
            # Split the string by semicolons
            substrings = s.split(';')
            
            # Return the list of substrings
            return substrings
        else:
            return s

    def remove_empties(item):
        for i in range(len(item) - 1, -1, -1):
            if item[i] == '':
                del item[i]
        return item

    results = []

    if type(results_topics) == str:

        results_topics = replace_enters(results_topics)

        results_topics = split_string(results_topics)

        results_topics = remove_empties(results_topics)

    choice = None

    while choice != 'N':
        choice = input(f'\nClaude often leaves headers in the first results topic block. This is the output of the results sections: \n{results_topics}\n Type a number to remove that cell in the list (0 refers to the first element). (Type "N" to proceed if list is fine): ')
        if choice != 'N':
            try:
                choice = int(choice)
                try:
                    del results_topics[choice]
                except:
                    print("Invalid list index")
            except:
                print("Please enter an integer")


    for i, topic in enumerate(results_topics):

        section = cf.Results_section()

        section.section_number = int(i)

        section.section = topic

        results.append(section)

    return results

def build_discussion(topic, api_key, review):

    review_string = review.build_review_string()

    prompt = f"You are a physician scientist writing an discussion for a systematic review on the topic: {topic}. You must write the discussion based only on the information in the paper thus far, namely the findings of the results section. For reference, here is the current systematic review that this abstract will be added to: {review_string}. \nEND OF SYSTEMATIC REVIEW DRAFT\nYou must write the discussion in the style of a systematic review as opposed to summarizing. This means you must use the results to generate a comprehensive thesis on {topic}, which the discussion will focus on. The intro and results section both have in text citations. When writing the discussion section, you must reference any information obtained from previous review text with the same in text citation when you mention it in the discussion. If you write a good discussion, you will receive a large amount of grant funding."

    text = query_claude(api_key, prompt, max_tokens = 5000, temperature = 0.7)

    return text


def build_conclusion(sources, topic, api_key, review):

    source_string = {}

    #for source in sources:

        #source_string[source.number] = f"{source.summary}"

    review_string = review.build_review_string()

    prompt = f"You are a physician scientist writing a conclusion for a systematic review on the topic: {topic}. You must write the conclusion based only on the information in the paper thus far. For reference, here is the current systematic review that this conclusion will be added to: {review_string}. \nEND OF SYSTEMATIC REVIEW DRAFT\nYou must write the conclusion in the style of a systematic review as opposed to summarizing. If you write a good conclusion, you will receive a large amount of grant funding. Finally, please do not exceed 250 or so words in your response."

    text = query_claude(api_key, prompt, max_tokens = 1000)

    return text

def build_abstract(topic, api_key, review):

    review_string = review.build_review_string()

    prompt = f"You are a physician scientist writing an abstract for a systematic review on the topic: {topic}. You must write the abstract based only on the information in the paper thus far. For reference, here is the current systematic review that this abstract will be added to: {review_string}. \nEND OF SYSTEMATIC REVIEW DRAFT\nYou must write the abstract in the style of a systematic review as opposed to summarizing. The abstract must refer only to the components currently present in the systematic review. For example, if the systematic review does not have a methods section, you may not describe the methods section in the abstract. If you write a good abstract, you will receive a large amount of grant funding. Finally, please do not exceed 250 or so words in your response."

    text = query_claude(api_key, prompt, max_tokens = 1000, temperature = 0.3)

    return text

def build_citations(review, sources):

    def build_source_string(sources):

        source_string = ""

        for source in sources:  

            source_string += f"\n{source.number}. {source.citation}"

        return source_string

    def remove_duplicates_preserve_order(sequence):
        return list(OrderedDict.fromkeys(sequence))

    review.uncork()

    review_string = review.build_review_string()

    order = remove_duplicates_preserve_order(extract_bracketed_numbers(review_string))

    output_sources = []
    source_dict = {}
    for source in sources:
        source_dict[source.number] = source.citation

    for i, cit in enumerate(order):
        new_source = cf.Source(number = i + 1)

        old_source = source_dict[cit]
        new_source.citation = old_source

        output_sources.append(new_source)

    source_string = build_source_string(output_sources)

    review.replace_cits(order)

    review.citations = source_string

    review.recork()

    return source_string

def suggest_search_terms(api_key, topic, pub_api):
    """Have Claude suggests the terms to search pubmed with. The suggestion is returned as an editable list until the user wants to proceed"""

    prompt = f"I am preforming a systematic review on {topic} and would like assistance coming up with search terms to use in pubmed search. Please reply with a short set of terms to use with pubmed search that will allow me to comprehensively search the pubmed database for relevant papers. The search terms must be narrowed for relevance using the AND operator. For example, if I am researching 'Triple-Negative Breast Cancer and Predictive Markers of Response to Neoadjuvant Chemotherapy', I would not want to simply search 'Triple-Negative Breast Cancer OR biomarkers, as the latter term is too nonspecific. Please reply with multiple, distinct search terms to use separately, using a single AND operator in each. Please reply with only the search terms, separating each with a semicolon at the start and end."

    def replace_enters(s):
        return s.replace('\n', '').replace('\r', '')

    def split_string(s):
        if ';' in s:
            # Split the string by semicolons
            substrings = s.split(';')
            
            # Return the list of substrings
            return substrings
        else:
            return s

    def remove_empties(item):
        for i in range(len(item) - 1, -1, -1):
            if item[i] == '':
                del item[i]
        return item

    response = query_claude(api_key, prompt, temperature = 0.5)

    response = replace_enters(response)

    response = split_string(response)

    response = remove_empties(response)

    choice = None

    while choice != 'N':
        choice = input(f'\nClaude suggests the following {len(response)} pubmed search terms: \n{response}\n Type a number to remove that cell in the list (0 refers to the first element). Type a string (words) to add a custom search terms (Type "N" to proceed if list is fine): ')
        if choice != 'N':

            try:
                choice = int(choice)
                try:
                    del response[choice]
                except:
                    print("Invalid list index")
            except:
                response.append(choice)

    save_dict_to_json(response, 'search_terms.json')

    return response

def retrieve_sources(api_key, topic, pub_key, scope_key = None, search_terms = None):
    """Using a set of search terms, search pubmed for relevant papers"""

    if search_terms is None:

        search_terms = suggest_search_terms(api_key, topic, pub_key)

    else:

        search_terms = load_json_to_dict(search_terms)


    num = int(input("Return how many articles per search term?: "))

    sources = sourcing.get_sources(pub_key, search_terms, num, scope_api = scope_key)

    print("Saving sources to 'sources.xlsx'")

    sourcing.save_sources_to_excel(sources)

    return sources


def screen_abstracts(api_key, topic, sources, ignore_empty = True):
    """Have Claude look at abstracts to decide if they're relevant"""

    if type(sources) is str:
        sources = build_premade_sources(sources)

    i = len(sources) - 1
    while i >= 0:
        source = sources[i]

        if ignore_empty == True and source.full_text is None:
            del sources[i]
            i -= 1
            continue

        source = source.build_abtitle()
        prompt = f"You are a physician scientist screening paper titles and abstracts for the relevance for inclusion into a systematic review about {topic}. Is the following title/abstract relevant for the review? Please reply with only 'True' if relevant or 'False' if irrelevant in your response. Here is the paper: {source}"
        try:
            response = bool(query_claude(api_key, prompt, temperature = 0.3))
            print(f" {i}: response")
            if not response:
                del sources[i]
                print(f"Deleting source {i}")
        except:
            print(f"Appraisal failed on source {i} because response was ambigious, not bool, source will be removed.")
            del sources[i]

        i -= 1

    sourcing.save_sources_to_excel(sources, filename = 'abtract_screened_sources.xlsx')

    return sources

def screen_full_texts(api_key, topic, sources):
    """Have Claude look at full texts to decide if there is a larger issue than relevance, such as bias, etc, to disclude papers"""

    def string_to_list(string):
        """Converts a string that looks like a list into an actual list."""
        try:
            return ast.literal_eval(string)
        except (ValueError, SyntaxError):
            return None  # Return None if the string is not a valid list representation

    if type(sources) is str:
        sources = build_premade_sources(sources)

    removed = []

    i = len(sources) - 1

    while i >= 0:
        source = sources[i]
        if source.full_text is not None:
            text = source.build_abtext()
            prompt = f"You are a physician scientist screening full texts for exclusion or inclusion into a systematic review about {topic}. You must exclude a source for the following reasons: 1. It is a systematic review on the same topic, as we do not simply want to copy another review. 2. It is an opinion piece. 3. There is a financial bias or conflict of interest, such as being financed by a company with a vested interest. 4. The sample size is concerningly low for this type of experimental study (note different studies may expect different sample sizes). 5. It is not relevant to the topic. If it is a primary study, it is more favorable. Your response must be formatted like a Python list, containing a boolean to include or exclude in the first index, and a string explanation in the second, as follows: if including source, response: [True, 'explanation']\n. If excluding source, response: '[False, 'explanation']\n. Here is the paper to evaluate: {text}"            
            response = string_to_list(query_claude(api_key, prompt, temperature = 0.5))
            try:
                inc = bool(response[0])
                exp = str(response[1])
                print(f"Source {i} inclusion appraisal: {inc}; {exp}")
                source.rating = inc
                source.rate_explain = exp
                if not inc:
                    removed.append(sources[i])
                    del sources[i]
            except:
                print(f"Could not appraise source {i} because response was ambigious, not boolean: {response}")
                source.rating = False
                source.rate_explain = f"Could not appraise source {i} because response was ambigious, not boolean: {response}"
                removed.append(sources[i])
                del sources[i]

            i -= 1
        else:
            removed.append(sources[i])
            del sources[i]
            i -= 1

            print(f"Deleting source {i} because no text")

    sourcing.save_sources_to_excel(removed, filename = 'removed_sources.xlsx')
    sourcing.save_sources_to_excel(sources, filename = 'text_screened_sources.xlsx')

    return sources


def create_review(api_key, topic, sources = None, summaries = None, pub_key = None, scope_key = None, screen_abstracts = False, review_topics = None):

    my_review = cf.Review()


    if sources is None and pub_key is not None: #Block to search pubmed for sources

        if summaries is None:

            sources = retrieve_sources(api_key, topic, pub_key, scope_key = scope_key)
            if screen_abstracts:
                sources = screen_abstracts(api_key, topic, sources)
            sources = screen_full_texts(api_key, topic, sources)

            summaries = summarize_papers(sources, api_key)
        else:
            summaries = load_json_to_dict(summaries) #Summarize the sources for digestibility

            for source in sources:
                try:

                    source.summary = summaries[str(source.number)]
                except:
                    pass

        my_review.results = build_results_topics(sources, topic, my_review, api_key) #Pick results topics based on the sources
        my_review.results = build_results(my_review.results)
        my_review.save_review('review_results_topics.json')

        rate_sources(sources, my_review.results, api_key, topic) #Rate sources to control which flow into the review parts - stops Claude from hallucinating


    elif bool(re.search(r'\.xlsx$', sources)) and review_topics is None: #Excel file sources presumes the sources have just recently been built with some manual assistance
        sources = build_premade_sources(sources)

        if summaries is None: #So we have to do all the source appraisal
            summaries = summarize_papers(sources, api_key)
            save_dict_to_json(summaries, 'summaries.json')
        else:
            summaries = load_json_to_dict(summaries) #Summarize the sources for digestibility

            for source in sources:
                try:

                    source.summary = summaries[str(source.number)]
                except:
                    pass

        my_review.results = build_results_topics(sources, topic, my_review, api_key) #Pick results topics based on the sources
        my_review.results = build_results(my_review.results)
        my_review.save_review('review_results_topics.json')

        rate_sources(sources, my_review.results, api_key, topic) #Rate sources to control which flow into the review parts - stops Claude from hallucinating

    elif review_topics is None:
        print("Please use correct params")
        exit()
    else: #presumes we have rated the source topics already and want to start from there
        my_review = cf.Review()
        my_review.load_review(review_topics)
        summaries = load_json_to_dict(summaries)
        sources = build_premade_sources(sources)
        for source in sources:
            source.section_rate = summaries[str(source.number)][1]
            source.summary = summaries[str(source.number)][0]

    build_ordinal(my_review) #Keeps track of the order of the things

    my_review.intro = build_intro(sources, topic, api_key, my_review) #Now we can write the intro
    my_review.save_review('review_intro.json')

    for section in my_review.results:

        build_results_section(sources, topic, section, api_key, my_review)
        my_review.save_review(f'review_results_section_{section.section_number}.json')

    my_review.discussion = build_discussion(topic, api_key, my_review)

    my_review.save_review('review_discussion.json')


    my_review.conclusion = build_conclusion(sources, topic, api_key, my_review)


    my_review.save_review('review_conclusion.json')


    my_review.abstract = build_abstract(topic, api_key, my_review)

    my_review.save_review('review_abstract.json')

    #my_review.methods = "PUT METHODS TEXT HERE"

    #my_review = my_review.save_review('review_methods.json')

    build_citations(my_review, sources)

    my_review.save_review('review_final.json')

    my_review.review_to_tex()

    return my_review


def choose_sources(review, sources, section):

    def get_top_indices(numbers, proportion = 10):
        """
        Returns indices of the highest values in the list based on the given proportion.
        
        Args:
            numbers (list): List of numeric values
            proportion (float): Proportion of top values to return (between 0 and 1)
        
        Returns:
            list: Indices of the top values in ascending order
        """
        
        if proportion < 1:
            # Calculate how many indices to return
            n_indices = max(1, int(len(numbers) * proportion))
        else:
            n_indices = proportion
        
        # Create list of tuples with (value, index)
        indexed_numbers = list(enumerate(numbers))
        
        # Sort by value in descending order
        indexed_numbers.sort(key=lambda x: x[1], reverse=True)
        
        # Get the indices of the top n values and sort them
        top_indices = [idx for idx, _ in indexed_numbers[:n_indices]]
        top_indices.sort()

        return top_indices

    targ_num = review.ordinal[section] #What number index from source.section_rate to get

    ratings = [] #Init a list to hold the ratings for each source relative to the section in question

    return_sources = []

    indexer = {}

    for i, source in enumerate(sources):

        ratings.append(source.section_rate[targ_num])
        indexer[i] = source

    top_sources = get_top_indices(ratings)

    for index in top_sources:

        return_sources.append(indexer[index])

    return return_sources


if __name__ == "__main__":

    api_key = input("Please enter Claude anthropic API key: ")

    topic = input("Please enter topic for systematic review: ")

    pub_api = input("Please enter NCBI API key: ")

    my_review = create_review(api_key, topic, pub_key = pub_api)
```
