## Supplementary material for "In Search of Ethical Procedures for LLM-Assisted Systematic Review Production: A Proof-of-Concept Evaluation of Selected Review Components": source_data: README.md

# automated_systematic_review_claude
Scripts to create an end-to-end systematic review by scraping the NCBI API for sources on a topic, then having Anthropic's Claude Sonnet 4.0 LLM model iteratively query itself to summarize, evaluate and write a complete systematic review on some topic.
The supplementary files folder contains the data/reviews/surveys from the associated study.

These scripts are arranged to utilize the Claude Anthropic API together with the NCBI API to write a sytematic review on some topic with just the push of a button. The outputted review will contain accurate citations to real papers in the topic evaluation. As a disclaimer, these scripts were created as a proof of concept and for the purpose of evaluation of AI-assisted systematic review creation, and not as a means to create actually publishable systematic reviews.

Before running this program, you will need to obtain the following:

1. Download and install python version 3.11+. Make sure it is setup to as an environment variable on your PATH.

2. You will need to install two additional Python packages that are not part of the main library. These are 'pandas' which is a spreadsheet manager and 'requests' which helps handle calls to an external API. Within your Python environment, use the commands 'pip install requests' and 'pip install pandas' to set this up.

3. You will need to obtain an API key for both Anthropic and the NCBI pubmed database.
    - The pubmed API can be set up for free here: https://library.cumc.columbia.edu/kb/getting-started-pubmed-api
    - A Claude API can be set up here: https://www.claude.com/platform/api. Note that you will need to add funds to pay for any API-based queries to Claude, although it's probably only a few dollars or so to write a complete systematic review using this. A larger limitation is that you may face rate daily querying limits with a new account. This may prevent the evaluation of a large amount of sources for systematic review writing, although in my case I was able to get Anthropic to massively increase my API rates by just asking them.

After setting up the above 3 things, you will be able to run the scripts.

These scripts are designed to be run from the command line. To use them, place all three scripts (main.py, sourcing.py, and class_file.py) in some folder/directory and open the windows cmd within that directory. Next, give the command 'python main.py' to run the main code block that executes the pipeline. It will ask you to provide a topic for the systematic review, your Anthropic API, and your NCBI API keys. 

The pipeline works the following way:

  1. You provide a topic for your review.
  2. Claude creates 'Boolean Search Terms' (just search phrases with 'AND' or 'OR' operators) to be used for pubmed search. You as the user will be prompted to add or remove any of these terms as you feel is necessary. 
  3. You provide the number of sources to get for each search term.
  4. The search terms are used to pull papers from the pubmed API.
  5. Claude reads the papers and decides if they are worthy for inclusion in the systematic review. Those showing evidence of bias, competing interests, low sample size, irrelevance, etc, or are systematic reviews themselves tend to get tossed. The exact behavior of this can be tweaked by editing the prompt it its respective section of the 'main.py' file. Any papers that didn't yield a full-text (usually due to being paywalled) will be tossed. Although such papers would be ideal to retrieve, no workaround was set up for this proof of concept.
  6. All included papers are summarized to pull out pertinent findings.
  7. Claude reads the summaries and comes up with a series of subsection topics to address in the 'results' section of the review. You as the user will be prompted to add or remove any of these 'subsections' as you feel necessary -- usually the first one needs to be removed as it will just be Claude talking about what it's doing.
  8. For each summary, Claude rates how relevant it thinks it is for writing the intro and the results subsections.
  9. Next, Claude will write the intro and results subsections. The top 10 summaries for each subsection are pulled when writing that section, to use as references. This text-reduction pipeline was necessary as the only consistent way to get the LLM to accurately cite its sources, as it tends to get confused when reading too much text at once.
  10. Claude will write the discussion section, but will only use the intro and results sections as references when writing this, to ensure the discussion is consistent with the results.
  11. Next, some of the auxiliary sections are written, namely the abstract, conclusion, and references sections. No methods section is produced.
  12. The totality of the review is saved as a Latex file. It can be edited and then converted into a nice-looking pdf with a latex-compiling text editor such as TeXworks.

Some final things to note:
  1. The program will attempt to save a bunch of files to your directory, which are just .json files containing the growing review data, to be used for your reference, as well as some .xlsx microsoft excel spreadsheets, to compile some of the iterative results (such as the summaries). Please make sure your computer will let you save these.
  2. This program does not employ many try/except blocks so it will probably crash if it runs into a problem, which would be related to how you set up the environment to run the program. For example, if it can't save a file due to some permissions reasons, it might just exit the program. If you run out of Claude tokens (hit the API limit for your account), it will sleep for a bit and try again, up to five sleeps, with longer delays between each sleep. If it goes beyond that, it will just skip to the next step of the program and may eventually not work correctly if there are a lot of holes in the responses. You can update the max_retries param in the 'query_claude' function if you want it to sleep for longer but I would instead recommend just remaining aware of your API limits and not letting the program hit them. In my case, I was able to get anthropic to give me a huge API limit by just asking them so I didn't have this issue really.
  3. This program uses the 'Claude Sonnet 4.0' model, mainly because it was the most recent one at the time. You can update this in the 'query_clade' function if you want to use a different model instead. Additionally, many of Claude's responses are dependent on the prompt used. These prompts can be found within the code written out manually as strings, if you would like to try changing them. But note that parts of the pipeline expect Claude to respond in a certain format, and that removing the request for response formatting will probably break the pipeline.
  4. The AI can still make mistakes. Please do not use this pipeline to create systematic reviews and then claim they are your own in an attempt to publish them.
