## Supplementary material for "In Search of Ethical Procedures for LLM-Assisted Systematic Review Production: A Proof-of-Concept Evaluation of Selected Review Components": source_data: sourcing.py

```
import requests
import xml.etree.ElementTree as ET
import json
import main
import class_file as cf
import pandas as pd
from typing import List, Dict, Optional
from time import sleep


# THE FOLLOWING METHODS ARE RELATED TO SEARCHING THE PUBMED API

        
def search_pubmed(query, api_key, retmax=100, sort='relevance', attempt_fulltext=True, umn_credentials=None):
    """
    Search PubMed articles with specified sorting options and optional full-text access.
    
    Args:
        query (str): Search query
        api_key (str): NCBI API key
        retmax (int): Maximum number of results to return
        sort (str): Sort order for results. Options:
            - 'relevance': Sort by relevance score
            - 'pub_date': Sort by publication date (most recent first)
            - 'first_author': Sort by first author
            - 'journal': Sort by journal name
        attempt_fulltext (bool): Whether to attempt full-text retrieval
        umn_credentials (dict): UMN credentials {'username': 'user', 'password': 'pass'}
    
    Returns:
        list: List of dictionaries containing article information
    """
    base_url = "https://eutils.ncbi.nlm.nih.gov/entrez/eutils/"
    
    # Map sort parameter to PubMed's sort field
    sort_map = {
        'relevance': 'relevance',
        'pub_date': 'date',
        'first_author': 'author',
        'journal': 'journal'
    }
    
    # Validate sort parameter
    if sort not in sort_map:
        raise ValueError(f"Invalid sort option. Choose from: {', '.join(sort_map.keys())}")
    
    # Perform the search
    search_url = f"{base_url}esearch.fcgi"
    search_params = {
        "db": "pubmed",
        "term": query,
        "retmax": retmax,
        "usehistory": "y",
        "api_key": api_key,
        "sort": sort_map[sort]
    }
    
    response = requests.get(search_url, params=search_params)
    search_tree = ET.fromstring(response.content)
    
    # Extract WebEnv and QueryKey
    web_env = search_tree.find("WebEnv").text
    query_key = search_tree.find("QueryKey").text
    
    # Fetch the results
    fetch_url = f"{base_url}efetch.fcgi"
    fetch_params = {
        "db": "pubmed",
        "query_key": query_key,
        "WebEnv": web_env,
        "retmax": retmax,
        "retmode": "xml",
        "api_key": api_key
    }
    
    response = requests.get(fetch_url, params=fetch_params)
    fetch_tree = ET.fromstring(response.content)
    
    # Initialize session for UMN access if credentials provided
    umn_session = None
    if attempt_fulltext and umn_credentials:
        umn_session = requests.Session()
        authenticate_umn(umn_session, umn_credentials)
    
    # Parse and return the results
    results = []
    for article in fetch_tree.findall(".//PubmedArticle"):
        title = article.find(".//ArticleTitle").text
        abstract = article.find(".//Abstract/AbstractText")
        abstract_text = abstract.text if abstract is not None else "No abstract available"
        pmid = article.find(".//PMID").text
        
        # Check for PMC ID (for full-text availability)
        pmc_id = article.find(".//ArticleId[@IdType='pmc']")
        pmc_id = pmc_id.text if pmc_id is not None else None
        
        # Get DOI
        doi = article.find(".//ArticleId[@IdType='doi']")
        doi = doi.text if doi is not None else None
        
        # Construct citation
        citation, journal, year = construct_citation(article, doi)
        
        # Initialize article data
        article_data = {
            "title": title,
            "abstract": abstract_text,
            "pmid": pmid,
            "pmc_id": pmc_id,
            "citation": citation,
            "doi": normalize_doi(doi),
            "publication_year": year,
            "journal": journal,
            "full_text": None,
            "full_text_source": None,
            "access_url": None
        }
        
        # Attempt full-text retrieval if requested
        if attempt_fulltext:
            # Try PMC first (free access)
            if pmc_id:
                full_text = get_full_text(pmc_id, api_key)
                if full_text:
                    article_data["full_text"] = full_text
                    article_data["full_text_source"] = "PMC_Free"
                    article_data["access_url"] = f"https://www.ncbi.nlm.nih.gov/pmc/articles/{pmc_id}/"
            
            # If no PMC access and we have UMN credentials, try UMN proxy
            if not article_data["full_text"] and doi and umn_session:
                umn_url, umn_text = try_umn_access(normalize_doi(doi), umn_session)
                if umn_text:
                    article_data["full_text"] = umn_text
                    article_data["full_text_source"] = "UMN_Proxy"
                    article_data["access_url"] = umn_url
                elif umn_url:  # Store URL even if text retrieval failed
                    article_data["access_url"] = umn_url
        
        results.append(article_data)
    
    return results

def get_full_text(pmc_id, api_key):
    """
    Get full text from PMC for a given PMC ID.
    
    Args:
        pmc_id (str): PMC ID (with or without PMC prefix)
        api_key (str): NCBI API key
    
    Returns:
        str or None: Full text content or None if not available
    """
    if not pmc_id:
        return None
        
    # Ensure PMC ID has proper format
    if not pmc_id.startswith('PMC'):
        pmc_id = f'PMC{pmc_id}'
    
    base_url = "https://eutils.ncbi.nlm.nih.gov/entrez/eutils/"
    fetch_url = f"{base_url}efetch.fcgi"
    fetch_params = {
        "db": "pmc",
        "id": pmc_id,
        "retmode": "xml",
        "api_key": api_key
    }
    
    try:
        response = requests.get(fetch_url, params=fetch_params, timeout=10)
        if response.status_code != 200:
            return None
            
        article_tree = ET.fromstring(response.content)
        
        # Extract full text from various sections
        full_text = ""
        
        # Get abstract
        abstract_sections = article_tree.findall(".//abstract//p")
        if abstract_sections:
            full_text += "ABSTRACT:\n"
            for para in abstract_sections:
                if para.text:
                    full_text += para.text + "\n"
            full_text += "\n"
        
        # Get body text
        body_paras = article_tree.findall(".//body//p")
        if body_paras:
            full_text += "MAIN TEXT:\n"
            for para in body_paras:
                if para.text:
                    full_text += para.text + "\n\n"
        
        # If no body paragraphs, try getting all text content
        if not body_paras:
            all_paras = article_tree.findall(".//p")
            for para in all_paras:
                if para.text:
                    full_text += para.text + "\n\n"
        
        return full_text.strip() if full_text.strip() else None
        
    except Exception as e:
        print(f"Error retrieving full text for {pmc_id}: {e}")
        return None

def construct_citation(article, doi):
    # Extract relevant information
    authors = article.findall(".//Author")
    author_list = []
    for author in authors:
        last_name = author.find("LastName")
        initials = author.find("Initials")
        if last_name is not None and initials is not None:
            author_list.append(f"{last_name.text} {initials.text}")
    
    title = article.find(".//ArticleTitle").text
    journal = article.find(".//Journal/Title")
    journal = journal.text if journal is not None else "Unknown Journal"
    year = article.find(".//PubDate/Year")
    year = year.text if year is not None else "Unknown Year"
    volume = article.find(".//Volume")
    volume = volume.text if volume is not None else ""
    issue = article.find(".//Issue")
    issue = issue.text if issue is not None else ""
    pages = article.find(".//MedlinePgn")
    pages = pages.text if pages is not None else ""
    
    # Construct the citation
    if author_list:
        authors_str = ", ".join(author_list[:6])  # Limit to first 6 authors
        if len(author_list) > 6:
            authors_str += ", et al."
    else:
        authors_str = "Unknown Authors"
    
    citation = f"{authors_str}. {title}. {journal}. {year}"
    if volume:
        citation += f";{volume}"
    if issue:
        citation += f"({issue})"
    if pages:
        citation += f":{pages}"
    citation += "."
    if doi is not None:
        citation += f" doi: {doi}."
    
    return citation, journal, year


# THE FOLLOWING METHODS ARE RELATED TO SEARCHING SCOPUS:


def search_scopus(query: str, api_key: str, retmax: int = 25, sort: str = 'relevance') -> List[Dict]:
    """
    Search Scopus articles with specified sorting options.
    
    Args:
        query (str): Search query
        api_key (str): Elsevier API key
        retmax (int): Maximum number of results to return (will paginate if > 25)
        sort (str): Sort order for results. Options:
            - 'relevance': Sort by relevance score
            - 'date': Sort by publication date (most recent first)
            - 'citedby-count': Sort by citation count
    
    Returns:
        list: List of dictionaries containing article information
    """
    base_url = "https://api.elsevier.com/content/search/scopus"
    
    # Map sort parameter to Scopus's sort field
    sort_map = {
        'relevance': 'relevancy',
        'date': 'date',
        'citedby-count': 'citedby-count'
    }
    
    # Validate sort parameter
    if sort not in sort_map:
        raise ValueError(f"Invalid sort option. Choose from: {', '.join(sort_map.keys())}")
    
    # Prepare headers
    headers = {
        "X-ELS-APIKey": api_key,
        "Accept": "application/json"
    }
    
    results = []
    start_index = 0
    page_size = 25  # Maximum allowed per request
    p = 1
    
    while start_index < retmax:
        # Prepare search parameters for this page
        search_params = {
            "query": query,
            "count": min(page_size, retmax - start_index),  # Don't exceed desired total
            "start": start_index,  # Starting position for this page
            "sort": sort_map[sort],
            "field": "title,abstract,doi,author,publicationName,volume,issueIdentifier,pageRange,coverDate,citedby-count"
        }
        
        try:
            response = requests.get(base_url, headers=headers, params=search_params)
            response.raise_for_status()
            data = response.json()
            
            if "service-error" in data:
                raise Exception(f"Scopus API error: {data['service-error']}")
            
            # Process results for this page
            entries = data.get("search-results", {}).get("entry", [])
            if not entries:  # No more results available
                break
                
            for i, entry in enumerate(entries):
                # Extract and format citation information
                citation, journal = construct_scopus_citation(entry)
                
                # Build result dictionary
                result = {
                    "title": entry.get("dc:title", "No title available"),
                    "abstract": entry.get("dc:description", "No abstract available"),
                    "doi": normalize_doi(entry.get("prism:doi")),
                    "scopus_id": entry.get("dc:identifier", "").replace("SCOPUS_ID:", ""),
                    "citation": citation,
                    "cited_by_count": entry.get("citedby-count", "0"),
                    "publication_year": entry.get("prism:coverDate", "")[:4],
                    "journal": journal
                }
                results.append(result)
                print(f'Obtained Scopus source {i} of page {p}')
            
            # Update start index for next page
            start_index += page_size
            
            # Respect rate limits (maximum 6 requests per second)
            sleep(0.2)  # Wait 200ms between requests
            
            # If we've got all requested results, break
            if len(results) >= retmax:
                p += 1
                break
                
        except requests.exceptions.RequestException as e:
            raise Exception(f"Error querying Scopus API: {str(e)}")
    
    # Trim to exact number requested if we got more
    return results[:retmax]

def get_scopus_text(doi: str, api_key: str) -> Optional[str]:
    """
    Attempt to retrieve full text from ScienceDirect if available.
    Note: This will only work for open access articles or if your API key
    has institutional full-text access rights.
    
    Args:
        doi (str): DOI of the article
        api_key (str): Elsevier API key
    
    Returns:
        str or None: Full text if available, None otherwise
    """
    base_url = f"https://api.elsevier.com/content/article/doi/{doi}"
    
    headers = {
        "X-ELS-APIKey": api_key,
        "Accept": "application/json"
    }
    
    try:
        response = requests.get(base_url, headers=headers)
        response.raise_for_status()
        data = response.json()
        
        # Check if full text is available
        if "full-text-retrieval-response" not in data:
            return None
        
        # Extract full text sections
        full_text = ""
        sections = data["full-text-retrieval-response"].get("originalText", "")
        
        return sections if sections else None
        
    except requests.exceptions.RequestException:
        return None

def construct_scopus_citation(entry: Dict) -> str:
    """
    Construct a citation string from Scopus article metadata.
    
    Args:
        entry (dict): Article metadata from Scopus API
    
    Returns:
        str: Formatted citation
    """
    # Extract authors
    authors = entry.get("author", [])
    if isinstance(authors, dict):  # Handle single author case
        author_str = authors.get("authname", "Unknown Author")
    else:  # Handle multiple authors
        author_names = [author.get("authname", "") for author in authors[:6]]
        author_str = ", ".join(filter(None, author_names))
        if len(authors) > 6:
            author_str += ", et al."
    
    # Extract other citation components
    title = entry.get("dc:title", "Unknown Title")
    journal = entry.get("prism:publicationName", "Unknown Journal")
    year = entry.get("prism:coverDate", "")[:4]
    volume = entry.get("prism:volume", "")
    issue = entry.get("prism:issueIdentifier", "")
    pages = entry.get("prism:pageRange", "")
    doi = entry.get("prism:doi", "")
    
    # Construct citation
    citation = f"{author_str}. {title}. {journal}. {year}"
    if volume:
        citation += f";{volume}"
    if issue:
        citation += f"({issue})"
    if pages:
        citation += f":{pages}"
    citation += "."
    if doi:
        citation += f" doi: {doi}."
    
    return citation, journal


# THE FOLLOWING METHODS ARE RELATED TO GENERICALLY OBTAINING/INTERACTING WITH SOURCES

def normalize_doi(doi):
    """
    Normalize DOI by converting to lowercase and handling None values.
    
    Args:
        doi (str or None): DOI string to normalize
    
    Returns:
        str or None: Normalized DOI or None if input was None
    """
    return doi.lower().strip() if doi else None

def get_sources(pub_api, search_query, ret_max = 100, scope_api = None):

    if type(search_query) is str:

        results = search_pubmed(search_query, pub_api, ret_max)

    elif type(search_query) is list:

        results = []

        for query in search_query:
            qresult = search_pubmed(query, pub_api, ret_max)
            results += qresult

    textless = []
    abless = []
    sources = []
    doi_tracking = []

    x = 1
    for i, result in enumerate(results):

        if result['doi'] in doi_tracking:
            continue

        full_text = get_full_text(result['pmc_id'], pub_api)
        new_source = cf.Source(title = result['title'], number = x, abstract = result['abstract'], pmid = result['pmid'], pmc_id = result['pmc_id'], citation = result['citation'], doi = result['doi'], full_text = full_text, journal = result['journal'], year = result['publication_year'])
        sources.append(new_source)
        doi_tracking.append(new_source.doi)
        x += 1
        if new_source.full_text is None:
            textless.append(new_source.number)
        if new_source.abstract == 'No abstract available':
            new_source.abstract = None
            abless.append(new_source.number)

        print(f"Obtained Pubmed Source {i + 1}")

    if scope_api is not None:
        if type(search_query) is str:
            results = search_scopus(search_query, scope_api, retmax = ret_max)
        elif type(search_query) is list:
            results = []

            for query in search_query:
                qresult = search_scopus(query, scope_api, retmax = ret_max)
                results += qresult

        for result in results:
            if result['doi'] in doi_tracking:
                continue
            full_text = get_scopus_text(result['doi'], scope_api)
            new_source = cf.Source(title = result['title'], number = x, abstract = result['abstract'], pmid = result['scopus_id'], citation = result['citation'], cited_by = result['cited_by_count'], doi = result['doi'], full_text = full_text, journal = result['journal'], year = result['publication_year'])
            sources.append(new_source)
            doi_tracking.append(new_source.doi)
            x += 1
            if new_source.full_text is None:
                textless.append(new_source.number)
            if new_source.abstract == 'No abstract available':
                new_source.abstract = None
                abless.append(new_source.number)

    print("(Duplicate sources were excluded)")

    print(f"The following articles are missing full texts: {textless}")
    print(f"The following articles are missing abstracts: {abless}")


    return sources

def save_relevant_parts(sources, filename = 'source_summaries_rated.json'):
    out_dict = {}
    for source in sources:
        out_dict[source.number] = [source.summary, source.section_rate]

    main.save_dict_to_json(out_dict, filename)

def load_relevant_parts(filename):

    sources = []

    my_dict = main.load_json_to_dict(filename)

    for source in my_dict:
        new_source = cf.Source()
        val = my_dict[source]
        new_source.summary = val[0]
        new_source.section_rate = val[1]
        new_source.number = int(source)
        sources.append(new_source)

    return sources

def add_relevant_parts(filename, sources):
    my_dict = main.load_json_to_dict(filename)

    for source in sources:
        targ = my_dict[str(source.number)]
        source.summary = targ[0]
        source.section_rate = targ[1]

    return sources

def save_sources_to_excel(sources, filename='sources.xlsx'):
    """
    Save data from a list of class objects to an Excel file using pandas.
    Each class object should have source_number, citation, and text attributes.
    
    Parameters:
    class_list (list): List of class objects containing the data
    filename (str): Name of the output Excel file (default: 'output.xlsx')
    
    Returns:
    None
    """
    # Initialize lists to store the data
    numbers = []
    citations = []
    texts = []
    abstracts = []
    ratings = []
    explanations = []

    
    # Extract data from each class object
    for source in sources:

        numbers.append(source.number)
        citations.append(source.citation)
        texts.append(source.full_text)
        abstracts.append(source.abstract)
        ratings.append(source.rating)
        explanations.append(source.rate_explain)
 
    
    # Create a dictionary with the data
    data = {
        'Source': numbers,
        'Citation': citations,
        'Text': texts,
        'Abstract': abstracts,
        'Ratings': ratings,
        'Explanations': explanations
    }
    
    # Convert to DataFrame
    df = pd.DataFrame(data)
    
    # Save to Excel
    df.to_excel(filename, index=False)
    
    print(f"Data successfully saved to {filename}")
```
