## Supplementary material for "In Search of Ethical Procedures for LLM-Assisted Systematic Review Production: A Proof-of-Concept Evaluation of Selected Review Components": source_data: readme.txt

python_scripts_for_the_pipeline - This folder contains the original scripts used to generate the fully-AI systematic reviews shown to the survey respondents. These scripts predate ReviewFoundry and are generally meant to be run to automatically generate an AI systematic review with one script execution, without making it easy to break down the subcomponent processes without coding.

review_foundry_source_code - The source code for ReviewFoundry version 1.0.5. The application can be started by running main_app.py in Python.

statistical_testing_code - The code used to run all statistical tests in the paper.

ReviewFoundry_Installer - Windows installer for ReviewFoundry, if you want to run it as a local app.

Supplemental Tables - Contains all the supplemental tables noted in the paper.

The remaining PDFs represent the AI-generated reviews, as described in the pdf's file name, or the survey used.
