## Supplementary material for "In Search of Ethical Procedures for LLM-Assisted Systematic Review Production: A Proof-of-Concept Evaluation of Selected Review Components": source_data: api_manager.py

```
"""
API manager for querying Claude and Ollama LLMs.
Provides a unified interface for both backends.
"""
import requests
import time
import random
import json
from typing import Optional, Dict, Any

from PyQt6.QtCore import QObject, pyqtSignal, QThread
from config_manager import build_claude_payload, extract_text


class LLMWorker(QThread):
    """Background worker for LLM API calls to keep UI responsive."""
    finished = pyqtSignal(str)
    error = pyqtSignal(str)

    def __init__(self, api_manager, prompt: str, params: Dict[str, Any], parent=None):
        super().__init__(parent)
        self.api_manager = api_manager
        self.prompt = prompt
        self.params = params

    def run(self):
        try:
            result = self.api_manager.query(self.prompt, **self.params)
            self.finished.emit(result)
        except Exception as e:
            self.error.emit(str(e))


class APIManager:
    """Manages LLM API connections for Claude and Ollama."""

    def __init__(self, config_manager):
        self.config = config_manager

    def query(self, prompt: str, max_tokens: int = 2000, temperature: float = 0.3,
              top_p: float = None, max_retries: int = 5, initial_delay: int = 1,
              num_ctx: int = 32768, **kwargs) -> str:
        """Route query to the active API backend."""
        active = self.config.active_api
        if active == "claude":
            return self._query_claude(prompt, max_tokens, temperature, top_p,
                                       max_retries, initial_delay)
        elif active == "ollama":
            return self._query_ollama(prompt, max_tokens, temperature, top_p,
                                      max_retries, initial_delay, num_ctx)
        else:
            raise ValueError(f"Unknown API backend: {active}")

    def _query_claude(self, prompt: str, max_tokens: int, temperature: float,
                      top_p: float, max_retries: int, initial_delay: int,
                      model: str = None) -> str:
        api_key = self.config.claude_api_key
        if not api_key:
            raise ValueError(
                "No Claude API key configured. Please set it in Prompt Settings.")
     
        # Caller may override, but default to the configured model so this
        # stays consistent with every other call path.
        model = model or self.config.claude_model
        if not model:
            raise ValueError(
                "No Claude model selected. Open Prompt Settings and choose one.")
     
        url = "https://api.anthropic.com/v1/messages"
        headers = {
            "Content-Type": "application/json",
            "x-api-key": api_key,
            "anthropic-version": "2023-06-01",
        }
     
        # Decides which of temperature / top_p (if any) this model accepts,
        # and clamps max_tokens to its ceiling.
        data = build_claude_payload(model, prompt, max_tokens,
                                    temperature=temperature, top_p=top_p)
     
        retries = 0
        while retries < max_retries:
            try:
                response = requests.post(url, headers=headers, json=data,
                                         timeout=120)
                response.raise_for_status()
                # NOT result['content'][0]['text'] -- on thinking models the
                # first block is a thinking block, so that raises
                # KeyError('text'), which surfaces as the bare message 'text'.
                return extract_text(response.json())
     
            except requests.exceptions.HTTPError as e:
                status = e.response.status_code
                if status in (429, 529):
                    if retries == max_retries - 1:
                        raise RuntimeError(
                            f"Rate limit exceeded after {max_retries} retries.")
                    sleep_time = (2 ** retries) * initial_delay + random.uniform(0, 1)
                    time.sleep(sleep_time)
                    retries += 1
                else:
                    hint = ""
                    if status == 400 and not is_known_model(model):
                        hint = (f"\n\nNote: '{model}' is not in this build's "
                                f"model table, so only generic parameters were "
                                f"sent. It may require parameters this version "
                                f"does not know about.")
                    raise RuntimeError(
                        f"Claude API error: {status} — {e.response.text}{hint}")
     
            except requests.RequestException as e:
                raise RuntimeError(f"Network error querying Claude: {e}")
     
        raise RuntimeError("Max retries exceeded.")


    def _query_ollama(self, prompt: str, max_tokens: int, temperature: float,
                       top_p: float, max_retries: int, initial_delay: int,
                       num_ctx: int) -> str:
        model = self.config.active_ollama_model
        if not model:
            raise ValueError("No Ollama model selected. Please configure in Prompt Settings.")

        url = f"{self.config.ollama_url}/api/chat"
        headers = {"Content-Type": "application/json"}
        data = {
            "model": model,
            "messages": [{"role": "user", "content": prompt}],
            "stream": False,
            "options": {
                "num_predict": max_tokens,
                "num_ctx": num_ctx,
            }
        }
        if temperature is not None:
            data["options"]["temperature"] = temperature
        if top_p is not None:
            data["options"]["top_p"] = top_p

        retries = 0
        while retries < max_retries:
            try:
                response = requests.post(url, headers=headers, json=data, timeout=300)
                response.raise_for_status()
                result = response.json()
                if 'message' in result and 'content' in result['message']:
                    return result['message']['content']
                raise RuntimeError(f"Unexpected Ollama response: {result}")
            except requests.exceptions.ConnectionError:
                if retries == max_retries - 1:
                    raise RuntimeError("Cannot connect to Ollama. Is it running?")
                sleep_time = (2 ** retries) * initial_delay + random.uniform(0, 1)
                time.sleep(sleep_time)
                retries += 1
            except requests.exceptions.HTTPError as e:
                raise RuntimeError(f"Ollama HTTP error: {e}")
            except requests.RequestException as e:
                raise RuntimeError(f"Ollama error: {e}")
        raise RuntimeError("Max retries exceeded.")

    def get_claude_usage_info(self) -> Optional[Dict]:
        """Attempt to get token usage info from Claude API (free endpoint)."""
        # The Anthropic API doesn't have a free usage/limits endpoint currently
        # Returning None indicates this info isn't available
        return None

    def test_connection(self) -> str:
        """Test the active API connection."""
        active = self.config.active_api
        try:
            if active == "claude":
                if not self.config.claude_api_key:
                    return "No Claude API key set."
                result = self.query("Reply with only the word 'connected'.",
                                     max_tokens=10, max_retries=1)
                return f"Claude connected. Response: {result}"
            elif active == "ollama":
                if not self.config.active_ollama_model:
                    return "No Ollama model selected."
                result = self.query("Reply with only the word 'connected'.",
                                     max_tokens=10, max_retries=1)
                return f"Ollama connected. Response: {result}"
        except Exception as e:
            return f"Connection failed: {e}"

    def list_ollama_models(self) -> list:
        """Fetch available models from Ollama."""
        try:
            url = f"{self.config.ollama_url}/api/tags"
            response = requests.get(url, timeout=5)
            response.raise_for_status()
            data = response.json()
            return [m['name'] for m in data.get('models', [])]
        except Exception:
            return []
```
