## Supplementary material for "In Search of Ethical Procedures for LLM-Assisted Systematic Review Production: A Proof-of-Concept Evaluation of Selected Review Components": source_data: citation_manager.py

```
"""
Citation management and validation for the Systematic Review Foundry.
Handles citation parsing, validation (manual, regex-based, AI-based),
and citation reordering for export.
"""
import re
from typing import List, Tuple, Optional, Dict
from collections import OrderedDict
from difflib import SequenceMatcher

from models import Source, ReviewSession


def extract_bracketed_numbers(text: str) -> List[int]:
    """Extract all integers inside brackets: '[5] and [6]' -> [5, 6]"""
    return [int(m) for m in re.findall(r'\[(\d+)\]', text)]


def separate_bracketed_lists(text: str) -> str:
    """Split combined citations: '[5,6]' -> '[5][6]'"""
    def replace_bracket(match):
        numbers = [n.strip() for n in match.group(1).split(',')]
        return ''.join(f'[{n}]' for n in numbers)
    return re.sub(r'[\[\(]([\d,\s]+)[\]\)]', replace_bracket, text)


def combine_adjacent_brackets(text: str) -> str:
    """Combine adjacent citations: '[5][6]' -> '[5,6]'"""
    def replace_adjacent(match):
        numbers = sorted(int(n) for n in match.group(0).replace('][', ' ').strip('[]').split())
        return '[' + ','.join(str(x) for x in numbers) + ']'
    return re.sub(r'(?:\[(\d+)\])+', replace_adjacent, text)


def find_citation_spans(text: str) -> List[dict]:
    """
    Find all citation spans in text. Returns list of dicts with:
    - 'match': the full match string like '[5]' or '[3,7,12]'
    - 'numbers': list of ints
    - 'start': start position in text
    - 'end': end position in text
    """
    results = []
    # Match both [x] and [x,y,z] patterns
    for m in re.finditer(r'\[(\d+(?:\s*,\s*\d+)*)\]', text):
        numbers = [int(n.strip()) for n in m.group(1).split(',')]
        results.append({
            'match': m.group(0),
            'numbers': numbers,
            'start': m.start(),
            'end': m.end(),
        })
    return results


def get_preceding_text(full_text: str, citation_start: int) -> str:
    """
    Get the text that a citation is attributing.
    This is the text after the preceding citation up to this one,
    but not across paragraph breaks.
    """
    # Find the paragraph containing this citation
    para_start = full_text.rfind('\n', 0, citation_start)
    para_start = para_start + 1 if para_start >= 0 else 0

    # Find preceding citation in this paragraph
    preceding_text = full_text[para_start:citation_start]
    prev_cit = list(re.finditer(r'\[\d+(?:\s*,\s*\d+)*\]', preceding_text))
    if prev_cit:
        last = prev_cit[-1]
        return preceding_text[last.end():].strip()
    return preceding_text.strip()


def get_preceding_sentence(full_text: str, citation_start: int) -> str:
    """
    Get only the sentence immediately preceding a citation.
    Walks backward from citation_start to find the nearest sentence
    terminator (. ! ? or a preceding citation bracket), then returns
    everything between that and the citation.
    """
    # Don't cross paragraph breaks
    para_start = full_text.rfind('\n', 0, citation_start)
    para_start = para_start + 1 if para_start >= 0 else 0

    chunk = full_text[para_start:citation_start]

    # Find the last sentence boundary before the citation.
    # A "boundary" is: a period/exclamation/question followed by a space,
    # OR the end of a previous citation bracket ']'.
    best = -1
    # Sentence terminators
    for m in re.finditer(r'[.!?]\s', chunk):
        if m.end() <= len(chunk):
            best = max(best, m.end())
    # Previous citation bracket
    for m in re.finditer(r'\]\s*', chunk):
        # Make sure this is a citation bracket, not random ]
        preceding_bit = chunk[:m.start() + 1]
        if re.search(r'\[\d+(?:\s*,\s*\d+)*\]$', preceding_bit):
            best = max(best, m.end())

    if best >= 0:
        return chunk[best:].strip()
    return chunk.strip()


def find_best_source_match(preceding_text: str, sources: list, mode: int,
                           exclude_numbers: list = None
                           ) -> Optional[Tuple[int, str, float]]:
    """
    Search ALL source summaries to find the best-matching source for
    a piece of attributed text.

    Args:
        preceding_text: the text to match against
        sources: list of source dicts from session.sources
        mode: informs whether to use full texts or summaries, 0 or 1
        exclude_numbers: source numbers to skip (e.g. the current citation)

    Returns:
        (source_number, best_excerpt, score) or None if nothing found
    """
    if not preceding_text:
        return None

    exclude = set(exclude_numbers or [])
    best_overall = None  # (source_number, excerpt, score)

    for s in sources:
        src = s if isinstance(s, dict) else s
        num = src.get('number', 0)
        if num in exclude:
            continue
        if src.get('rating') is False:
            continue
        if mode == 0:
            source_text = (src.get('summary')
                           or src.get('full_text')
                           or src.get('abstract') or '')
        elif mode == 1:
            source_text = (src.get('full_text') or '')

        if not source_text:
            continue
        is_match, excerpt, score = auto_detect_match(
            preceding_text, source_text)
        if best_overall is None or score > best_overall[2]:
            best_overall = (num, excerpt, score)

    return best_overall


def auto_detect_match(preceding_text: str, source_text: str,
                      threshold: float = 0.3) -> Tuple[bool, str, float]:
    """
    Non-AI citation validation using text similarity matching.
    Uses SequenceMatcher to find the best matching passage in the source.
    
    Returns: (is_match, best_match_excerpt, similarity_score)
    """
    if not preceding_text or not source_text:
        return False, "", 0.0

    preceding_lower = preceding_text.lower().strip()
    source_lower = source_text.lower()

    # Try to find matching windows in the source text
    best_score = 0.0
    best_excerpt = ""
    window_size = len(preceding_lower)

    # Use SequenceMatcher for overall similarity
    words_preceding = preceding_lower.split()

    # Slide a window across the source text
    source_words = source_lower.split()
    window = max(len(words_preceding), 10)

    for i in range(max(1, len(source_words) - window + 1)):
        chunk = ' '.join(source_words[i:i + window])
        score = SequenceMatcher(None, preceding_lower, chunk).ratio()
        if score > best_score:
            best_score = score
            # Get the original-case version
            orig_words = source_text.split()
            start = max(0, i - 2)
            end = min(len(orig_words), i + window + 2)
            best_excerpt = ' '.join(orig_words[start:end])

    is_match = best_score >= threshold
    return is_match, best_excerpt, best_score


def reorder_citations_for_export(session: ReviewSession) -> ReviewSession:
    """
    Reorder all citations in the review to be in ascending numerical order
    based on order of first appearance. Updates all section text and
    generates the citations/references list.
    """
    import copy
    export_session = copy.deepcopy(session)

    # Build full review text to find citation order
    all_text = ""
    if export_session.intro:
        all_text += export_session.intro + "\n"
    for rs in export_session.results:
        if rs.get('text'):
            all_text += rs['text'] + "\n"
    if export_session.discussion:
        all_text += export_session.discussion + "\n"
    if export_session.conclusion:
        all_text += export_session.conclusion + "\n"

    # Separate combined citations first
    all_text_sep = separate_bracketed_lists(all_text)
    order = list(OrderedDict.fromkeys(extract_bracketed_numbers(all_text_sep)))

    if not order:
        return export_session

    # Build mapping: old_number -> new_number
    mapping = {old: new + 1 for new, old in enumerate(order)}

    def remap_text(text):
        if not text:
            return text
        text = separate_bracketed_lists(text)
        for old_num in sorted(mapping.keys(), reverse=True):
            text = text.replace(f'[{old_num}]', f'[{mapping[old_num]},]')
        for old_num in sorted(mapping.keys(), reverse=True):
            new_num = mapping[old_num]
            text = text.replace(f'[{new_num},]', f'[{new_num}]')
        text = combine_adjacent_brackets(text)
        return text

    export_session.intro = remap_text(export_session.intro)
    for rs in export_session.results:
        rs['text'] = remap_text(rs.get('text'))
    export_session.discussion = remap_text(export_session.discussion)
    export_session.conclusion = remap_text(export_session.conclusion)

    # Build references list
    source_dict = {}
    for s in export_session.sources:
        src = s if isinstance(s, dict) else s
        source_dict[src.get('number')] = src.get('citation', 'Citation unavailable')

    ref_lines = []
    for old_num in order:
        new_num = mapping[old_num]
        cit = source_dict.get(old_num, f"Source {old_num} — citation not found")
        ref_lines.append(f"{new_num}. {cit}")

    export_session.citations = "\n".join(ref_lines)
    return export_session
```
