## Supplementary material for "In Search of Ethical Procedures for LLM-Assisted Systematic Review Production: A Proof-of-Concept Evaluation of Selected Review Components": source_data: config_manager.py

```
"""
Configuration manager for the Systematic Review Foundry.
Handles user preferences, API keys, custom prompts, and model parameters.

This module is the single source of truth for Claude model discovery:
the /v1/models fetch and the family-grouping helpers live here, and the
UI imports them rather than reimplementing them.
"""
import json
import os
import copy
import urllib.error
import urllib.request
from pathlib import Path
from typing import Optional, Dict, Any, List, Tuple

from default_prompts import DEFAULT_PROMPTS, DEFAULT_MODEL_PARAMS


# ═══════════════════════════════════════════════════════════════════
#  Claude model discovery
# ═══════════════════════════════════════════════════════════════════

ANTHROPIC_MODELS_URL = "https://api.anthropic.com/v1/models"
ANTHROPIC_VERSION = "2023-06-01"

# Family preferred when no model has been chosen yet. This is a hint,
# not a pinned version -- if no model of this family exists the newest
# model overall is used instead, so it can never go stale the way a
# hardcoded model id does.
PREFERRED_FAMILY = "sonnet"


def model_family(model_id: str) -> str:
    """
    Extract the family token ('sonnet', 'opus', 'haiku', ...) from a model
    id without hardcoding a list of families, so new families are picked
    up automatically. Handles both the 'claude-sonnet-4-6' and the older
    'claude-3-5-haiku-20241022' naming schemes.
    """
    for token in model_id.split('-'):
        low = token.lower()
        if low and low != 'claude' and not low.isdigit():
            return low
    return model_id.lower()


def newest_per_family(models: List[Dict[str, str]]) -> List[Dict[str, str]]:
    """
    One model per family. The API lists newest first, so the first id
    seen for a family is that family's current release.
    """
    seen, out = set(), []
    for m in models:
        fam = model_family(m['id'])
        if fam not in seen:
            seen.add(fam)
            out.append(m)
    return out


def normalize_models(raw) -> List[Dict[str, str]]:
    """
    Coerce a cached model list into [{'id', 'display_name'}, ...].
    Tolerates the older cache format, which was a plain list of id
    strings, so an existing config.json keeps working.
    """
    out = []
    for m in raw or []:
        if isinstance(m, str):
            out.append({'id': m, 'display_name': m})
        elif isinstance(m, dict) and m.get('id'):
            out.append({
                'id': m['id'],
                'display_name': m.get('display_name') or m['id'],
            })
    return out


# ═══════════════════════════════════════════════════════════════════
#  Known models and their parameter profiles (as of 2026-08-31)
# ═══════════════════════════════════════════════════════════════════
#
# Anthropic removed the sampling controls (temperature, top_p, top_k)
# starting with Claude Opus 4.7. Affected models return
#     400  `temperature` is deprecated for this model.
# rather than ignoring the value. Support is NOT reported by the
# /v1/models capabilities object, and 'effort' is not a usable proxy
# (Sonnet 4.6 supports effort AND accepts temperature), so it has to be
# recorded here by hand.
#
# 'sampling'   -- False means never send temperature/top_p/top_k.
# 'max_output' -- ceiling for max_tokens; None means do not clamp.
#
# Listed newest first. Update via the Refresh button in Prompt Settings
# when Anthropic ships something newer than this table.

# Date this table was last verified against Anthropic's docs. Shown in
# the startup notice so users can judge how stale it is. Update it when
# you update KNOWN_MODELS.
MODEL_TABLE_DATE = "2026-08-31"

SAMPLING_PARAMS = ("temperature", "top_p", "top_k")

KNOWN_MODELS: List[Dict[str, Any]] = [
    {'id': 'claude-opus-5',              'display_name': 'Claude Opus 5',
     'sampling': False, 'max_output': 128000},
    {'id': 'claude-fable-5',             'display_name': 'Claude Fable 5',
     'sampling': False, 'max_output': 128000},
    {'id': 'claude-sonnet-5',            'display_name': 'Claude Sonnet 5',
     'sampling': False, 'max_output': 128000},
    {'id': 'claude-opus-4-8',            'display_name': 'Claude Opus 4.8',
     'sampling': False, 'max_output': 128000},
    {'id': 'claude-opus-4-7',            'display_name': 'Claude Opus 4.7',
     'sampling': False, 'max_output': None},
    {'id': 'claude-haiku-4-5-20251001',  'display_name': 'Claude Haiku 4.5',
     'sampling': True,  'max_output': 64000},
    {'id': 'claude-opus-4-6',            'display_name': 'Claude Opus 4.6',
     'sampling': True,  'max_output': None},
    {'id': 'claude-sonnet-4-6',          'display_name': 'Claude Sonnet 4.6',
     'sampling': True,  'max_output': None},
]

# Claude Mythos 5 is deliberately absent: it is gated to Anthropic's
# Project Glasswing and would 404 for almost every key. Add it by hand,
# with 'sampling': False, if you have access.

# Used for any model not in the table -- i.e. anything Anthropic ships
# after this date. Sends only the fields that have been required on
# every version of /v1/messages (model, max_tokens, messages) and no
# sampling controls, since the trend is that newer models drop them.
# Worst case the request is plainer than it needed to be; it will not
# 400.
GENERIC_PROFILE: Dict[str, Any] = {'sampling': False, 'max_output': None}


def extract_text(result: Dict[str, Any]) -> str:
    """
    Pull the assistant's text out of a /v1/messages response.

    result['content'] is a LIST OF BLOCKS, and on models with adaptive
    thinking the thinking blocks come FIRST:

        [{"type": "thinking", "thinking": "...", "signature": "..."},
         {"type": "text",     "text": "the actual answer"}]

    So content[0]['text'] raises KeyError('text') -- which prints as just
    'text' -- on Opus 5, Fable 5 and any other thinking model. Thinking
    cannot be turned off on Fable 5, and Opus 5 defaults to effort
    'high', so they hit it nearly every call; Sonnet 4.6 and Haiku 4.5
    send no thinking blocks and appear to work fine.

    Collect every text block instead and ignore the rest.
    """
    blocks = result.get('content') or []
    parts = [b.get('text', '') for b in blocks
             if isinstance(b, dict) and b.get('type') == 'text']
    text = "\n".join(p for p in parts if p).strip()
    if text:
        return text

    # No text came back at all -- say why, rather than KeyError-ing.
    stop = result.get('stop_reason')
    if stop == 'max_tokens':
        raise RuntimeError(
            "The response hit its max_tokens limit before producing any "
            "text. Thinking tokens count toward max_tokens, so a long "
            "thinking pass can consume the whole budget. Raise Max Tokens "
            "for this prompt in Prompt Settings.")
    kinds = sorted({b.get('type') for b in blocks
                    if isinstance(b, dict)}) or ['none']
    raise RuntimeError(
        f"No text content in the Claude response "
        f"(stop_reason={stop!r}, blocks={', '.join(kinds)}).")


def profile_for_model(model_id: str) -> Dict[str, Any]:
    """
    Look up the parameter profile for a model id.

    Falls back through: exact match -> longest prefix match (so dated
    snapshots such as 'claude-sonnet-4-6-20260401' inherit from
    'claude-sonnet-4-6') -> GENERIC_PROFILE for anything unrecognised.
    """
    if not model_id:
        return dict(GENERIC_PROFILE)
    for m in KNOWN_MODELS:
        if m['id'] == model_id:
            return {'sampling': m['sampling'], 'max_output': m['max_output']}
    best = None
    for m in KNOWN_MODELS:
        if model_id.startswith(m['id']) and (
                best is None or len(m['id']) > len(best['id'])):
            best = m
    if best:
        return {'sampling': best['sampling'],
                'max_output': best['max_output']}
    return dict(GENERIC_PROFILE)


def is_known_model(model_id: str) -> bool:
    """False for anything that will fall back to GENERIC_PROFILE."""
    return any(model_id == m['id'] or model_id.startswith(m['id'])
               for m in KNOWN_MODELS)


def default_claude_model() -> str:
    """
    Newest model of PREFERRED_FAMILY in the table, else the newest
    overall. A position in the list, never a literal id, so updating
    KNOWN_MODELS moves the default forward on its own.
    """
    for m in KNOWN_MODELS:
        if model_family(m['id']) == PREFERRED_FAMILY:
            return m['id']
    return KNOWN_MODELS[0]['id'] if KNOWN_MODELS else ""


def build_claude_payload(model: str, prompt: str, max_tokens: int,
                         temperature=None, top_p=None) -> Dict[str, Any]:
    """
    Assemble a /v1/messages body with only the parameters this model
    accepts. Single place where "what can this model take" is decided,
    so the API layer never has to know.
    """
    profile = profile_for_model(model)

    if profile['max_output']:
        max_tokens = min(int(max_tokens), profile['max_output'])

    payload: Dict[str, Any] = {
        'model': model,
        'max_tokens': int(max_tokens),
        'messages': [{'role': 'user', 'content': prompt}],
    }
    if profile['sampling']:
        # At most one sampling control, as Anthropic recommends.
        if temperature is not None:
            payload['temperature'] = temperature
        elif top_p is not None:
            payload['top_p'] = top_p
    return payload


def get_config_dir() -> Path:
    """Return the platform-appropriate config directory."""
    if os.name == 'nt':
        base = Path(os.environ.get('APPDATA', Path.home()))
    elif os.name == 'posix' and 'darwin' in os.uname().sysname.lower():
        base = Path.home() / 'Library' / 'Application Support'
    else:
        base = Path(os.environ.get('XDG_CONFIG_HOME', Path.home() / '.config'))
    config_dir = base / 'SystematicReviewFoundry'
    config_dir.mkdir(parents=True, exist_ok=True)
    return config_dir


def get_default_session_dir() -> Path:
    """Return the default directory for saving sessions."""
    d = Path.home() / 'Documents' / 'SystematicReviewFoundry'
    d.mkdir(parents=True, exist_ok=True)
    return d


class ConfigManager:
    """Manages application configuration, custom prompts, and API settings."""

    def __init__(self):
        self.config_path = get_config_dir() / 'config.json'
        self._config: Dict[str, Any] = {}
        self._load()

    def _default_config(self) -> Dict[str, Any]:
        return {
            "active_session_path": str(get_default_session_dir() / "untitled.json"),
            "active_api": "claude",
            "claude_api_key": "",
            "claude_models": [],
            "claude_model": "",
            # False = "keep me on the newest model automatically".
            # Set True only when the user explicitly picks a model.
            "claude_model_pinned": False,
            "show_model_notice": True,
            "ncbi_api_key": "",
            "ollama_models": [],
            "ollama_url": "http://localhost:11434",
            "active_ollama_model": "",
            "custom_prompts": {},
            "custom_params": {},
            "master_params": {},
            "window_geometry": None,
            "recent_sessions": [],
            "auto_save_interval_seconds": 120,
            "institutional_credentials": {
                "username": "",
                "password": ""
            }
        }

    def _load(self):
        if self.config_path.exists():
            try:
                with open(self.config_path, 'r', encoding='utf-8') as f:
                    saved = json.load(f)
                # Merge with defaults so new keys are always present
                self._config = self._default_config()
                self._config.update(saved)
            except (json.JSONDecodeError, IOError):
                self._config = self._default_config()
        else:
            self._config = self._default_config()

    def save(self):
        with open(self.config_path, 'w', encoding='utf-8') as f:
            json.dump(self._config, f, indent=2, ensure_ascii=False)

    # --- Getters / Setters ---

    @property
    def active_session_path(self) -> str:
        return self._config.get("active_session_path",
                                str(get_default_session_dir() / "untitled.json"))

    @active_session_path.setter
    def active_session_path(self, value: str):
        self._config["active_session_path"] = value
        # Track recent sessions
        recents = self._config.get("recent_sessions", [])
        if value in recents:
            recents.remove(value)
        recents.insert(0, value)
        self._config["recent_sessions"] = recents[:10]
        self.save()

    @property
    def active_api(self) -> str:
        return self._config.get("active_api", "claude")

    @active_api.setter
    def active_api(self, value: str):
        self._config["active_api"] = value
        self.save()

    @property
    def claude_api_key(self) -> str:
        return self._config.get("claude_api_key", "")

    @claude_api_key.setter
    def claude_api_key(self, value: str):
        self._config["claude_api_key"] = value
        self.save()

    @property
    def claude_model(self) -> str:
        """
        The active model. Falls back to the newest preferred entry in
        KNOWN_MODELS rather than a literal id, so the fallback moves
        forward whenever the table is updated.
        """
        return self._config.get("claude_model") or default_claude_model()

    @claude_model.setter
    def claude_model(self, value: str):
        self._config["claude_model"] = value
        self.save()

    @property
    def claude_model_pinned(self) -> bool:
        """
        True when the user deliberately chose a model and it should be
        left alone. False (the default, including for configs written by
        older versions that predate this flag) means the app is free to
        move the selection onto the newest available model.
        """
        return bool(self._config.get("claude_model_pinned", False))

    @claude_model_pinned.setter
    def claude_model_pinned(self, value: bool):
        self._config["claude_model_pinned"] = bool(value)
        self.save()

    @property
    def show_model_notice(self) -> bool:
        """Whether to show the model/parameter notice at startup."""
        return bool(self._config.get("show_model_notice", True))

    @show_model_notice.setter
    def show_model_notice(self, value: bool):
        self._config["show_model_notice"] = bool(value)
        self.save()

    def set_claude_model(self, value: str, pinned: bool = True):
        """Record an explicit model choice. Use this for user selections."""
        self._config["claude_model"] = value
        self._config["claude_model_pinned"] = bool(pinned)
        self.save()

    @property
    def claude_models(self) -> List[Dict[str, str]]:
        """
        Models to offer in the UI: the fetched list if the user has ever
        pressed Refresh, otherwise the hardcoded table. Never empty, so
        the app works offline and on first run.
        """
        fetched = normalize_models(self._config.get("claude_models", []))
        return fetched or normalize_models(KNOWN_MODELS)

    @claude_models.setter
    def claude_models(self, value):
        self._config["claude_models"] = normalize_models(value)
        self.save()

    def fetch_claude_models(self, api_key: Optional[str] = None,
                            limit: int = 100, timeout: float = 10.0
                            ) -> Tuple[List[Dict[str, str]], Optional[str]]:
        """
        Ask the Anthropic API which models the key can actually use.

        GET /v1/models is a metadata lookup, not an inference call, so it
        consumes no tokens and costs nothing. It does require a working
        key, which makes it a reasonable proxy for "is this API usable at
        all" -- there is therefore no hardcoded fallback list. Results
        come back newest-first and that order is preserved.

        Pass api_key to test a key that has not been saved yet; otherwise
        the stored key is used.

        Returns (models, error):
          models  list of {'id', 'display_name'}, newest first
          error   None on success, else a short human-readable message
        """
        key = (self.claude_api_key if api_key is None else api_key).strip()
        if not key:
            return [], "No API key set"

        req = urllib.request.Request(
            f"{ANTHROPIC_MODELS_URL}?limit={int(limit)}",
            headers={
                "x-api-key": key,
                "anthropic-version": ANTHROPIC_VERSION,
            },
            method="GET",
        )
        try:
            with urllib.request.urlopen(req, timeout=timeout) as resp:
                payload = json.load(resp)
        except urllib.error.HTTPError as e:
            if e.code in (401, 403):
                return [], "API key rejected"
            if e.code == 429:
                return [], "Rate limited — try again in a moment"
            return [], f"API error {e.code}"
        except urllib.error.URLError as e:
            return [], f"Could not reach api.anthropic.com ({e.reason})"
        except (json.JSONDecodeError, OSError, ValueError) as e:
            return [], f"Model fetch failed: {e}"

        models = normalize_models(payload.get("data", []))
        if not models:
            return [], "API returned no models"

        self.claude_models = models
        # Move onto the newest model unless the user pinned a choice, and
        # always move if the saved model has been retired. Preference is
        # the newest of PREFERRED_FAMILY, else newest overall -- a family
        # name, never a pinned version that can go out of date.
        available = {m['id'] for m in models}
        if not self.claude_model_pinned or self.claude_model not in available:
            newest = newest_per_family(models)
            pick = next((m['id'] for m in newest
                         if model_family(m['id']) == PREFERRED_FAMILY), None)
            self._config["claude_model"] = pick or models[0]['id']
            self.save()
        return models, None

    @property
    def ncbi_api_key(self) -> str:
        return self._config.get("ncbi_api_key", "")

    @ncbi_api_key.setter
    def ncbi_api_key(self, value: str):
        self._config["ncbi_api_key"] = value
        self.save()

    @property
    def ollama_url(self) -> str:
        return self._config.get("ollama_url", "http://localhost:11434")

    @ollama_url.setter
    def ollama_url(self, value: str):
        self._config["ollama_url"] = value
        self.save()

    @property
    def ollama_models(self) -> list:
        return self._config.get("ollama_models", [])

    @ollama_models.setter
    def ollama_models(self, value: list):
        self._config["ollama_models"] = value
        self.save()

    @property
    def active_ollama_model(self) -> str:
        return self._config.get("active_ollama_model", "")

    @active_ollama_model.setter
    def active_ollama_model(self, value: str):
        self._config["active_ollama_model"] = value
        self.save()

    @property
    def auto_save_interval(self) -> int:
        return self._config.get("auto_save_interval_seconds", 120)

    # --- Prompt Management ---

    def get_prompt(self, prompt_key: str) -> str:
        """Get the active prompt: custom if set, else default."""
        custom = self._config.get("custom_prompts", {}).get(prompt_key)
        if custom:
            return custom
        return DEFAULT_PROMPTS.get(prompt_key, "")

    def set_custom_prompt(self, prompt_key: str, prompt_text: str):
        if "custom_prompts" not in self._config:
            self._config["custom_prompts"] = {}
        self._config["custom_prompts"][prompt_key] = prompt_text
        self.save()

    def reset_prompt(self, prompt_key: str):
        customs = self._config.get("custom_prompts", {})
        if prompt_key in customs:
            del customs[prompt_key]
            self.save()

    def is_prompt_customized(self, prompt_key: str) -> bool:
        return prompt_key in self._config.get("custom_prompts", {})

    def get_all_prompt_keys(self) -> list:
        return list(DEFAULT_PROMPTS.keys())

    # --- Model Parameters ---

    def get_params_for_prompt(self, prompt_key: str) -> Dict[str, Any]:
        """Get parameters for a specific prompt. Falls back to master, then defaults."""
        custom = self._config.get("custom_params", {}).get(prompt_key)
        if custom:
            return custom
        master = self._config.get("master_params", {})
        if master:
            return master
        return copy.deepcopy(DEFAULT_MODEL_PARAMS)

    def set_params_for_prompt(self, prompt_key: str, params: Dict[str, Any]):
        if "custom_params" not in self._config:
            self._config["custom_params"] = {}
        self._config["custom_params"][prompt_key] = params
        self.save()

    def reset_params_for_prompt(self, prompt_key: str):
        customs = self._config.get("custom_params", {})
        if prompt_key in customs:
            del customs[prompt_key]
            self.save()

    def get_master_params(self) -> Dict[str, Any]:
        master = self._config.get("master_params", {})
        if master:
            return master
        return copy.deepcopy(DEFAULT_MODEL_PARAMS)

    def set_master_params(self, params: Dict[str, Any]):
        self._config["master_params"] = params
        self.save()

    def reset_master_params(self):
        self._config["master_params"] = {}
        self.save()

    @property
    def institutional_credentials(self) -> Dict[str, str]:
        return self._config.get("institutional_credentials", {"username": "", "password": ""})

    @institutional_credentials.setter
    def institutional_credentials(self, value: Dict[str, str]):
        self._config["institutional_credentials"] = value
        self.save()
```
