## Supplementary material for "In Search of Ethical Procedures for LLM-Assisted Systematic Review Production: A Proof-of-Concept Evaluation of Selected Review Components": source_data: default_prompts.py

```
"""
Default prompts and model parameters for the Systematic Review Foundry.
All prompts use Python f-string syntax with named variables.
"""

DEFAULT_MODEL_PARAMS = {
    "max_tokens": 4096,
    "temperature": 0.3,
    "top_p": None,
    "max_retries": 5,
    "initial_delay": 1,
    "num_ctx": 32768,  # Ollama context window
}

DEFAULT_PROMPTS = {
    # === SOURCE RETRIEVAL ===
    "suggest_search_terms": (
        "I am performing a systematic review on {topic} and would like assistance "
        "coming up with search terms to use in PubMed search. Please reply with a "
        "short set of terms to use with PubMed search that will allow me to "
        "comprehensively search the PubMed database for relevant papers. The search "
        "terms must be narrowed for relevance using the AND operator. For example, if "
        "I am researching 'Triple-Negative Breast Cancer and Predictive Markers of "
        "Response to Neoadjuvant Chemotherapy', I would not want to simply search "
        "'Triple-Negative Breast Cancer OR biomarkers', as the latter term is too "
        "nonspecific. Please reply with multiple, distinct search terms to use "
        "separately, using a single AND operator in each. Please reply with ONLY the "
        "search terms, separating each with a semicolon at the start and end. Do not "
        "include any other text."
    ),

    # === SOURCE SUMMARIZATION ===
    "summarize_source": (
        "I am doing a literature analysis. Please produce a detailed bullet point "
        "summary of all contents in this paper: {source_text}"
    ),

    # === SOURCE INCLUSION — SUBJECTIVE (default when no criteria) ===
    "screen_source_subjective": (
        "You are a physician scientist screening paper titles and abstracts for "
        "relevance for inclusion into a systematic review about {topic}. Is the "
        "following title/abstract relevant for the review? Reply with ONLY a Python "
        "list: [True, 'explanation'] to include, or [False, 'explanation'] to exclude. "
        "Do not include any other text. Here is the paper: {source_text}"
    ),

    # === SOURCE INCLUSION — CRITERIA-BASED ===
    "screen_source_criteria": (
        "IMPORTANT MESSAGE FROM USER: "
        "Reply with ONLY a Python list: [True, 'explanation'] to include, or "
        "[False, 'explanation'] to exclude. Do not include any other text as I need to immediately use the response in code or it will break. "
        "You are a physician scientist screening papers for inclusion into a "
        "systematic review about {topic}. Evaluate the following paper against "
        "the criteria below.\n\n{criteria}\n\n"
        "For INCLUSION CRITERIA the paper must satisfy ALL listed conditions to "
        "be included. For EXCLUSION CRITERIA, if ANY single condition applies "
        "the paper must be excluded. If only one type of criteria is provided, "
        "evaluate against that type alone.\n\n"
        "IMPORTANT MESSAGE FROM USER: "
        "Reply with ONLY a Python list: [True, 'explanation'] to include, or "
        "[False, 'explanation'] to exclude. Do not include any other text as I need to immediately use the response in code or it will break. "
        "Here is the paper:\n{source_text}"
    ),

    # === SOURCE RATING FOR SECTIONS ===
    "rate_source_sections": (
        "You are a physician scientist trying to rate the relevance of a source for "
        "different subsections of a systematic review. The topics of the subsections "
        "will be presented to you in a list. You must respond with a Python style list "
        "where you rate the source on a scale of 1-10 on its relevance to include as "
        "a source for each section shown in the list. For example, if presented "
        "['Hip Replacement Techniques', 'Patient Outcomes', 'Future Directions'], "
        "your reply could be [5,9,2]. Relevance is the primary indicator for ranking, "
        "but the following qualities may indicate a higher ranking: 1. A robust study "
        "design. 2. Comprehensive reporting. 3. Novel contributions. 4. Prestigious "
        "journal. Reply with ONLY a Python list of equivalent indices. Do not add "
        "any code formatting or other text — just return a string that resembles a "
        "Python list. Here is the list of topics: {topics}. Here is the source, "
        "published in {journal}: ({summary})"
    ),

    # === SOURCE RATING FOR TOPICS ===
    "rate_source_topics": (
        "You are a physician scientist rating the relevance of a source to a specific "
        "research topic. Rate on a scale of 1-10 how relevant this source is to the "
        "topic: '{topic_title}'. Reply with ONLY a single integer (1-10). Do not "
        "include any other text. Here is the source summary: {summary}"
    ),

    # === RESULTS TOPIC GENERATION ===
    "generate_results_topics": (
        "You are a physician scientist coming up with subsection topics for the "
        "results section for a systematic review on the topic: {topic}. You must "
        "come up with the five most relevant subsection headers to organize the "
        "results section based only on the information in the following sources. "
        "{thesis_clause}"
        "In your response, begin and end each subsection header with a semicolon "
        "character (;), to make them separable (never placing a ; within a header). "
        "Do not include any other text. Here are the sources: {source_string}"
    ),

    # === THESIS GENERATION ===
    "generate_thesis": (
        "You are a physician scientist. Based on the following source summaries "
        "about the topic '{topic}', synthesize what the bulk of the literature is "
        "concluding about this topic into a concise thesis statement (2-4 sentences). "
        "This thesis will guide the organization of a systematic review. "
        "Here are the source summaries: {context}"
    ),

    # === SECTION WRITING ===
    "write_intro": (
        "You are a physician scientist writing the intro for a systematic review on "
        "the topic: {topic}. You must write the intro based only on the information "
        "in the following sources. Each source is numerically categorized in a "
        "dictionary. When you write the intro, all information you draw from the "
        "sources must be referenced by its numerical identifier in your text response "
        "using [x] style citations. You must write an intro with citations in the "
        "style of a systematic review rather than a summary. All information you draw "
        "upon must be from the sources. You must try to cite each unique source at "
        "least once. End your intro with a statement about the purpose of this "
        "systematic review on {topic}. Please do not exceed 400 words. "
        "Here are the sources: {context}"
    ),

    "write_results_section": (
        "You are a physician scientist writing a subsection for the results of a "
        "systematic review on the topic: {topic}. The subsection you are writing is "
        "titled: {section_title}. You must write systematic review contents on this "
        "subsection based only on the information in the following sources. Each "
        "source is numerically categorized. When you write the subsection, all "
        "information you draw from the sources must be referenced by its numerical "
        "identifier using [x] style citations. {thesis_clause}"
        "For reference, here is the current review draft: {review_string}\n"
        "END OF SYSTEMATIC REVIEW DRAFT\n"
        "You must write in the style of a systematic review with citations rather "
        "than a summary. Reference each source at least once unless very irrelevant. "
        "Please do not exceed 400 words. Here are the sources: {context}"
    ),

    "write_discussion": (
        "You are a physician scientist writing the discussion for a systematic "
        "review on the topic: {topic}. You must write the discussion based only on "
        "the information in the paper thus far, namely the findings of the results "
        "section. Here is the current systematic review: {review_string}\n"
        "END OF SYSTEMATIC REVIEW DRAFT\n"
        "Write the discussion in the style of a systematic review. Use the results "
        "to generate a comprehensive thesis on {topic}. Reference any information "
        "from previous review text with the same in-text citations. "
        "{context}"
    ),

    "write_conclusion": (
        "You are a physician scientist writing a conclusion for a systematic review "
        "on the topic: {topic}. You must write the conclusion based only on the "
        "information in the paper thus far. Here is the current systematic review: "
        "{review_string}\nEND OF SYSTEMATIC REVIEW DRAFT\n"
        "Write the conclusion in the style of a systematic review. "
        "Please do not exceed 250 words. {context}"
    ),

    "write_abstract": (
        "You are a physician scientist writing an abstract for a systematic review "
        "on the topic: {topic}. Write the abstract based only on the information in "
        "the paper thus far. Here is the current systematic review: {review_string}\n"
        "END OF SYSTEMATIC REVIEW DRAFT\n"
        "Write the abstract in the style of a systematic review. The abstract must "
        "refer only to components currently present in the review. "
        "Please do not exceed 250 words. {context}"
    ),

    "write_methods": (
        "You are a physician scientist writing a methods section for a systematic "
        "review on the topic: {topic}. The review was conducted using the following "
        "approach: Sources were retrieved from the NCBI PubMed database using Boolean "
        "search terms. {methods_notes} Sources were screened for inclusion based on "
        "{screening_approach}. Included sources were summarized and rated for "
        "relevance to each results subsection. The review contains the following "
        "sections: {section_list}. Write a concise methods section in the style of "
        "a systematic review."
    ),

    # === TEXT IMPROVEMENT ===
    "improve_text": (
        "You are a physician scientist editing a systematic review. Please improve "
        "the following text by polishing the language, improving clarity and flow, "
        "while maintaining all in-text citations exactly as they are. Do not add or "
        "remove any citations. Return only the improved text. "
        "Here is the text to improve: {selected_text}"
    ),

    # === TOPIC GENERATION ===
    "generate_topic": (
        "You are a physician scientist. Based on the following sources about "
        "'{topic}', write a detailed evidence-based response to the following "
        "research question: '{topic_title}'. All information must be cited using "
        "[x] style citations referencing the source numbers provided. "
        "Here are the sources: {context}"
    ),

    "auto_generate_topics": (
        "You are a physician scientist working on a systematic review about "
        "'{topic}'. Based on the following information, suggest 5 specific research "
        "questions that would be valuable to investigate as topics within this "
        "review. Reply with ONLY the questions, each separated by a semicolon at "
        "the start and end. {thesis_clause} Here is the context: {context}"
    ),

    # === AI-GENERATED TOPIC / STAT QUESTIONS ===
    "auto_generate_topic_questions": (
        "You are a physician scientist working on a systematic review about "
        "'{topic}'. Based on the following context, generate 8-12 succinct "
        "research questions that would be valuable to investigate as evidence-"
        "gathering topics. Each question should be specific, answerable from "
        "the literature, and relevant to the review's scope. Reply with ONLY "
        "the questions, each separated by a semicolon at the start and end. "
        "Do not include numbering or other text.\n\n{context}"
    ),

    "auto_generate_stat_questions": (
        "You are a physician scientist working on a systematic review about "
        "'{topic}'. Based on the following context, generate 6-10 specific "
        "statistical or numerical questions that could be answered by extracting "
        "data from the literature. Focus on quantifiable outcomes, rates, means, "
        "sample sizes, effect sizes, and comparisons. Each question should be "
        "succinct and answerable with specific numbers. Reply with ONLY the "
        "questions, each separated by a semicolon at the start and end. "
        "Do not include numbering or other text.\n\n{context}"
    ),

    # === BATCHED TOPIC RATING ===
    "rate_source_topics_batch": (
        "You are a physician scientist rating the relevance of a source to "
        "different research topics for a systematic review. The topics will be "
        "presented as a list. You must respond with a Python style list where you "
        "rate the source on a scale of 1-10 for its relevance to each topic. "
        "For example, if presented ['Drug efficacy', 'Side effects', 'Dosing'], "
        "your reply could be [7,3,5]. Reply with ONLY a Python list of integers. "
        "Do not add any code formatting or other text. "
        "Here is the list of topics: {topics}. "
        "Here is the source, published in {journal}: ({summary})"
    ),

    # === BATCHED STAT RATING ===
    "rate_source_stats_batch": (
        "You are a physician scientist rating the relevance of a source to "
        "different statistical questions for a systematic review. The questions "
        "will be presented as a list. You must respond with a Python style list "
        "where you rate the source on a scale of 1-10 for how likely it contains "
        "numerical data or statistics relevant to each question. "
        "For example, if presented ['Mean survival rate', 'Sample sizes'], "
        "your reply could be [8,4]. Reply with ONLY a Python list of integers. "
        "Do not add any code formatting or other text. "
        "Here is the list of questions: {topics}. "
        "Here is the source, published in {journal}: ({summary})"
    ),

    # === STATISTICS (BETA) ===
    "stat_query_text": (
        "You are a physician scientist. Based on the following sources, answer this "
        "statistical/mathematical question: '{question}'. Provide specific numerical "
        "data with citations using [x] style. Here are the sources: {context}"
    ),

    "stat_query_python": (
        "You are a physician scientist. Based on the following sources, answer this "
        "statistical question: '{question}'. Reply with ONLY a Python dictionary "
        "containing the numerical data, suitable for ast.literal_eval(). Do not "
        "include any other text, code formatting, or backticks. "
        "Here are the sources: {context}"
    ),

    # === RISK OF BIAS ASSESSMENT ===
    "risk_of_bias": (
        "You are a physician scientist performing a risk of bias assessment "
        "using the Cochrane Risk of Bias (RoB 2) tool. Assess the following "
        "source for risk of bias across these domains:\n"
        "1. Bias arising from the randomization process\n"
        "2. Bias due to deviations from intended interventions\n"
        "3. Bias due to missing outcome data\n"
        "4. Bias in measurement of the outcome\n"
        "5. Bias in selection of the reported result\n\n"
        "For each domain, judge the risk as 'Low risk', 'Some concerns', or "
        "'High risk'. Then provide an overall risk of bias judgement.\n\n"
        "Reply with ONLY a Python list: [overall_judgement, 'explanation "
        "covering each domain']. The overall_judgement must be one of: 'Low', "
        "'Some concerns', or 'High'. Do not include any other text.\n\n"
        "The systematic review topic is: {topic}\n\n"
        "Here is the source:\nTitle: {title}\n{source_text}"
    ),

    # === CITATION VALIDATION ===
    "ai_validate_citation": (
        "You are a physician scientist validating citations in a systematic review. "
        "The following text appears before citation [{citation_number}]:\n"
        "'{preceding_text}'\n\n"
        "Here are the source materials for source [{citation_number}]:\n"
        "{source_context}\n\n"
        "Does the information in the preceding text accurately come from this source? "
        "Reply with ONLY a Python list: [True/False, 'explanation of where the match "
        "was found or why no match exists']. Do not include any other text."
    ),

    "ai_validate_citation_batch": (
        "You are a physician scientist validating a citation in a systematic review. "
        "The following text appears before citation [{citation_number}]:\n"
        "'{preceding_text}'\n\n"
        "Here is the source material:\n{source_context}\n\n"
        "Does this text accurately come from this source? Reply with ONLY 'True' or "
        "'False'. Do not include any other text."
    ),

    # === ITERATIVE TOPIC/STAT GENERATION ===
    "generate_topic_iterative": (
        "You are a physician scientist investigating the research question: "
        "'{topic_title}' as part of a systematic review on '{topic}'. "
        "You are examining sources one at a time to build a comprehensive, "
        "evidence-based response.\n\n"
        "Here is source [{source_number}]:\n{source_text}\n\n"
        "{existing_text_clause}"
        "Extract all information from this source that is relevant to the "
        "research question. Cite all information using [{source_number}]. "
        "If there is nothing relevant, reply with ONLY the word 'SKIP'. "
        "Do not repeat information already covered in the existing text."
    ),

    "generate_stat_iterative": (
        "You are a physician scientist extracting statistical data for the "
        "question: '{question}' as part of a systematic review.\n\n"
        "Here is source [{source_number}]:\n{source_text}\n\n"
        "{existing_text_clause}"
        "Extract any specific numerical data, statistics, or quantitative "
        "findings from this source that answer or relate to the question. "
        "Cite all data using [{source_number}]. "
        "If there is no relevant numerical data, reply with ONLY the word 'SKIP'."
    ),

    # === DISTILLATION ===
    "distill_topic": (
        "You are a physician scientist editing a topic summary for a systematic "
        "review. The following text was assembled from multiple sources and may be "
        "verbose or repetitive. Distill it into a concise, well-organized summary "
        "that preserves all crucial findings and maintains every in-text citation "
        "exactly as written (do not add, remove, or change citation numbers). "
        "Remove redundancy, unnecessary phrasing, and filler while keeping all "
        "unique data points and conclusions.\n\n"
        "Here is the text to distill:\n{text}"
    ),

    "distill_stat": (
        "You are a physician scientist editing a statistical summary for a "
        "systematic review. The following text contains numerical data extracted "
        "from multiple sources. Distill it into a concise summary that preserves "
        "all specific numbers, statistics, and quantitative findings. Maintain "
        "every in-text citation exactly as written. Remove redundancy and "
        "unnecessary prose while keeping all unique data points.\n\n"
        "Here is the text to distill:\n{text}"
    ),
}
```
