## Supplementary material for "In Search of Ethical Procedures for LLM-Assisted Systematic Review Production: A Proof-of-Concept Evaluation of Selected Review Components": source_data: dialogs.py

```
"""
Dialogs for the Systematic Review Foundry.
Includes prompt approval, context configuration, output approval, and settings.
"""
import ast
import re
from PyQt6.QtWidgets import (
    QDialog, QVBoxLayout, QHBoxLayout, QTextEdit, QLabel, QPushButton,
    QDialogButtonBox, QMessageBox, QToolTip, QGroupBox, QLineEdit,
    QComboBox, QListWidget, QListWidgetItem, QSpinBox, QDoubleSpinBox,
    QTabWidget, QWidget, QScrollArea, QCheckBox, QSplitter, QAbstractItemView,
    QFormLayout, QApplication
)
from PyQt6.QtCore import Qt, QPoint, pyqtSignal
from PyQt6.QtGui import QFont, QCursor, QColor, QTextCursor, QTextDocument

# Model discovery lives in config_manager; the UI only presents it.
from config_manager import (newest_per_family, is_known_model,
                            profile_for_model, MODEL_TABLE_DATE)


# ═══════════════════════════════════════════════════════════════════
#  Startup model notice
# ═══════════════════════════════════════════════════════════════════

def show_model_notice(config, parent=None):
    """
    One-time-per-launch notice telling the user which model is active and
    how much this build actually knows about it.

    Call from the main window once the UI is up:

        from dialogs import show_model_notice
        show_model_notice(self.config, self)

    Silently does nothing if the user ticked "Don't show this again".
    """
    if not config.show_model_notice:
        return

    model = config.claude_model
    known = is_known_model(model)
    profile = profile_for_model(model)

    if known:
        recognised = (
            f"This model is in the built-in table, which was last checked "
            f"against Anthropic's documentation on <b>{MODEL_TABLE_DATE}</b>.")
    else:
        recognised = (
            f"This model is <b>not</b> in the built-in table (last checked "
            f"{MODEL_TABLE_DATE}), so only generic parameters will be sent. "
            f"That is the safe default, but it may not be optimal.")

    if profile['sampling']:
        sampling = ("Requests to it <b>will</b> include your Temperature "
                    "(or Top-P) setting.")
    else:
        sampling = ("Requests to it will <b>not</b> include Temperature or "
                    "Top-P. Anthropic removed those controls on newer "
                    "models, which reject them outright.")

    box = QMessageBox(parent)
    box.setWindowTitle("Claude Model In Use")
    box.setIcon(QMessageBox.Icon.Information)
    box.setTextFormat(Qt.TextFormat.RichText)
    box.setText(f"<b>Active model: {model}</b>")
    box.setInformativeText(
        f"{recognised}<br><br>"
        f"{sampling}<br><br>"
        f"Anthropic retires models and changes which parameters they "
        f"accept over time, so depending on today's date this model may no "
        f"longer be available, and its parameter support may have changed. "
        f"<b>Please verify against Anthropic's current documentation "
        f"rather than relying on this build.</b><br><br>"
        f"To switch models, or to ask the API which models your key can "
        f"use right now, open <b>Prompt Settings → API Settings</b> and "
        f"press <b>Check for New Models</b>.")

    dont_show = QCheckBox("Don't show this again")
    box.setCheckBox(dont_show)
    box.exec()

    if dont_show.isChecked():
        config.show_model_notice = False


class PromptApprovalDialog(QDialog):
    """
    Dialog shown before sending any prompt to the LLM.
    Displays the prompt template with f-string variables abbreviated.
    Double-clicking a variable shows its full content in a popup.
    """

    def __init__(self, prompt_key: str, prompt_template: str,
                 variables: dict, config_manager, parent=None):
        super().__init__(parent)
        self.prompt_key = prompt_key
        self.original_template = prompt_template
        self.variables = variables
        self.config = config_manager
        self.result_action = None

        self.setWindowTitle(f"Approve Prompt: {prompt_key}")
        self.setMinimumSize(700, 500)
        self._build_ui()

    def _build_ui(self):
        layout = QVBoxLayout(self)
        info = QLabel(f"<b>Prompt:</b> {self.prompt_key}")
        layout.addWidget(info)

        if self.variables:
            var_group = QGroupBox("Variables (click to expand)")
            var_layout = QVBoxLayout(var_group)
            for name, value in self.variables.items():
                val_str = str(value)
                short = (val_str[:80] + "..."
                         if len(val_str) > 80 else val_str)
                btn = QPushButton(f"  {{{name}}} = {short}")
                btn.setStyleSheet("text-align: left; padding: 4px;")
                btn.setToolTip("Click to see full value")
                full_value = val_str

                def make_handler(n, v):
                    def handler():
                        dlg = QDialog(self)
                        dlg.setWindowTitle(f"Variable: {{{n}}}")
                        dlg.setMinimumSize(500, 400)
                        lay = QVBoxLayout(dlg)
                        te = QTextEdit()
                        te.setReadOnly(True)
                        te.setPlainText(v)
                        lay.addWidget(te)
                        close_btn = QPushButton("Close")
                        close_btn.clicked.connect(dlg.close)
                        lay.addWidget(close_btn)
                        dlg.exec()
                    return handler

                btn.clicked.connect(make_handler(name, full_value))
                var_layout.addWidget(btn)
            layout.addWidget(var_group)

        layout.addWidget(QLabel("Prompt Template (editable):"))
        self.prompt_edit = QTextEdit()
        self.prompt_edit.setPlainText(self.original_template)
        self.prompt_edit.setFont(QFont("Consolas", 10))
        layout.addWidget(self.prompt_edit)

        btn_layout = QHBoxLayout()
        for label, handler in [
            ("Send As-Is", self._send),
            ("Send Edited (Don't Save)", self._send_edited),
            ("Save & Send", self._save_and_send),
            ("Reset to Default", self._reset),
            ("Cancel", self.reject),
        ]:
            btn = QPushButton(label)
            btn.clicked.connect(handler)
            btn_layout.addWidget(btn)
        layout.addLayout(btn_layout)

    def _validate_fstring(self, template: str) -> bool:
        try:
            template.format(**{k: "" for k in self.variables})
            return True
        except (KeyError, ValueError, IndexError):
            return False

    def _send(self):
        self.result_action = "send"
        self.accept()

    def _send_edited(self):
        edited = self.prompt_edit.toPlainText()
        if not self._validate_fstring(edited):
            QMessageBox.warning(
                self, "Invalid Template",
                "The edited prompt has invalid f-string variables. "
                "Edit was rejected.")
            return
        self.original_template = edited
        self.result_action = "send_edited"
        self.accept()

    def _save_and_send(self):
        edited = self.prompt_edit.toPlainText()
        if not self._validate_fstring(edited):
            QMessageBox.warning(
                self, "Invalid Template",
                "The edited prompt has invalid f-string variables. "
                "Edit was rejected.")
            return
        self.config.set_custom_prompt(self.prompt_key, edited)
        self.original_template = edited
        self.result_action = "save_and_send"
        self.accept()

    def _reset(self):
        from default_prompts import DEFAULT_PROMPTS
        default = DEFAULT_PROMPTS.get(self.prompt_key, "")
        self.prompt_edit.setPlainText(default)
        self.config.reset_prompt(self.prompt_key)

    def get_formatted_prompt(self) -> str:
        return self.original_template.format(**self.variables)


class OutputApprovalDialog(QDialog):
    """Dialog to approve or reject LLM output before writing to a section."""

    def __init__(self, section_name: str, output_text: str, parent=None):
        super().__init__(parent)
        self.setWindowTitle(f"Approve Output: {section_name}")
        self.setMinimumSize(600, 450)
        self.approved = False

        layout = QVBoxLayout(self)
        layout.addWidget(
            QLabel(f"<b>LLM output for:</b> {section_name}"))

        self.output_view = QTextEdit()
        self.output_view.setPlainText(output_text)
        self.output_view.setReadOnly(True)
        layout.addWidget(self.output_view)

        btn_layout = QHBoxLayout()
        approve_btn = QPushButton("✓ Approve && Apply")
        approve_btn.clicked.connect(self._approve)
        btn_layout.addWidget(approve_btn)
        reject_btn = QPushButton("✗ Reject")
        reject_btn.clicked.connect(self.reject)
        btn_layout.addWidget(reject_btn)
        layout.addLayout(btn_layout)

    def _approve(self):
        self.approved = True
        self.accept()


# ═══════════════════════════════════════════════════════════════════
#  Helper: resolve the section_rate index for a section_key
# ═══════════════════════════════════════════════════════════════════

def _section_rate_index(section_key: str) -> int:
    """
    Map a section_key to the index in source['section_rate'].
    section_rate is built as: [intro, results_0, results_1, ...]
    Returns -1 for sections without a specific rating index.
    """
    if section_key == 'intro':
        return 0
    if section_key.startswith('results_'):
        try:
            return int(section_key.split('_')[1]) + 1
        except (ValueError, IndexError):
            return -1
    return -1


def _get_source_rating_for_section(src: dict, section_key: str,
                                    topic_id: str = None,
                                    stat_id: str = None) -> int:
    """
    Return the rating (1-10) of a source for a given section, topic,
    or stat. Returns 0 if no rating exists.
    """
    if topic_id:
        tr = src.get('topic_ratings') or {}
        return tr.get(topic_id, 0)

    if stat_id:
        sr = src.get('stat_ratings') or {}
        return sr.get(stat_id, 0)

    idx = _section_rate_index(section_key)
    if idx < 0:
        return 0
    sr = src.get('section_rate') or []
    if idx < len(sr):
        return sr[idx]
    return 0


# ═══════════════════════════════════════════════════════════════════
#  Filterable list with rating-aware auto-select
# ═══════════════════════════════════════════════════════════════════

class _FilterableList(QWidget):
    """A list widget with search, bulk buttons, and selection status label."""

    def __init__(self, parent=None):
        super().__init__(parent)
        layout = QVBoxLayout(self)
        layout.setContentsMargins(0, 0, 0, 0)

        # Search bar
        search_row = QHBoxLayout()
        search_row.addWidget(QLabel("🔍"))
        self.search_edit = QLineEdit()
        self.search_edit.setPlaceholderText("Filter items…")
        self.search_edit.textChanged.connect(self._filter)
        search_row.addWidget(self.search_edit)
        layout.addLayout(search_row)

        # Bulk buttons
        bulk_row = QHBoxLayout()
        add_all = QPushButton("Select All")
        add_all.clicked.connect(self._select_all)
        bulk_row.addWidget(add_all)
        remove_all = QPushButton("Deselect All")
        remove_all.clicked.connect(self._deselect_all)
        bulk_row.addWidget(remove_all)
        bulk_row.addStretch()
        # Selection count label
        self.count_label = QLabel("")
        self.count_label.setStyleSheet("color: #666; font-style: italic;")
        bulk_row.addWidget(self.count_label)
        layout.addLayout(bulk_row)

        # List
        self.list_widget = QListWidget()
        self.list_widget.setSelectionMode(
            QAbstractItemView.SelectionMode.MultiSelection)
        self.list_widget.itemSelectionChanged.connect(self._update_count)
        layout.addWidget(self.list_widget)

    def add_item(self, text: str, data, selected: bool = False,
                 rating: int = 0):
        """Add an item. If rating > 0, it is shown as a prefix."""
        if rating > 0:
            display = f"[Rating: {rating}] {text}"
        else:
            display = text
        item = QListWidgetItem(display)
        item.setData(Qt.ItemDataRole.UserRole, data)
        item.setData(Qt.ItemDataRole.UserRole + 1, rating)
        # Color-code by rating
        if rating >= 7:
            item.setForeground(QColor("#27ae60"))
        elif 0 < rating <= 3:
            item.setForeground(QColor("#c0392b"))
        # MUST add to list before setting selection — Qt ignores
        # setSelected() on items that don't belong to a widget yet.
        self.list_widget.addItem(item)
        if selected:
            item.setSelected(True)

    def get_selected_data(self) -> list:
        return [self.list_widget.item(i).data(Qt.ItemDataRole.UserRole)
                for i in range(self.list_widget.count())
                if self.list_widget.item(i).isSelected()]

    def select_top_n(self, n: int):
        """Select the top N items by rating (descending), deselect others."""
        items_with_ratings = []
        for i in range(self.list_widget.count()):
            item = self.list_widget.item(i)
            rating = item.data(Qt.ItemDataRole.UserRole + 1) or 0
            items_with_ratings.append((i, rating))

        # Sort by rating descending
        items_with_ratings.sort(key=lambda x: x[1], reverse=True)

        for rank, (idx, rating) in enumerate(items_with_ratings):
            item = self.list_widget.item(idx)
            item.setSelected(rank < n and rating > 0)

        self._update_count()

    def has_any_ratings(self) -> bool:
        for i in range(self.list_widget.count()):
            r = self.list_widget.item(i).data(Qt.ItemDataRole.UserRole + 1)
            if r and r > 0:
                return True
        return False

    def _filter(self, text: str):
        text_lower = text.lower()
        for i in range(self.list_widget.count()):
            item = self.list_widget.item(i)
            item.setHidden(text_lower not in item.text().lower())

    def _select_all(self):
        for i in range(self.list_widget.count()):
            item = self.list_widget.item(i)
            if not item.isHidden():
                item.setSelected(True)
        self._update_count()

    def _deselect_all(self):
        for i in range(self.list_widget.count()):
            item = self.list_widget.item(i)
            if not item.isHidden():
                item.setSelected(False)
        self._update_count()

    def _update_count(self):
        total = self.list_widget.count()
        selected = len(self.get_selected_data())
        self.count_label.setText(f"{selected} / {total} included in context")


# ═══════════════════════════════════════════════════════════════════
#  Context Configuration Dialog
# ═══════════════════════════════════════════════════════════════════

class ContextConfigDialog(QDialog):
    """
    Dialog for configuring what context is sent to the LLM for a section.
    Knows which section_key it's configuring so it can look up relevant
    section ratings and auto-select top-rated sources.
    """

    def __init__(self, section_key: str, session,
                 current_config: dict = None, parent=None,
                 topic_id: str = None, stat_id: str = None):
        """
        section_key: e.g. 'intro', 'results_0', 'discussion', or a topic/stat id
        topic_id:    if set, use topic_ratings instead of section_rate
        stat_id:     if set, use stat_ratings instead of section_rate
        """
        super().__init__(parent)
        self.setWindowTitle(f"Context Configuration: {section_key}")
        self.setMinimumSize(850, 650)
        self.session = session
        self.section_key = section_key
        self.topic_id = topic_id
        self.stat_id = stat_id
        self.config = current_config or {}
        self._is_first_open = not bool(current_config)
        # Sections that rely on review_string in their prompts and should
        # NOT include source summaries by default
        self._no_default_summaries = section_key in (
            'discussion', 'conclusion', 'abstract')
        self._build_ui()

    def _build_ui(self):
        layout = QVBoxLayout(self)

        # Legend
        legend = QLabel(
            "<b>Legend:</b> "
            "<span style='color:#27ae60'>■ Green = high rating (7-10)</span> · "
            "<span style='color:#c0392b'>■ Red = low rating (1-3)</span> · "
            "Selected items = <b>included</b> in context. "
            "Ratings come from AI section/topic relevance scoring.")
        legend.setWordWrap(True)
        legend.setStyleSheet(
            "background: #f0f0f0; padding: 6px; border-radius: 4px;")
        layout.addWidget(legend)

        tabs = QTabWidget()

        # ── Source Summaries (with rating-aware auto-select) ──
        summary_tab = QWidget()
        summary_layout = QVBoxLayout(summary_tab)

        # Top-N controls
        topn_row = QHBoxLayout()
        topn_row.addWidget(QLabel("Auto-select top"))
        self.top_n_spin = QSpinBox()
        self.top_n_spin.setRange(1, 999)
        self.top_n_spin.setValue(
            self.config.get('top_n', 10))
        topn_row.addWidget(self.top_n_spin)
        topn_row.addWidget(QLabel("rated sources"))
        auto_btn = QPushButton("Auto-Select Top N")
        auto_btn.setToolTip(
            "Deselect all, then select only the top N sources by rating")
        auto_btn.clicked.connect(self._auto_select_top_n)
        topn_row.addWidget(auto_btn)
        topn_row.addStretch()

        # Summary limit
        topn_row.addWidget(QLabel("Hard limit on summaries sent:"))
        self.summary_limit = QSpinBox()
        self.summary_limit.setRange(0, 9999)
        self.summary_limit.setValue(self.config.get('summary_limit', 0))
        self.summary_limit.setSpecialValueText("No limit")
        self.summary_limit.setToolTip(
            "Even among selected sources, cap how many summaries "
            "are included (0 = no cap)")
        topn_row.addWidget(self.summary_limit)
        summary_layout.addLayout(topn_row)

        self.summary_flist = _FilterableList()

        # Determine whether the user has previously saved a context config.
        # This is the key distinction: if a config was saved, we honour it
        # exactly (even if the user selected zero summaries). If no config
        # was saved we apply defaults.
        _has_saved_config = not self._is_first_open
        selected_summaries = set(self.config.get('summaries', []))

        # Determine which rating to show per source
        has_ratings = False
        for s in self.session.sources:
            src = (s if isinstance(s, dict)
                   else s.to_dict() if hasattr(s, 'to_dict') else s)
            num = src.get('number', '?')
            title = src.get('title', 'Untitled')[:55]
            has_summary = "✓" if src.get('summary') else "✗"
            rating = _get_source_rating_for_section(
                src, self.section_key, self.topic_id, self.stat_id)
            if rating > 0:
                has_ratings = True

            # Selection logic:
            # 1) User previously saved a config → restore exactly
            # 2) First open + discussion/conclusion/abstract → empty
            # 3) First open + ratings exist → defer to auto-select below
            # 4) First open + no ratings → include non-rejected only
            #    (rating=True or rating=None/unscreened; never rating=False)
            is_excluded = src.get('rating') is False
            if _has_saved_config:
                sel = num in selected_summaries
            elif self._no_default_summaries:
                sel = False
            elif has_ratings:
                sel = False  # will be set by auto-select below
            else:
                sel = not is_excluded  # first open, no ratings → include non-rejected

            self.summary_flist.add_item(
                f"[{num}] {has_summary} {title}",
                num, selected=sel, rating=rating)

        summary_layout.addWidget(self.summary_flist)
        tabs.addTab(summary_tab, "Source Summaries")

        # Auto-select top 10 on first open if ratings exist
        # and not a no-summary section (discussion/conclusion/abstract)
        if self._is_first_open and has_ratings \
           and not self._no_default_summaries:
            self.summary_flist.select_top_n(self.top_n_spin.value())

        # ── Full Texts ──
        ft_tab = QWidget()
        ft_layout = QVBoxLayout(ft_tab)
        self.fulltext_flist = _FilterableList()
        selected_ft = set(self.config.get('full_texts', []))
        for s in self.session.sources:
            src = (s if isinstance(s, dict)
                   else s.to_dict() if hasattr(s, 'to_dict') else s)
            if src.get('full_text'):
                num = src.get('number', '?')
                title = src.get('title', 'Untitled')[:60]
                rating = _get_source_rating_for_section(
                    src, self.section_key, self.topic_id, self.stat_id)
                self.fulltext_flist.add_item(
                    f"[{num}] {title}", num,
                    selected=(num in selected_ft), rating=rating)
        ft_layout.addWidget(self.fulltext_flist)
        tabs.addTab(ft_tab, "Full Texts")

        # ── Topics ──
        topic_tab = QWidget()
        topic_layout = QVBoxLayout(topic_tab)
        self.topic_flist = _FilterableList()
        selected_topics = set(self.config.get('topics', []))
        for t in self.session.topics:
            top = (t if isinstance(t, dict)
                   else t.to_dict() if hasattr(t, 'to_dict') else t)
            tid = top.get('topic_id', '?')
            title = top.get('title', 'Untitled')[:60]
            has_text = "✓" if top.get('text') else "✗"
            self.topic_flist.add_item(
                f"{tid}: {has_text} {title}", tid,
                selected=(tid in selected_topics))
        topic_layout.addWidget(self.topic_flist)
        tabs.addTab(topic_tab, "Topics")

        # ── Statistics ──
        stat_tab = QWidget()
        stat_layout = QVBoxLayout(stat_tab)
        self.stat_flist = _FilterableList()
        selected_stats = set(self.config.get('statistics', []))
        for st in self.session.statistics:
            stat = (st if isinstance(st, dict)
                    else st.to_dict() if hasattr(st, 'to_dict') else st)
            sid = stat.get('stat_id', '?')
            q = stat.get('question', 'Untitled')[:60]
            self.stat_flist.add_item(
                f"{sid}: {q}", sid,
                selected=(sid in selected_stats))
        stat_layout.addWidget(self.stat_flist)
        tabs.addTab(stat_tab, "Statistics")

        # ── Review Sections ──
        section_tab = QWidget()
        section_layout = QVBoxLayout(section_tab)
        self.section_flist = _FilterableList()
        selected_sections = set(self.config.get('review_sections', []))
        for sec_name in ['abstract', 'intro', 'methods',
                         'discussion', 'conclusion']:
            self.section_flist.add_item(
                sec_name.capitalize(), sec_name,
                selected=(sec_name in selected_sections))
        for rs in self.session.results:
            r = rs if isinstance(rs, dict) else rs
            sec = r.get('section', 'Results')
            key = f"results_{r.get('section_number', 0)}"
            self.section_flist.add_item(
                f"Results: {sec}", key,
                selected=(key in selected_sections))
        section_layout.addWidget(self.section_flist)
        tabs.addTab(section_tab, "Review Sections")

        layout.addWidget(tabs)

        # ── Included summary at bottom ──
        self.included_label = QLabel("")
        self.included_label.setStyleSheet("font-weight: bold; padding: 4px;")
        layout.addWidget(self.included_label)
        self._update_included_summary()

        # Connect change signals for live summary
        self.summary_flist.list_widget.itemSelectionChanged.connect(
            self._update_included_summary)
        self.fulltext_flist.list_widget.itemSelectionChanged.connect(
            self._update_included_summary)
        self.topic_flist.list_widget.itemSelectionChanged.connect(
            self._update_included_summary)

        # Buttons
        btn_layout = QHBoxLayout()
        ok_btn = QPushButton("Apply")
        ok_btn.clicked.connect(self.accept)
        btn_layout.addWidget(ok_btn)
        cancel_btn = QPushButton("Cancel")
        cancel_btn.clicked.connect(self.reject)
        btn_layout.addWidget(cancel_btn)
        layout.addLayout(btn_layout)

    def _auto_select_top_n(self):
        self.summary_flist.select_top_n(self.top_n_spin.value())
        self._update_included_summary()

    def _update_included_summary(self):
        n_sum = len(self.summary_flist.get_selected_data())
        n_ft = len(self.fulltext_flist.get_selected_data())
        n_top = len(self.topic_flist.get_selected_data())
        self.included_label.setText(
            f"Context will include: {n_sum} summaries, "
            f"{n_ft} full texts, {n_top} topics")

    def get_config(self) -> dict:
        return {
            'summaries': self.summary_flist.get_selected_data(),
            'summary_limit': self.summary_limit.value(),
            'top_n': self.top_n_spin.value(),
            'full_texts': self.fulltext_flist.get_selected_data(),
            'topics': self.topic_flist.get_selected_data(),
            'statistics': self.stat_flist.get_selected_data(),
            'review_sections': self.section_flist.get_selected_data(),
        }


# ═══════════════════════════════════════════════════════════════════
#  Link to Sections Dialog
# ═══════════════════════════════════════════════════════════════════

class LinkToSectionsDialog(QDialog):
    """
    Dialog for linking a topic or statistic to one or more review sections.
    Shows all available sections as checkboxes so the user can click rather
    than type.  On accept, returns the list of selected section keys and
    updates ``session.section_contexts`` so each chosen section's context
    includes the linked topic/stat — exactly as if the user had configured
    it through ContextConfigDialog.
    """

    def __init__(self, item_label: str, session,
                 current_linked: list = None, parent=None, *,
                 topic_id: str = None, stat_id: str = None):
        """
        Parameters
        ----------
        item_label : str
            Human-readable name shown in the title (e.g. "T3: Mortality rates").
        session : Session
            The live session object.
        current_linked : list[str]
            Section keys already linked (used to pre-check boxes).
        topic_id / stat_id : str
            Exactly one should be set — determines whether the link adds a
            topic or a statistic to each section's context.
        """
        super().__init__(parent)
        self.setWindowTitle(f"Link to Sections — {item_label}")
        self.setMinimumWidth(420)
        self.session = session
        self.topic_id = topic_id
        self.stat_id = stat_id
        self._current_linked = set(current_linked or [])
        self._checkboxes: list[tuple[QCheckBox, str]] = []  # (widget, key)
        self._build_ui()

    # ── UI ──

    def _build_ui(self):
        layout = QVBoxLayout(self)

        hint = QLabel(
            "Select the review sections this item should be included in.\n"
            "Linked items are automatically added to each section's "
            "AI context configuration.")
        hint.setWordWrap(True)
        hint.setStyleSheet(
            "background: #eaf0fb; padding: 8px; border-radius: 4px;")
        layout.addWidget(hint)

        # ── Fixed sections ──
        group = QGroupBox("Sections")
        g_layout = QVBoxLayout(group)

        for key, display in [("intro", "Introduction"),
                              ("discussion", "Discussion"),
                              ("conclusion", "Conclusion")]:
            cb = QCheckBox(display)
            cb.setChecked(key in self._current_linked)
            g_layout.addWidget(cb)
            self._checkboxes.append((cb, key))

        # ── Results subsections (dynamic) ──
        if self.session.results:
            g_layout.addWidget(QLabel(""))  # spacer
            g_layout.addWidget(QLabel("Results subsections:"))
            for i, rs in enumerate(self.session.results):
                sec = rs.get('section', f'Subsection {i + 1}')
                key = f"results_{i}"
                # Also recognise the legacy format "results: <name>"
                legacy_key = f"results: {sec}"
                pre_checked = (key in self._current_linked
                               or legacy_key in self._current_linked)
                cb = QCheckBox(f"Results: {sec}")
                cb.setChecked(pre_checked)
                g_layout.addWidget(cb)
                self._checkboxes.append((cb, key))

        layout.addWidget(group)

        # ── Quick-select helpers ──
        helper_row = QHBoxLayout()
        all_btn = QPushButton("Select All")
        all_btn.clicked.connect(self._select_all)
        helper_row.addWidget(all_btn)
        none_btn = QPushButton("Deselect All")
        none_btn.clicked.connect(self._deselect_all)
        helper_row.addWidget(none_btn)
        helper_row.addStretch()
        layout.addLayout(helper_row)

        # ── OK / Cancel ──
        btn_layout = QHBoxLayout()
        ok_btn = QPushButton("OK")
        ok_btn.setDefault(True)
        ok_btn.clicked.connect(self.accept)
        btn_layout.addWidget(ok_btn)
        cancel_btn = QPushButton("Cancel")
        cancel_btn.clicked.connect(self.reject)
        btn_layout.addWidget(cancel_btn)
        layout.addLayout(btn_layout)

    # ── helpers ──

    def _select_all(self):
        for cb, _ in self._checkboxes:
            cb.setChecked(True)

    def _deselect_all(self):
        for cb, _ in self._checkboxes:
            cb.setChecked(False)

    # ── public API ──

    def get_selected_keys(self) -> list[str]:
        """Return the canonical section keys that were checked."""
        return [key for cb, key in self._checkboxes if cb.isChecked()]

    def apply_to_session(self):
        """
        Write the link into ``session.section_contexts`` so that each
        selected section's context includes this topic/stat.  Sections
        that were *un*-checked are cleaned up as well.
        """
        selected = set(self.get_selected_keys())

        # Every section key that *could* be linked
        all_keys = {key for _, key in self._checkboxes}

        for key in all_keys:
            ctx = self.session.section_contexts.setdefault(key, {})

            if self.topic_id:
                topics = ctx.setdefault('topics', [])
                if key in selected:
                    if self.topic_id not in topics:
                        topics.append(self.topic_id)
                else:
                    if self.topic_id in topics:
                        topics.remove(self.topic_id)

            if self.stat_id:
                stats = ctx.setdefault('statistics', [])
                if key in selected:
                    if self.stat_id not in stats:
                        stats.append(self.stat_id)
                else:
                    if self.stat_id in stats:
                        stats.remove(self.stat_id)


# ═══════════════════════════════════════════════════════════════════
#  Prompt Settings Dialog
# ═══════════════════════════════════════════════════════════════════

class PromptSettingsDialog(QDialog):
    """
    Master prompt settings dialog.
    Allows viewing/editing all prompts, configuring APIs, and model params.
    """

    def __init__(self, config_manager, api_manager, parent=None):
        super().__init__(parent)
        self.config = config_manager
        self.api = api_manager
        self.setWindowTitle("Prompt & API Settings")
        self.setMinimumSize(900, 650)
        self._build_ui()

    def _build_ui(self):
        layout = QVBoxLayout(self)
        tabs = QTabWidget()

        # === API Configuration Tab ===
        api_tab = QWidget()
        api_layout = QVBoxLayout(api_tab)

        api_select = QHBoxLayout()
        api_select.addWidget(QLabel("Active API:"))
        self.api_combo = QComboBox()
        self.api_combo.addItems(["claude", "ollama"])
        self.api_combo.setCurrentText(self.config.active_api)
        api_select.addWidget(self.api_combo)
        api_layout.addLayout(api_select)

        # Claude settings
        claude_group = QGroupBox("Claude API")
        claude_layout = QFormLayout(claude_group)
        self.claude_key_edit = QLineEdit(self.config.claude_api_key)
        self.claude_key_edit.setEchoMode(QLineEdit.EchoMode.Password)
        claude_layout.addRow("API Key:", self.claude_key_edit)
        self.claude_model_combo = QComboBox()
        self.claude_model_combo.setEditable(True)
        # Hardcoded table, or the fetched list if Check for New Models
        # has been used. No network access happens on open.
        self._claude_models_all = self.config.claude_models
        claude_model_row = QHBoxLayout()
        claude_model_row.addWidget(self.claude_model_combo, 1)
        self.claude_refresh_btn = QPushButton("Check for New Models")
        self.claude_refresh_btn.clicked.connect(
            lambda: self._refresh_claude_models())
        claude_model_row.addWidget(self.claude_refresh_btn)
        claude_layout.addRow("Model:", claude_model_row)

        self.claude_show_old = QCheckBox("Show older / dated versions")
        self.claude_show_old.toggled.connect(
            lambda: self._populate_claude_combo())
        claude_layout.addRow("", self.claude_show_old)

        self.claude_auto_newest = QCheckBox(
            "Always use the newest available model")
        self.claude_auto_newest.setChecked(not self.config.claude_model_pinned)
        self.claude_auto_newest.toggled.connect(self._on_auto_newest_toggled)
        claude_layout.addRow("", self.claude_auto_newest)

        # activated fires only on user interaction, not on programmatic
        # repopulation, so it is the right signal for "the user picked one".
        self.claude_model_combo.activated.connect(
            lambda: self.claude_auto_newest.setChecked(False))

        self.claude_model_status = QLabel("")
        self.claude_model_status.setStyleSheet("color: #888;")
        self.claude_model_status.setWordWrap(True)
        claude_layout.addRow("", self.claude_model_status)

        self._populate_claude_combo()
        self.claude_model_status.setText(
            "Built-in model list. Press Check for New Models to ask the "
            "API what else is available.")
        api_layout.addWidget(claude_group)

        # Ollama settings
        ollama_group = QGroupBox("Ollama (Local)")
        ollama_layout = QFormLayout(ollama_group)
        self.ollama_url_edit = QLineEdit(self.config.ollama_url)
        ollama_layout.addRow("URL:", self.ollama_url_edit)
        model_row = QHBoxLayout()
        self.ollama_model_combo = QComboBox()
        self.ollama_model_combo.setEditable(True)
        for m in self.config.ollama_models:
            self.ollama_model_combo.addItem(m)
        if self.config.active_ollama_model:
            self.ollama_model_combo.setCurrentText(
                self.config.active_ollama_model)
        refresh_btn = QPushButton("Refresh Models")
        refresh_btn.clicked.connect(self._refresh_ollama_models)
        model_row.addWidget(self.ollama_model_combo)
        model_row.addWidget(refresh_btn)
        ollama_layout.addRow("Model:", model_row)

        add_model_row = QHBoxLayout()
        self.new_model_edit = QLineEdit()
        self.new_model_edit.setPlaceholderText("model_name:tag")
        add_btn = QPushButton("Add")
        add_btn.clicked.connect(self._add_ollama_model)
        add_model_row.addWidget(self.new_model_edit)
        add_model_row.addWidget(add_btn)
        ollama_layout.addRow("Add Model:", add_model_row)
        api_layout.addWidget(ollama_group)

        # NCBI API Key
        ncbi_group = QGroupBox("NCBI PubMed")
        ncbi_layout = QFormLayout(ncbi_group)
        self.ncbi_key_edit = QLineEdit(self.config.ncbi_api_key)
        self.ncbi_key_edit.setEchoMode(QLineEdit.EchoMode.Password)
        ncbi_layout.addRow("API Key:", self.ncbi_key_edit)
        cred = self.config.institutional_credentials
        self.inst_user_edit = QLineEdit(cred.get('username', ''))
        ncbi_layout.addRow("Institutional User (Beta):",
                           self.inst_user_edit)
        self.inst_pass_edit = QLineEdit(cred.get('password', ''))
        self.inst_pass_edit.setEchoMode(QLineEdit.EchoMode.Password)
        ncbi_layout.addRow("Institutional Pass (Beta):",
                           self.inst_pass_edit)
        api_layout.addWidget(ncbi_group)

        test_btn = QPushButton("Test Active API Connection")
        test_btn.clicked.connect(self._test_connection)
        api_layout.addWidget(test_btn)
        self.startup_notice_check = QCheckBox(
            "Show the model notice at startup")
        self.startup_notice_check.setChecked(self.config.show_model_notice)
        api_layout.addWidget(self.startup_notice_check)
        self.test_label = QLabel("")
        api_layout.addWidget(self.test_label)
        api_layout.addStretch()
        tabs.addTab(api_tab, "API Settings")

        # === Prompts Tab ===
        prompt_tab = QWidget()
        prompt_layout = QHBoxLayout(prompt_tab)

        self.prompt_list = QListWidget()
        for key in self.config.get_all_prompt_keys():
            item = QListWidgetItem(key)
            if self.config.is_prompt_customized(key):
                item.setText(f"★ {key}")
            self.prompt_list.addItem(item)
        self.prompt_list.currentRowChanged.connect(
            self._on_prompt_selected)
        prompt_layout.addWidget(self.prompt_list, 1)

        right = QVBoxLayout()
        self.prompt_key_label = QLabel("")
        right.addWidget(self.prompt_key_label)
        self.prompt_text_edit = QTextEdit()
        self.prompt_text_edit.setFont(QFont("Consolas", 10))
        right.addWidget(self.prompt_text_edit)

        param_group = QGroupBox("Parameters for this prompt")
        param_layout = QFormLayout(param_group)
        self.p_max_tokens = QSpinBox()
        self.p_max_tokens.setRange(100, 100000)
        param_layout.addRow("Max Tokens:", self.p_max_tokens)
        self.p_temperature = QDoubleSpinBox()
        self.p_temperature.setRange(0.0, 2.0)
        self.p_temperature.setSingleStep(0.1)
        param_layout.addRow("Temperature:", self.p_temperature)
        right.addWidget(param_group)

        prompt_btn_layout = QHBoxLayout()
        save_prompt_btn = QPushButton("Save Prompt")
        save_prompt_btn.clicked.connect(self._save_current_prompt)
        prompt_btn_layout.addWidget(save_prompt_btn)
        reset_prompt_btn = QPushButton("Reset to Default")
        reset_prompt_btn.clicked.connect(self._reset_current_prompt)
        prompt_btn_layout.addWidget(reset_prompt_btn)
        right.addLayout(prompt_btn_layout)

        prompt_layout.addLayout(right, 2)
        tabs.addTab(prompt_tab, "Prompts")

        # === Master Parameters Tab ===
        master_tab = QWidget()
        master_layout = QFormLayout(master_tab)
        master_params = self.config.get_master_params()

        self.m_max_tokens = QSpinBox()
        self.m_max_tokens.setRange(100, 100000)
        self.m_max_tokens.setValue(
            master_params.get('max_tokens', 4096))
        master_layout.addRow("Default Max Tokens:", self.m_max_tokens)

        self.m_temperature = QDoubleSpinBox()
        self.m_temperature.setRange(0.0, 2.0)
        self.m_temperature.setSingleStep(0.1)
        self.m_temperature.setValue(
            master_params.get('temperature', 0.3))
        master_layout.addRow("Default Temperature:", self.m_temperature)

        self.m_max_retries = QSpinBox()
        self.m_max_retries.setRange(1, 20)
        self.m_max_retries.setValue(
            master_params.get('max_retries', 5))
        master_layout.addRow("Max Retries:", self.m_max_retries)

        self.m_num_ctx = QSpinBox()
        self.m_num_ctx.setRange(2048, 131072)
        self.m_num_ctx.setValue(
            master_params.get('num_ctx', 32768))
        master_layout.addRow("Ollama Context Window:", self.m_num_ctx)

        tabs.addTab(master_tab, "Master Parameters")
        layout.addWidget(tabs)

        # Dialog buttons
        btn_layout = QHBoxLayout()
        apply_btn = QPushButton("Apply && Close")
        apply_btn.clicked.connect(self._apply_and_close)
        btn_layout.addWidget(apply_btn)
        cancel_btn = QPushButton("Cancel")
        cancel_btn.clicked.connect(self.reject)
        btn_layout.addWidget(cancel_btn)
        layout.addLayout(btn_layout)

    def _on_auto_newest_toggled(self, on: bool):
        """Re-select the newest model when auto mode is switched back on."""
        if on and self.claude_model_combo.count():
            self.claude_model_combo.setCurrentIndex(0)

    def _populate_claude_combo(self):
        """
        Rebuild the model dropdown from whatever the last fetch returned.

        When auto mode is on the newest entry wins, so a stale saved model
        is not carried forward. When the user has pinned a choice, that
        choice is always kept as an entry -- even if a fetch failed -- so
        it can never be silently blanked.
        """
        pinned = not self.claude_auto_newest.isChecked()
        current = ((self.claude_model_combo.currentText().strip()
                    or self.config.claude_model) if pinned else "")
        models = (self._claude_models_all if self.claude_show_old.isChecked()
                  else newest_per_family(self._claude_models_all))

        self.claude_model_combo.blockSignals(True)
        self.claude_model_combo.clear()
        for m in models:
            self.claude_model_combo.addItem(m['id'])
            self.claude_model_combo.setItemData(
                self.claude_model_combo.count() - 1,
                m['display_name'], Qt.ItemDataRole.ToolTipRole)

        if current:
            idx = self.claude_model_combo.findText(current)
            if idx < 0:
                self.claude_model_combo.insertItem(0, current)
                idx = 0
            self.claude_model_combo.setCurrentIndex(idx)
        elif self.claude_model_combo.count():
            self.claude_model_combo.setCurrentIndex(0)
        self.claude_model_combo.blockSignals(False)

    def _refresh_claude_models(self):
        """Pull the live model list using the key currently in the field."""
        self.claude_model_status.setText("Fetching models…")
        self.claude_refresh_btn.setEnabled(False)
        QApplication.setOverrideCursor(Qt.CursorShape.WaitCursor)
        QApplication.processEvents()
        try:
            models, error = self.config.fetch_claude_models(
                api_key=self.claude_key_edit.text())
        finally:
            QApplication.restoreOverrideCursor()
            self.claude_refresh_btn.setEnabled(True)

        if error:
            # Keep whatever the cache holds rather than emptying the list.
            self.claude_model_status.setText(
                f"⚠ {error}"
                + (" — showing last known models"
                   if self._claude_models_all else ""))
            self._populate_claude_combo()
            return

        self._claude_models_all = models
        self._populate_claude_combo()
        unknown = [m['id'] for m in models if not is_known_model(m['id'])]
        note = ""
        if unknown:
            note = (f" · {len(unknown)} newer than this build "
                    f"(generic parameters will be used)")
        self.claude_model_status.setText(
            f"{len(models)} models available · showing "
            f"{self.claude_model_combo.count()}{note}")

    def _refresh_ollama_models(self):
        models = self.api.list_ollama_models()
        self.ollama_model_combo.clear()
        for m in models:
            self.ollama_model_combo.addItem(m)
        if models:
            self.config.ollama_models = models

    def _add_ollama_model(self):
        name = self.new_model_edit.text().strip()
        if name:
            self.ollama_model_combo.addItem(name)
            models = self.config.ollama_models
            if name not in models:
                models.append(name)
                self.config.ollama_models = models
            self.new_model_edit.clear()

    def _test_connection(self):
        self._apply_api_settings()
        result = self.api.test_connection()
        self.test_label.setText(result)

    def _on_prompt_selected(self, row):
        if row < 0:
            return
        key = self.config.get_all_prompt_keys()[row]
        self.prompt_key_label.setText(f"<b>{key}</b>")
        self.prompt_text_edit.setPlainText(self.config.get_prompt(key))
        params = self.config.get_params_for_prompt(key)
        self.p_max_tokens.setValue(params.get('max_tokens', 4096))
        self.p_temperature.setValue(params.get('temperature', 0.3))

    def _save_current_prompt(self):
        row = self.prompt_list.currentRow()
        if row < 0:
            return
        key = self.config.get_all_prompt_keys()[row]
        self.config.set_custom_prompt(
            key, self.prompt_text_edit.toPlainText())
        self.config.set_params_for_prompt(key, {
            'max_tokens': self.p_max_tokens.value(),
            'temperature': self.p_temperature.value(),
        })
        self.prompt_list.item(row).setText(f"★ {key}")

    def _reset_current_prompt(self):
        row = self.prompt_list.currentRow()
        if row < 0:
            return
        key = self.config.get_all_prompt_keys()[row]
        self.config.reset_prompt(key)
        self.config.reset_params_for_prompt(key)
        from default_prompts import DEFAULT_PROMPTS
        self.prompt_text_edit.setPlainText(
            DEFAULT_PROMPTS.get(key, ""))
        self.prompt_list.item(row).setText(key)

    def _apply_api_settings(self):
        self.config.active_api = self.api_combo.currentText()
        self.config.claude_api_key = self.claude_key_edit.text()
        self.config.set_claude_model(
            self.claude_model_combo.currentText(),
            pinned=not self.claude_auto_newest.isChecked())
        self.config.show_model_notice = \
            self.startup_notice_check.isChecked()
        self.config.ollama_url = self.ollama_url_edit.text()
        self.config.active_ollama_model = \
            self.ollama_model_combo.currentText()
        self.config.ncbi_api_key = self.ncbi_key_edit.text()
        self.config.institutional_credentials = {
            'username': self.inst_user_edit.text(),
            'password': self.inst_pass_edit.text(),
        }
        self.config.set_master_params({
            'max_tokens': self.m_max_tokens.value(),
            'temperature': self.m_temperature.value(),
            'max_retries': self.m_max_retries.value(),
            'num_ctx': self.m_num_ctx.value(),
        })

    def _apply_and_close(self):
        self._apply_api_settings()
        self.accept()


# ═══════════════════════════════════════════════════════════════════
#  Find Dialog  (Ctrl+F search for any QTextEdit)
# ═══════════════════════════════════════════════════════════════════

class FindDialog(QDialog):
    """
    Modeless find dialog that searches within the currently focused
    QTextEdit. Stays open so the user can keep pressing Next/Previous
    while reading the document.
    """

    def __init__(self, parent=None):
        super().__init__(parent)
        self.setWindowTitle("Find")
        self.setModal(False)
        self.setFixedHeight(50)
        self.setMinimumWidth(420)
        self._target = None
        self._build_ui()

    def _build_ui(self):
        layout = QHBoxLayout(self)
        layout.setContentsMargins(8, 4, 8, 4)

        layout.addWidget(QLabel("Find:"))

        self.search_edit = QLineEdit()
        self.search_edit.setPlaceholderText("Search text…")
        self.search_edit.textChanged.connect(self._on_text_changed)
        self.search_edit.returnPressed.connect(self._find_next)
        layout.addWidget(self.search_edit, 1)

        self.count_label = QLabel("")
        self.count_label.setStyleSheet(
            "color: #666; min-width: 60px;")
        layout.addWidget(self.count_label)

        prev_btn = QPushButton("▲ Prev")
        prev_btn.setFixedWidth(65)
        prev_btn.clicked.connect(self._find_prev)
        prev_btn.setShortcut("Shift+Return")
        layout.addWidget(prev_btn)

        next_btn = QPushButton("▼ Next")
        next_btn.setFixedWidth(65)
        next_btn.clicked.connect(self._find_next)
        layout.addWidget(next_btn)

        close_btn = QPushButton("✕")
        close_btn.setFixedWidth(28)
        close_btn.clicked.connect(self._close_and_clear)
        layout.addWidget(close_btn)

    def activate_for(self, text_edit):
        """Show the dialog and bind it to the given QTextEdit."""
        self._target = text_edit
        # If there's selected text in the editor, pre-fill the search
        cursor = text_edit.textCursor()
        if cursor.hasSelection():
            selected = cursor.selectedText()
            if len(selected) < 200:
                self.search_edit.setText(selected)
        self.show()
        self.raise_()
        self.search_edit.setFocus()
        self.search_edit.selectAll()

    def _on_text_changed(self, text):
        if not self._target or not text:
            self.count_label.setText("")
            self._clear_highlight()
            return
        self._highlight_all(text)
        # Move cursor to start and find first match
        cursor = self._target.textCursor()
        cursor.movePosition(QTextCursor.MoveOperation.Start)
        self._target.setTextCursor(cursor)
        self._find_next()

    def _find_next(self):
        if not self._target:
            return
        text = self.search_edit.text()
        if not text:
            return
        found = self._target.find(text)
        if not found:
            # Wrap around to beginning
            cursor = self._target.textCursor()
            cursor.movePosition(QTextCursor.MoveOperation.Start)
            self._target.setTextCursor(cursor)
            found = self._target.find(text)
        if not found:
            self.count_label.setText("0 matches")
            self.count_label.setStyleSheet(
                "color: #c0392b; min-width: 60px;")
        else:
            self.count_label.setStyleSheet(
                "color: #666; min-width: 60px;")

    def _find_prev(self):
        if not self._target:
            return
        text = self.search_edit.text()
        if not text:
            return
        found = self._target.find(
            text, QTextDocument.FindFlag.FindBackward)
        if not found:
            # Wrap around to end
            cursor = self._target.textCursor()
            cursor.movePosition(QTextCursor.MoveOperation.End)
            self._target.setTextCursor(cursor)
            found = self._target.find(
                text, QTextDocument.FindFlag.FindBackward)

    def _highlight_all(self, text):
        """Count matches and update label (highlighting is done by
        the cursor selection from find())."""
        if not self._target:
            return
        # Count occurrences
        content = self._target.toPlainText()
        count = content.lower().count(text.lower())
        self.count_label.setText(
            f"{count} match{'es' if count != 1 else ''}")

    def _clear_highlight(self):
        """Remove any selection in the target."""
        if self._target:
            cursor = self._target.textCursor()
            cursor.clearSelection()
            self._target.setTextCursor(cursor)

    def _close_and_clear(self):
        self._clear_highlight()
        self.hide()

    def keyPressEvent(self, event):
        if event.key() == Qt.Key.Key_Escape:
            self._close_and_clear()
        else:
            super().keyPressEvent(event)
```
