## Supplementary material for "In Search of Ethical Procedures for LLM-Assisted Systematic Review Production: A Proof-of-Concept Evaluation of Selected Review Components": source_data: export_manager.py

```
"""
Export manager for the Systematic Review Foundry.
Handles export to .docx, .pdf, and .tex formats,
plus CSV/XLSX import and export for sources, topics, and statistics.
"""
import re
import io
import csv
import unicodedata
from typing import Dict, Any, List

from models import ReviewSession


# ═══════════════════════════════════════════════════════════════════
#  CSV / XLSX Export
# ═══════════════════════════════════════════════════════════════════

_SOURCE_CSV_COLUMNS = [
    'number', 'title', 'abstract', 'pmid', 'pmc_id', 'citation',
    'doi', 'year', 'journal', 'full_text', 'summary', 'rating',
    'rate_explain', 'bias_assessment', 'bias_explanation',
]

_TOPIC_CSV_COLUMNS = [
    'topic_id', 'title', 'text', 'linked_sections',
]

_STAT_CSV_COLUMNS = [
    'stat_id', 'question', 'text_response', 'python_response',
    'linked_sections',
]


def _ensure_openpyxl():
    """Import openpyxl or raise a helpful error."""
    try:
        import openpyxl
        return openpyxl
    except ImportError:
        raise ImportError(
            "openpyxl is required for spreadsheet export. "
            "Install with: pip install openpyxl")


def _sanitize_cell_value(val):
    """Convert a value to a clean string suitable for a spreadsheet cell.
    Returns the original type for numbers/bools but cleans strings."""
    if val is None:
        return ''
    if isinstance(val, bool):
        return val
    if isinstance(val, (int, float)):
        return val
    s = str(val)
    # Remove null bytes and control chars (except tab) that corrupt cells
    s = ''.join(ch for ch in s if ch == '\t' or ch == '\n' or
                (ord(ch) >= 32) or ord(ch) > 127)
    return s


def export_sources_spreadsheet(session: ReviewSession, filepath: str):
    """Export all source data to an XLSX spreadsheet."""
    openpyxl = _ensure_openpyxl()
    wb = openpyxl.Workbook()
    ws = wb.active
    ws.title = "Sources"

    # Header
    ws.append(_SOURCE_CSV_COLUMNS)

    for s in session.sources:
        src = s if isinstance(s, dict) else s
        row = []
        for col in _SOURCE_CSV_COLUMNS:
            val = src.get(col)
            if col == 'rating':
                if val is True:
                    row.append('TRUE')
                elif val is False:
                    row.append('FALSE')
                else:
                    row.append('')
            else:
                row.append(_sanitize_cell_value(val))
        ws.append(row)

    # Auto-size the narrower columns (skip large text fields)
    _auto_size_columns(ws, skip_wide={'abstract', 'full_text', 'summary',
                                       'citation', 'rate_explain',
                                       'bias_explanation'})
    wb.save(filepath)


def export_topics_spreadsheet(session: ReviewSession, filepath: str):
    """Export all topics to an XLSX spreadsheet."""
    openpyxl = _ensure_openpyxl()
    wb = openpyxl.Workbook()
    ws = wb.active
    ws.title = "Topics"

    ws.append(_TOPIC_CSV_COLUMNS)

    for t in session.topics:
        top = t if isinstance(t, dict) else t
        row = []
        for col in _TOPIC_CSV_COLUMNS:
            if col == 'linked_sections':
                ls = top.get('linked_sections', [])
                row.append(';'.join(ls) if ls else '')
            else:
                row.append(_sanitize_cell_value(top.get(col)))
        ws.append(row)

    _auto_size_columns(ws, skip_wide={'text'})
    wb.save(filepath)


def export_stats_spreadsheet(session: ReviewSession, filepath: str):
    """Export all statistics to an XLSX spreadsheet."""
    openpyxl = _ensure_openpyxl()
    wb = openpyxl.Workbook()
    ws = wb.active
    ws.title = "Statistics"

    ws.append(_STAT_CSV_COLUMNS)

    for st in session.statistics:
        stat = st if isinstance(st, dict) else st
        row = []
        for col in _STAT_CSV_COLUMNS:
            if col == 'linked_sections':
                ls = stat.get('linked_sections', [])
                row.append(';'.join(ls) if ls else '')
            else:
                row.append(_sanitize_cell_value(stat.get(col)))
        ws.append(row)

    _auto_size_columns(ws, skip_wide={'text_response', 'python_response'})
    wb.save(filepath)


def _auto_size_columns(ws, skip_wide=None, max_width=50):
    """Auto-size columns, capping wide text fields."""
    skip_wide = skip_wide or set()
    from openpyxl.utils import get_column_letter
    for col_idx, cell in enumerate(ws[1], 1):
        col_name = cell.value or ''
        if col_name in skip_wide:
            ws.column_dimensions[get_column_letter(col_idx)].width = 30
        else:
            # Measure from first ~20 rows
            max_len = len(str(col_name))
            for row in ws.iter_rows(min_row=2, max_row=min(20, ws.max_row),
                                     min_col=col_idx, max_col=col_idx):
                for c in row:
                    if c.value:
                        max_len = max(max_len, min(len(str(c.value)), max_width))
            ws.column_dimensions[get_column_letter(col_idx)].width = \
                min(max_len + 2, max_width)


# ═══════════════════════════════════════════════════════════════════
#  CSV / XLSX Import
# ═══════════════════════════════════════════════════════════════════

def _read_tabular_file(filepath: str) -> List[Dict]:
    """Read a CSV or XLSX file and return a list of row dicts."""
    if filepath.lower().endswith(('.xlsx', '.xls')):
        try:
            import openpyxl
        except ImportError:
            raise ImportError(
                "openpyxl is required to import XLSX files. "
                "Install with: pip install openpyxl")
        wb = openpyxl.load_workbook(filepath, read_only=True,
                                     data_only=True)
        ws = wb.active
        rows_iter = ws.iter_rows(values_only=True)
        headers = [str(h).strip() if h else '' for h in next(rows_iter)]
        records = []
        for row_vals in rows_iter:
            row_dict = {}
            for col, val in zip(headers, row_vals):
                if col:
                    row_dict[col] = val if val is not None else ''
            records.append(row_dict)
        wb.close()
        return records
    else:
        # CSV
        with open(filepath, 'r', newline='', encoding='utf-8') as f:
            reader = csv.DictReader(f)
            return list(reader)


def import_sources_csv(filepath: str) -> List[Dict]:
    """Import sources from a CSV or XLSX file. Returns list of source dicts."""
    records = _read_tabular_file(filepath)
    sources = []
    for i, row in enumerate(records):
        source = {}
        for col in _SOURCE_CSV_COLUMNS:
            val = row.get(col, '')
            if val is None:
                val = ''
            source[col] = str(val) if val != '' else None

        # Parse number
        num_str = source.get('number')
        try:
            source['number'] = int(num_str) if num_str else i + 1
        except (ValueError, TypeError):
            source['number'] = i + 1

        # Parse boolean rating
        rating_str = str(source.get('rating', '') or '').strip().lower()
        if rating_str == 'true':
            source['rating'] = True
        elif rating_str == 'false':
            source['rating'] = False
        else:
            source['rating'] = None

        # Ensure empty strings become None for optional text fields
        for field in ('abstract', 'full_text', 'summary', 'citation',
                      'doi', 'pmid', 'pmc_id', 'rate_explain',
                      'bias_assessment', 'bias_explanation'):
            if not source.get(field):
                source[field] = None

        sources.append(source)
    return sources


def import_topics_csv(filepath: str) -> List[Dict]:
    """Import topics from a CSV or XLSX file. Returns list of topic dicts."""
    records = _read_tabular_file(filepath)
    topics = []
    for i, row in enumerate(records):
        topic = {}
        for col in _TOPIC_CSV_COLUMNS:
            val = row.get(col, '')
            if val is None:
                val = ''
            topic[col] = str(val) if val != '' else None

        if not topic.get('topic_id'):
            topic['topic_id'] = f"T{i + 1}"
        if not topic.get('title'):
            topic['title'] = 'Untitled'

        # Parse linked_sections from semicolon-separated string
        ls_str = topic.get('linked_sections') or ''
        topic['linked_sections'] = (
            [s.strip() for s in ls_str.split(';') if s.strip()]
            if ls_str else [])
        topic['context_config'] = None
        topics.append(topic)
    return topics


def import_stats_csv(filepath: str) -> List[Dict]:
    """Import statistics from a CSV or XLSX file. Returns list of stat dicts."""
    records = _read_tabular_file(filepath)
    stats = []
    for i, row in enumerate(records):
        stat = {}
        for col in _STAT_CSV_COLUMNS:
            val = row.get(col, '')
            if val is None:
                val = ''
            stat[col] = str(val) if val != '' else None

        if not stat.get('stat_id'):
            stat['stat_id'] = f"S{i + 1}"
        if not stat.get('question'):
            stat['question'] = 'Untitled'

        ls_str = stat.get('linked_sections') or ''
        stat['linked_sections'] = (
            [s.strip() for s in ls_str.split(';') if s.strip()]
            if ls_str else [])
        stat['context_config'] = None
        stats.append(stat)
    return stats


def sanitize_for_latex(text: str) -> str:
    """Make a string block compatible with LaTeX."""
    replacements = [
        ('\\', r'\textbackslash{}'),
        ('&', r'\&'), ('%', r'\%'), ('$', r'\$'), ('#', r'\#'),
        ('_', r'\_'), ('{', r'\{'), ('}', r'\}'),
        ('~', r'\textasciitilde{}'), ('^', r'\textasciicircum{}'),
        ('≥', r'$\geq$'), ('≤', r'$\leq$'),
        ('>', r'$>$'), ('<', r'$<$'),
    ]

    def replace_unescaped(text, char, replacement):
        result = ""
        i = 0
        while i < len(text):
            if text[i:i+len(char)] == char:
                if i == 0 or text[i-1] != '\\':
                    result += replacement
                    i += len(char)
                else:
                    result += char
                    i += len(char)
            else:
                result += text[i]
                i += 1
        return result

    def replace_greek(text):
        result = ""
        for char in text:
            try:
                name = unicodedata.name(char, '')
                if name.startswith('GREEK SMALL LETTER'):
                    latex_name = name.split()[-1].lower()
                    result += f'$\\{latex_name}$'
                elif name.startswith('GREEK CAPITAL LETTER'):
                    latex_name = name.split()[-1].lower()
                    result += f'$\\{latex_name.capitalize()}$'
                else:
                    result += char
            except ValueError:
                result += char
        return result

    for char, replacement in replacements:
        text = replace_unescaped(text, char, replacement)
    text = replace_greek(text)
    return text


def export_to_latex(session: ReviewSession, filepath: str, title: str = "Systematic Review"):
    """Export the review session to a .tex file."""
    parts = []
    parts.append("\\documentclass[12pt,a4paper]{article}")
    parts.append("\\usepackage[utf8]{inputenc}")
    parts.append("\\usepackage{hyperref}")
    parts.append("\\usepackage{geometry}")
    parts.append("\\geometry{margin=1in}")
    parts.append("\\usepackage{lmodern}")
    parts.append("\\begin{document}")
    parts.append(f"\\title{{{sanitize_for_latex(title)}}}")
    parts.append("\\author{Systematic Review Team}")
    parts.append("\\maketitle")

    if session.abstract:
        parts.append("\\section{Abstract}")
        parts.append(sanitize_for_latex(session.abstract))

    if session.intro:
        parts.append("\\section{Introduction}")
        parts.append(sanitize_for_latex(session.intro))

    if session.methods:
        parts.append("\\section{Methods}")
        parts.append(sanitize_for_latex(session.methods))

    if session.results:
        parts.append("\\section{Results}")
        for rs in session.results:
            sec_title = rs.get('section', 'Untitled')
            sec_title = re.sub(r'^\d+\.\s*', '', sec_title)
            text = rs.get('text', '')
            if text:
                parts.append(f"\\subsection{{{sanitize_for_latex(sec_title)}}}")
                parts.append(sanitize_for_latex(text))

    if session.discussion:
        parts.append("\\section{Discussion}")
        parts.append(sanitize_for_latex(session.discussion))

    if session.conclusion:
        parts.append("\\section{Conclusion}")
        parts.append(sanitize_for_latex(session.conclusion))

    if session.citations:
        parts.append("\\section{References}")
        parts.append(sanitize_for_latex(session.citations))

    parts.append("\\end{document}")

    with io.open(filepath, 'w', encoding='utf-8') as f:
        f.write("\n\n".join(parts))


def export_to_docx(session: ReviewSession, filepath: str, title: str = "Systematic Review"):
    """Export the review session to a .docx file."""
    try:
        from docx import Document
        from docx.shared import Pt, Inches
        from docx.enum.text import WD_ALIGN_PARAGRAPH
    except ImportError:
        raise ImportError("python-docx is required for DOCX export. Install with: pip install python-docx")

    doc = Document()

    # Title
    title_para = doc.add_heading(title, level=0)
    title_para.alignment = WD_ALIGN_PARAGRAPH.CENTER

    sections = [
        ("Abstract", session.abstract),
        ("Introduction", session.intro),
        ("Methods", session.methods),
    ]

    for heading, content in sections:
        if content:
            doc.add_heading(heading, level=1)
            doc.add_paragraph(content)

    if session.results:
        doc.add_heading("Results", level=1)
        for rs in session.results:
            sec_title = rs.get('section', 'Untitled')
            sec_title = re.sub(r'^\d+\.\s*', '', sec_title)
            text = rs.get('text', '')
            if text:
                doc.add_heading(sec_title, level=2)
                doc.add_paragraph(text)

    more_sections = [
        ("Discussion", session.discussion),
        ("Conclusion", session.conclusion),
        ("References", session.citations),
    ]

    for heading, content in more_sections:
        if content:
            doc.add_heading(heading, level=1)
            doc.add_paragraph(content)

    doc.save(filepath)


def export_to_pdf(session: ReviewSession, filepath: str, title: str = "Systematic Review"):
    """Export via generating LaTeX then compiling, or use reportlab as fallback."""
    try:
        from reportlab.lib.pagesizes import letter
        from reportlab.lib.styles import getSampleStyleSheet, ParagraphStyle
        from reportlab.platypus import SimpleDocTemplate, Paragraph, Spacer
        from reportlab.lib.units import inch
    except ImportError:
        raise ImportError("reportlab is required for PDF export. Install with: pip install reportlab")

    doc = SimpleDocTemplate(filepath, pagesize=letter,
                            topMargin=inch, bottomMargin=inch,
                            leftMargin=inch, rightMargin=inch)
    styles = getSampleStyleSheet()

    title_style = ParagraphStyle('ReviewTitle', parent=styles['Title'], fontSize=18, spaceAfter=20)
    heading_style = ParagraphStyle('ReviewHeading', parent=styles['Heading1'], fontSize=14, spaceAfter=10)
    subheading_style = ParagraphStyle('ReviewSubheading', parent=styles['Heading2'], fontSize=12, spaceAfter=8)
    body_style = ParagraphStyle('ReviewBody', parent=styles['Normal'], fontSize=11, spaceAfter=6, leading=14)

    story = []
    story.append(Paragraph(title, title_style))
    story.append(Spacer(1, 0.3 * inch))

    def add_section(heading, content):
        if content:
            story.append(Paragraph(heading, heading_style))
            for para in content.split('\n'):
                if para.strip():
                    # Escape XML special chars for reportlab
                    safe = para.replace('&', '&amp;').replace('<', '&lt;').replace('>', '&gt;')
                    story.append(Paragraph(safe, body_style))
            story.append(Spacer(1, 0.15 * inch))

    add_section("Abstract", session.abstract)
    add_section("Introduction", session.intro)
    add_section("Methods", session.methods)

    if session.results:
        story.append(Paragraph("Results", heading_style))
        for rs in session.results:
            sec_title = rs.get('section', 'Untitled')
            sec_title = re.sub(r'^\d+\.\s*', '', sec_title)
            text = rs.get('text', '')
            if text:
                story.append(Paragraph(sec_title, subheading_style))
                for para in text.split('\n'):
                    if para.strip():
                        safe = para.replace('&', '&amp;').replace('<', '&lt;').replace('>', '&gt;')
                        story.append(Paragraph(safe, body_style))
        story.append(Spacer(1, 0.15 * inch))

    add_section("Discussion", session.discussion)
    add_section("Conclusion", session.conclusion)
    add_section("References", session.citations)

    doc.build(story)
```
