## Supplementary material for "In Search of Ethical Procedures for LLM-Assisted Systematic Review Production: A Proof-of-Concept Evaluation of Selected Review Components": source_data: main_app.py

```
"""
Systematic Review Foundry — Main Application Window.
QMainWindow with detachable/dockable tabs, File menu (Save/Save As/Load/Export),
Prompt Settings menu, auto-save timer, and session management.
"""
import sys
import os
from pathlib import Path

from PyQt6.QtWidgets import (
    QApplication, QMainWindow, QTabWidget, QTabBar, QMenuBar, QMenu,
    QFileDialog, QMessageBox, QStatusBar, QWidget, QVBoxLayout,
    QDockWidget, QStyle, QTextEdit, QDialog
)
from PyQt6.QtCore import Qt, QTimer, QByteArray
from PyQt6.QtGui import QAction, QKeySequence, QIcon, QShortcut

from config_manager import ConfigManager, get_default_session_dir
from api_manager import APIManager
from models import ReviewSession
from tabs.main_tab import MainReviewTab
from tabs.sources_tab import SourcesTab
from tabs.topics_stats_tab import TopicsTab, StatsTab
from dialogs import PromptSettingsDialog, FindDialog
from export_manager import (
    export_to_latex, export_to_docx, export_to_pdf,
    export_sources_spreadsheet, export_topics_spreadsheet,
    export_stats_spreadsheet,
    import_sources_csv, import_topics_csv, import_stats_csv,
)
from source_import import ImportSourcesDialog


class DetachableTabWidget(QTabWidget):
    """A QTabWidget that supports popping tabs out into floating windows
    and restoring them back into the tab bar."""

    def __init__(self, parent=None):
        super().__init__(parent)
        self.setTabsClosable(False)
        self.setMovable(True)
        self._detached_windows: dict[str, QDockWidget] = {}

    def detach_tab(self, index: int):
        widget = self.widget(index)
        title = self.tabText(index)

        if title in self._detached_windows:
            self._detached_windows[title].raise_()
            self._detached_windows[title].activateWindow()
            return

        self.removeTab(index)

        dock = QDockWidget(title, self.parent())
        dock.setWidget(widget)
        dock.setFeatures(
            QDockWidget.DockWidgetFeature.DockWidgetClosable |
            QDockWidget.DockWidgetFeature.DockWidgetMovable |
            QDockWidget.DockWidgetFeature.DockWidgetFloatable
        )
        dock.setFloating(True)
        dock.resize(900, 700)

        def _on_dock_close(event, t=title, w=widget, d=dock):
            self._restore_tab(t, w, d)
            event.accept()

        dock.closeEvent = _on_dock_close

        dock.show()
        self._detached_windows[title] = dock

    def _restore_tab(self, title: str, widget: QWidget, dock: QDockWidget):
        dock.setWidget(None)
        self.addTab(widget, title)
        self._detached_windows.pop(title, None)

    def reattach_all(self):
        for title in list(self._detached_windows.keys()):
            dock = self._detached_windows[title]
            dock.close()


class MainWindow(QMainWindow):
    """Main application window for Systematic Review Foundry."""

    APP_TITLE = "ReviewFoundry"

    def __init__(self):
        super().__init__()

        # Core managers
        self.config = ConfigManager()
        self.api = APIManager(self.config)
        self.session = ReviewSession()

        self._load_initial_session()
        self._dirty = False

        self._build_menus()
        self._build_tabs()
        self._build_statusbar()

        # Auto-save timer
        self._auto_save_timer = QTimer(self)
        self._auto_save_timer.timeout.connect(self._auto_save)
        interval_ms = self.config.auto_save_interval * 1000
        self._auto_save_timer.start(interval_ms)

        # Global Ctrl+F find dialog
        self._find_dialog = FindDialog(self)
        find_shortcut = QShortcut(
            QKeySequence("Ctrl+F"), self,
            context=Qt.ShortcutContext.ApplicationShortcut)
        find_shortcut.activated.connect(self._show_find)

        self.setWindowTitle(self._window_title())
        self.resize(1280, 860)

        geom = self.config._config.get("window_geometry")
        if geom:
            try:
                self.restoreGeometry(QByteArray.fromHex(geom.encode()))
            except Exception:
                pass


    def _window_title(self) -> str:
        path = self.config.active_session_path
        name = Path(path).stem if path else "untitled"
        dirty_marker = " *" if self._dirty else ""
        return f"{name}{dirty_marker} — {self.APP_TITLE}"
 
    def _mark_dirty(self):
        self._dirty = True
        self.setWindowTitle(self._window_title())

    # ── Menu bar ─────────────────────────────────────────────────────

    def _build_menus(self):
        menubar = self.menuBar()

        # File menu
        file_menu = menubar.addMenu("&File")

        new_action = QAction("&New Project", self)
        new_action.setShortcut(QKeySequence("Ctrl+N"))
        new_action.triggered.connect(self._new_project)
        file_menu.addAction(new_action)

        save_action = QAction("&Save", self)
        save_action.setShortcut(QKeySequence.StandardKey.Save)
        save_action.triggered.connect(self._save)
        file_menu.addAction(save_action)

        save_as_action = QAction("Save &As…", self)
        save_as_action.setShortcut(QKeySequence("Ctrl+Shift+S"))
        save_as_action.triggered.connect(self._save_as)
        file_menu.addAction(save_as_action)

        load_action = QAction("&Load…", self)
        load_action.setShortcut(QKeySequence("Ctrl+O"))
        load_action.triggered.connect(self._load)
        file_menu.addAction(load_action)

        file_menu.addSeparator()

        export_menu = file_menu.addMenu("&Export")
        for fmt, label in [("docx", "Word Document (.docx)"),
                           ("pdf", "PDF (.pdf)"),
                           ("tex", "LaTeX (.tex)")]:
            act = QAction(label, self)
            act.triggered.connect(lambda checked, f=fmt: self._export(f))
            export_menu.addAction(act)

        # Spreadsheet export submenu
        xlsx_export_menu = file_menu.addMenu("Export &Spreadsheet")
        for data_type, label in [
            ("sources", "Sources (.xlsx)"),
            ("topics", "Topics (.xlsx)"),
            ("statistics", "Statistics (.xlsx)"),
        ]:
            act = QAction(label, self)
            act.triggered.connect(
                lambda checked, dt=data_type: self._export_xlsx(dt))
            xlsx_export_menu.addAction(act)

        # Source import (RIS / CSV / XLSX with auto-detect or column mapper)
        import_sources_action = QAction("&Import Sources…", self)
        import_sources_action.triggered.connect(self._import_sources)
        file_menu.addAction(import_sources_action)

        # CSV/XLSX import submenu for topics and statistics
        csv_import_menu = file_menu.addMenu("&Import CSV/XLSX")
        for data_type, label in [
            ("topics", "Topics"),
            ("statistics", "Statistics"),
        ]:
            act = QAction(label, self)
            act.triggered.connect(
                lambda checked, dt=data_type: self._import_csv(dt))
            csv_import_menu.addAction(act)

        file_menu.addSeparator()

        quit_action = QAction("&Quit", self)
        quit_action.setShortcut(QKeySequence("Ctrl+Q"))
        quit_action.triggered.connect(self.close)
        file_menu.addAction(quit_action)

        # Prompt Settings menu
        settings_menu = menubar.addMenu("&Prompt Settings")
        open_settings_action = QAction("Open Prompt &Manager…", self)
        open_settings_action.triggered.connect(self._open_prompt_settings)
        settings_menu.addAction(open_settings_action)

        # View menu
        view_menu = menubar.addMenu("&View")
        self._pop_actions = {}
        for tab_name in ["Main Review", "Sources", "Topics",
                         "Statistics (Beta)"]:
            act = QAction(f"Pop Out: {tab_name}", self)
            act.triggered.connect(
                lambda checked, n=tab_name: self._pop_out_tab_by_name(n))
            view_menu.addAction(act)
            self._pop_actions[tab_name] = act

        view_menu.addSeparator()
        reattach_action = QAction("Reattach All Tabs", self)
        reattach_action.triggered.connect(self._reattach_all)
        view_menu.addAction(reattach_action)

    # ── Tabs ─────────────────────────────────────────────────────────

    def _build_tabs(self):
        self.tab_widget = DetachableTabWidget(self)

        self.main_tab = MainReviewTab(
            self.config, self.api, self.session, self)
        self.sources_tab = SourcesTab(
            self.config, self.api, self.session, self)
        self.topics_tab = TopicsTab(
            self.config, self.api, self.session, self)
        self.stats_tab = StatsTab(
            self.config, self.api, self.session, self)

        self.tab_widget.addTab(self.main_tab, "Main Review")
        self.tab_widget.addTab(self.sources_tab, "Sources")
        self.tab_widget.addTab(self.topics_tab, "Topics")
        self.tab_widget.addTab(self.stats_tab, "Statistics (Beta)")

        self.tab_widget.tabBar().tabBarDoubleClicked.connect(
            self._on_tab_double_click)

        self.setCentralWidget(self.tab_widget)

        # Inter-tab wiring
        self.main_tab.section_changed.connect(self._mark_dirty)
        self.main_tab.request_sources_pop.connect(
            self._pop_sources_for_citations)
        self.main_tab.request_navigate_to_source.connect(
            self._navigate_to_source)
        try:
            self.sources_tab.sources_changed.connect(self._mark_dirty)
        except AttributeError:
            pass

    def _on_tab_double_click(self, index):
        if index >= 0:
            self.tab_widget.detach_tab(index)

    def _pop_out_tab_by_name(self, name: str):
        for i in range(self.tab_widget.count()):
            if self.tab_widget.tabText(i) == name:
                self.tab_widget.detach_tab(i)
                return

    def _reattach_all(self):
        self.tab_widget.reattach_all()

    def _pop_sources_for_citations(self, numbers: list):
        for i in range(self.tab_widget.count()):
            if self.tab_widget.tabText(i) == "Sources":
                self.tab_widget.detach_tab(i)
                break
        try:
            self.sources_tab.show_sources_for_citations(numbers)
        except AttributeError:
            pass

    def _navigate_to_source(self, source_number: int, excerpt: str,
                            detail_tab_index: int):
        """Pop out the Sources tab and navigate to the matched excerpt."""
        # Pop out Sources tab if not already detached
        for i in range(self.tab_widget.count()):
            if self.tab_widget.tabText(i) == "Sources":
                self.tab_widget.detach_tab(i)
                break
        # If already detached, bring it to front
        dock = self.tab_widget._detached_windows.get("Sources")
        if dock:
            dock.raise_()
            dock.activateWindow()
        self.sources_tab.navigate_to_source_excerpt(
            source_number, excerpt, detail_tab_index)

    # ── Status bar ───────────────────────────────────────────────────

    def _build_statusbar(self):
        self.statusbar = QStatusBar(self)
        self.setStatusBar(self.statusbar)
        self._update_status()

    def _update_status(self):
        n_sources = len(self.session.sources)
        n_topics = len(self.session.topics)
        api = self.config.active_api.capitalize()
        path = Path(self.config.active_session_path).name
        self.statusbar.showMessage(
            f"Session: {path}  |  Sources: {n_sources}  |  "
            f"Topics: {n_topics}  |  API: {api}")

    # ── Session I/O ──────────────────────────────────────────────────

    def _load_initial_session(self):
        path = self.config.active_session_path
        if path and Path(path).exists():
            try:
                self.session = ReviewSession.load_from_file(path)
            except Exception:
                self.session = ReviewSession()
        else:
            self.session = ReviewSession()

    def _new_project(self):
        """Create a fresh, empty project."""
        if self._dirty:
            reply = QMessageBox.question(
                self, "Unsaved Changes",
                "You have unsaved changes. Save before creating "
                "a new project?",
                QMessageBox.StandardButton.Yes |
                QMessageBox.StandardButton.No |
                QMessageBox.StandardButton.Cancel)
            if reply == QMessageBox.StandardButton.Cancel:
                return
            if reply == QMessageBox.StandardButton.Yes:
                self._save()

        default_dir = str(get_default_session_dir())
        path, _ = QFileDialog.getSaveFileName(
            self, "New Project — Choose Location", default_dir,
            "JSON Files (*.json);;All Files (*)")
        if not path:
            return
        if not path.endswith('.json'):
            path += '.json'

        # Create a blank session, write it to disk, and switch to it
        self.session = ReviewSession()
        try:
            Path(path).parent.mkdir(parents=True, exist_ok=True)
            self.session.save_to_file(path)
            self.config.active_session_path = path
            self._refresh_all_tabs()
            self._dirty = False
            self.setWindowTitle(self._window_title())
            self._update_status()
            self.statusbar.showMessage(f"New project created: {path}", 5000)
        except Exception as e:
            QMessageBox.critical(self, "Error", str(e))

    def _save(self):
        path = self.config.active_session_path
        if not path:
            return self._save_as()
        Path(path).parent.mkdir(parents=True, exist_ok=True)
        try:
            self.session.save_to_file(path)
            self._dirty = False
            self.setWindowTitle(self._window_title())
            self.statusbar.showMessage(f"Saved to {path}", 3000)
        except Exception as e:
            QMessageBox.critical(self, "Save Error", str(e))

    def _save_as(self):
        default_dir = str(get_default_session_dir())
        path, _ = QFileDialog.getSaveFileName(
            self, "Save Session As", default_dir,
            "JSON Files (*.json);;All Files (*)")
        if path:
            if not path.endswith('.json'):
                path += '.json'
            self.config.active_session_path = path
            self._save()

    def _load(self):
        if self._dirty:
            reply = QMessageBox.question(
                self, "Unsaved Changes",
                "You have unsaved changes. Save before loading?",
                QMessageBox.StandardButton.Yes |
                QMessageBox.StandardButton.No |
                QMessageBox.StandardButton.Cancel)
            if reply == QMessageBox.StandardButton.Cancel:
                return
            if reply == QMessageBox.StandardButton.Yes:
                self._save()

        default_dir = str(get_default_session_dir())
        path, _ = QFileDialog.getOpenFileName(
            self, "Load Session", default_dir,
            "JSON Files (*.json);;All Files (*)")
        if path:
            try:
                self.session = ReviewSession.load_from_file(path)
                self.config.active_session_path = path
                self._refresh_all_tabs()
                self._dirty = False
                self.setWindowTitle(self._window_title())
                self._update_status()
                self.statusbar.showMessage(f"Loaded {path}", 3000)
            except Exception as e:
                QMessageBox.critical(self, "Load Error", str(e))

    def _refresh_all_tabs(self):
        self.main_tab.session = self.session
        self.sources_tab.session = self.session
        self.topics_tab.session = self.session
        self.stats_tab.session = self.session

        self.main_tab.refresh_from_session()
        for tab in (self.sources_tab, self.topics_tab, self.stats_tab):
            try:
                tab.refresh_from_session()
            except AttributeError:
                pass

    # ── Export (FIX: correct reorder_citations_for_export signature) ─

    def _check_citation_validations(self) -> bool:
        """Check whether all citations are approved.

        Returns True if the export should proceed (all approved, or user
        chose to continue anyway).  Returns False if the user cancelled.
        """
        try:
            from citation_manager import extract_bracketed_numbers
        except ImportError:
            return True  # can't check — allow export

        # Gather every citation number that appears in the review text
        all_text = ""
        if self.session.intro:
            all_text += self.session.intro + "\n"
        for rs in self.session.results:
            all_text += rs.get('text', '') + "\n"
        if self.session.discussion:
            all_text += self.session.discussion + "\n"
        if self.session.conclusion:
            all_text += self.session.conclusion + "\n"

        all_numbers = sorted(set(extract_bracketed_numbers(all_text)))
        if not all_numbers:
            return True  # nothing to validate

        validations = self.session.citation_validations
        disapproved = []
        unevaluated = []
        for n in all_numbers:
            entry = validations.get(str(n))
            if entry is None:
                unevaluated.append(n)
            elif entry.get('status') != 'approved':
                disapproved.append(n)

        if not disapproved and not unevaluated:
            return True  # all approved — proceed

        # Build a descriptive warning message
        parts = []
        if disapproved:
            parts.append(
                f"  • {len(disapproved)} disapproved: "
                f"[{', '.join(str(n) for n in disapproved)}]")
        if unevaluated:
            parts.append(
                f"  • {len(unevaluated)} unevaluated: "
                f"[{', '.join(str(n) for n in unevaluated)}]")
        detail = "\n".join(parts)

        total_issues = len(disapproved) + len(unevaluated)
        msg = QMessageBox(self)
        msg.setIcon(QMessageBox.Icon.Warning)
        msg.setWindowTitle("Unapproved Citations")
        msg.setText(
            f"{total_issues} of {len(all_numbers)} citation(s) have "
            f"not been approved:")
        msg.setInformativeText(
            f"{detail}\n\n"
            "Do you want to export anyway, or go back and "
            "review them first?")
        export_btn = msg.addButton(
            "Export Anyway", QMessageBox.ButtonRole.AcceptRole)
        msg.addButton(
            "Go Back", QMessageBox.ButtonRole.RejectRole)
        msg.setDefaultButton(export_btn)
        msg.exec()

        return msg.clickedButton() == export_btn

    def _export(self, fmt: str):
        # ── Citation-approval gate ──────────────────────────────────
        if not self._check_citation_validations():
            return

        ext_map = {"docx": "Word (*.docx)",
                   "pdf": "PDF (*.pdf)",
                   "tex": "LaTeX (*.tex)"}
        default_name = Path(self.config.active_session_path).stem or "review"
        path, _ = QFileDialog.getSaveFileName(
            self, f"Export as .{fmt}",
            str(Path.home() / f"{default_name}.{fmt}"),
            f"{ext_map.get(fmt, '')};;All Files (*)")
        if not path:
            return

        try:
            from citation_manager import reorder_citations_for_export

            # reorder_citations_for_export takes a ReviewSession
            # and returns a ReviewSession with renumbered citations
            export_session = reorder_citations_for_export(self.session)

            title = self.session.paper_topic or "Systematic Review"

            if fmt == "tex":
                export_to_latex(export_session, path, title)
            elif fmt == "docx":
                export_to_docx(export_session, path, title)
            elif fmt == "pdf":
                export_to_pdf(export_session, path, title)

            self.statusbar.showMessage(f"Exported to {path}", 5000)
            QMessageBox.information(
                self, "Export Complete",
                f"Review exported to:\n{path}")
        except Exception as e:
            QMessageBox.critical(self, "Export Error", str(e))

    def _export_xlsx(self, data_type: str):
        """Export sources, topics, or statistics as XLSX spreadsheet."""
        default_name = Path(self.config.active_session_path).stem or "review"
        path, _ = QFileDialog.getSaveFileName(
            self, f"Export {data_type.capitalize()} as Spreadsheet",
            str(Path.home() / f"{default_name}_{data_type}.xlsx"),
            "Excel Files (*.xlsx);;All Files (*)")
        if not path:
            return
        if not path.lower().endswith('.xlsx'):
            path += '.xlsx'
        try:
            if data_type == "sources":
                export_sources_spreadsheet(self.session, path)
            elif data_type == "topics":
                export_topics_spreadsheet(self.session, path)
            elif data_type == "statistics":
                export_stats_spreadsheet(self.session, path)
            self.statusbar.showMessage(
                f"Exported {data_type} to {path}", 5000)
            QMessageBox.information(
                self, "Export Complete",
                f"{data_type.capitalize()} exported to:\n{path}")
        except Exception as e:
            QMessageBox.critical(self, "Export Error", str(e))

    def _import_sources(self):
        """Import sources via RIS, auto-mapped CSV/XLSX, or column mapper."""
        dlg = ImportSourcesDialog(self)
        if dlg.exec() != QDialog.DialogCode.Accepted:
            return

        imported = dlg.get_sources()
        if not imported:
            return

        count = len(imported)
        msg = QMessageBox(self)
        msg.setWindowTitle("Import Sources")
        msg.setText(f"Found {count} sources.\n\n"
                    f"Replace existing sources, or append to them?")
        replace_btn = msg.addButton(
            "Replace", QMessageBox.ButtonRole.YesRole)
        append_btn = msg.addButton(
            "Append", QMessageBox.ButtonRole.NoRole)
        msg.addButton("Cancel", QMessageBox.ButtonRole.RejectRole)
        msg.exec()

        clicked = msg.clickedButton()
        if clicked == replace_btn:
            self.session.sources = imported
        elif clicked == append_btn:
            max_num = max(
                (s.get('number', 0) if isinstance(s, dict) else 0
                 for s in self.session.sources), default=0)
            for s in imported:
                max_num += 1
                s['number'] = max_num
            self.session.sources.extend(imported)
        else:
            return

        self.sources_tab.refresh_from_session()
        self.sources_tab.sources_changed.emit()
        self._mark_dirty()
        self._update_status()
        self.statusbar.showMessage(
            f"Imported {count} sources", 5000)

    def _import_csv(self, data_type: str):
        """Import topics or statistics from CSV or XLSX."""
        path, _ = QFileDialog.getOpenFileName(
            self, f"Import {data_type.capitalize()}",
            str(Path.home()),
            "CSV/XLSX Files (*.csv *.xlsx *.xls);;All Files (*)")
        if not path:
            return
        try:
            if data_type == "topics":
                imported = import_topics_csv(path)
                count = len(imported)
                reply = QMessageBox.question(
                    self, "Import Topics",
                    f"Found {count} topics. Replace or append?",
                    QMessageBox.StandardButton.Yes |
                    QMessageBox.StandardButton.No |
                    QMessageBox.StandardButton.Cancel)
                if reply == QMessageBox.StandardButton.Cancel:
                    return
                if reply == QMessageBox.StandardButton.Yes:
                    self.session.topics = imported
                else:
                    # Renumber topic IDs
                    for t in imported:
                        t['topic_id'] = (
                            f"T{len(self.session.topics) + 1}")
                        self.session.topics.append(t)
                self.topics_tab.refresh_from_session()
                try:
                    self.topics_tab.topics_changed.emit()
                except AttributeError:
                    pass

            elif data_type == "statistics":
                imported = import_stats_csv(path)
                count = len(imported)
                reply = QMessageBox.question(
                    self, "Import Statistics",
                    f"Found {count} statistics. Replace or append?",
                    QMessageBox.StandardButton.Yes |
                    QMessageBox.StandardButton.No |
                    QMessageBox.StandardButton.Cancel)
                if reply == QMessageBox.StandardButton.Cancel:
                    return
                if reply == QMessageBox.StandardButton.Yes:
                    self.session.statistics = imported
                else:
                    for s in imported:
                        s['stat_id'] = (
                            f"S{len(self.session.statistics) + 1}")
                        self.session.statistics.append(s)
                self.stats_tab.refresh_from_session()
                try:
                    self.stats_tab.stats_changed.emit()
                except AttributeError:
                    pass

            self._mark_dirty()
            self._update_status()
            self.statusbar.showMessage(
                f"Imported {data_type} from {path}", 5000)
        except Exception as e:
            QMessageBox.critical(self, "Import Error", str(e))

    # ── Prompt Settings ──────────────────────────────────────────────

    def _open_prompt_settings(self):
        dlg = PromptSettingsDialog(self.config, self.api, self)
        dlg.exec()

    # ── Find (Ctrl+F) ───────────────────────────────────────────────

    def _show_find(self):
        """Open the find dialog for whichever QTextEdit has focus."""
        focused = QApplication.focusWidget()
        # Walk up the widget tree to find the nearest QTextEdit
        widget = focused
        while widget is not None:
            if isinstance(widget, QTextEdit):
                self._find_dialog.activate_for(widget)
                return
            widget = widget.parentWidget()

    # ── Auto-save ────────────────────────────────────────────────────

    def _auto_save(self):
        if self._dirty:
            path = self.config.active_session_path
            if path:
                try:
                    Path(path).parent.mkdir(parents=True, exist_ok=True)
                    self.session.save_to_file(path)
                    self._dirty = False
                    self.setWindowTitle(self._window_title())
                    self.statusbar.showMessage("Auto-saved", 2000)
                except Exception:
                    pass

    # ── Close event ──────────────────────────────────────────────────

    def closeEvent(self, event):
        self.tab_widget.reattach_all()

        if self._dirty:
            reply = QMessageBox.question(
                self, "Unsaved Changes", "Save before closing?",
                QMessageBox.StandardButton.Yes |
                QMessageBox.StandardButton.No |
                QMessageBox.StandardButton.Cancel)
            if reply == QMessageBox.StandardButton.Cancel:
                event.ignore()
                return
            if reply == QMessageBox.StandardButton.Yes:
                self._save()

        self.config._config["window_geometry"] = bytes(
            self.saveGeometry().toHex()).decode()
        self.config.save()
        event.accept()


def main():
    app = QApplication(sys.argv)
    app.setApplicationName("Systematic Review Foundry")
    app.setOrganizationName("SystematicReviewFoundry")
    app.setStyle("Fusion")
    window = MainWindow()
    window.show()
    from dialogs import show_model_notice
    show_model_notice(window.config, window)
    sys.exit(app.exec())


if __name__ == "__main__":
    main()
```
