## Supplementary material for "In Search of Ethical Procedures for LLM-Assisted Systematic Review Production: A Proof-of-Concept Evaluation of Selected Review Components": source_data: models.py

```
"""
Data models for the Systematic Review Foundry application.
"""
import re
import json
from dataclasses import dataclass, field, asdict
from typing import Optional, List, Dict, Any


@dataclass
class Source:
    """Represents a single literature source."""
    number: int = 0
    title: Optional[str] = None
    abstract: Optional[str] = None
    pmid: Optional[str] = None
    pmc_id: Optional[str] = None
    citation: Optional[str] = None
    doi: Optional[str] = None
    year: Optional[str] = None
    journal: Optional[str] = None
    full_text: Optional[str] = None
    cited_by: Optional[str] = None
    summary: Optional[str] = None
    rating: Optional[bool] = None
    rate_explain: Optional[str] = None
    section_rate: Optional[List[int]] = None
    topic_ratings: Optional[Dict[str, int]] = None  # topic_id -> rating
    stat_ratings: Optional[Dict[str, int]] = None   # stat_id -> rating
    bias_assessment: Optional[str] = None   # e.g. "Low", "Some concerns", "High"
    bias_explanation: Optional[str] = None  # detailed RoB reasoning

    def build_abtext(self) -> Optional[str]:
        if self.abstract is None and self.full_text is not None:
            return self.full_text
        elif self.abstract is not None and self.full_text is None:
            return self.abstract
        elif self.abstract is not None and self.full_text is not None:
            return f'Abstract: {self.abstract}\nFull Text: {self.full_text}'
        return None

    def build_abtitle(self) -> Optional[str]:
        if self.abstract is None and self.title is not None:
            return f'Title: {self.title}'
        elif self.abstract is not None and self.title is None:
            return f'Abstract: {self.abstract}'
        elif self.abstract is not None and self.title is not None:
            return f'Title: {self.title}\nAbstract: {self.abstract}'
        return None

    def to_dict(self) -> dict:
        d = asdict(self)
        return d

    @classmethod
    def from_dict(cls, d: dict) -> 'Source':
        return cls(**{k: v for k, v in d.items() if k in cls.__dataclass_fields__})


@dataclass
class ResultsSection:
    """A subsection within the results."""
    section_number: int = 0
    section: str = ""
    text: Optional[str] = None
    context_config: Optional[Dict[str, Any]] = None  # tracks what context was used

    def to_dict(self) -> dict:
        return asdict(self)

    @classmethod
    def from_dict(cls, d: dict) -> 'ResultsSection':
        return cls(**{k: v for k, v in d.items() if k in cls.__dataclass_fields__})


@dataclass
class Topic:
    """A research topic/question for evidence gathering."""
    topic_id: str = ""
    title: str = ""
    text: Optional[str] = None
    linked_sections: List[str] = field(default_factory=list)
    context_config: Optional[Dict[str, Any]] = None

    def to_dict(self) -> dict:
        return asdict(self)

    @classmethod
    def from_dict(cls, d: dict) -> 'Topic':
        return cls(**{k: v for k, v in d.items() if k in cls.__dataclass_fields__})


@dataclass
class Statistic:
    """A statistical question/data point (beta feature)."""
    stat_id: str = ""
    question: str = ""
    text_response: Optional[str] = None
    python_response: Optional[str] = None
    linked_sections: List[str] = field(default_factory=list)
    context_config: Optional[Dict[str, Any]] = None

    def to_dict(self) -> dict:
        return asdict(self)

    @classmethod
    def from_dict(cls, d: dict) -> 'Statistic':
        return cls(**{k: v for k, v in d.items() if k in cls.__dataclass_fields__})


@dataclass
class CitationValidation:
    """Tracks validation state of a citation reference."""
    citation_number: int = 0
    status: str = "unvalidated"  # "approved", "disapproved", "unvalidated"
    validation_method: Optional[str] = None  # "manual", "auto_detect", "ai_detect", "batch_ai"
    match_text: Optional[str] = None


@dataclass
class ReviewSession:
    """The master session object containing the entire review state."""
    paper_topic: str = ""
    thesis: Optional[str] = None

    # Review sections
    abstract: Optional[str] = None
    intro: Optional[str] = None
    methods: Optional[str] = None
    results: List[Dict] = field(default_factory=list)  # list of ResultsSection dicts
    discussion: Optional[str] = None
    conclusion: Optional[str] = None
    citations: Optional[str] = None
    ordinal: Dict[str, int] = field(default_factory=dict)

    # Sources
    sources: List[Dict] = field(default_factory=list)
    search_terms: List[str] = field(default_factory=list)
    inclusion_criteria: List[str] = field(default_factory=list)
    exclusion_criteria: List[str] = field(default_factory=list)

    # Topics & Stats
    topics: List[Dict] = field(default_factory=list)
    statistics: List[Dict] = field(default_factory=list)

    # Context configs per section
    section_contexts: Dict[str, Dict] = field(default_factory=dict)

    # Citation validations
    citation_validations: Dict[str, Dict] = field(default_factory=dict)

    # Metadata
    version: str = "1.0"

    def to_dict(self) -> dict:
        return asdict(self)

    @classmethod
    def from_dict(cls, d: dict) -> 'ReviewSession':
        return cls(**{k: v for k, v in d.items() if k in cls.__dataclass_fields__})

    def get_sources_as_objects(self) -> List[Source]:
        return [Source.from_dict(s) for s in self.sources]

    def get_results_as_objects(self) -> List[ResultsSection]:
        return [ResultsSection.from_dict(r) for r in self.results]

    def get_topics_as_objects(self) -> List[Topic]:
        return [Topic.from_dict(t) for t in self.topics]

    def get_stats_as_objects(self) -> List[Statistic]:
        return [Statistic.from_dict(s) for s in self.statistics]

    def build_review_string(self) -> str:
        parts = []
        if self.abstract:
            parts.append(f"ABSTRACT:\n{self.abstract}")
        if self.intro:
            parts.append(f"INTRO:\n{self.intro}")
        if self.methods:
            parts.append(f"METHODS:\n{self.methods}")
        if self.results:
            for rs in self.results:
                if rs.get('text'):
                    parts.append(f"RESULTS SECTION {rs.get('section_number', '?')} - {rs.get('section', '')}:\n{rs['text']}")
        if self.discussion:
            parts.append(f"DISCUSSION:\n{self.discussion}")
        if self.conclusion:
            parts.append(f"CONCLUSION:\n{self.conclusion}")
        return "\n".join(parts)

    def save_to_file(self, filepath: str):
        with open(filepath, 'w', encoding='utf-8') as f:
            json.dump(self.to_dict(), f, indent=2, ensure_ascii=False)

    @classmethod
    def load_from_file(cls, filepath: str) -> 'ReviewSession':
        with open(filepath, 'r', encoding='utf-8') as f:
            data = json.load(f)
        return cls.from_dict(data)
```
