## Supplementary material for "In Search of Ethical Procedures for LLM-Assisted Systematic Review Production: A Proof-of-Concept Evaluation of Selected Review Components": source_data: ncbi_retrieval.py

```
"""
NCBI PubMed source retrieval for the Systematic Review Foundry.
Adapted from the original sourcing.py with improvements.
"""
import requests
import xml.etree.ElementTree as ET
import unicodedata
from typing import List, Dict, Optional, Tuple
from time import sleep

from models import Source


BASE_URL = "https://eutils.ncbi.nlm.nih.gov/entrez/eutils/"


def normalize_doi(doi: Optional[str]) -> Optional[str]:
    return doi.lower().strip() if doi else None


# ── XML helpers ───────────────────────────────────────────────────


def _get_element_text(element) -> str:
    """Recursively get all text content from an XML element,
    including text inside nested inline tags (<i>, <sub>, etc.)."""
    texts = []
    if element.text:
        texts.append(element.text)
    for child in element:
        texts.append(_get_element_text(child))
        if child.tail:
            texts.append(child.tail)
    return ''.join(texts)


def _extract_abstract(parent) -> Optional[str]:
    """Extract abstract text from an element containing Abstract/AbstractText.

    Handles both plain abstracts (single <AbstractText>) and structured
    abstracts (multiple <AbstractText Label="..."> elements, e.g.
    Objective, Methods, Results, Conclusions).

    *parent* can be a PubmedArticle, MedlineCitation, or Article element —
    the descendant search (.//) works from any of these levels.
    """
    abstract_els = parent.findall(".//Abstract/AbstractText")
    if not abstract_els:
        return None
    parts = []
    for ab in abstract_els:
        text = _get_element_text(ab).strip()
        if text:
            label = ab.get("Label")
            if label:
                parts.append(f"{label}: {text}")
            else:
                parts.append(text)
    return "\n".join(parts) if parts else None


def _construct_citation(article, doi) -> Tuple[str, str, str]:
    """Build a formatted citation from a PubmedArticle XML element.

    Returns (citation_string, journal_name, year).
    """
    authors = article.findall(".//Author")
    author_list = []
    for author in authors:
        last = author.find("LastName")
        init = author.find("Initials")
        if last is not None and init is not None:
            author_list.append(f"{last.text} {init.text}")

    title_el = article.find(".//ArticleTitle")
    title = (_get_element_text(title_el).strip()
             if title_el is not None else "Unknown Title")

    journal_el = article.find(".//Journal/Title")
    journal = journal_el.text if journal_el is not None else "Unknown Journal"

    year_el = article.find(".//PubDate/Year")
    if year_el is not None and year_el.text:
        year = year_el.text
    else:
        medline_date = article.find(".//PubDate/MedlineDate")
        if medline_date is not None and medline_date.text:
            year = medline_date.text.strip()[:4]
        else:
            year = "Unknown Year"

    volume_el = article.find(".//Volume")
    volume = volume_el.text if volume_el is not None else ""
    issue_el = article.find(".//Issue")
    issue = issue_el.text if issue_el is not None else ""
    pages_el = article.find(".//MedlinePgn")
    pages = pages_el.text if pages_el is not None else ""

    if author_list:
        authors_str = ", ".join(author_list[:6])
        if len(author_list) > 6:
            authors_str += ", et al."
    else:
        authors_str = "Unknown Authors"

    citation = f"{authors_str}. {title}. {journal}. {year}"
    if volume:
        citation += f";{volume}"
    if issue:
        citation += f"({issue})"
    if pages:
        citation += f":{pages}"
    citation += "."
    if doi:
        citation += f" doi: {doi}."

    return citation, journal, year


# ── Search functions ──────────────────────────────────────────────


def search_pubmed(query: str, api_key: str, retmax: int = 100,
                  sort: str = 'relevance') -> List[Dict]:
    """Search PubMed by query string and return parsed article metadata."""
    sort_map = {
        'relevance': 'relevance',
        'pub_date': 'date',
        'first_author': 'author',
        'journal': 'journal'
    }
    if sort not in sort_map:
        sort = 'relevance'

    search_params = {
        "db": "pubmed", "term": query, "retmax": retmax,
        "usehistory": "y", "api_key": api_key, "sort": sort_map[sort]
    }

    try:
        response = requests.get(
            f"{BASE_URL}esearch.fcgi", params=search_params, timeout=30)
        response.raise_for_status()
        search_tree = ET.fromstring(response.content)

        web_env_el = search_tree.find("WebEnv")
        query_key_el = search_tree.find("QueryKey")
        if web_env_el is None or query_key_el is None:
            return []

        fetch_params = {
            "db": "pubmed", "query_key": query_key_el.text,
            "WebEnv": web_env_el.text, "retmax": retmax,
            "retmode": "xml", "api_key": api_key
        }
        response = requests.get(
            f"{BASE_URL}efetch.fcgi", params=fetch_params, timeout=60)
        response.raise_for_status()
        fetch_tree = ET.fromstring(response.content)
    except Exception as e:
        print(f"PubMed search error: {e}")
        return []

    results = []
    for article in fetch_tree.findall(".//PubmedArticle"):
        title_el = article.find(".//ArticleTitle")
        title = (_get_element_text(title_el).strip()
                 if title_el is not None else "No title")

        abstract_text = _extract_abstract(article)

        pmid_el = article.find(".//PMID")
        pmid = pmid_el.text if pmid_el is not None else None

        pmc_el = article.find(".//ArticleId[@IdType='pmc']")
        pmc_id = pmc_el.text if pmc_el is not None else None

        doi_el = article.find(".//ArticleId[@IdType='doi']")
        doi = doi_el.text if doi_el is not None else None

        citation, journal, year = _construct_citation(article, doi)

        results.append({
            "title": title, "abstract": abstract_text,
            "pmid": pmid, "pmc_id": pmc_id,
            "citation": citation, "doi": normalize_doi(doi),
            "year": year, "journal": journal,
            "full_text": None
        })

    return results


def search_by_doi(doi: str, api_key: str) -> List[str]:
    """Search PubMed by DOI and return matching PMID strings."""
    params = {
        "db": "pubmed",
        "term": f"{doi}[DOI]",
        "retmode": "json",
        "retmax": 1,
        "api_key": api_key,
    }
    try:
        response = requests.get(
            f"{BASE_URL}esearch.fcgi", params=params, timeout=30)
        response.raise_for_status()
        data = response.json()
        return data.get("esearchresult", {}).get("idlist", [])
    except Exception as e:
        print(f"DOI search error: {e}")
        return []


# ── Fetch functions ───────────────────────────────────────────────


def fetch_article_by_pmid(pmid: str, api_key: str) -> Dict:
    """Fetch article XML from PubMed by PMID and return a metadata dict.

    Returns a dict with keys: title, journal, year, abstract, doi, pmid,
    pmc_id, citation.  Returns an empty dict on failure.

    This consolidates the per-article parsing that was previously duplicated
    between search_pubmed (bulk retrieval) and SourceInfoFillWorker
    (single-DOI lookup).
    """
    params = {
        "db": "pubmed",
        "id": pmid,
        "rettype": "xml",
        "retmode": "xml",
        "api_key": api_key,
    }
    try:
        response = requests.get(
            f"{BASE_URL}efetch.fcgi", params=params, timeout=30)
        response.raise_for_status()
    except Exception as e:
        print(f"Article fetch error for PMID {pmid}: {e}")
        return {}

    root = ET.fromstring(response.content)
    article_node = root.find(".//PubmedArticle")
    if article_node is None:
        return {}

    medline = article_node.find("MedlineCitation")
    if medline is None:
        return {}
    article = medline.find("Article")
    if article is None:
        return {}

    fields: Dict = {}

    # Title — _get_element_text handles inline markup (<i>, <sub>, etc.)
    title_node = article.find("ArticleTitle")
    if title_node is not None:
        fields["title"] = _get_element_text(title_node).strip()

    # Journal
    journal_node = article.find("Journal/Title")
    if journal_node is not None and journal_node.text:
        fields["journal"] = journal_node.text.strip()

    # Year (with MedlineDate fallback for journals that lack a clean Year)
    year_node = article.find("Journal/JournalIssue/PubDate/Year")
    if year_node is not None and year_node.text:
        fields["year"] = year_node.text.strip()
    else:
        medline_date = article.find(
            "Journal/JournalIssue/PubDate/MedlineDate")
        if medline_date is not None and medline_date.text:
            fields["year"] = medline_date.text.strip()[:4]

    # Abstract — handles structured (multi-section) abstracts
    abstract = _extract_abstract(article)
    if abstract:
        fields["abstract"] = abstract

    # DOI — check ELocationID first, then ArticleIdList
    for eid in article.findall("ELocationID"):
        if eid.get("EIdType") == "doi" and eid.text:
            fields["doi"] = eid.text.strip()
            break
    if "doi" not in fields:
        pubmed_data = article_node.find("PubmedData")
        if pubmed_data is not None:
            for aid in pubmed_data.findall("ArticleIdList/ArticleId"):
                if aid.get("IdType") == "doi" and aid.text:
                    fields["doi"] = aid.text.strip()
                    break

    # PMID
    pmid_node = medline.find("PMID")
    if pmid_node is not None and pmid_node.text:
        fields["pmid"] = pmid_node.text.strip()

    # PMC ID — needed for full-text retrieval
    pubmed_data = article_node.find("PubmedData")
    if pubmed_data is not None:
        for aid in pubmed_data.findall("ArticleIdList/ArticleId"):
            if aid.get("IdType") == "pmc" and aid.text:
                fields["pmc_id"] = aid.text.strip()
                break

    # Citation string
    author_list = article.findall("AuthorList/Author")
    if author_list:
        names = []
        for au in author_list:
            last_n = au.find("LastName")
            init_n = au.find("Initials")
            last = (last_n.text.strip()
                    if last_n is not None and last_n.text else "")
            initials = (init_n.text.strip()
                        if init_n is not None and init_n.text else "")
            if last:
                names.append(f"{last} {initials}".strip())
        if names:
            j_short = article.find("Journal/ISOAbbreviation")
            j_abbrev = (j_short.text.strip()
                        if j_short is not None and j_short.text
                        else fields.get("journal", ""))
            year = fields.get("year", "")
            vol_n = article.find("Journal/JournalIssue/Volume")
            volume = (vol_n.text.strip()
                      if vol_n is not None and vol_n.text else "")
            iss_n = article.find("Journal/JournalIssue/Issue")
            issue = (f"({iss_n.text.strip()})"
                     if iss_n is not None and iss_n.text else "")
            pg_n = article.find("Pagination/MedlinePgn")
            pages = (pg_n.text.strip()
                     if pg_n is not None and pg_n.text else "")

            author_str = ", ".join(names[:6])
            if len(names) > 6:
                author_str += ", et al."
            cite_parts = [
                author_str + ".",
                fields.get("title", "") + ".",
            ]
            tail = f"{j_abbrev}. {year}"
            if volume:
                tail += f";{volume}{issue}"
            if pages:
                tail += f":{pages}"
            tail += "."
            if fields.get("doi"):
                tail += f" doi:{fields['doi']}"
            cite_parts.append(tail)
            fields["citation"] = " ".join(cite_parts)

    return fields


def get_full_text(pmc_id: str, api_key: str) -> Optional[str]:
    """Retrieve full article body text from PMC.

    Returns the body text only — the abstract is stored separately and
    is NOT duplicated here.  Returns None if no body content is available
    (e.g. the article is not open-access in PMC).
    """
    if not pmc_id:
        return None
    if not pmc_id.upper().startswith('PMC'):
        pmc_id = f'PMC{pmc_id}'

    params = {
        "db": "pmc", "id": pmc_id,
        "rettype": "xml", "retmode": "xml", "api_key": api_key
    }

    try:
        response = requests.get(
            f"{BASE_URL}efetch.fcgi", params=params, timeout=60)
        if response.status_code != 200:
            return None
        root = ET.fromstring(response.content)

        # Body is required — if the PMC record has no body (abstract-only),
        # return None so we don't duplicate the abstract field.
        body = root.find(".//body")
        if body is None:
            return None

        body_paragraphs = []
        for el in body.iter():
            if el.tag == "title":
                title_text = _get_element_text(el).strip()
                if title_text:
                    body_paragraphs.append(f"\n{title_text}\n")
            elif el.tag == "p":
                p_text = _get_element_text(el).strip()
                if p_text:
                    body_paragraphs.append(p_text)

        if not body_paragraphs:
            return None

        # Include the abstract at the top when body content is present
        parts = []
        abstract_paragraphs = []
        for p in root.findall(".//abstract//p"):
            text = _get_element_text(p).strip()
            if text:
                abstract_paragraphs.append(text)
        if abstract_paragraphs:
            parts.append("ABSTRACT:\n" + "\n".join(abstract_paragraphs))

        parts.append("MAIN TEXT:\n" + "\n\n".join(body_paragraphs))

        full = "\n\n".join(parts).strip()
        return full if full else None
    except Exception as e:
        print(f"Error retrieving full text for {pmc_id}: {e}")
        return None


# ── High-level retrieval ──────────────────────────────────────────


def retrieve_sources(api_key: str, search_terms: List[str],
                     retmax_per_term: int = 100,
                     per_term_limits: Optional[Dict[str, int]] = None,
                     progress_callback=None
                     ) -> Tuple[List[Source], List[int], List[int]]:
    """Retrieve sources from PubMed for all search terms.

    Deduplicates by DOI.  Returns (sources, textless_nums, abstractless_nums).
    """
    all_results = []
    for i, term in enumerate(search_terms):
        limit = retmax_per_term
        if per_term_limits and term in per_term_limits:
            limit = per_term_limits[term]
        results = search_pubmed(term, api_key, retmax=limit)
        all_results.extend(results)
        if progress_callback:
            progress_callback(
                f"Retrieved {len(results)} results for term "
                f"{i+1}/{len(search_terms)}")
        sleep(0.35)  # NCBI rate limit

    # Deduplicate and build Source objects
    sources = []
    seen_dois = set()
    num = 1
    textless = []
    abless = []

    for result in all_results:
        doi = result.get('doi')
        if doi and doi in seen_dois:
            continue
        if doi:
            seen_dois.add(doi)

        # Attempt full text retrieval
        full_text = None
        if result.get('pmc_id'):
            full_text = get_full_text(result['pmc_id'], api_key)
            sleep(0.35)

        source = Source(
            number=num,
            title=result.get('title'),
            abstract=result.get('abstract'),
            pmid=result.get('pmid'),
            pmc_id=result.get('pmc_id'),
            citation=result.get('citation'),
            doi=doi,
            year=result.get('year'),
            journal=result.get('journal'),
            full_text=full_text
        )
        sources.append(source)
        if source.full_text is None:
            textless.append(num)
        if source.abstract is None:
            abless.append(num)
        num += 1

        if progress_callback:
            progress_callback(f"Processed source {num-1} (DOI dedup active)")

    return sources, textless, abless
```
