## Supplementary material for "In Search of Ethical Procedures for LLM-Assisted Systematic Review Production: A Proof-of-Concept Evaluation of Selected Review Components": source_data: source_import.py

```
"""
Source Import Module for Systematic Review Foundry.

Provides three import pathways for bringing external database exports
into the application's source format:

  1. RIS import  — parses .ris files (universal across all academic databases)
  2. Auto-mapped CSV/XLSX import — recognises column names from Scopus,
     Web of Science, Embase, PubMed, CINAHL, and other databases
  3. Assisted column-mapper dialog — QDialog that lets users manually
     assign columns from any spreadsheet to the expected fields

All three produce the same List[Dict] output compatible with
ReviewSession.sources.
"""
import csv
import re
from pathlib import Path
from typing import Dict, List, Optional, Tuple

from PyQt6.QtWidgets import (
    QDialog, QVBoxLayout, QHBoxLayout, QLabel, QComboBox,
    QPushButton, QTableWidget, QTableWidgetItem, QHeaderView,
    QFileDialog, QMessageBox, QGroupBox, QScrollArea, QWidget,
    QSizePolicy, QAbstractItemView,
)
from PyQt6.QtCore import Qt
from PyQt6.QtGui import QFont


# ═══════════════════════════════════════════════════════════════════
#  Canonical columns (must match _SOURCE_CSV_COLUMNS in export_manager)
# ═══════════════════════════════════════════════════════════════════

SOURCE_COLUMNS = [
    'number', 'title', 'abstract', 'pmid', 'pmc_id', 'citation',
    'doi', 'year', 'journal', 'full_text', 'summary', 'rating',
    'rate_explain', 'bias_assessment', 'bias_explanation',
]

# Human-readable labels for the dialog
_COLUMN_LABELS = {
    'number':       'Number (#)',
    'title':        'Title',
    'abstract':     'Abstract',
    'pmid':         'PubMed ID',
    'pmc_id':       'PMC ID',
    'citation':     'Citation / Reference',
    'doi':          'DOI',
    'year':         'Year',
    'journal':      'Journal / Source',
    'full_text':    'Full Text',
    'summary':      'Summary',
    'rating':       'Rating (True/False)',
    'rate_explain':  'Rating Explanation',
    'bias_assessment': 'Bias Assessment',
    'bias_explanation': 'Bias Explanation',
    # Extra fields not stored directly but used to build citation
    'authors':      'Authors',
    'volume':       'Volume',
    'issue':        'Issue',
    'pages':        'Pages',
    'start_page':   'Start Page',
    'end_page':     'End Page',
}


# ═══════════════════════════════════════════════════════════════════
#  Column alias map — covers Scopus, WoS, Embase, PubMed, CINAHL
# ═══════════════════════════════════════════════════════════════════

# Keys are internal field names.  Values are lowercase aliases that
# incoming CSV/XLSX headers get matched against.
_COLUMN_ALIASES: Dict[str, List[str]] = {
    'title': [
        'title', 'article title', 'document title', 'ti',
        'primary title', 'item title',
    ],
    'abstract': [
        'abstract', 'ab', 'abstract note', 'n2',
    ],
    'authors': [
        'authors', 'author', 'author names', 'author full names',
        'author(s)', 'au', 'first author', 'book authors',
        'author name', 'author, year',
    ],
    'doi': [
        'doi', 'digital object identifier', 'di', 'doi link',
    ],
    'year': [
        'year', 'publication year', 'pub year', 'py',
        'publication date', 'date', 'year of publication',
        'date of publication',
    ],
    'journal': [
        'journal', 'source title', 'source', 'journal title',
        'so', 'publication name', 'journal/book', 'journal name',
        'abbreviated source title', 'journal abbreviation',
        'journal iso abbreviation', 'jf', 'jo', 'j2', 't2',
        'secondary title',
    ],
    'pmid': [
        'pmid', 'pubmed id', 'pubmed_id', 'pubmedid',
    ],
    'pmc_id': [
        'pmc id', 'pmcid', 'pmc', 'pmc_id',
    ],
    'citation': [
        'citation', 'reference', 'cited reference', 'full citation',
    ],
    'volume': [
        'volume', 'vol', 'vl',
    ],
    'issue': [
        'issue', 'issue number', 'is', 'number',
    ],
    'pages': [
        'pages', 'page range', 'art. no.', 'article number',
    ],
    'start_page': [
        'start page', 'page start', 'sp', 'beginning page',
    ],
    'end_page': [
        'end page', 'page end', 'ep', 'ending page',
    ],
    'number': [
        'number', '#', 'no.', 'num', 'record number',
    ],
    'full_text': [
        'full_text', 'full text', 'body', 'content',
    ],
    'summary': [
        'summary',
    ],
    'rating': [
        'rating', 'include', 'included', 'relevant',
    ],
    'rate_explain': [
        'rate_explain', 'rating explanation', 'exclusion reason',
        'notes', 'reason',
    ],
    'bias_assessment': [
        'bias_assessment', 'bias assessment', 'risk of bias', 'rob',
    ],
    'bias_explanation': [
        'bias_explanation', 'bias explanation', 'rob explanation',
    ],
}


# ═══════════════════════════════════════════════════════════════════
#  RIS tag → internal field mapping
# ═══════════════════════════════════════════════════════════════════

_RIS_TAG_MAP = {
    'TI': 'title',
    'T1': 'title',
    'AB': 'abstract',
    'N2': 'abstract',
    'AU': 'authors',     # repeatable
    'A1': 'authors',
    'PY': 'year',
    'Y1': 'year',
    'DA': 'year',
    'JO': 'journal',
    'JF': 'journal',
    'J2': 'journal',
    'T2': 'journal',
    'DO': 'doi',
    'VL': 'volume',
    'IS': 'issue',
    'SP': 'start_page',
    'EP': 'end_page',
    'AN': 'accession',   # may be PMID depending on DB field
    'DB': 'database',
    'UR': 'url',
    'L2': 'url',
    'KW': 'keywords',
    'SN': 'issn',
}

# Tags that can appear multiple times per record
_RIS_REPEATABLE = {'AU', 'A1', 'KW'}


# ═══════════════════════════════════════════════════════════════════
#  RIS parser
# ═══════════════════════════════════════════════════════════════════

def parse_ris_file(filepath: str) -> List[Dict]:
    """Parse a .ris file and return a list of raw record dicts.

    Repeatable fields (e.g. AU) are stored as lists.
    """
    records: List[Dict] = []
    current: Dict = {}

    with open(filepath, 'r', encoding='utf-8-sig') as f:
        for raw_line in f:
            line = raw_line.rstrip('\n\r')

            # RIS tag line: two uppercase letters, two spaces, hyphen, space
            match = re.match(r'^([A-Z][A-Z0-9])\s\s-\s?(.*)', line)
            if match:
                tag, value = match.group(1), match.group(2).strip()

                if tag == 'ER':
                    # End of record
                    if current:
                        records.append(current)
                    current = {}
                    continue

                if tag == 'TY':
                    current['_type'] = value
                    continue

                field = _RIS_TAG_MAP.get(tag)
                if not field:
                    continue

                if tag in _RIS_REPEATABLE:
                    current.setdefault(field, [])
                    current[field].append(value)
                else:
                    # First value wins (don't overwrite title with alt)
                    if field not in current:
                        current[field] = value

        # Handle file without trailing ER
        if current:
            records.append(current)

    return records


def _extract_year(raw: str) -> Optional[str]:
    """Pull a four-digit year from RIS date fields like '2024/03/15'."""
    m = re.search(r'(\d{4})', raw or '')
    return m.group(1) if m else None


def _build_citation(rec: Dict) -> str:
    """Build a formatted citation string from RIS fields."""
    parts = []

    # Authors
    authors = rec.get('authors', [])
    if isinstance(authors, list) and authors:
        if len(authors) <= 3:
            parts.append(', '.join(authors))
        else:
            parts.append(f"{authors[0]}, et al.")
    elif isinstance(authors, str) and authors:
        parts.append(authors)

    # Year
    year = _extract_year(rec.get('year', ''))
    if year:
        parts.append(f"({year})")

    # Title
    title = rec.get('title', '')
    if title:
        parts.append(f"{title}.")

    # Journal, Volume(Issue), Pages
    journal_parts = []
    journal = rec.get('journal', '')
    if journal:
        journal_parts.append(journal)

    vol = rec.get('volume', '')
    issue = rec.get('issue', '')
    if vol and issue:
        journal_parts.append(f"{vol}({issue})")
    elif vol:
        journal_parts.append(vol)

    sp = rec.get('start_page', '')
    ep = rec.get('end_page', '')
    if sp and ep:
        journal_parts.append(f"{sp}-{ep}")
    elif sp:
        journal_parts.append(sp)

    if journal_parts:
        parts.append(', '.join(journal_parts) + '.')

    # DOI
    doi = rec.get('doi', '')
    if doi:
        parts.append(f"doi:{doi}")

    return ' '.join(parts)


def _detect_pmid_from_ris(rec: Dict) -> Optional[str]:
    """Try to extract a PMID from the accession number or URL fields."""
    # Some databases put the PMID in the AN field
    accession = rec.get('accession', '')
    db = (rec.get('database', '') or '').lower()

    if db in ('pubmed', 'medline', 'nlm') and accession:
        return accession

    # Check if accession looks like a plain numeric PMID
    if accession and re.match(r'^\d{5,10}$', accession):
        return accession

    # Try to extract from URL
    url = rec.get('url', '')
    m = re.search(r'pubmed[./](\d+)', url or '')
    if m:
        return m.group(1)

    return None


def import_sources_ris(filepath: str) -> List[Dict]:
    """Import sources from a RIS file into the standard source dict format."""
    raw_records = parse_ris_file(filepath)
    sources = []

    for i, rec in enumerate(raw_records):
        source = {col: None for col in SOURCE_COLUMNS}
        source['number'] = i + 1
        source['title'] = rec.get('title')
        source['abstract'] = rec.get('abstract')
        source['year'] = _extract_year(rec.get('year', ''))
        source['journal'] = rec.get('journal')
        source['doi'] = rec.get('doi')
        source['pmid'] = _detect_pmid_from_ris(rec)
        source['citation'] = _build_citation(rec)
        source['rating'] = None
        source['rate_explain'] = None
        source['full_text'] = None
        source['summary'] = None
        source['pmc_id'] = None
        source['bias_assessment'] = None
        source['bias_explanation'] = None

        sources.append(source)

    return sources


# ═══════════════════════════════════════════════════════════════════
#  CSV / XLSX auto-mapped import
# ═══════════════════════════════════════════════════════════════════

def _normalize_header(h: str) -> str:
    """Lowercase, strip, collapse whitespace."""
    return re.sub(r'\s+', ' ', h.strip().lower())


def auto_map_columns(headers: List[str]) -> Dict[str, Optional[str]]:
    """Given a list of CSV/XLSX column headers, return a mapping of
    internal field name → original header name for every field that
    could be matched.  Returns None for unmatched fields.
    """
    mapping: Dict[str, Optional[str]] = {}
    normalised = {_normalize_header(h): h for h in headers}

    # All fields we might want to map (including auxiliary ones)
    all_fields = list(_COLUMN_ALIASES.keys())

    used_headers = set()

    for field in all_fields:
        aliases = _COLUMN_ALIASES[field]
        matched = None
        for alias in aliases:
            if alias in normalised and normalised[alias] not in used_headers:
                matched = normalised[alias]
                used_headers.add(matched)
                break
        mapping[field] = matched

    return mapping


def _build_citation_from_csv_row(
        row: Dict, mapping: Dict[str, Optional[str]]) -> Optional[str]:
    """Construct a citation string from auxiliary CSV columns."""
    def _get(field):
        col = mapping.get(field)
        if col is None:
            return ''
        val = row.get(col, '')
        return str(val).strip() if val else ''

    authors = _get('authors')
    year = _get('year')
    title = _get('title')
    journal = _get('journal')
    volume = _get('volume')
    issue = _get('issue')
    pages = _get('pages')
    sp = _get('start_page')
    ep = _get('end_page')
    doi = _get('doi')

    if not any([authors, title]):
        return None

    parts = []
    if authors:
        parts.append(authors)
    if year:
        # Extract just the year if it's a full date
        m = re.search(r'(\d{4})', year)
        parts.append(f"({m.group(1)})" if m else f"({year})")
    if title:
        parts.append(f"{title}.")

    journal_bits = []
    if journal:
        journal_bits.append(journal)
    if volume and issue:
        journal_bits.append(f"{volume}({issue})")
    elif volume:
        journal_bits.append(volume)

    if pages:
        journal_bits.append(pages)
    elif sp and ep:
        journal_bits.append(f"{sp}-{ep}")
    elif sp:
        journal_bits.append(sp)

    if journal_bits:
        parts.append(', '.join(journal_bits) + '.')
    if doi:
        clean_doi = doi.replace('https://doi.org/', '').replace(
            'http://doi.org/', '')
        parts.append(f"doi:{clean_doi}")

    return ' '.join(parts) if parts else None


def _read_tabular_file(filepath: str) -> Tuple[List[str], List[Dict]]:
    """Read a CSV or XLSX file. Returns (headers, rows).

    Unlike the version in export_manager, this returns raw headers
    separately so auto_map_columns can work on them.
    """
    if filepath.lower().endswith(('.xlsx', '.xls')):
        try:
            import openpyxl
        except ImportError:
            raise ImportError(
                "openpyxl is required to import XLSX files. "
                "Install with: pip install openpyxl")
        wb = openpyxl.load_workbook(filepath, read_only=True,
                                     data_only=True)
        ws = wb.active
        rows_iter = ws.iter_rows(values_only=True)
        headers = [str(h).strip() if h else '' for h in next(rows_iter)]
        records = []
        for row_vals in rows_iter:
            row_dict = {}
            for col, val in zip(headers, row_vals):
                if col:
                    row_dict[col] = val if val is not None else ''
            records.append(row_dict)
        wb.close()
        return headers, records
    else:
        with open(filepath, 'r', newline='', encoding='utf-8-sig') as f:
            reader = csv.DictReader(f)
            records = list(reader)
            headers = reader.fieldnames or []
            return list(headers), records


def import_sources_auto(filepath: str) -> Tuple[List[Dict], Dict[str, Optional[str]]]:
    """Auto-import sources from CSV/XLSX using alias-based column mapping.

    Returns (sources, mapping_used) so callers can inspect what was mapped.
    """
    headers, records = _read_tabular_file(filepath)
    mapping = auto_map_columns(headers)

    sources = []
    for i, row in enumerate(records):
        source = {col: None for col in SOURCE_COLUMNS}
        source['number'] = i + 1

        # Direct-mapped fields
        for field in SOURCE_COLUMNS:
            col = mapping.get(field)
            if col is not None:
                val = row.get(col, '')
                if val is None or str(val).strip() == '':
                    source[field] = None
                else:
                    source[field] = str(val).strip()

        # Parse number
        num_str = source.get('number')
        try:
            source['number'] = int(num_str) if num_str else i + 1
        except (ValueError, TypeError):
            source['number'] = i + 1

        # Extract year from date-like strings
        if source.get('year'):
            m = re.search(r'(\d{4})', str(source['year']))
            source['year'] = m.group(1) if m else source['year']

        # Build citation if none was directly mapped
        if not source.get('citation'):
            source['citation'] = _build_citation_from_csv_row(row, mapping)

        # Parse boolean rating
        rating_str = str(source.get('rating', '') or '').strip().lower()
        if rating_str in ('true', 'yes', '1', 'include', 'included'):
            source['rating'] = True
        elif rating_str in ('false', 'no', '0', 'exclude', 'excluded'):
            source['rating'] = False
        else:
            source['rating'] = None

        # Ensure empty strings become None
        for field in ('abstract', 'full_text', 'summary', 'citation',
                      'doi', 'pmid', 'pmc_id', 'rate_explain',
                      'bias_assessment', 'bias_explanation'):
            if not source.get(field):
                source[field] = None

        sources.append(source)

    return sources, mapping


# ═══════════════════════════════════════════════════════════════════
#  Column Mapper Dialog (assisted GUI import)
# ═══════════════════════════════════════════════════════════════════

class ColumnMapperDialog(QDialog):
    """A dialog that loads a CSV/XLSX file and lets the user map its
    columns to the application's expected source fields.

    Layout:
      - Top: file info and preview table (first 5 rows)
      - Middle: two-column mapping grid (their column → our field),
        pre-populated with auto-detected guesses
      - Bottom: OK / Cancel
    """

    # Fields the user can map to, in display order.
    # Includes auxiliary fields used for citation construction.
    MAPPABLE_FIELDS = [
        'title', 'abstract', 'authors', 'doi', 'year', 'journal',
        'pmid', 'pmc_id', 'citation', 'volume', 'issue', 'pages',
        'start_page', 'end_page', 'rating', 'rate_explain',
        'full_text', 'summary', 'bias_assessment', 'bias_explanation',
    ]

    def __init__(self, filepath: str, parent=None):
        super().__init__(parent)
        self.setWindowTitle("Column Mapper — Source Import")
        self.setMinimumSize(820, 620)
        self.resize(900, 700)

        self._filepath = filepath
        self._headers: List[str] = []
        self._records: List[Dict] = []
        self._combos: Dict[str, QComboBox] = {}
        self._result_sources: Optional[List[Dict]] = None

        self._load_file()
        self._build_ui()

    # ── File loading ────────────────────────────────────────────

    def _load_file(self):
        try:
            self._headers, self._records = _read_tabular_file(
                self._filepath)
        except Exception as e:
            QMessageBox.critical(self, "Import Error",
                                 f"Failed to read file:\n{e}")
            self._headers = []
            self._records = []

    # ── UI construction ─────────────────────────────────────────

    def _build_ui(self):
        layout = QVBoxLayout(self)

        # File info
        info_label = QLabel(
            f"<b>File:</b> {Path(self._filepath).name} — "
            f"{len(self._records)} records, "
            f"{len(self._headers)} columns detected")
        layout.addWidget(info_label)

        # Preview table
        preview_group = QGroupBox("Data Preview (first 5 rows)")
        preview_layout = QVBoxLayout(preview_group)
        self._preview_table = QTableWidget()
        self._preview_table.setEditTriggers(
            QAbstractItemView.EditTrigger.NoEditTriggers)
        self._preview_table.setMaximumHeight(180)
        self._populate_preview()
        preview_layout.addWidget(self._preview_table)
        layout.addWidget(preview_group)

        # Mapping section
        mapping_group = QGroupBox(
            "Column Mapping — assign your file's columns to source fields")
        mapping_outer = QVBoxLayout(mapping_group)

        scroll = QScrollArea()
        scroll.setWidgetResizable(True)
        scroll_content = QWidget()
        mapping_layout = QVBoxLayout(scroll_content)
        mapping_layout.setSpacing(6)

        # Header row
        header_row = QHBoxLayout()
        lbl_field = QLabel("Source Field")
        lbl_field.setFont(QFont(lbl_field.font().family(), -1, QFont.Weight.Bold))
        lbl_field.setFixedWidth(200)
        header_row.addWidget(lbl_field)
        lbl_col = QLabel("Your Column")
        lbl_col.setFont(QFont(lbl_col.font().family(), -1, QFont.Weight.Bold))
        header_row.addWidget(lbl_col)
        mapping_layout.addLayout(header_row)

        # Get auto-mapping suggestions
        auto_map = auto_map_columns(self._headers)

        for field in self.MAPPABLE_FIELDS:
            row_layout = QHBoxLayout()

            label_text = _COLUMN_LABELS.get(field, field)
            # Mark key fields
            if field in ('title',):
                label_text += '  ★'
            label = QLabel(label_text)
            label.setFixedWidth(200)
            row_layout.addWidget(label)

            combo = QComboBox()
            combo.addItem("— skip —", None)
            for h in self._headers:
                combo.addItem(h, h)
            combo.setSizePolicy(
                QSizePolicy.Policy.Expanding, QSizePolicy.Policy.Fixed)

            # Pre-select auto-detected match
            suggested = auto_map.get(field)
            if suggested:
                idx = combo.findData(suggested)
                if idx >= 0:
                    combo.setCurrentIndex(idx)

            self._combos[field] = combo
            row_layout.addWidget(combo)
            mapping_layout.addLayout(row_layout)

        mapping_layout.addStretch()
        scroll_content.setLayout(mapping_layout)
        scroll.setWidget(scroll_content)
        mapping_outer.addWidget(scroll)
        layout.addWidget(mapping_group)

        # Buttons
        btn_layout = QHBoxLayout()
        btn_layout.addStretch()

        reset_btn = QPushButton("Reset to Auto-Detect")
        reset_btn.clicked.connect(self._reset_to_auto)
        btn_layout.addWidget(reset_btn)

        cancel_btn = QPushButton("Cancel")
        cancel_btn.clicked.connect(self.reject)
        btn_layout.addWidget(cancel_btn)

        ok_btn = QPushButton("Import")
        ok_btn.setDefault(True)
        ok_btn.clicked.connect(self._do_import)
        btn_layout.addWidget(ok_btn)

        layout.addLayout(btn_layout)

    def _populate_preview(self):
        if not self._headers or not self._records:
            return

        preview_rows = self._records[:5]
        self._preview_table.setColumnCount(len(self._headers))
        self._preview_table.setRowCount(len(preview_rows))
        self._preview_table.setHorizontalHeaderLabels(self._headers)

        for r, row in enumerate(preview_rows):
            for c, h in enumerate(self._headers):
                val = str(row.get(h, ''))
                # Truncate long values for preview
                if len(val) > 80:
                    val = val[:77] + '...'
                item = QTableWidgetItem(val)
                self._preview_table.setItem(r, c, item)

        self._preview_table.horizontalHeader().setSectionResizeMode(
            QHeaderView.ResizeMode.ResizeToContents)

    def _reset_to_auto(self):
        auto_map = auto_map_columns(self._headers)
        for field, combo in self._combos.items():
            suggested = auto_map.get(field)
            if suggested:
                idx = combo.findData(suggested)
                combo.setCurrentIndex(idx if idx >= 0 else 0)
            else:
                combo.setCurrentIndex(0)

    # ── Import execution ────────────────────────────────────────

    def _do_import(self):
        # Build the user's mapping
        user_mapping: Dict[str, Optional[str]] = {}
        for field, combo in self._combos.items():
            user_mapping[field] = combo.currentData()

        # Check that at least title is mapped
        if not user_mapping.get('title'):
            QMessageBox.warning(
                self, "Missing Mapping",
                "Please map at least the 'Title' column before importing.")
            return

        sources = []
        for i, row in enumerate(self._records):
            source = {col: None for col in SOURCE_COLUMNS}
            source['number'] = i + 1

            # Apply the mapping
            for field in SOURCE_COLUMNS:
                col = user_mapping.get(field)
                if col is not None:
                    val = row.get(col, '')
                    if val is None or str(val).strip() == '':
                        source[field] = None
                    else:
                        source[field] = str(val).strip()

            # Parse number
            num_str = source.get('number')
            try:
                source['number'] = int(num_str) if num_str else i + 1
            except (ValueError, TypeError):
                source['number'] = i + 1

            # Extract year
            if source.get('year'):
                m = re.search(r'(\d{4})', str(source['year']))
                source['year'] = m.group(1) if m else source['year']

            # Build citation if not directly mapped
            if not source.get('citation'):
                source['citation'] = _build_citation_from_csv_row(
                    row, user_mapping)

            # Parse rating
            rating_str = str(
                source.get('rating', '') or '').strip().lower()
            if rating_str in ('true', 'yes', '1', 'include', 'included'):
                source['rating'] = True
            elif rating_str in (
                    'false', 'no', '0', 'exclude', 'excluded'):
                source['rating'] = False
            else:
                source['rating'] = None

            # Normalise empty → None
            for field in ('abstract', 'full_text', 'summary', 'citation',
                          'doi', 'pmid', 'pmc_id', 'rate_explain',
                          'bias_assessment', 'bias_explanation'):
                if not source.get(field):
                    source[field] = None

            sources.append(source)

        self._result_sources = sources
        self.accept()

    def get_sources(self) -> Optional[List[Dict]]:
        """Returns imported sources after dialog accepted, or None."""
        return self._result_sources


# ═══════════════════════════════════════════════════════════════════
#  Import method chooser dialog
# ═══════════════════════════════════════════════════════════════════

class ImportSourcesDialog(QDialog):
    """Top-level dialog that lets the user choose an import method:

      • Auto Import (RIS or CSV/XLSX with auto column detection)
      • Assisted Import (opens the column mapper dialog)

    After a successful import the caller retrieves sources via
    get_sources().
    """

    def __init__(self, parent=None):
        super().__init__(parent)
        self.setWindowTitle("Import Sources")
        self.setMinimumWidth(460)
        self._sources: Optional[List[Dict]] = None
        self._build_ui()

    def _build_ui(self):
        layout = QVBoxLayout(self)

        desc = QLabel(
            "Choose how to import sources from an external database export.\n\n"
            "Auto Import detects file format and column names automatically.\n"
            "Assisted Import lets you manually assign columns from your file.")
        desc.setWordWrap(True)
        layout.addWidget(desc)

        layout.addSpacing(12)

        # Auto import
        auto_btn = QPushButton("Auto Import (RIS / CSV / XLSX)")
        auto_btn.setMinimumHeight(38)
        auto_btn.setToolTip(
            "Automatically detect file format and map columns.\n"
            "Works with exports from PubMed, Scopus, Web of Science,\n"
            "Embase, CINAHL, Cochrane, and most other databases.")
        auto_btn.clicked.connect(self._auto_import)
        layout.addWidget(auto_btn)

        # Assisted import
        assist_btn = QPushButton("Assisted Import (Column Mapper)")
        assist_btn.setMinimumHeight(38)
        assist_btn.setToolTip(
            "Open a CSV or XLSX file and manually map its columns\n"
            "to source fields using a visual interface.")
        assist_btn.clicked.connect(self._assisted_import)
        layout.addWidget(assist_btn)

        layout.addSpacing(8)

        cancel_btn = QPushButton("Cancel")
        cancel_btn.clicked.connect(self.reject)
        layout.addWidget(cancel_btn)

    def _pick_file(self, filter_str: str) -> Optional[str]:
        path, _ = QFileDialog.getOpenFileName(
            self, "Select File to Import",
            str(Path.home()),
            filter_str)
        return path if path else None

    def _auto_import(self):
        path = self._pick_file(
            "All Supported (*.ris *.csv *.xlsx *.xls);;"
            "RIS Files (*.ris);;"
            "CSV Files (*.csv);;"
            "Excel Files (*.xlsx *.xls);;"
            "All Files (*)")
        if not path:
            return

        try:
            ext = Path(path).suffix.lower()
            if ext == '.ris':
                sources = import_sources_ris(path)
                method = "RIS"
            else:
                sources, mapping = import_sources_auto(path)
                method = "CSV/XLSX (auto-mapped)"

                # Report what was mapped
                mapped = [f for f in SOURCE_COLUMNS
                          if mapping.get(f) is not None]
                unmapped_important = [
                    f for f in ('title', 'abstract', 'doi', 'year',
                                'journal', 'pmid')
                    if mapping.get(f) is None]

                if not mapping.get('title'):
                    reply = QMessageBox.warning(
                        self, "No Title Column Detected",
                        "Could not auto-detect a 'Title' column in this "
                        "file. Would you like to use the Assisted Import "
                        "column mapper instead?",
                        QMessageBox.StandardButton.Yes |
                        QMessageBox.StandardButton.No)
                    if reply == QMessageBox.StandardButton.Yes:
                        return self._run_column_mapper(path)
                    # Otherwise proceed with what we have

            self._sources = sources
            QMessageBox.information(
                self, "Import Successful",
                f"Imported {len(sources)} sources via {method}.")
            self.accept()

        except Exception as e:
            QMessageBox.critical(
                self, "Import Error",
                f"Failed to import file:\n{e}")

    def _assisted_import(self):
        path = self._pick_file(
            "CSV/XLSX Files (*.csv *.xlsx *.xls);;All Files (*)")
        if not path:
            return
        self._run_column_mapper(path)

    def _run_column_mapper(self, path: str):
        dlg = ColumnMapperDialog(path, parent=self)
        if dlg.exec() == QDialog.DialogCode.Accepted:
            self._sources = dlg.get_sources()
            if self._sources is not None:
                QMessageBox.information(
                    self, "Import Successful",
                    f"Imported {len(self._sources)} sources via "
                    f"assisted column mapping.")
                self.accept()

    def get_sources(self) -> Optional[List[Dict]]:
        """Returns imported sources if dialog was accepted, else None."""
        return self._sources
```
