## Supplementary material for "In Search of Ethical Procedures for LLM-Assisted Systematic Review Production: A Proof-of-Concept Evaluation of Selected Review Components": source_data: __init__.py

```
"""
Tabs package for Systematic Review Foundry.
"""
from tabs.main_tab import MainReviewTab
from tabs.sources_tab import SourcesTab
from tabs.topics_stats_tab import TopicsTab, StatsTab

__all__ = ['MainReviewTab', 'SourcesTab', 'TopicsTab', 'StatsTab']
```
