## Supplementary material for "In Search of Ethical Procedures for LLM-Assisted Systematic Review Production: A Proof-of-Concept Evaluation of Selected Review Components": source_data: main_tab.py

```
"""
Main Review Tab — the primary text editing workspace.
Contains piecemeal section editors for abstract, intro, methods, results, 
discussion, conclusion, and citations. Supports AI writing, context 
configuration, citation validation, and text improvement.
"""
import re
import ast
from functools import partial

from PyQt6.QtWidgets import (
    QWidget, QVBoxLayout, QHBoxLayout, QScrollArea, QTextEdit, QLabel,
    QPushButton, QGroupBox, QMenu, QComboBox, QCheckBox, QLineEdit,
    QMessageBox, QInputDialog, QFrame, QSplitter, QToolBar, QSizePolicy,
    QDoubleSpinBox, QFormLayout, QDialog
)
from PyQt6.QtCore import Qt, pyqtSignal, QTimer
from PyQt6.QtGui import (
    QAction, QTextCursor, QTextCharFormat, QColor, QFont, QSyntaxHighlighter
)

from dialogs import (
    PromptApprovalDialog, OutputApprovalDialog, ContextConfigDialog
)


class CitationHighlighter(QSyntaxHighlighter):
    """Highlights [x] style citations in the text with color based on validation."""

    def __init__(self, parent, validation_data=None):
        super().__init__(parent)
        self.validation_data = validation_data or {}

    def highlightBlock(self, text):
        pattern = re.compile(r'\[\d+(?:\s*,\s*\d+)*\]')
        for match in pattern.finditer(text):
            fmt = QTextCharFormat()
            numbers = [int(n.strip()) for n in match.group(0).strip('[]').split(',')]
            statuses = [self.validation_data.get(str(n), {}).get('status', 'unvalidated')
                        for n in numbers]

            if all(s == 'approved' for s in statuses):
                fmt.setForeground(QColor("#2ecc71"))
            elif any(s == 'disapproved' for s in statuses):
                fmt.setForeground(QColor("#e74c3c"))
            else:
                fmt.setForeground(QColor("#3498db"))

            fmt.setFontWeight(QFont.Weight.Bold)
            fmt.setFontUnderline(True)
            self.setFormat(match.start(), match.end() - match.start(), fmt)


class SectionEditor(QWidget):
    """
    Editor widget for a single review section (e.g., Introduction).
    Uses a plain QWidget with a collapsible content area — avoids the
    QGroupBox.setCheckable crash entirely.
    """
    content_changed = pyqtSignal(str, str)

    def __init__(self, section_key: str, display_name: str,
                 parent_tab=None, auto_only: bool = False):
        super().__init__()
        self.section_key = section_key
        self.display_name = display_name
        self.parent_tab = parent_tab
        self._collapsed = False
        self._auto_only = auto_only
        self._build_ui()

    def _build_ui(self):
        outer = QVBoxLayout(self)
        outer.setContentsMargins(0, 0, 0, 6)

        # ── Header row ──
        header = QHBoxLayout()
        self.collapse_btn = QPushButton("▼")
        self.collapse_btn.setFixedSize(24, 24)
        self.collapse_btn.clicked.connect(self._toggle_collapse)
        header.addWidget(self.collapse_btn)

        header.addWidget(QLabel(f"<b>{self.display_name}</b>"))
        header.addStretch()

        self.write_btn = QPushButton("✦ AI Write")
        self.write_btn.setToolTip("Generate this section using AI")
        self.write_btn.clicked.connect(
            lambda: self.parent_tab.ai_write_section(self.section_key))
        header.addWidget(self.write_btn)

        self.context_btn = QPushButton("📋 Context")
        self.context_btn.setToolTip("Configure what context is sent to the AI")
        self.context_btn.clicked.connect(
            lambda: self.parent_tab.configure_context(self.section_key))
        header.addWidget(self.context_btn)

        # Hide AI buttons for auto-assembled sections (e.g. Citations)
        if self._auto_only:
            self.write_btn.setVisible(False)
            self.context_btn.setVisible(False)

        outer.addLayout(header)

        # ── Collapsible content ──
        self.content_widget = QWidget()
        cw_layout = QVBoxLayout(self.content_widget)
        cw_layout.setContentsMargins(4, 2, 4, 2)

        self.editor = QTextEdit()
        self.editor.setMinimumHeight(150)
        self.editor.setFont(QFont("Georgia", 11))
        self.editor.textChanged.connect(self._on_text_changed)
        self.editor.setContextMenuPolicy(Qt.ContextMenuPolicy.CustomContextMenu)
        self.editor.customContextMenuRequested.connect(self._context_menu)
        self.highlighter = CitationHighlighter(self.editor.document())
        cw_layout.addWidget(self.editor)

        outer.addWidget(self.content_widget)

        self.setStyleSheet(
            "SectionEditor { border: 1px solid #bbb; border-radius: 4px; "
            "padding: 4px; margin-bottom: 2px; }")

    # ── collapse helpers ──
    def _toggle_collapse(self):
        self._collapsed = not self._collapsed
        self.content_widget.setVisible(not self._collapsed)
        self.collapse_btn.setText("▶" if self._collapsed else "▼")

    def set_collapsed(self, collapsed: bool):
        self._collapsed = collapsed
        self.content_widget.setVisible(not collapsed)
        self.collapse_btn.setText("▶" if collapsed else "▼")

    # ── text helpers ──
    def _on_text_changed(self):
        self.content_changed.emit(self.section_key, self.editor.toPlainText())

    def set_text(self, text: str):
        self.editor.blockSignals(True)
        self.editor.setPlainText(text or "")
        self.editor.blockSignals(False)

    def get_text(self) -> str:
        return self.editor.toPlainText()

    def update_validation_data(self, data: dict):
        self.highlighter.validation_data = data
        self.highlighter.rehighlight()

    # ── context menu with citation actions ──
    def _context_menu(self, pos):
        menu = self.editor.createStandardContextMenu()
        menu.addSeparator()

        cursor = self.editor.textCursor()
        selected = cursor.selectedText()
        if selected:
            improve_action = QAction("✦ Improve Selected Text", self)
            improve_action.triggered.connect(
                lambda: self.parent_tab.improve_text(self.section_key, selected))
            menu.addAction(improve_action)

        # Citation detection at click position
        cursor_pos = self.editor.cursorForPosition(pos)
        block_text = cursor_pos.block().text()
        char_in_block = cursor_pos.positionInBlock()

        citation_match = None
        for m in re.finditer(r'\[\d+(?:\s*,\s*\d+)*\]', block_text):
            if m.start() <= char_in_block <= m.end():
                citation_match = m
                break

        if citation_match:
            numbers = [int(n.strip())
                       for n in citation_match.group(0).strip('[]').split(',')]
            menu.addSeparator()

            view_act = QAction(f"View Source(s) {citation_match.group(0)}", self)
            view_act.triggered.connect(
                lambda: self.parent_tab.view_citation_sources(numbers))
            menu.addAction(view_act)

            app_act = QAction("✓ Approve Citation", self)
            app_act.triggered.connect(
                lambda: self.parent_tab.validate_citation(numbers, "approved"))
            menu.addAction(app_act)

            dis_act = QAction("✗ Disapprove Citation", self)
            dis_act.triggered.connect(
                lambda: self.parent_tab.validate_citation(numbers, "disapproved"))
            menu.addAction(dis_act)

            auto_act = QAction("🔍 Auto-Detect (regex)", self)
            auto_act.triggered.connect(
                lambda: self.parent_tab.auto_detect_citation(
                    self.section_key, citation_match, numbers))
            menu.addAction(auto_act)

            ai_act = QAction("✦ AI-Detect", self)
            ai_act.triggered.connect(
                lambda: self.parent_tab.ai_detect_citation(
                    self.section_key, citation_match, numbers))
            menu.addAction(ai_act)

            swap_act = QAction("🔄 Find Best Source & Swap", self)
            swap_act.triggered.connect(
                lambda: self.parent_tab.swap_citation(
                    self.section_key, citation_match, numbers))
            menu.addAction(swap_act)

        menu.exec(self.editor.mapToGlobal(pos))


# ═══════════════════════════════════════════════════════════════════
#  Main Review Tab
# ═══════════════════════════════════════════════════════════════════

class MainReviewTab(QWidget):
    """The main review editing tab."""

    section_changed = pyqtSignal()
    request_sources_pop = pyqtSignal(list)
    request_navigate_to_source = pyqtSignal(int, str, int)  # source_number, excerpt, detail_tab_index

    def __init__(self, config_manager, api_manager, session, parent=None):
        super().__init__(parent)
        self.config = config_manager
        self.api = api_manager
        self.session = session
        self.section_editors = {}
        self._build_ui()
        self._load_from_session()

    # ── UI construction ──────────────────────────────────────────────

    def _build_ui(self):
        layout = QVBoxLayout(self)

        # Top toolbar
        top_bar = QHBoxLayout()
        top_bar.addWidget(QLabel("Paper Topic:"))
        self.topic_edit = QLineEdit()
        self.topic_edit.setPlaceholderText(
            "Enter the topic of your systematic review")
        self.topic_edit.textChanged.connect(self._on_topic_changed)
        top_bar.addWidget(self.topic_edit)

        self.citation_val_box = QComboBox()
        self.citation_val_box.addItems(['Validate Citations Using Summaries', 'Validate Citations Using Full-Texts'])
        self.citation_val_box.setCurrentIndex(0)
        top_bar.addWidget(self.citation_val_box)

        thesis_btn = QPushButton("Thesis")
        thesis_btn.clicked.connect(self._manage_thesis)
        top_bar.addWidget(thesis_btn)

        write_all_btn = QPushButton("✦ Write All Sections")
        write_all_btn.setToolTip(
            "Auto-write the entire review using current context")
        write_all_btn.clicked.connect(self._write_all)
        top_bar.addWidget(write_all_btn)

        validate_all_btn = QPushButton("Validate All Citations")
        validate_all_btn.clicked.connect(self._validate_all_citations)
        top_bar.addWidget(validate_all_btn)
        layout.addLayout(top_bar)

        # View controls
        view_bar = QHBoxLayout()
        view_bar.addWidget(QLabel("View:"))
        all_btn = QPushButton("All Sections")
        all_btn.clicked.connect(self._show_all_sections)
        view_bar.addWidget(all_btn)
        for name in ["Abstract", "Intro", "Methods", "Results",
                      "Discussion", "Conclusion", "Citations"]:
            btn = QPushButton(name)
            btn.clicked.connect(partial(self._show_single_section,
                                        name.lower()))
            view_bar.addWidget(btn)
        # Results subsection isolator
        self.results_sub_combo = QComboBox()
        self.results_sub_combo.setSizeAdjustPolicy(
            QComboBox.SizeAdjustPolicy.AdjustToContents)
        self.results_sub_combo.setMinimumWidth(140)
        self.results_sub_combo.setPlaceholderText("Results subsection…")
        view_bar.addWidget(self.results_sub_combo)
        self.results_sub_go_btn = QPushButton("Go")
        self.results_sub_go_btn.setToolTip(
            "Isolate the selected results subsection")
        self.results_sub_go_btn.clicked.connect(
            self._show_selected_results_subsection)
        view_bar.addWidget(self.results_sub_go_btn)

        view_bar.addStretch()
        layout.addLayout(view_bar)

        # Scrollable section area
        scroll = QScrollArea()
        scroll.setWidgetResizable(True)
        scroll.setHorizontalScrollBarPolicy(
            Qt.ScrollBarPolicy.ScrollBarAlwaysOff)

        self.sections_container = QWidget()
        self.sections_layout = QVBoxLayout(self.sections_container)

        for key, display in [("abstract", "Abstract"),
                              ("intro", "Introduction"),
                              ("methods", "Methods")]:
            ed = SectionEditor(key, display, parent_tab=self)
            ed.content_changed.connect(self._on_section_changed)
            self.section_editors[key] = ed
            self.sections_layout.addWidget(ed)

        # Results: header + dynamic sub-editors
        self.results_header_widget = QWidget()
        rh_layout = QHBoxLayout(self.results_header_widget)
        rh_layout.setContentsMargins(0, 8, 0, 0)
        rh_layout.addWidget(QLabel("<b>── Results ──</b>"))
        rh_layout.addStretch()
        gen_btn = QPushButton("✦ Generate Results Topics")
        gen_btn.clicked.connect(self._generate_results_topics)
        rh_layout.addWidget(gen_btn)
        add_btn = QPushButton("+ Add Subsection")
        add_btn.clicked.connect(self._add_results_subsection)
        rh_layout.addWidget(add_btn)
        rm_btn = QPushButton("- Remove Subsection")
        rm_btn.clicked.connect(self._remove_results_subsection)
        rh_layout.addWidget(rm_btn)
        self.sections_layout.addWidget(self.results_header_widget)

        self.results_editors_container = QWidget()
        self.results_editors_layout = QVBoxLayout(
            self.results_editors_container)
        self.results_editors_layout.setContentsMargins(20, 0, 0, 0)
        self.sections_layout.addWidget(self.results_editors_container)

        for key, display in [("discussion", "Discussion"),
                              ("conclusion", "Conclusion")]:
            ed = SectionEditor(key, display, parent_tab=self)
            ed.content_changed.connect(self._on_section_changed)
            self.section_editors[key] = ed
            self.sections_layout.addWidget(ed)

        # Citations section — auto-assembled, no AI Write / Context
        citations_ed = SectionEditor(
            "citations", "Citations (auto-assembled)",
            parent_tab=self, auto_only=True)
        citations_ed.editor.setReadOnly(True)
        citations_ed.editor.setStyleSheet(
            "QTextEdit { background-color: #f8f8f0; }")
        citations_ed.content_changed.connect(self._on_section_changed)
        self.section_editors["citations"] = citations_ed
        self.sections_layout.addWidget(citations_ed)

        # Spacer widget that can be hidden when a single section is isolated
        self._bottom_spacer = QWidget()
        self._bottom_spacer.setSizePolicy(
            QSizePolicy.Policy.Preferred, QSizePolicy.Policy.Expanding)
        self.sections_layout.addWidget(self._bottom_spacer)
        scroll.setWidget(self.sections_container)
        layout.addWidget(scroll)

    # ── Session I/O ──────────────────────────────────────────────────

    def _load_from_session(self):
        self.topic_edit.setText(self.session.paper_topic or "")
        for key in ["abstract", "intro", "methods",
                     "discussion", "conclusion"]:
            val = getattr(self.session, key, None)
            if key in self.section_editors:
                self.section_editors[key].set_text(val or "")
                self.section_editors[key].update_validation_data(
                    self.session.citation_validations)
        # Update citation validation highlighting on the citations editor
        if 'citations' in self.section_editors:
            self.section_editors['citations'].update_validation_data(
                self.session.citation_validations)
        self._rebuild_results_editors()
        self._rebuild_citations()

    def _rebuild_results_editors(self):
        for k in [k for k in self.section_editors if k.startswith('results_')]:
            ed = self.section_editors.pop(k)
            self.results_editors_layout.removeWidget(ed)
            ed.setParent(None)
            ed.deleteLater()

        for i, rs in enumerate(self.session.results):
            key = f"results_{i}"
            title = rs.get('section', f'Subsection {i+1}')
            ed = SectionEditor(key, f"Results: {title}", parent_tab=self)
            ed.content_changed.connect(self._on_section_changed)
            ed.set_text(rs.get('text', ''))
            ed.update_validation_data(self.session.citation_validations)
            self.section_editors[key] = ed
            self.results_editors_layout.addWidget(ed)

        self._update_results_sub_combo()

    def refresh_from_session(self):
        self._load_from_session()

    # ── Signals ──────────────────────────────────────────────────────

    def _on_topic_changed(self, text):
        self.session.paper_topic = text
        self.section_changed.emit()

    def _on_section_changed(self, section_key: str, text: str):
        if section_key.startswith('results_'):
            idx = int(section_key.split('_')[1])
            if idx < len(self.session.results):
                self.session.results[idx]['text'] = text
        else:
            setattr(self.session, section_key, text if text else None)
        # Auto-rebuild the citations list whenever any content section changes
        # (but not when citations itself changes, to avoid recursion)
        if section_key != 'citations':
            self._rebuild_citations()
        self.section_changed.emit()

    def _rebuild_citations(self):
        """
        Scan all review text for [N] style citations, look up each
        source's citation string, and auto-populate the Citations editor.
        Uses original source numbers (no reordering — that's export-only).
        """
        from citation_manager import extract_bracketed_numbers, \
            separate_bracketed_lists

        # Gather all text from content sections
        all_text = ""
        if self.session.intro:
            all_text += self.session.intro + "\n"
        for rs in self.session.results:
            if rs.get('text'):
                all_text += rs['text'] + "\n"
        if self.session.discussion:
            all_text += self.session.discussion + "\n"
        if self.session.conclusion:
            all_text += self.session.conclusion + "\n"

        # Separate combined citations ([3,5] → [3][5]) then extract numbers
        all_text_sep = separate_bracketed_lists(all_text)
        cited_numbers = sorted(set(extract_bracketed_numbers(all_text_sep)))

        if not cited_numbers:
            self._set_citations_text("")
            return

        # Build a lookup from source number → citation string
        source_citations = {}
        for s in self.session.sources:
            src = s if isinstance(s, dict) else s
            num = src.get('number')
            if num is not None:
                source_citations[num] = src.get(
                    'citation', f'Source {num} — citation unavailable')

        # Assemble references list
        ref_lines = []
        for num in cited_numbers:
            cit = source_citations.get(
                num, f"Source {num} — citation not found")
            ref_lines.append(f"{num}. {cit}")

        citations_text = "\n".join(ref_lines)
        self._set_citations_text(citations_text)

    def _set_citations_text(self, text: str):
        """Set the citations editor text without triggering a change loop."""
        if 'citations' not in self.section_editors:
            return
        ed = self.section_editors['citations']
        ed.editor.blockSignals(True)
        ed.editor.setPlainText(text)
        ed.editor.blockSignals(False)
        self.session.citations = text if text else None

    # ── View toggling ────────────────────────────────────────────────

    def _show_all_sections(self):
        for ed in self.section_editors.values():
            ed.setVisible(True)
            ed.set_collapsed(False)
            ed.editor.setMinimumHeight(150)
            ed.setSizePolicy(
                QSizePolicy.Policy.Preferred, QSizePolicy.Policy.Preferred)
        self.results_header_widget.setVisible(True)
        self.results_editors_container.setVisible(True)
        self._bottom_spacer.setVisible(True)

    def _show_single_section(self, section_key: str):
        is_results = section_key == "results"
        for key, ed in self.section_editors.items():
            if is_results:
                show = key.startswith("results_")
            else:
                show = (key == section_key)
            ed.setVisible(show)
            if show:
                ed.set_collapsed(False)
                ed.setSizePolicy(
                    QSizePolicy.Policy.Preferred,
                    QSizePolicy.Policy.Expanding)
            else:
                ed.setSizePolicy(
                    QSizePolicy.Policy.Preferred,
                    QSizePolicy.Policy.Preferred)
        self.results_header_widget.setVisible(is_results)
        self.results_editors_container.setVisible(is_results)
        # Hide the bottom spacer so the visible editor(s) can expand
        self._bottom_spacer.setVisible(False)

    def _show_selected_results_subsection(self):
        """Isolate the results subsection currently selected in the combo."""
        idx = self.results_sub_combo.currentIndex()
        if idx < 0 or idx >= len(self.session.results):
            return
        target_key = f"results_{idx}"
        for key, ed in self.section_editors.items():
            show = (key == target_key)
            ed.setVisible(show)
            if show:
                ed.set_collapsed(False)
                ed.setSizePolicy(
                    QSizePolicy.Policy.Preferred,
                    QSizePolicy.Policy.Expanding)
            else:
                ed.setSizePolicy(
                    QSizePolicy.Policy.Preferred,
                    QSizePolicy.Policy.Preferred)
        self.results_header_widget.setVisible(False)
        self.results_editors_container.setVisible(True)
        self._bottom_spacer.setVisible(False)

    def _update_results_sub_combo(self):
        """Refresh the results-subsection combo box from session data."""
        self.results_sub_combo.blockSignals(True)
        self.results_sub_combo.clear()
        for i, rs in enumerate(self.session.results):
            title = rs.get('section', f'Subsection {i + 1}')
            self.results_sub_combo.addItem(title)
        self.results_sub_combo.blockSignals(False)

    # ── Thesis ───────────────────────────────────────────────────────

    def _manage_thesis(self):
        current = self.session.thesis or ""
        dlg = QInputDialog(self)
        dlg.setWindowTitle("Thesis Statement")
        dlg.setLabelText(
            "Thesis (optional — guides results subsection generation):")
        dlg.setTextValue(current)
        dlg.setOption(
            QInputDialog.InputDialogOption.UsePlainTextEditForTextInput)
        dlg.resize(600, 300)
        if dlg.exec():
            self.session.thesis = dlg.textValue()
            self.section_changed.emit()

    # ── Context building ─────────────────────────────────────────────

    def _build_context_string(self, section_key: str) -> str:
        config = self.session.section_contexts.get(section_key, {})
        parts = []

        # Sections that should NOT include source summaries by default
        # (they already receive review_string in their prompts)
        no_default_summaries = {'discussion', 'conclusion', 'abstract'}
        has_saved_config = bool(config)

        # Source summaries
        summary_nums = config.get('summaries', [])
        limit = config.get('summary_limit', 0)
        source_dict = {}

        # Only include summaries if there's a saved config with explicit
        # selections, OR if this is a section that defaults to including them
        if has_saved_config and summary_nums:
            for s in self.session.sources:
                src = s if isinstance(s, dict) else s
                num = src.get('number')
                if num in summary_nums:
                    summary = src.get('summary')
                    if summary:
                        source_dict[num] = summary
        elif not has_saved_config and section_key not in no_default_summaries:
            # Default: include non-rejected summaries for intro/results sections
            for s in self.session.sources:
                src = s if isinstance(s, dict) else s
                # Never include rejected sources by default
                if src.get('rating') is False:
                    continue
                num = src.get('number')
                summary = src.get('summary')
                if summary:
                    source_dict[num] = summary
        # else: no saved config + discussion/conclusion/abstract = empty

        if limit > 0:
            source_dict = dict(list(source_dict.items())[:limit])
        if source_dict:
            parts.append(f"Source Summaries: {source_dict}")

        # Full texts
        for s in self.session.sources:
            src = s if isinstance(s, dict) else s
            if src.get('number') in config.get('full_texts', []) \
               and src.get('full_text'):
                parts.append(
                    f"Full Text [{src['number']}]: "
                    f"{src['full_text'][:3000]}")

        # Topics
        for t in self.session.topics:
            top = t if isinstance(t, dict) else t
            if top.get('topic_id') in config.get('topics', []) \
               and top.get('text'):
                parts.append(f"Topic — {top['title']}: {top['text']}")

        # Statistics
        for st in self.session.statistics:
            stat = st if isinstance(st, dict) else st
            if stat.get('stat_id') in config.get('statistics', []):
                parts.append(
                    f"Statistic — {stat['question']}: "
                    f"{stat.get('text_response', '')}")

        # Review sections as context
        for ref in config.get('review_sections', []):
            if ref.startswith('results_'):
                idx = int(ref.split('_')[1])
                if idx < len(self.session.results):
                    rs = self.session.results[idx]
                    parts.append(
                        f"Results - {rs.get('section', '')}: "
                        f"{rs.get('text', '')}")
            else:
                val = getattr(self.session, ref, None)
                if val:
                    parts.append(f"{ref.capitalize()}: {val}")

        return "\n\n".join(parts) if parts else ""

    # ── AI write ─────────────────────────────────────────────────────

    def ai_write_section(self, section_key: str):
        if not self.session.paper_topic:
            QMessageBox.warning(self, "No Topic",
                                "Please set a paper topic first.")
            return
        if not self.config.claude_api_key \
           and self.config.active_api == "claude":
            from dialogs import PromptSettingsDialog
            PromptSettingsDialog(self.config, self.api, self).exec()
            return

        context = self._build_context_string(section_key)

        prompt_key_map = {
            'abstract': 'write_abstract',
            'intro': 'write_intro',
            'methods': 'write_methods',
            'discussion': 'write_discussion',
            'conclusion': 'write_conclusion',
        }

        if section_key.startswith('results_'):
            prompt_key = 'write_results_section'
            idx = int(section_key.split('_')[1])
            section_title = self.session.results[idx].get('section', 'Untitled')
            thesis_clause = ""
            if self.session.thesis:
                thesis_clause = (
                    f"The thesis of this review is: "
                    f"{self.session.thesis}. ")
            variables = {
                'topic': self.session.paper_topic,
                'section_title': section_title,
                'thesis_clause': thesis_clause,
                'review_string': self.session.build_review_string(),
                'context': context,
            }
        elif section_key == 'methods':
            prompt_key = 'write_methods'
            sections_list = ', '.join(
                rs.get('section', 'Untitled')
                for rs in self.session.results)
            variables = {
                'topic': self.session.paper_topic,
                'methods_notes': '',
                'screening_approach':
                    'relevance assessment and inclusion criteria',
                'section_list': sections_list or 'TBD',
            }
        else:
            prompt_key = prompt_key_map.get(section_key, 'write_intro')
            variables = {
                'topic': self.session.paper_topic,
                'review_string': self.session.build_review_string(),
                'context': context,
            }

        prompt_template = self.config.get_prompt(prompt_key)
        dlg = PromptApprovalDialog(
            prompt_key, prompt_template, variables, self.config, self)
        if dlg.exec():
            formatted_prompt = dlg.get_formatted_prompt()
            params = self.config.get_params_for_prompt(prompt_key)
            try:
                from api_manager import LLMWorker
                self._active_worker = LLMWorker(
                    self.api, formatted_prompt, params)
                self._active_section_key = section_key
                self._active_worker.finished.connect(
                    self._on_ai_write_complete)
                self._active_worker.error.connect(self._on_ai_error)
                self.section_editors[section_key].write_btn.setEnabled(False)
                self.section_editors[section_key].write_btn.setText(
                    "⏳ Writing...")
                self._active_worker.start()
            except Exception as e:
                QMessageBox.critical(self, "Error", str(e))

    def _on_ai_write_complete(self, result: str):
        key = self._active_section_key
        if key in self.section_editors:
            self.section_editors[key].write_btn.setEnabled(True)
            self.section_editors[key].write_btn.setText("✦ AI Write")

        dlg = OutputApprovalDialog(key, result, self)
        if dlg.exec() and dlg.approved:
            if key in self.section_editors:
                self.section_editors[key].set_text(result)
                self._on_section_changed(key, result)

    def _on_ai_error(self, error_msg: str):
        key = self._active_section_key
        if key in self.section_editors:
            self.section_editors[key].write_btn.setEnabled(True)
            self.section_editors[key].write_btn.setText("✦ AI Write")
        QMessageBox.critical(self, "AI Error", error_msg)

    # ── Context config dialog ────────────────────────────────────────

    def configure_context(self, section_key: str):
        current = self.session.section_contexts.get(section_key, {})
        dlg = ContextConfigDialog(section_key, self.session, current, self)
        if dlg.exec():
            self.session.section_contexts[section_key] = dlg.get_config()
            self.section_changed.emit()

    # ── Text improvement ─────────────────────────────────────────────

    def improve_text(self, section_key: str, selected_text: str):
        prompt_template = self.config.get_prompt('improve_text')
        variables = {'selected_text': selected_text}
        dlg = PromptApprovalDialog(
            'improve_text', prompt_template, variables, self.config, self)
        if dlg.exec():
            formatted = dlg.get_formatted_prompt()
            params = self.config.get_params_for_prompt('improve_text')
            try:
                result = self.api.query(formatted, **params)
                out_dlg = OutputApprovalDialog(
                    "Text Improvement", result, self)
                if out_dlg.exec() and out_dlg.approved:
                    editor = self.section_editors[section_key].editor
                    cursor = editor.textCursor()
                    cursor.insertText(result)
            except Exception as e:
                QMessageBox.critical(self, "Error", str(e))

    # ── Citation actions ─────────────────────────────────────────────

    def view_citation_sources(self, numbers: list):
        self.request_sources_pop.emit(numbers)

    def validate_citation(self, numbers: list, status: str):
        for n in numbers:
            self.session.citation_validations[str(n)] = {
                'status': status, 'validation_method': 'manual'}
        for ed in self.section_editors.values():
            ed.update_validation_data(self.session.citation_validations)
        self.section_changed.emit()

    def _get_citation_preceding_text(self, section_key, citation_match):
        """
        Extract the preceding sentence for a citation and let the user
        adjust the amount of text to match against.
        Returns the user-approved text, or None if cancelled.
        """
        from citation_manager import get_preceding_sentence
        full_text = self.section_editors[section_key].get_text()
        # Block text offset: citation_match is within the block, but
        # we need it relative to the full text
        block = self.section_editors[section_key].editor.document().findBlockByNumber(0)
        # get_preceding_sentence works on full_text with absolute position
        # citation_match.start() is relative to the block_text.
        # We need to find the absolute position in full_text.
        block_text = None
        abs_start = 0
        for blk_idx in range(self.section_editors[section_key].editor.document().blockCount()):
            blk = self.section_editors[section_key].editor.document().findBlockByNumber(blk_idx)
            if citation_match.group(0) in blk.text():
                # Check if the match position lines up
                local_match = re.search(
                    re.escape(citation_match.group(0)), blk.text())
                if local_match and local_match.start() == citation_match.start():
                    abs_start = blk.position() + citation_match.start()
                    break
            abs_start = blk.position()

        sentence = get_preceding_sentence(full_text, abs_start)
        if not sentence:
            sentence = full_text[max(0, abs_start - 200):abs_start].strip()

        dlg = QInputDialog(self)
        dlg.setWindowTitle("Citation Text to Match")
        dlg.setLabelText(
            "The text below will be matched against sources.\n"
            "You can edit it to include more or less context:")
        dlg.setTextValue(sentence)
        dlg.setOption(
            QInputDialog.InputDialogOption.UsePlainTextEditForTextInput)
        dlg.resize(600, 250)
        if dlg.exec():
            return dlg.textValue().strip()
        return None

    def auto_detect_citation(self, section_key, citation_match, numbers):
        from citation_manager import auto_detect_match
        preceding = self._get_citation_preceding_text(
            section_key, citation_match)
        if preceding is None:
            return

        # Pre-compute results for all citation numbers
        pending = []
        for num in numbers:
            source_text = ""
            for s in self.session.sources:
                src = s if isinstance(s, dict) else s
                if src.get('number') == num:
                    if self.citation_val_box.currentIndex() == 0:
                        source_text = (src.get('summary')
                                       or src.get('full_text')
                                       or src.get('abstract') or "")
                    if self.citation_val_box.currentIndex() == 1:
                        source_text = (src.get('full_text') or "")
                    break
            is_match, excerpt, score = auto_detect_match(
                preceding, source_text)
            pending.append((num, is_match, excerpt, score))

        self._auto_detect_pending = pending
        self._auto_detect_preceding = preceding
        self._show_next_auto_detect_dialog()

    def _show_next_auto_detect_dialog(self):
        """Show a non-modal dialog for the next queued auto-detect result."""
        if not self._auto_detect_pending:
            return

        num, is_match, excerpt, score = self._auto_detect_pending.pop(0)
        preceding = self._auto_detect_preceding

        dlg = QDialog(self)
        dlg.setWindowTitle(f"Auto-Detect: Citation [{num}]")
        dlg.setModal(False)
        layout = QVBoxLayout(dlg)

        msg_label = QLabel(
            f"Text: \"{preceding[:200]}...\"\n\n"
            f"Best match (score: {score:.2f}):\n"
            f"\"{excerpt[:300]}...\"\n\n"
            f"Match found: {'Yes' if is_match else 'No'}\n\n"
            f"Approve this citation?")
        msg_label.setWordWrap(True)
        layout.addWidget(msg_label)

        btn_layout = QHBoxLayout()
        yes_btn = QPushButton("✓ Approve")
        no_btn = QPushButton("✗ Disapprove")
        take_btn = QPushButton("🔍 Take Me There")
        cancel_btn = QPushButton("Cancel")
        btn_layout.addWidget(yes_btn)
        btn_layout.addWidget(no_btn)
        btn_layout.addWidget(take_btn)
        btn_layout.addWidget(cancel_btn)
        layout.addLayout(btn_layout)

        def on_approve():
            self.validate_citation([num], "approved")
            dlg.close()
            self._show_next_auto_detect_dialog()

        def on_disapprove():
            self.validate_citation([num], "disapproved")
            dlg.close()
            self._show_next_auto_detect_dialog()

        def on_cancel():
            self._auto_detect_pending.clear()
            dlg.close()

        def on_take_me_there():
            self._navigate_to_citation(num, excerpt)

        yes_btn.clicked.connect(on_approve)
        no_btn.clicked.connect(on_disapprove)
        cancel_btn.clicked.connect(on_cancel)
        take_btn.clicked.connect(on_take_me_there)

        # Prevent garbage collection while dialog is open
        self._active_citation_dialog = dlg
        dlg.resize(500, 300)
        dlg.show()
        dlg.raise_()

    def _navigate_to_citation(self, citation_number, excerpt):
        """Navigate to the matched excerpt in the Sources tab."""
        # Map citation_val_box index to the sources detail_tabs index
        # 0 = summaries → detail_tabs index 2 (Summary tab)
        # 1 = full texts → detail_tabs index 1 (Full Text tab)
        val_mode = self.citation_val_box.currentIndex()
        detail_tab_index = 2 if val_mode == 0 else 1
        self.request_navigate_to_source.emit(
            citation_number, excerpt, detail_tab_index)

    def ai_detect_citation(self, section_key, citation_match, numbers):
        preceding = self._get_citation_preceding_text(
            section_key, citation_match)
        if preceding is None:
            return

        for num in numbers:
            source_text = ""
            for s in self.session.sources:
                src = s if isinstance(s, dict) else s
                if src.get('number') == num:
                    if self.citation_val_box.currentIndex() == 0:
                        source_text = (src.get('summary')
                                       or src.get('full_text')
                                       or src.get('abstract') or "")
                    if self.citation_val_box.currentIndex() == 1:
                        source_text = (src.get('full_text') or "")
                    break
            prompt_template = self.config.get_prompt('ai_validate_citation')
            variables = {
                'citation_number': num,
                'preceding_text': preceding[:500],
                'source_context': source_text[:2000],
            }
            formatted = prompt_template.format(**variables)
            try:
                result = self.api.query(formatted, max_tokens=500)
                reply = QMessageBox.question(
                    self, f"AI-Detect: Citation [{num}]",
                    f"AI Analysis:\n{result}\n\nApprove this citation?",
                    QMessageBox.StandardButton.Yes |
                    QMessageBox.StandardButton.No |
                    QMessageBox.StandardButton.Cancel)
                if reply == QMessageBox.StandardButton.Yes:
                    self.validate_citation([num], "approved")
                elif reply == QMessageBox.StandardButton.No:
                    self.validate_citation([num], "disapproved")
            except Exception as e:
                QMessageBox.critical(self, "Error", str(e))

    def swap_citation(self, section_key, citation_match, numbers):
        """
        For each citation number, search ALL source summaries to find
        the best-matching source. Show the user a comparison between
        the current source's match and the best alternative, and offer
        to swap the citation number in the text.
        Uses a non-modal dialog so the user can inspect sources while
        the dialog remains open.
        """
        from citation_manager import (
            auto_detect_match, find_best_source_match)

        preceding = self._get_citation_preceding_text(
            section_key, citation_match)
        if preceding is None:
            return

        # Pre-compute swap data for all citation numbers
        pending = []
        for num in numbers:
            # Score the CURRENT citation
            current_text = ""
            for s in self.session.sources:
                if s.get('rating') is False:
                    continue
                src = s if isinstance(s, dict) else s
                if src.get('number') == num:
                    if self.citation_val_box.currentIndex() == 0:
                        current_text = (src.get('summary')
                                       or src.get('full_text')
                                       or src.get('abstract') or "")
                    if self.citation_val_box.currentIndex() == 1:
                        current_text = (src.get('full_text') or "")
                    break

            _, current_excerpt, current_score = auto_detect_match(
                preceding, current_text)

            # Find best across ALL sources (excluding current)
            best = find_best_source_match(
                preceding, self.session.sources,
                self.citation_val_box.currentIndex(),
                exclude_numbers=[num])

            if best is None:
                QMessageBox.information(
                    self, "No Alternative",
                    f"No alternative sources found for citation [{num}].")
                continue

            best_num, best_excerpt, best_score = best
            pending.append({
                'num': num,
                'current_excerpt': current_excerpt,
                'current_score': current_score,
                'best_num': best_num,
                'best_excerpt': best_excerpt,
                'best_score': best_score,
            })

        self._swap_pending = pending
        self._swap_state = {
            'section_key': section_key,
            'citation_match': citation_match,
            'numbers': numbers,
            'preceding': preceding,
        }
        self._show_next_swap_dialog()

    def _show_next_swap_dialog(self):
        """Show a non-modal dialog for the next queued swap comparison."""
        if not self._swap_pending:
            return

        item = self._swap_pending.pop(0)
        state = self._swap_state
        preceding = state['preceding']
        section_key = state['section_key']
        citation_match = state['citation_match']
        numbers = state['numbers']
        num = item['num']
        current_excerpt = item['current_excerpt']
        current_score = item['current_score']
        best_num = item['best_num']
        best_excerpt = item['best_excerpt']
        best_score = item['best_score']

        # Build comparison message
        msg = (
            f"Matching text:\n\"{preceding[:200]}\"\n\n"
            f"── Current: [{num}] (score: {current_score:.3f}) ──\n"
            f"\"{current_excerpt[:250]}\"\n\n"
            f"── Best alternative: [{best_num}] "
            f"(score: {best_score:.3f}) ──\n"
            f"\"{best_excerpt[:250]}\"\n\n")

        if best_score > current_score:
            msg += (
                f"⬆ Source [{best_num}] is a better match "
                f"(+{best_score - current_score:.3f}).\n"
                f"Swap [{num}] → [{best_num}]?")
        else:
            msg += (
                f"Current source [{num}] is already the best or "
                f"equal match. Swap anyway to [{best_num}]?")

        dlg = QDialog(self)
        dlg.setWindowTitle(f"Swap Citation [{num}]?")
        dlg.setModal(False)
        layout = QVBoxLayout(dlg)

        msg_label = QLabel(msg)
        msg_label.setWordWrap(True)
        layout.addWidget(msg_label)

        btn_layout = QHBoxLayout()
        yes_btn = QPushButton("✓ Swap")
        no_btn = QPushButton("✗ Keep")
        view_current_btn = QPushButton(f"View Current [{num}]")
        view_best_btn = QPushButton(f"View Alternative [{best_num}]")
        cancel_btn = QPushButton("Cancel")
        btn_layout.addWidget(yes_btn)
        btn_layout.addWidget(no_btn)
        btn_layout.addWidget(view_current_btn)
        btn_layout.addWidget(view_best_btn)
        btn_layout.addWidget(cancel_btn)
        layout.addLayout(btn_layout)

        def on_swap():
            editor = self.section_editors[section_key]
            text = editor.get_text()
            old_citation = citation_match.group(0)
            if len(numbers) == 1:
                new_citation = f'[{best_num}]'
            else:
                new_nums = [best_num if n == num else n
                            for n in numbers]
                new_citation = '[' + ','.join(
                    str(n) for n in new_nums) + ']'
            new_text = (text[:text.find(old_citation)] +
                        new_citation +
                        text[text.find(old_citation) +
                             len(old_citation):])
            editor.set_text(new_text)
            self._on_section_changed(section_key, new_text)
            dlg.close()
            QMessageBox.information(
                self, "Swapped",
                f"Citation [{num}] swapped to [{best_num}].")
            self._show_next_swap_dialog()

        def on_keep():
            dlg.close()
            self._show_next_swap_dialog()

        def on_cancel():
            self._swap_pending.clear()
            dlg.close()

        def on_view_current():
            self._navigate_to_citation(num, current_excerpt)

        def on_view_best():
            self._navigate_to_citation(best_num, best_excerpt)

        yes_btn.clicked.connect(on_swap)
        no_btn.clicked.connect(on_keep)
        cancel_btn.clicked.connect(on_cancel)
        view_current_btn.clicked.connect(on_view_current)
        view_best_btn.clicked.connect(on_view_best)

        # Prevent garbage collection while dialog is open
        self._active_swap_dialog = dlg
        dlg.resize(550, 350)
        dlg.show()
        dlg.raise_()

    # ── Results topic generation ─────────────────────────────────────

    def _generate_results_topics(self):
        if not self.session.paper_topic:
            QMessageBox.warning(self, "No Topic",
                                "Please set a paper topic first.")
            return

        source_dict = {}
        for i, s in enumerate(self.session.sources):
            src = s if isinstance(s, dict) else s
            if src.get('rating') is False:
                continue
            summary = src.get('summary')
            if summary:
                source_dict[src.get('number', i)] = summary
            if len(source_dict) > 100: # Limit to stop AI from losing context
                break

        thesis_clause = ""
        if self.session.thesis:
            thesis_clause = (
                f"The thesis of this review is: {self.session.thesis}. "
                "Generate subsection headers that explicitly address "
                "this thesis. ")

        variables = {
            'topic': self.session.paper_topic,
            'thesis_clause': thesis_clause,
            'source_string': str(source_dict),
        }
        prompt_template = self.config.get_prompt('generate_results_topics')
        dlg = PromptApprovalDialog(
            'generate_results_topics', prompt_template,
            variables, self.config, self)
        if dlg.exec():
            formatted = dlg.get_formatted_prompt()
            try:
                result = self.api.query(formatted, max_tokens=1000)
                topics = [t.strip() for t in result.split(';') if t.strip()]
                reply = QMessageBox.question(
                    self, "Results Topics",
                    f"Generated {len(topics)} topics:\n" +
                    "\n".join(f"  {i+1}. {t}"
                              for i, t in enumerate(topics)) +
                    "\n\nApply these as results subsections?",
                    QMessageBox.StandardButton.Yes |
                    QMessageBox.StandardButton.No)
                if reply == QMessageBox.StandardButton.Yes:
                    self.session.results = []
                    for i, topic in enumerate(topics):
                        self.session.results.append({
                            'section_number': i,
                            'section': topic, 'text': None})
                    self._rebuild_results_editors()
                    self.section_changed.emit()
            except Exception as e:
                QMessageBox.critical(self, "Error", str(e))

    def _add_results_subsection(self):
        title, ok = QInputDialog.getText(
            self, "New Results Subsection", "Subsection title:")
        if ok and title:
            idx = len(self.session.results)
            self.session.results.append({
                'section_number': idx, 'section': title, 'text': None})
            self._rebuild_results_editors()
            self.section_changed.emit()

    def _remove_results_subsection(self):
        if not self.session.results:
            QMessageBox.information(
                self, "No Subsections",
                "There are no results subsections to remove.")
            return

        # Build a list of subsection titles for the user to pick from
        items = [f"{i+1}. {rs.get('section', 'Untitled')}"
                 for i, rs in enumerate(self.session.results)]
        choice, ok = QInputDialog.getItem(
            self, "Remove Results Subsection",
            "Select a subsection to remove:", items, 0, False)
        if ok and choice:
            idx = items.index(choice)
            title = self.session.results[idx].get('section', 'Untitled')
            reply = QMessageBox.question(
                self, "Confirm Removal",
                f"Remove results subsection \"{title}\"?\n\n"
                f"Any text in this subsection will be lost.",
                QMessageBox.StandardButton.Yes |
                QMessageBox.StandardButton.No)
            if reply == QMessageBox.StandardButton.Yes:
                old_key = f"results_{idx}"
                del self.session.results[idx]
                # Clean up section_rate on every source:
                # section_rate index 0 = intro, index idx+1 = this subsection
                rate_idx = idx + 1
                for s in self.session.sources:
                    src = s if isinstance(s, dict) else s
                    sr = src.get('section_rate')
                    if sr and rate_idx < len(sr):
                        del sr[rate_idx]
                # Clean up section_contexts:
                # 1) Remove the config entry for this subsection
                self.session.section_contexts.pop(old_key, None)
                # 2) Remove from 'review_sections' lists in other configs
                for cfg in self.session.section_contexts.values():
                    ref_list = cfg.get('review_sections', [])
                    if old_key in ref_list:
                        ref_list.remove(old_key)
                # 3) Renumber results_N keys above the deleted index
                n_remaining = len(self.session.results)
                new_contexts = {}
                for key, val in self.session.section_contexts.items():
                    if key.startswith('results_'):
                        old_idx = int(key.split('_')[1])
                        if old_idx > idx:
                            new_contexts[f'results_{old_idx - 1}'] = val
                        else:
                            new_contexts[key] = val
                    else:
                        new_contexts[key] = val
                self.session.section_contexts = new_contexts
                # Renumber remaining subsections
                for i, rs in enumerate(self.session.results):
                    rs['section_number'] = i
                self._rebuild_results_editors()
                self.section_changed.emit()

    def _write_all(self):
        reply = QMessageBox.question(
            self, "Write All Sections",
            "This will auto-write all sections using AI with current "
            "context. Results are auto-applied without individual "
            "approval dialogs. Continue?",
            QMessageBox.StandardButton.Yes | QMessageBox.StandardButton.No)
        if reply == QMessageBox.StandardButton.Yes:
            self._write_all_queue = [
                k for k in
                (['intro'] +
                 [f'results_{i}'
                  for i in range(len(self.session.results))] +
                 ['discussion', 'conclusion', 'abstract'])
                if k in self.section_editors
            ]
            self._write_all_active = True
            self._write_next_queued_section()

    def _write_next_queued_section(self):
        """Pop the next section from the write-all queue and write it."""
        if not self._write_all_queue:
            self._write_all_active = False
            QMessageBox.information(
                self, "Done", "All sections have been written.")
            return

        section_key = self._write_all_queue.pop(0)
        if section_key not in self.section_editors:
            self._write_next_queued_section()
            return

        # Build prompt (same logic as ai_write_section but no dialog)
        context = self._build_context_string(section_key)

        prompt_key_map = {
            'abstract': 'write_abstract',
            'intro': 'write_intro',
            'methods': 'write_methods',
            'discussion': 'write_discussion',
            'conclusion': 'write_conclusion',
        }

        if section_key.startswith('results_'):
            prompt_key = 'write_results_section'
            idx = int(section_key.split('_')[1])
            section_title = self.session.results[idx].get(
                'section', 'Untitled')
            thesis_clause = ""
            if self.session.thesis:
                thesis_clause = (
                    f"The thesis of this review is: "
                    f"{self.session.thesis}. ")
            variables = {
                'topic': self.session.paper_topic,
                'section_title': section_title,
                'thesis_clause': thesis_clause,
                'review_string': self.session.build_review_string(),
                'context': context,
            }
        elif section_key == 'methods':
            prompt_key = 'write_methods'
            sections_list = ', '.join(
                rs.get('section', 'Untitled')
                for rs in self.session.results)
            variables = {
                'topic': self.session.paper_topic,
                'methods_notes': '',
                'screening_approach':
                    'relevance assessment and inclusion criteria',
                'section_list': sections_list or 'TBD',
            }
        else:
            prompt_key = prompt_key_map.get(section_key, 'write_intro')
            variables = {
                'topic': self.session.paper_topic,
                'review_string': self.session.build_review_string(),
                'context': context,
            }

        prompt_template = self.config.get_prompt(prompt_key)
        try:
            formatted_prompt = prompt_template.format(**variables)
        except (KeyError, ValueError) as e:
            QMessageBox.warning(
                self, "Prompt Error",
                f"Error formatting prompt for {section_key}: {e}")
            self._write_next_queued_section()
            return
        params = self.config.get_params_for_prompt(prompt_key)

        try:
            from api_manager import LLMWorker
            worker = LLMWorker(self.api, formatted_prompt, params)
            # Bind the section_key into the callbacks via default arg
            worker.finished.connect(
                lambda result, k=section_key:
                    self._on_write_all_section_done(k, result))
            worker.error.connect(
                lambda err, k=section_key:
                    self._on_write_all_section_error(k, err))
            self.section_editors[section_key].write_btn.setEnabled(False)
            self.section_editors[section_key].write_btn.setText(
                "⏳ Writing...")
            worker.start()
            # Keep a reference so the worker isn't garbage-collected
            self._active_worker = worker
        except Exception as e:
            QMessageBox.critical(self, "Error", str(e))
            self._write_next_queued_section()

    def _on_write_all_section_done(self, section_key: str, result: str):
        """Auto-apply result for a write-all section, then continue."""
        if section_key in self.section_editors:
            self.section_editors[section_key].write_btn.setEnabled(True)
            self.section_editors[section_key].write_btn.setText(
                "✦ AI Write")
            self.section_editors[section_key].set_text(result)
            self._on_section_changed(section_key, result)
        # Continue to next section in queue
        self._write_next_queued_section()

    def _on_write_all_section_error(self, section_key: str, error: str):
        """Handle error during write-all, continue to next section."""
        if section_key in self.section_editors:
            self.section_editors[section_key].write_btn.setEnabled(True)
            self.section_editors[section_key].write_btn.setText(
                "✦ AI Write")
        QMessageBox.warning(
            self, f"Error writing {section_key}",
            f"Skipping {section_key}: {error}")
        self._write_next_queued_section()

    def _validate_all_citations(self):
        from PyQt6.QtWidgets import QDialog, QDialogButtonBox

        # Ask user which validation method to use
        msg = QMessageBox(self)
        msg.setWindowTitle("Validate All Citations")
        msg.setText(
            "Choose a validation method for all citations:")
        msg.setInformativeText(
            "<b>AI Validation</b> — uses the LLM to check each "
            "citation against its source (slower, higher accuracy).\n\n"
            "<b>Automated (regex)</b> — uses text similarity matching "
            "to score each citation (faster, requires a threshold).")
        ai_btn = msg.addButton(
            "AI Validation", QMessageBox.ButtonRole.AcceptRole)
        regex_btn = msg.addButton(
            "Automated (regex)", QMessageBox.ButtonRole.ActionRole)
        cancel_btn = msg.addButton(QMessageBox.StandardButton.Cancel)
        msg.exec()

        clicked = msg.clickedButton()
        if clicked == cancel_btn:
            return
        elif clicked == ai_btn:
            self._validate_all_citations_ai()
        elif clicked == regex_btn:
            self._validate_all_citations_regex()

    def _validate_all_citations_ai(self):
        """Validate all citations using AI (original behavior)."""
        reply = QMessageBox.question(
            self, "AI Validation",
            "This will use AI to validate all citations. "
            "This may take a while. Continue?",
            QMessageBox.StandardButton.Yes | QMessageBox.StandardButton.No)
        if reply != QMessageBox.StandardButton.Yes:
            return

        from citation_manager import find_citation_spans, get_preceding_text
        for key, editor in self.section_editors.items():
            text = editor.get_text()
            if not text:
                continue
            spans = find_citation_spans(text)
            for span in spans:
                preceding = get_preceding_text(text, span['start'])
                for num in span['numbers']:
                    source_text = ""
                    for s in self.session.sources:
                        src = s if isinstance(s, dict) else s
                        if src.get('number') == num:
                            if self.citation_val_box.currentIndex() == 0:
                                source_text = (src.get('summary')
                                               or src.get('full_text')
                                               or src.get('abstract') or "")
                            elif self.citation_val_box.currentIndex() == 1:
                                source_text = (src.get('full_text') or "")
                            break
                    prompt = self.config.get_prompt(
                        'ai_validate_citation_batch')
                    formatted = prompt.format(
                        citation_number=num,
                        preceding_text=preceding[:500],
                        source_context=source_text[:2000])
                    try:
                        result = self.api.query(
                            formatted, max_tokens=10).strip()
                        status = ('approved'
                                  if result.lower().startswith('true')
                                  else 'disapproved')
                        self.session.citation_validations[str(num)] = {
                            'status': status,
                            'validation_method': 'batch_ai'}
                    except Exception:
                        pass

        for ed in self.section_editors.values():
            ed.update_validation_data(self.session.citation_validations)
        self.section_changed.emit()
        QMessageBox.information(
            self, "Done", "AI citation validation complete.")

    def _validate_all_citations_regex(self):
        """Validate all citations using automated text similarity matching."""
        from PyQt6.QtWidgets import QDoubleSpinBox, QDialog, QFormLayout

        # Ask for threshold
        dlg = QDialog(self)
        dlg.setWindowTitle("Set Similarity Threshold")
        layout = QVBoxLayout(dlg)
        layout.addWidget(QLabel(
            "Set the minimum similarity score (0.0–1.0) for a citation "
            "to be approved.\n\n"
            "Lower values are more lenient, higher values are stricter.\n"
            "Recommended: 0.25–0.40"))
        form = QFormLayout()
        threshold_spin = QDoubleSpinBox()
        threshold_spin.setRange(0.0, 1.0)
        threshold_spin.setSingleStep(0.05)
        threshold_spin.setValue(0.30)
        threshold_spin.setDecimals(2)
        form.addRow("Threshold:", threshold_spin)
        layout.addLayout(form)
        btn_layout = QHBoxLayout()
        ok_btn = QPushButton("Run Validation")
        ok_btn.clicked.connect(dlg.accept)
        btn_layout.addWidget(ok_btn)
        cancel_btn = QPushButton("Cancel")
        cancel_btn.clicked.connect(dlg.reject)
        btn_layout.addWidget(cancel_btn)
        layout.addLayout(btn_layout)
        if not dlg.exec():
            return

        threshold = threshold_spin.value()

        from citation_manager import (
            find_citation_spans, get_preceding_text, auto_detect_match)

        validated_count = 0
        approved_count = 0
        disapproved_count = 0

        for key, editor in self.section_editors.items():
            text = editor.get_text()
            if not text:
                continue
            spans = find_citation_spans(text)
            for span in spans:
                preceding = get_preceding_text(text, span['start'])
                if not preceding:
                    continue
                for num in span['numbers']:
                    source_text = ""
                    for s in self.session.sources:
                        src = s if isinstance(s, dict) else s
                        if src.get('number') == num:
                            if self.citation_val_box.currentIndex() == 0:
                                source_text = (src.get('summary')
                                               or src.get('full_text')
                                               or src.get('abstract') or "")
                            elif self.citation_val_box.currentIndex() == 1:
                                source_text = (src.get('full_text') or "")
                            break
                    if not source_text:
                        continue
                    

                    is_match, excerpt, score = auto_detect_match(
                        preceding, source_text, threshold=threshold)

                    status = 'approved' if score >= threshold \
                        else 'disapproved'
                    self.session.citation_validations[str(num)] = {
                        'status': status,
                        'validation_method': 'auto_detect',
                        'match_text': f"score={score:.3f}"}
                    validated_count += 1
                    if status == 'approved':
                        approved_count += 1
                    else:
                        disapproved_count += 1


        for ed in self.section_editors.values():
            ed.update_validation_data(self.session.citation_validations)
        self.section_changed.emit()
        QMessageBox.information(
            self, "Done",
            f"Automated citation validation complete.\n\n"
            f"Validated: {validated_count}\n"
            f"Approved (≥{threshold:.2f}): {approved_count}\n"
            f"Disapproved (<{threshold:.2f}): {disapproved_count}")
```
