## Supplementary material for "In Search of Ethical Procedures for LLM-Assisted Systematic Review Production: A Proof-of-Concept Evaluation of Selected Review Components": source_data: sources_tab.py

````
"""
Sources Tab — manages source retrieval, summarization, inclusion screening,
and section relevance rating.
"""
import webbrowser
from functools import partial

from PyQt6.QtWidgets import (
    QWidget, QVBoxLayout, QHBoxLayout, QSplitter, QTableWidget, QTableWidgetItem,
    QHeaderView, QPushButton, QGroupBox, QLabel, QTextEdit, QComboBox, QSpinBox,
    QMessageBox, QInputDialog, QListWidget, QListWidgetItem, QTabWidget,
    QLineEdit, QCheckBox, QProgressBar, QMenu, QAbstractItemView, QFormLayout
)
from PyQt6.QtCore import Qt, pyqtSignal, QThread
from PyQt6.QtGui import QAction, QColor

from dialogs import PromptApprovalDialog, OutputApprovalDialog


class SourceRetrievalWorker(QThread):
    """Background worker for source retrieval."""
    progress = pyqtSignal(str)
    finished = pyqtSignal(object, list, list)
    error = pyqtSignal(str)

    def __init__(self, api_key, search_terms, retmax, per_term_limits=None):
        super().__init__()
        self.api_key = api_key
        self.search_terms = search_terms
        self.retmax = retmax
        self.per_term_limits = per_term_limits

    def run(self):
        try:
            from ncbi_retrieval import retrieve_sources
            sources, textless, abless = retrieve_sources(
                self.api_key, self.search_terms, self.retmax,
                self.per_term_limits,
                progress_callback=lambda msg: self.progress.emit(msg)
            )
            self.finished.emit(sources, textless, abless)
        except Exception as e:
            self.error.emit(str(e))


class SourceInfoFillWorker(QThread):
    """Background worker that fetches source metadata from NCBI by DOI.

    Retrieves article metadata (title, journal, year, abstract, authors,
    citation) from PubMed, and — when available — full text from PubMed
    Central (PMC).  All NCBI logic lives in ncbi_retrieval.py.
    """
    finished = pyqtSignal(int, dict)   # (row_index, field_dict)
    error = pyqtSignal(int, str)

    def __init__(self, api_key: str, row_index: int, doi: str):
        super().__init__()
        self.api_key = api_key
        self.row_index = row_index
        self.doi = doi.strip()

    def run(self):
        try:
            from ncbi_retrieval import (
                search_by_doi, fetch_article_by_pmid, get_full_text)

            pmids = search_by_doi(self.doi, self.api_key)
            if not pmids:
                self.error.emit(
                    self.row_index,
                    "No PubMed results found for this DOI.")
                return

            fields = fetch_article_by_pmid(pmids[0], self.api_key)
            if not fields:
                self.error.emit(
                    self.row_index,
                    "PubMed returned an empty record for this DOI.")
                return

            # If a PMCID is available, try to pull the full text from PMC
            pmc_id = fields.get("pmc_id", "")
            if pmc_id:
                try:
                    full_text = get_full_text(pmc_id, self.api_key)
                    if full_text:
                        fields["full_text"] = full_text
                except Exception:
                    pass  # full text is best-effort; metadata still succeeds

            self.finished.emit(self.row_index, fields)

        except Exception as e:
            self.error.emit(self.row_index, str(e))


class SourcesTab(QWidget):
    """Tab for managing sources."""

    sources_changed = pyqtSignal()

    def __init__(self, config_manager, api_manager, session, parent=None):
        super().__init__(parent)
        self.config = config_manager
        self.api = api_manager
        self.session = session
        self._per_term_limits = {}  # term -> custom limit
        self._loading = False  # suppress auto-save while populating widgets
        self._build_ui()
        self._load_from_session()

    def _build_ui(self):
        layout = QVBoxLayout(self)
        splitter = QSplitter(Qt.Orientation.Horizontal)

        # ── Left panel: Source list ──
        left = QWidget()
        left_layout = QVBoxLayout(left)

        toolbar = QHBoxLayout()
        retrieve_btn = QPushButton("🔍 Retrieve Sources")
        retrieve_btn.clicked.connect(self._retrieve_sources)
        toolbar.addWidget(retrieve_btn)

        fill_out_btn = QPushButton("Fill out All Sources")
        fill_out_btn.clicked.connect(self._fill_out_sources)
        toolbar.addWidget(fill_out_btn)

        add_btn = QPushButton("+ Add Manual Source")
        add_btn.clicked.connect(self._add_manual_source)
        toolbar.addWidget(add_btn)

        remove_src_btn = QPushButton("- Remove Source")
        remove_src_btn.clicked.connect(self._remove_source)
        toolbar.addWidget(remove_src_btn)

        summarize_btn = QPushButton("✦ Batch Summarize")
        summarize_btn.clicked.connect(self._batch_summarize)
        summarize_btn.setToolTip("Summarize All Non Excluded Abstracts")
        toolbar.addWidget(summarize_btn)

        screen_btn = QPushButton("✦ Screen for Inclusion")
        screen_btn.clicked.connect(self._screen_inclusion)
        screen_btn.setToolTip("Screens all sources for included that do not yet have an inclusion verdict")
        toolbar.addWidget(screen_btn)

        rate_btn = QPushButton("✦ Rate for Sections")
        rate_btn.clicked.connect(self._rate_for_sections)
        toolbar.addWidget(rate_btn)

        bias_btn = QPushButton("✦ Risk of Bias")
        bias_btn.clicked.connect(self._risk_of_bias)
        bias_btn.setToolTip("Assess risk of bias for all included sources using Cochrane RoB 2 tool. This tool is mainly to appraise randomized control trials. The default prompt is designed to be broad and nonspecific. For higher specificity to some source or for other study types, please edit the prompt as needed.")
        toolbar.addWidget(bias_btn)


        left_layout.addLayout(toolbar)

        self.status_label = QLabel("Ready")
        left_layout.addWidget(self.status_label)
        self.progress_bar = QProgressBar()
        self.progress_bar.setVisible(False)
        left_layout.addWidget(self.progress_bar)

        self.source_table = QTableWidget()
        self.source_table.setColumnCount(6)
        self.source_table.setHorizontalHeaderLabels(
            ["#", "Title", "Year", "Journal", "Rating", "Summary"])
        self.source_table.horizontalHeader().setSectionResizeMode(
            1, QHeaderView.ResizeMode.Stretch)
        self.source_table.setSelectionBehavior(
            QAbstractItemView.SelectionBehavior.SelectRows)
        self.source_table.itemSelectionChanged.connect(
            self._on_source_selected_from_selection)
        self.source_table.cellChanged.connect(self._on_table_cell_changed)
        self.source_table.setContextMenuPolicy(
            Qt.ContextMenuPolicy.CustomContextMenu)
        self.source_table.customContextMenuRequested.connect(
            self._show_source_context_menu)
        left_layout.addWidget(self.source_table)
        splitter.addWidget(left)

        # ── Right panel: Source detail ──
        right = QWidget()
        right_layout = QVBoxLayout(right)

        self.detail_title = QLabel("<b>Select a source</b>")
        right_layout.addWidget(self.detail_title)
        self.detail_tabs = QTabWidget()

        # Abstract tab
        abstract_tab = QWidget()
        ab_layout = QVBoxLayout(abstract_tab)
        self.abstract_edit = QTextEdit()
        self.abstract_edit.textChanged.connect(lambda: self._auto_save_field('abstract'))
        ab_layout.addWidget(self.abstract_edit)
        self.detail_tabs.addTab(abstract_tab, "Abstract")

        # Full Text tab
        ft_tab = QWidget()
        ft_layout = QVBoxLayout(ft_tab)
        self.fulltext_edit = QTextEdit()
        self.fulltext_edit.textChanged.connect(lambda: self._auto_save_field('full_text'))
        ft_layout.addWidget(self.fulltext_edit)
        self.detail_tabs.addTab(ft_tab, "Full Text")

        # Summary tab
        sum_tab = QWidget()
        sum_layout = QVBoxLayout(sum_tab)
        self.summary_edit = QTextEdit()
        self.summary_edit.textChanged.connect(lambda: self._auto_save_field('summary'))
        sum_layout.addWidget(self.summary_edit)
        sum_btn_layout = QHBoxLayout()
        gen_sum_btn = QPushButton("✦ Generate Summary")
        gen_sum_btn.clicked.connect(self._generate_summary_single)
        sum_btn_layout.addWidget(gen_sum_btn)
        sum_layout.addLayout(sum_btn_layout)
        self.detail_tabs.addTab(sum_tab, "Summary")

        # Info tab
        info_tab = QWidget()
        info_layout = QFormLayout(info_tab)
        self.info_title = QLineEdit()
        self.info_title.textChanged.connect(lambda text: self._auto_save_field('title'))
        info_layout.addRow("Title:", self.info_title)
        self.info_citation = QTextEdit()
        self.info_citation.setMaximumHeight(80)
        self.info_citation.textChanged.connect(lambda: self._auto_save_field('citation'))
        info_layout.addRow("Citation:", self.info_citation)
        self.info_doi = QLineEdit()
        self.info_doi.textChanged.connect(lambda text: self._auto_save_field('doi'))
        info_layout.addRow("DOI:", self.info_doi)
        doi_link_btn = QPushButton("Open DOI in Browser")
        doi_link_btn.clicked.connect(self._open_doi)
        info_layout.addRow("", doi_link_btn)
        self.info_pmid = QLineEdit()
        self.info_pmid.setReadOnly(True)
        info_layout.addRow("PMID:", self.info_pmid)
        self.info_rating = QLabel("")
        info_layout.addRow("Inclusion:", self.info_rating)
        self.info_explanation = QTextEdit()
        self.info_explanation.setMaximumHeight(80)
        self.info_explanation.setReadOnly(True)
        info_layout.addRow("Explanation:", self.info_explanation)
        include_btn = QPushButton("Include")
        include_btn.clicked.connect(lambda: self._set_inclusion(True))
        exclude_btn = QPushButton("Exclude")
        exclude_btn.clicked.connect(lambda: self._set_inclusion(False))
        ai_reassess = QPushButton("AI Assess ✦")
        ai_reassess.clicked.connect(self._ai_solo_screen)
        btn_row = QHBoxLayout()
        btn_row.addWidget(include_btn)
        btn_row.addWidget(exclude_btn)
        btn_row.addWidget(ai_reassess)
        info_layout.addRow("Manual:", btn_row)

        self.detail_tabs.addTab(info_tab, "Info")

        # ── NEW: Ratings tab ──
        ratings_tab = QWidget()
        ratings_layout = QVBoxLayout(ratings_tab)

        ratings_layout.addWidget(QLabel(
            "<b>Section Relevance Ratings</b> (1-10 per results subsection):"))
        self.section_ratings_table = QTableWidget()
        self.section_ratings_table.setColumnCount(2)
        self.section_ratings_table.setHorizontalHeaderLabels(
            ["Section", "Rating"])
        self.section_ratings_table.horizontalHeader().setSectionResizeMode(
            0, QHeaderView.ResizeMode.Stretch)
        self.section_ratings_table.setMaximumHeight(200)
        self.section_ratings_table.cellChanged.connect(
            lambda row, col: self._auto_save_rating('section_rate', self.section_ratings_table, row, col))
        ratings_layout.addWidget(self.section_ratings_table)

        ratings_layout.addWidget(QLabel(
            "<b>Topic Relevance Ratings</b> (1-10 per topic):"))
        self.topic_ratings_table = QTableWidget()
        self.topic_ratings_table.setColumnCount(2)
        self.topic_ratings_table.setHorizontalHeaderLabels(
            ["Topic", "Rating"])
        self.topic_ratings_table.horizontalHeader().setSectionResizeMode(
            0, QHeaderView.ResizeMode.Stretch)
        self.topic_ratings_table.setMaximumHeight(200)
        self.topic_ratings_table.cellChanged.connect(
            lambda row, col: self._auto_save_dict_rating('topic_ratings', self.topic_ratings_table, row, col))
        ratings_layout.addWidget(self.topic_ratings_table)

        ratings_layout.addWidget(QLabel(
            "<b>Stat Relevance Ratings</b> (1-10 per stat question):"))
        self.stat_ratings_table = QTableWidget()
        self.stat_ratings_table.setColumnCount(2)
        self.stat_ratings_table.setHorizontalHeaderLabels(
            ["Stat Question", "Rating"])
        self.stat_ratings_table.horizontalHeader().setSectionResizeMode(
            0, QHeaderView.ResizeMode.Stretch)
        self.stat_ratings_table.setMaximumHeight(200)
        self.stat_ratings_table.cellChanged.connect(
            lambda row, col: self._auto_save_dict_rating('stat_ratings', self.stat_ratings_table, row, col))
        ratings_layout.addWidget(self.stat_ratings_table)

        ratings_layout.addStretch()
        self.detail_tabs.addTab(ratings_tab, "Ratings")

        # ── Risk of Bias tab ──
        rob_tab = QWidget()
        rob_layout = QVBoxLayout(rob_tab)
        rob_layout.addWidget(QLabel(
            "<b>Cochrane Risk of Bias (RoB 2) Assessment</b>"))
        rob_form = QFormLayout()
        self.info_bias_assessment = QLabel("")
        rob_form.addRow("Overall Judgement:", self.info_bias_assessment)
        rob_layout.addLayout(rob_form)
        rob_layout.addWidget(QLabel("Explanation:"))
        self.info_bias_explanation = QTextEdit()
        self.info_bias_explanation.setReadOnly(True)
        rob_layout.addWidget(self.info_bias_explanation)
        rob_btn_row = QHBoxLayout()
        ai_rob_btn = QPushButton("AI Assess ✦")
        ai_rob_btn.setToolTip("Assess risk of bias for this source using the LLM")
        ai_rob_btn.clicked.connect(self._ai_solo_bias)
        rob_btn_row.addWidget(ai_rob_btn)
        clear_rob_btn = QPushButton("Clear")
        clear_rob_btn.setToolTip("Clear the risk of bias assessment for this source")
        clear_rob_btn.clicked.connect(self._clear_solo_bias)
        rob_btn_row.addWidget(clear_rob_btn)
        rob_layout.addLayout(rob_btn_row)
        self.detail_tabs.addTab(rob_tab, "Risk of Bias")

        right_layout.addWidget(self.detail_tabs)
        splitter.addWidget(right)
        splitter.setSizes([500, 400])

        # ── Search Terms section (with per-term limits) ──
        search_group = QGroupBox("Boolean Search Terms")
        search_layout = QVBoxLayout(search_group)

        search_toolbar = QHBoxLayout()
        gen_terms_btn = QPushButton("✦ Generate Search Terms")
        gen_terms_btn.clicked.connect(self._generate_search_terms)
        search_toolbar.addWidget(gen_terms_btn)

        add_term_btn = QPushButton("+ Add Term")
        add_term_btn.clicked.connect(self._add_search_term)
        search_toolbar.addWidget(add_term_btn)

        remove_term_btn = QPushButton("- Remove Selected")
        remove_term_btn.clicked.connect(self._remove_search_term)
        search_toolbar.addWidget(remove_term_btn)

        search_toolbar.addStretch()
        search_toolbar.addWidget(QLabel("Default per-term limit:"))
        self.default_retmax = QSpinBox()
        self.default_retmax.setRange(1, 1000)
        self.default_retmax.setValue(100)
        search_toolbar.addWidget(self.default_retmax)

        set_all_btn = QPushButton("Set All to Default")
        set_all_btn.setToolTip(
            "Reset all per-term limits to the default value")
        set_all_btn.clicked.connect(self._set_all_limits_to_default)
        search_toolbar.addWidget(set_all_btn)

        search_layout.addLayout(search_toolbar)

        # Search terms table with per-term limit column
        self.search_terms_table = QTableWidget()
        self.search_terms_table.setColumnCount(2)
        self.search_terms_table.setHorizontalHeaderLabels(
            ["Search Term", "Limit"])
        self.search_terms_table.horizontalHeader().setSectionResizeMode(
            0, QHeaderView.ResizeMode.Stretch)
        self.search_terms_table.setColumnWidth(1, 80)
        search_layout.addWidget(self.search_terms_table)

        # ── Screening Criteria section ──
        criteria_group = QGroupBox("Screening Criteria")
        criteria_outer = QVBoxLayout(criteria_group)

        # Screening mode toggle
        screen_mode_row = QHBoxLayout()
        screen_mode_row.addWidget(QLabel("Screening mode:"))
        self.screen_full_text_toggle = QCheckBox("Use full text instead of abstract")
        self.screen_full_text_toggle.setToolTip(
            "When checked, screening sends Title + Full Text to the AI.\n"
            "When unchecked (default), screening sends Title + Abstract.")
        screen_mode_row.addWidget(self.screen_full_text_toggle)
        screen_mode_row.addStretch()
        criteria_outer.addLayout(screen_mode_row)

        criteria_halves = QHBoxLayout()

        # --- Inclusion criteria (left) ---
        inc_box = QGroupBox("Inclusion Criteria (must meet ALL)")
        inc_layout = QVBoxLayout(inc_box)
        self.inclusion_list = QListWidget()
        self.inclusion_list.itemChanged.connect(self._on_criterion_edited)
        inc_layout.addWidget(self.inclusion_list)
        inc_btn_row = QHBoxLayout()
        add_inc_btn = QPushButton("+ Add")
        add_inc_btn.clicked.connect(self._add_inclusion_criterion)
        inc_btn_row.addWidget(add_inc_btn)
        rm_inc_btn = QPushButton("- Remove")
        rm_inc_btn.clicked.connect(self._remove_inclusion_criterion)
        inc_btn_row.addWidget(rm_inc_btn)
        inc_layout.addLayout(inc_btn_row)
        criteria_halves.addWidget(inc_box)

        # --- Exclusion criteria (right) ---
        exc_box = QGroupBox("Exclusion Criteria (any → exclude)")
        exc_layout = QVBoxLayout(exc_box)
        self.exclusion_list = QListWidget()
        self.exclusion_list.itemChanged.connect(self._on_criterion_edited)
        exc_layout.addWidget(self.exclusion_list)
        exc_btn_row = QHBoxLayout()
        add_exc_btn = QPushButton("+ Add")
        add_exc_btn.clicked.connect(self._add_exclusion_criterion)
        exc_btn_row.addWidget(add_exc_btn)
        rm_exc_btn = QPushButton("- Remove")
        rm_exc_btn.clicked.connect(self._remove_exclusion_criterion)
        exc_btn_row.addWidget(rm_exc_btn)
        exc_layout.addLayout(exc_btn_row)
        criteria_halves.addWidget(exc_box)

        criteria_outer.addLayout(criteria_halves)

        # ── Vertical splitter for resizable regions ──
        v_splitter = QSplitter(Qt.Orientation.Vertical)
        v_splitter.addWidget(splitter)
        v_splitter.addWidget(search_group)
        v_splitter.addWidget(criteria_group)
        v_splitter.setStretchFactor(0, 3)   # source tables get most space
        v_splitter.setStretchFactor(1, 1)   # search terms
        v_splitter.setStretchFactor(2, 1)   # criteria
        v_splitter.setChildrenCollapsible(False)
        layout.addWidget(v_splitter)

    # ── Right-click context menu ─────────────────────────────────────

    def _show_source_context_menu(self, pos):
        """Show a context menu when the user right-clicks in the source table."""
        row = self.source_table.rowAt(pos.y())
        if row < 0 or row >= len(self.session.sources):
            return
        src = self.session.sources[row]
        source_number = src.get('number', row + 1)

        menu = QMenu(self)
        fill_action = QAction(
            f"Fill Out Source {source_number} Information", self)
        fill_action.triggered.connect(lambda: self._fill_source_info(row))
        menu.addAction(fill_action)
        menu.exec(self.source_table.viewport().mapToGlobal(pos))

    def _fill_source_info(self, row: int):
        """Kick off an NCBI lookup to fill missing fields for the source at *row*."""
        if not self.config.ncbi_api_key:
            QMessageBox.warning(
                self, "No NCBI Key",
                "Please set your NCBI API key in Prompt Settings before "
                "using this feature.")
            return

        src = self.session.sources[row]
        doi = (src.get('doi') or "").strip()

        if not doi:
            QMessageBox.warning(
                self, "No DOI",
                "This source has no DOI filled in.\n\n"
                "A DOI is required to look up source information from "
                "PubMed. Please add one in the Info tab first.")
            return

        source_number = src.get('number', row + 1)
        self.status_label.setText(
            f"Looking up information for Source {source_number}...")

        worker = SourceInfoFillWorker(
            self.config.ncbi_api_key, row, doi)
        worker.finished.connect(self._on_fill_info_done)
        worker.error.connect(self._on_fill_info_error)
        # prevent garbage collection
        self._fill_info_worker = worker
        worker.start()

    def _on_fill_info_done(self, row: int, fields: dict):
        """Merge retrieved fields into the source, filling only what is missing."""
        if row < 0 or row >= len(self.session.sources):
            return
        src = self.session.sources[row]
        filled = []
        for key, value in fields.items():
            if not value:
                continue
            existing = src.get(key)
            if not existing or (isinstance(existing, str)
                                and not existing.strip()):
                src[key] = value
                filled.append(key)

        source_number = src.get('number', row + 1)
        if filled:
            self.status_label.setText(
                f"Source {source_number}: filled {', '.join(filled)}.")
        else:
            self.status_label.setText(
                f"Source {source_number}: all fields already populated "
                f"(nothing new to fill).")

        self._refresh_table()
        if self.source_table.currentRow() == row:
            self._on_source_selected(row)
        self.sources_changed.emit()

    def _on_fill_info_error(self, row: int, msg: str):
        """Handle a failed NCBI lookup gracefully."""
        source_number = "?"
        if 0 <= row < len(self.session.sources):
            source_number = self.session.sources[row].get('number', row + 1)
        self.status_label.setText(
            f"Source {source_number}: lookup failed — {msg}")
        QMessageBox.warning(
            self, "Lookup Failed",
            f"Could not retrieve information for Source {source_number}.\n\n"
            f"{msg}")

    # ── Helpers ───────────────────────────────────────────────────────

    def _on_source_selected_from_selection(self):
        self._on_source_selected(self.source_table.currentRow())

    def _on_table_cell_changed(self, row, col):
        """Handle direct edits in the source table (title, year, journal)."""
        if self._loading:
            return
        if row < 0 or row >= len(self.session.sources):
            return
        src = self.session.sources[row]
        if not isinstance(src, dict):
            return
        item = self.source_table.item(row, col)
        if not item:
            return
        text = item.text()
        field_map = {1: 'title', 2: 'year', 3: 'journal'}
        field = field_map.get(col)
        if not field:
            return
        src[field] = text if text else None
        if field == 'title' and row == self.source_table.currentRow():
            # Sync to the Info panel and header without retriggering
            self._loading = True
            self.info_title.setText(text or '')
            num = src.get('number', '')
            self.detail_title.setText(
                f"<b>[{num}] {text or 'Untitled'}</b>")
            self._loading = False
        self.sources_changed.emit()

    def _load_from_session(self):
        self._refresh_table()
        self._refresh_search_terms_table()
        self.inclusion_list.clear()
        for c in self.session.inclusion_criteria:
            item = QListWidgetItem(c)
            item.setFlags(item.flags() | Qt.ItemFlag.ItemIsEditable)
            self.inclusion_list.addItem(item)
        self.exclusion_list.clear()
        for c in self.session.exclusion_criteria:
            item = QListWidgetItem(c)
            item.setFlags(item.flags() | Qt.ItemFlag.ItemIsEditable)
            self.exclusion_list.addItem(item)

    def _refresh_table(self):
        self._loading = True
        self.source_table.setRowCount(0)
        for s in self.session.sources:
            src = s if isinstance(s, dict) else s
            row = self.source_table.rowCount()
            self.source_table.insertRow(row)

            self.source_table.setItem(
                row, 0, QTableWidgetItem(str(src.get('number', ''))))
            self.source_table.setItem(
                row, 1, QTableWidgetItem(str(src.get('title', ''))[:80]))
            self.source_table.setItem(
                row, 2, QTableWidgetItem(str(src.get('year', ''))))
            self.source_table.setItem(
                row, 3, QTableWidgetItem(str(src.get('journal', ''))[:30]))

            rating = src.get('rating')
            rating_text = ("✓" if rating is True
                           else "✗" if rating is False
                           else "?")
            rating_item = QTableWidgetItem(rating_text)
            if rating is True:
                rating_item.setForeground(QColor("#2ecc71"))
            elif rating is False:
                rating_item.setForeground(QColor("#e74c3c"))
            self.source_table.setItem(row, 4, rating_item)

            has_summary = "Yes" if src.get('summary') else "No"
            self.source_table.setItem(
                row, 5, QTableWidgetItem(has_summary))
        self._loading = False

    def _refresh_search_terms_table(self):
        """Populate the search terms table with per-term limits."""
        self.search_terms_table.setRowCount(0)
        default_limit = self.default_retmax.value()
        for term in self.session.search_terms:
            row = self.search_terms_table.rowCount()
            self.search_terms_table.insertRow(row)
            self.search_terms_table.setItem(
                row, 0, QTableWidgetItem(term))

            limit_spin = QSpinBox()
            limit_spin.setRange(1, 1000)
            limit_spin.setValue(
                self._per_term_limits.get(term, default_limit))
            limit_spin.valueChanged.connect(
                lambda val, t=term: self._on_term_limit_changed(t, val))
            self.search_terms_table.setCellWidget(row, 1, limit_spin)

    def _on_term_limit_changed(self, term: str, value: int):
        self._per_term_limits[term] = value

    def _set_all_limits_to_default(self):
        default_val = self.default_retmax.value()
        self._per_term_limits.clear()
        for row in range(self.search_terms_table.rowCount()):
            widget = self.search_terms_table.cellWidget(row, 1)
            if isinstance(widget, QSpinBox):
                widget.blockSignals(True)
                widget.setValue(default_val)
                widget.blockSignals(False)

    def _on_source_selected(self, row):
        if row < 0 or row >= len(self.session.sources):
            return
        self._loading = True
        src = self.session.sources[row]
        s = src if isinstance(src, dict) else (
            src.to_dict() if hasattr(src, 'to_dict') else src)

        self.detail_title.setText(
            f"<b>[{s.get('number')}] {s.get('title', 'Untitled')}</b>")
        self.info_title.setText(s.get('title') or "")
        self.abstract_edit.setPlainText(s.get('abstract') or "")
        self.fulltext_edit.setPlainText(s.get('full_text') or "")
        self.summary_edit.setPlainText(s.get('summary') or "")
        self.info_citation.setPlainText(s.get('citation') or "")
        self.info_doi.setText(s.get('doi') or "")
        self.info_pmid.setText(s.get('pmid') or "")

        rating = s.get('rating')
        if rating is True:
            self.info_rating.setText("✓ Included")
            self.info_rating.setStyleSheet(
                "color: #2ecc71; font-weight: bold;")
        elif rating is False:
            self.info_rating.setText("✗ Excluded")
            self.info_rating.setStyleSheet(
                "color: #e74c3c; font-weight: bold;")
        else:
            self.info_rating.setText("Not yet screened")
            self.info_rating.setStyleSheet("")

        self.info_explanation.setPlainText(s.get('rate_explain') or "")

        # Bias assessment display
        bias = s.get('bias_assessment')
        if bias:
            color_map = {
                'Low': '#2ecc71',
                'Some concerns': '#f39c12',
                'High': '#e74c3c',
            }
            color = color_map.get(bias, '')
            style = f"color: {color}; font-weight: bold;" if color else ""
            self.info_bias_assessment.setText(bias)
            self.info_bias_assessment.setStyleSheet(style)
        else:
            self.info_bias_assessment.setText("Not assessed")
            self.info_bias_assessment.setStyleSheet("")
        self.info_bias_explanation.setPlainText(
            s.get('bias_explanation') or "")

        # ── Populate Ratings tab ──
        self._populate_ratings(s)
        self._loading = False

    def _populate_ratings(self, src: dict):
        """Fill the section and topic ratings tables for a source."""
        read_only_flags = Qt.ItemFlag.ItemIsSelectable | Qt.ItemFlag.ItemIsEnabled

        # Section ratings
        self.section_ratings_table.setRowCount(0)
        section_rate = src.get('section_rate') or []
        if section_rate:
            # Build section names
            section_names = [f"Intro ({self.session.paper_topic or 'Topic'})"]
            for rs in self.session.results:
                section_names.append(rs.get('section', 'Untitled'))

            for i, rating in enumerate(section_rate):
                row = self.section_ratings_table.rowCount()
                self.section_ratings_table.insertRow(row)
                name = (section_names[i]
                        if i < len(section_names)
                        else f"Section {i}")
                name_item = QTableWidgetItem(name)
                name_item.setFlags(read_only_flags)
                self.section_ratings_table.setItem(row, 0, name_item)
                rating_item = QTableWidgetItem(str(rating))
                if rating >= 7:
                    rating_item.setForeground(QColor("#2ecc71"))
                elif rating <= 3:
                    rating_item.setForeground(QColor("#e74c3c"))
                self.section_ratings_table.setItem(row, 1, rating_item)

        # Topic ratings
        self.topic_ratings_table.setRowCount(0)
        topic_ratings = src.get('topic_ratings') or {}
        if topic_ratings:
            topic_names = {}
            for t in self.session.topics:
                top = t if isinstance(t, dict) else t
                topic_names[top.get('topic_id', '')] = top.get(
                    'title', 'Untitled')

            for tid, rating in topic_ratings.items():
                row = self.topic_ratings_table.rowCount()
                self.topic_ratings_table.insertRow(row)
                name = topic_names.get(tid, tid)
                name_item = QTableWidgetItem(name)
                name_item.setFlags(read_only_flags)
                self.topic_ratings_table.setItem(row, 0, name_item)
                rating_item = QTableWidgetItem(str(rating))
                if rating >= 7:
                    rating_item.setForeground(QColor("#2ecc71"))
                elif rating <= 3:
                    rating_item.setForeground(QColor("#e74c3c"))
                self.topic_ratings_table.setItem(row, 1, rating_item)

        # Stat ratings
        self.stat_ratings_table.setRowCount(0)
        stat_ratings = src.get('stat_ratings') or {}
        if stat_ratings:
            stat_names = {}
            for st in self.session.statistics:
                stat = st if isinstance(st, dict) else st
                stat_names[stat.get('stat_id', '')] = stat.get(
                    'question', 'Untitled')

            for sid, rating in stat_ratings.items():
                row = self.stat_ratings_table.rowCount()
                self.stat_ratings_table.insertRow(row)
                name = stat_names.get(sid, sid)
                name_item = QTableWidgetItem(name[:60])
                name_item.setFlags(read_only_flags)
                self.stat_ratings_table.setItem(row, 0, name_item)
                rating_item = QTableWidgetItem(str(rating))
                if rating >= 7:
                    rating_item.setForeground(QColor("#2ecc71"))
                elif rating <= 3:
                    rating_item.setForeground(QColor("#e74c3c"))
                self.stat_ratings_table.setItem(row, 1, rating_item)

    # ── Field save ───────────────────────────────────────────────────

    def _auto_save_field(self, field_name):
        """Auto-save a text field to the session whenever it changes."""
        if self._loading:
            return
        row = self.source_table.currentRow()
        if row < 0 or row >= len(self.session.sources):
            return
        widgets = {
            'abstract': self.abstract_edit,
            'full_text': self.fulltext_edit,
            'summary': self.summary_edit,
            'citation': self.info_citation,
        }
        if field_name in widgets:
            text = widgets[field_name].toPlainText()
        elif field_name == 'doi':
            text = self.info_doi.text()
        elif field_name == 'title':
            text = self.info_title.text()
        else:
            return
        if isinstance(self.session.sources[row], dict):
            self.session.sources[row][field_name] = text if text else None
        if field_name == 'title':
            # Keep the header label and table row in sync (guarded)
            num = self.session.sources[row].get('number', '')
            self.detail_title.setText(
                f"<b>[{num}] {text or 'Untitled'}</b>")
            self._loading = True
            title_item = self.source_table.item(row, 1)
            if title_item:
                title_item.setText((text or '')[:80])
            self._loading = False
        self.sources_changed.emit()

    def _auto_save_rating(self, key, table, row, col):
        """Auto-save list-based ratings (section_rate) when a cell is edited."""
        if self._loading or col != 1:
            return
        src_row = self.source_table.currentRow()
        if src_row < 0 or src_row >= len(self.session.sources):
            return
        src = self.session.sources[src_row]
        if not isinstance(src, dict):
            return
        item = table.item(row, col)
        if not item:
            return
        try:
            val = int(item.text())
            val = max(1, min(10, val))
        except ValueError:
            return
        ratings = src.get(key) or []
        while len(ratings) <= row:
            ratings.append(5)
        ratings[row] = val
        src[key] = ratings
        self.sources_changed.emit()

    def _auto_save_dict_rating(self, key, table, row, col):
        """Auto-save dict-based ratings (topic_ratings, stat_ratings) when a cell is edited."""
        if self._loading or col != 1:
            return
        src_row = self.source_table.currentRow()
        if src_row < 0 or src_row >= len(self.session.sources):
            return
        src = self.session.sources[src_row]
        if not isinstance(src, dict):
            return
        name_item = table.item(row, 0)
        val_item = table.item(row, col)
        if not name_item or not val_item:
            return
        try:
            val = int(val_item.text())
            val = max(1, min(10, val))
        except ValueError:
            return
        # Resolve the display name back to the ID
        rating_id = self._resolve_rating_id(key, name_item.text())
        if rating_id is None:
            return
        ratings = src.get(key) or {}
        ratings[rating_id] = val
        src[key] = ratings
        self.sources_changed.emit()

    def _resolve_rating_id(self, key, display_name):
        """Map a display name back to the original topic/stat ID."""
        if key == 'topic_ratings':
            for t in self.session.topics:
                top = t if isinstance(t, dict) else t
                if top.get('title', 'Untitled') == display_name:
                    return top.get('topic_id', '')
        elif key == 'stat_ratings':
            for st in self.session.statistics:
                stat = st if isinstance(st, dict) else st
                if stat.get('question', 'Untitled')[:60] == display_name:
                    return stat.get('stat_id', '')
        return None

    # ── Criteria management ─────────────────────────────────────────

    def _add_inclusion_criterion(self):
        text, ok = QInputDialog.getText(
            self, "Add Inclusion Criterion",
            "Criterion (paper must meet this to be included):")
        if ok and text.strip():
            item = QListWidgetItem(text.strip())
            item.setFlags(item.flags() | Qt.ItemFlag.ItemIsEditable)
            self.inclusion_list.addItem(item)
            self._sync_criteria_to_session()

    def _remove_inclusion_criterion(self):
        row = self.inclusion_list.currentRow()
        if row >= 0:
            self.inclusion_list.takeItem(row)
            self._sync_criteria_to_session()

    def _add_exclusion_criterion(self):
        text, ok = QInputDialog.getText(
            self, "Add Exclusion Criterion",
            "Criterion (any match → paper is excluded):")
        if ok and text.strip():
            item = QListWidgetItem(text.strip())
            item.setFlags(item.flags() | Qt.ItemFlag.ItemIsEditable)
            self.exclusion_list.addItem(item)
            self._sync_criteria_to_session()

    def _remove_exclusion_criterion(self):
        row = self.exclusion_list.currentRow()
        if row >= 0:
            self.exclusion_list.takeItem(row)
            self._sync_criteria_to_session()

    def _sync_criteria_to_session(self):
        """Push the current list-widget contents into the session."""
        self.session.inclusion_criteria = [
            self.inclusion_list.item(i).text()
            for i in range(self.inclusion_list.count())]
        self.session.exclusion_criteria = [
            self.exclusion_list.item(i).text()
            for i in range(self.exclusion_list.count())]
        self.sources_changed.emit()

    def _on_criterion_edited(self, item):
        """Save criteria whenever an item is edited inline."""
        if self._loading:
            return
        self._sync_criteria_to_session()

    def _build_criteria_text(self):
        """Format inclusion + exclusion criteria into a prompt-ready string."""
        parts = []
        inc = [self.inclusion_list.item(i).text()
               for i in range(self.inclusion_list.count())]
        exc = [self.exclusion_list.item(i).text()
               for i in range(self.exclusion_list.count())]
        if inc:
            parts.append(
                "INCLUSION CRITERIA (paper must meet ALL of these):\n"
                + "\n".join(f"- {c}" for c in inc))
        if exc:
            parts.append(
                "EXCLUSION CRITERIA (if ANY of these apply, exclude):\n"
                + "\n".join(f"- {c}" for c in exc))
        return "\n\n".join(parts)

    def _get_screening_text(self, src):
        """Return the text to send for screening based on the toggle."""
        title = src.get('title') or ''
        if self.screen_full_text_toggle.isChecked():
            #body = src.get('full_text') or src.get('abstract') or ''
            body = src.get('full_text') or ''
            label = 'Full Text'
        else:
            body = src.get('abstract') or ''
            label = 'Abstract'
        if not body and not title:
            return ''
        return f"Title: {title}\n{label}: {body}"

    def _open_doi(self):
        doi = self.info_doi.text().strip()
        if doi:
            url = (f"https://doi.org/{doi}"
                   if not doi.startswith('http') else doi)
            webbrowser.open(url)

    def _set_inclusion(self, included: bool):
        row = self.source_table.currentRow()
        if row < 0:
            return
        if isinstance(self.session.sources[row], dict):
            self.session.sources[row]['rating'] = included
            self.session.sources[row]['rate_explain'] = \
                'Manually set by user'
        self._refresh_table()
        self._on_source_selected(row)
        self.sources_changed.emit()

    def _ai_solo_screen(self):
        row = self.source_table.currentRow()
        if row < 0:
            return

        self._sync_criteria_to_session()
        criteria_text = self._build_criteria_text()
        has_criteria = bool(criteria_text)

        prompt_key = ('screen_source_criteria' if has_criteria
                      else 'screen_source_subjective')
        prompt_template = self.config.get_prompt(prompt_key)

        sample_vars = {
            'topic': self.session.paper_topic,
            'source_text': '(source text)',
        }
        if has_criteria:
            sample_vars['criteria'] = criteria_text

        dlg = PromptApprovalDialog(
            prompt_key, prompt_template, sample_vars, self.config, self)
        if not dlg.exec():
            return
        template = dlg.original_template

        s = self.session.sources[row]
        src = s if isinstance(s, dict) else s
        text = self._get_screening_text(src)
        if not text:
            return
        variables = {
            'topic': self.session.paper_topic,
            'source_text': text,
        }
        if has_criteria:
            variables['criteria'] = criteria_text
        formatted = template.format(**variables)
        try:
            import ast
            result = self.api.query(
                formatted, max_tokens=200, temperature=0.3)
            print(f"Result for AI screen: {result}")
            parsed = aggro_parse_reason(result)
            s['rating'] = bool(parsed[0])
            s['rate_explain'] = str(parsed[1])
            self.status_label.setText(
                f"Screened source {src.get('number')}: "
                f"{'Include' if parsed[0] else 'Exclude'}")
        except Exception as e:
            import traceback
            traceback.print_exc()
            print(f"Error screening source: {e}")
            #self.status_label.setText(
            #    f"Error screening source: {e}")
            s['rate_explain'] = result

        self._refresh_table()
        self._on_source_selected(row)
        self.sources_changed.emit()

    def _ai_solo_bias(self):
        """Assess risk of bias for the currently selected source."""
        row = self.source_table.currentRow()
        if row < 0:
            QMessageBox.warning(
                self, "No Selection",
                "Please select a source to assess.")
            return

        src = self.session.sources[row]
        source_text = src.get('full_text') or src.get('abstract') or ""
        if not source_text:
            QMessageBox.warning(
                self, "No Text",
                "This source has no full text or abstract to assess.")
            return

        title = src.get('title') or "Untitled"
        prompt_key = 'risk_of_bias'
        prompt_template = self.config.get_prompt(prompt_key)

        variables = {
            'topic': self.session.paper_topic,
            'title': title,
            'source_text': source_text,
        }

        dlg = PromptApprovalDialog(
            prompt_key, prompt_template, variables, self.config, self)
        if not dlg.exec():
            return
        template = dlg.original_template

        formatted = template.format(**variables)
        try:
            result = self.api.query(
                formatted, max_tokens=2000, temperature=0.3)
            print(f"Result for AI RoB: {result}")
            parsed = aggro_parse_bias(result)
            src['bias_assessment'] = str(parsed[0])
            src['bias_explanation'] = str(parsed[1])
            self.status_label.setText(
                f"RoB for source {src.get('number', row)}: {parsed[0]}")
        except Exception as e:
            import traceback
            traceback.print_exc()
            print(f"Error in solo RoB assessment: {e}")
            self.status_label.setText(
                f"Error assessing source {src.get('number', row)}: {e}")
            try:
                src['bias_explanation'] = result
            except Exception:
                pass

        self._on_source_selected(row)
        self.sources_changed.emit()

    def _clear_solo_bias(self):
        """Clear the risk of bias assessment for the currently selected source."""
        row = self.source_table.currentRow()
        if row < 0:
            return
        src = self.session.sources[row]
        src['bias_assessment'] = None
        src['bias_explanation'] = None
        self._on_source_selected(row)
        self.sources_changed.emit()

    def _add_manual_source(self):
        title, ok = QInputDialog.getText(
            self, "Add Manual Source", "Source title:")
        if not ok or not title.strip():
            return
        max_val = max(src["number"] for src in self.session.sources)
        source = {
            "number": max_val + 1,
            "title": title.strip(), "year": "", "journal": "",
            "abstract": "", "full_text": "", "summary": "",
            "citation": "", "doi": "", "pmid": "",
            "rating": None, "rate_explain": "",
        }
        self.session.sources.append(source)
        self._refresh_table()
        self.sources_changed.emit()

    def _remove_source(self):
        row = self.source_table.currentRow()
        if row < 0:
            QMessageBox.warning(
                self, "No Selection",
                "Please select a source to remove.")
            return
        src = self.session.sources[row]
        src_d = src if isinstance(src, dict) else src
        title = src_d.get('title', 'Untitled')[:50]
        num = src_d.get('number', '?')
        reply = QMessageBox.question(
            self, "Remove Source",
            f"Remove source [{num}] \"{title}\"?\n\n"
            f"Note: this will not update existing citation numbers "
            f"in the review text. You may need to fix citations "
            f"manually or re-export with reordering.",
            QMessageBox.StandardButton.Yes |
            QMessageBox.StandardButton.No)
        if reply == QMessageBox.StandardButton.Yes:
            # Clean up citation_validations for this source number
            num_str = str(num)
            self.session.citation_validations.pop(num_str, None)
            # Clean up section_contexts referencing this source
            for cfg in self.session.section_contexts.values():
                for key in ('summaries', 'full_texts'):
                    ref_list = cfg.get(key, [])
                    if num in ref_list:
                        ref_list.remove(num)
            del self.session.sources[row]
            self._refresh_table()
            self.sources_changed.emit()

    # ── Search terms ─────────────────────────────────────────────────

    def _generate_search_terms(self):
        if not self.session.paper_topic:
            QMessageBox.warning(
                self, "No Topic",
                "Please set a paper topic in the Main tab first.")
            return

        prompt_template = self.config.get_prompt('suggest_search_terms')
        variables = {'topic': self.session.paper_topic}
        dlg = PromptApprovalDialog(
            'suggest_search_terms', prompt_template,
            variables, self.config, self)
        if dlg.exec():
            formatted = dlg.get_formatted_prompt()
            try:
                result = self.api.query(
                    formatted, max_tokens=1000, temperature=0.5)
                terms = [t.strip() for t in result.split(';') if t.strip()]
                self.session.search_terms = terms
                self._per_term_limits.clear()
                self._refresh_search_terms_table()
                self.sources_changed.emit()
            except Exception as e:
                QMessageBox.critical(self, "Error", str(e))

    def _add_search_term(self):
        term, ok = QInputDialog.getText(
            self, "Add Search Term", "Boolean search term:")
        if ok and term:
            self.session.search_terms.append(term)
            self._refresh_search_terms_table()
            self.sources_changed.emit()

    def _remove_search_term(self):
        row = self.search_terms_table.currentRow()
        if row >= 0:
            item = self.search_terms_table.item(row, 0)
            if item:
                term = item.text()
                self._per_term_limits.pop(term, None)
            if row < len(self.session.search_terms):
                del self.session.search_terms[row]
            self._refresh_search_terms_table()
            self.sources_changed.emit()

    # ── Retrieval ────────────────────────────────────────────────────

    def _retrieve_sources(self):
        if not self.config.ncbi_api_key:
            QMessageBox.warning(
                self, "No NCBI Key",
                "Please set your NCBI API key in Prompt Settings.")
            return
        if not self.session.search_terms:
            QMessageBox.warning(
                self, "No Search Terms",
                "Please generate or add search terms first.")
            return

        # Build per-term limits from table
        per_term = {}
        for row in range(self.search_terms_table.rowCount()):
            item = self.search_terms_table.item(row, 0)
            widget = self.search_terms_table.cellWidget(row, 1)
            if item and isinstance(widget, QSpinBox):
                per_term[item.text()] = widget.value()

        self.status_label.setText("Retrieving sources...")
        self.progress_bar.setVisible(True)
        self.progress_bar.setRange(0, 0)

        self._retrieval_worker = SourceRetrievalWorker(
            self.config.ncbi_api_key,
            self.session.search_terms,
            self.default_retmax.value(),
            per_term_limits=per_term if per_term else None)
        self._retrieval_worker.progress.connect(
            lambda msg: self.status_label.setText(msg))
        self._retrieval_worker.finished.connect(self._on_retrieval_done)
        self._retrieval_worker.error.connect(self._on_retrieval_error)
        self._retrieval_worker.start()

    def _fill_out_sources(self):
        """Kick off NCBI lookups for every source that has a DOI,
        queuing them sequentially so each finishes before the next starts."""
        if not self.config.ncbi_api_key:
            QMessageBox.warning(
                self, "No NCBI Key",
                "Please set your NCBI API key in Prompt Settings before "
                "using this feature.")
            return

        # Build the queue: collect (row_index) for sources that have a DOI,
        # looking up the source number from the '#' column (column 0).
        self._fill_queue = []
        for row in range(self.source_table.rowCount()):
            num_item = self.source_table.item(row, 0)
            if not num_item:
                continue
            try:
                source_number = int(num_item.text())
            except (ValueError, TypeError):
                continue
            # Find the session index for this source number
            src_idx = None
            for idx, s in enumerate(self.session.sources):
                if s.get('number') == source_number:
                    src_idx = idx
                    break
            if src_idx is None:
                continue
            doi = (self.session.sources[src_idx].get('doi') or "").strip()
            if not doi:
                continue
            self._fill_queue.append(src_idx)

        if not self._fill_queue:
            QMessageBox.information(
                self, "Nothing to Fill",
                "No sources with a DOI were found to look up.")
            return

        self.status_label.setText(
            f"Filling out {len(self._fill_queue)} sources...")
        self._fill_next_in_queue()

    def _fill_next_in_queue(self):
        """Process the next source in the fill queue."""
        if not self._fill_queue:
            self.status_label.setText("Fill-out complete.")
            return
        row = self._fill_queue.pop(0)
        src = self.session.sources[row]
        doi = (src.get('doi') or "").strip()
        source_number = src.get('number', row + 1)
        self.status_label.setText(
            f"Looking up Source {source_number}... "
            f"({len(self._fill_queue)} remaining)")

        worker = SourceInfoFillWorker(
            self.config.ncbi_api_key, row, doi)
        worker.finished.connect(self._on_fill_all_done)
        worker.error.connect(self._on_fill_all_error)
        self._fill_info_worker = worker
        worker.start()

    def _on_fill_all_done(self, row: int, fields: dict):
        """Handle completion of one fill-out, then process next in queue."""
        self._on_fill_info_done(row, fields)
        self._fill_next_in_queue()

    def _on_fill_all_error(self, row: int, msg: str):
        """Handle failure of one fill-out, then continue with next."""
        self._on_fill_info_error(row, msg)
        self._fill_next_in_queue()


    def _on_retrieval_done(self, sources, textless, abless):
        self.progress_bar.setVisible(False)
        self.session.sources = [
            s.to_dict() if hasattr(s, 'to_dict') else s for s in sources]
        self._refresh_table()
        self.status_label.setText(
            f"Retrieved {len(sources)} sources. "
            f"{len(textless)} missing full text, "
            f"{len(abless)} missing abstracts.")
        self.sources_changed.emit()

    def _on_retrieval_error(self, msg):
        self.progress_bar.setVisible(False)
        self.status_label.setText(f"Error: {msg}")
        QMessageBox.critical(self, "Retrieval Error", msg)

    # ── Summarization ────────────────────────────────────────────────

    def _batch_summarize(self):
        sources_to_summarize = []
        for i, s in enumerate(self.session.sources):
            src = s if isinstance(s, dict) else s
            if src.get('rating') is False:
                continue
            if src.get('summary'):
                continue
            if src.get('abstract') or src.get('full_text'):
                sources_to_summarize.append(i)

        if not sources_to_summarize:
            QMessageBox.information(
                self, "Nothing to Summarize",
                "All applicable sources already have summaries.")
            return
        prompt_template = self.config.get_prompt('summarize_source')
        sample_text = "(source text will be inserted here)"
        dlg = PromptApprovalDialog(
            'summarize_source', prompt_template,
            {'source_text': sample_text}, self.config, self)
        if not dlg.exec():
            return

        template = dlg.original_template
        self.status_label.setText(
            f"Summarizing {len(sources_to_summarize)} sources...")

        for idx in sources_to_summarize:
            print(f"Summarizing source {idx}")
            src = self.session.sources[idx]
            text = src.get('full_text') or src.get('abstract') or ""
            if not text:
                continue
            formatted = template.format(source_text=text)
            try:
                result = self.api.query(
                    formatted, max_tokens=2000, temperature=0.3)
                self.session.sources[idx]['summary'] = result
                self.status_label.setText(
                    f"Summarized source {src.get('number', idx)}")
            except Exception as e:
                self.status_label.setText(
                    f"Error on source {src.get('number', idx)}: {e}")

        self._refresh_table()
        self.sources_changed.emit()
        self.status_label.setText("Batch summarization complete.")

    def _generate_summary_single(self):
        row = self.source_table.currentRow()
        if row < 0:
            return
        src = self.session.sources[row]
        text = src.get('full_text') or src.get('abstract') or ""
        if not text:
            QMessageBox.warning(
                self, "No Text",
                "This source has no text to summarize.")
            return

        prompt_template = self.config.get_prompt('summarize_source')
        variables = {'source_text': text}
        dlg = PromptApprovalDialog(
            'summarize_source', prompt_template,
            variables, self.config, self)
        if dlg.exec():
            formatted = dlg.get_formatted_prompt()
            try:
                result = self.api.query(
                    formatted, max_tokens=2000, temperature=0.3)
                self.session.sources[row]['summary'] = result
                self.summary_edit.setPlainText(result)
                self._refresh_table()
                self.sources_changed.emit()
            except Exception as e:
                QMessageBox.critical(self, "Error", str(e))

    # ── Screening ────────────────────────────────────────────────────

    def _screen_inclusion(self):
        self._sync_criteria_to_session()
        criteria_text = self._build_criteria_text()
        has_criteria = bool(criteria_text)

        prompt_key = ('screen_source_criteria' if has_criteria
                      else 'screen_source_subjective')
        prompt_template = self.config.get_prompt(prompt_key)

        sample_vars = {
            'topic': self.session.paper_topic,
            'source_text': '(source text)',
        }
        if has_criteria:
            sample_vars['criteria'] = criteria_text

        dlg = PromptApprovalDialog(
            prompt_key, prompt_template, sample_vars, self.config, self)
        if not dlg.exec():
            return
        template = dlg.original_template

        for i, s in enumerate(self.session.sources):
            src = s if isinstance(s, dict) else s
            if src.get('rating') is not None:
                continue
            text = self._get_screening_text(src)
            if not text:
                continue
            variables = {
                'topic': self.session.paper_topic,
                'source_text': text,
            }
            if has_criteria:
                variables['criteria'] = criteria_text
            formatted = template.format(**variables)
            try:
                import ast
                result = self.api.query(
                    formatted, max_tokens=200, temperature=0.3)
                parsed = aggro_parse_reason(result)
                self.session.sources[i]['rating'] = bool(parsed[0])
                self.session.sources[i]['rate_explain'] = str(parsed[1])
                self.status_label.setText(
                    f"Screened source {src.get('number', i)}: "
                    f"{'Include' if parsed[0] else 'Exclude'}")
                print(f'Screening source: {i}; Inclusion verdict: {result}')
            except Exception as e:
                print(f"Error in response: {e}")
                self.status_label.setText(
                    f"Error screening source {src.get('number', i)}: {e}")
                self.session.sources[i]['rate_explain'] = result

        self._refresh_table()
        self.sources_changed.emit()
        self.status_label.setText("Screening complete.")

    # ── Section rating ───────────────────────────────────────────────

    def _rate_for_sections(self):
        if not self.session.results:
            QMessageBox.warning(
                self, "No Results Sections",
                "Please generate results topics first in the Main tab.")
            return

        all_topics = [f"An introduction about {self.session.paper_topic}"]
        for rs in self.session.results:
            all_topics.append(rs.get('section', 'Untitled'))

        prompt_template = self.config.get_prompt('rate_source_sections')
        # Show prompt approval with a sample batch
        sample_batch = all_topics[:10]
        sample_vars = {
            'topics': str(sample_batch),
            'journal': '(journal)',
            'summary': '(summary)',
        }
        dlg = PromptApprovalDialog(
            'rate_source_sections', prompt_template,
            sample_vars, self.config, self)
        if not dlg.exec():
            return
        template = dlg.original_template

        # Batch the topics into chunks of 10
        BATCH_SIZE = 10
        batches = []
        for start in range(0, len(all_topics), BATCH_SIZE):
            batches.append(
                (start, all_topics[start:start + BATCH_SIZE]))

        for i, s in enumerate(self.session.sources):
            src = s if isinstance(s, dict) else s
            # Skip explicitly excluded sources
            if src.get('rating') is False:
                continue
            summary = src.get('summary')
            if not summary:
                continue

            # Collect ratings across all batches for this source
            all_ratings = []
            batch_failed = False

            for batch_start, batch_topics in batches:
                variables = {
                    'topics': str(batch_topics),
                    'journal': src.get('journal', 'Unknown'),
                    'summary': summary,
                }
                formatted = template.format(**variables)
                try:
                    import ast
                    result = self.api.query(formatted, max_tokens=200)
                    ratings = ast.literal_eval(result)
                    batch_ratings = [int(r) for r in ratings]
                    # Pad or truncate to match batch size
                    while len(batch_ratings) < len(batch_topics):
                        batch_ratings.append(5)
                    batch_ratings = batch_ratings[:len(batch_topics)]
                    all_ratings.extend(batch_ratings)
                except Exception as e:
                    self.status_label.setText(
                        f"Error rating source "
                        f"{src.get('number', i)} batch: {e}")
                    # Fill with neutral ratings so indices stay aligned
                    all_ratings.extend([5] * len(batch_topics))
                    batch_failed = True

            self.session.sources[i]['section_rate'] = all_ratings
            status = ("(partial)" if batch_failed else "")
            self.status_label.setText(
                f"Rated source {src.get('number', i)} "
                f"across {len(batches)} batch(es) {status}")

        self.sources_changed.emit()
        self.status_label.setText("Section rating complete.")

    # ── Risk of Bias Assessment ────────────────────────────────────

    def _risk_of_bias(self):
        """Assess risk of bias for all included sources using the
        Cochrane RoB 2 tool via the configured LLM."""

        QMessageBox.information(self, "Note:", "By Default this will Assess using Cochrane RoB 2 tool. This tool is mainly to appraise randomized control trials. The default prompt is designed to be broad and nonspecific. For higher specificity to some source or for other study types, please edit the prompt as needed.")

        included = [
            (i, s) for i, s in enumerate(self.session.sources)
            if s.get('rating') is True
        ]
        if not included:
            QMessageBox.warning(
                self, "No Included Sources",
                "No sources have been rated for inclusion (rating = True).\n"
                "Please screen sources for inclusion first.")
            return

        # Filter to those that have full text or abstract to assess
        assessable = [
            (i, s) for i, s in included
            if s.get('full_text') or s.get('abstract')
        ]
        if not assessable:
            QMessageBox.warning(
                self, "No Assessable Sources",
                "None of the included sources have full text or abstract "
                "available for bias assessment.")
            return

        prompt_key = 'risk_of_bias'
        prompt_template = self.config.get_prompt(prompt_key)

        sample_vars = {
            'topic': self.session.paper_topic,
            'title': '(source title)',
            'source_text': '(source text will be inserted here)',
        }

        dlg = PromptApprovalDialog(
            prompt_key, prompt_template, sample_vars, self.config, self)
        if not dlg.exec():
            return
        template = dlg.original_template

        self.status_label.setText(
            f"Assessing risk of bias for {len(assessable)} sources...")

        for idx, src in assessable:
            source_text = src.get('full_text') or src.get('abstract') or ""
            title = src.get('title') or "Untitled"
            variables = {
                'topic': self.session.paper_topic,
                'title': title,
                'source_text': source_text,
            }
            formatted = template.format(**variables)
            try:
                result = self.api.query(
                    formatted, max_tokens=2000, temperature=0.3)
                parsed = aggro_parse_bias(result)
                self.session.sources[idx]['bias_assessment'] = str(parsed[0])
                self.session.sources[idx]['bias_explanation'] = str(parsed[1])
                self.status_label.setText(
                    f"Assessed source {src.get('number', idx)}: {parsed[0]}")
                print(f"RoB source {src.get('number', idx)}: {parsed[0]}")
            except Exception as e:
                print(f"Error in RoB assessment for source "
                      f"{src.get('number', idx)}: {e}")
                self.status_label.setText(
                    f"Error assessing source {src.get('number', idx)}: {e}")
                # Store the raw result as explanation if parsing failed
                try:
                    self.session.sources[idx]['bias_explanation'] = result
                except Exception:
                    pass

        self._refresh_table()
        self.sources_changed.emit()
        self.status_label.setText("Risk of bias assessment complete.")

    # ── Citation linking ─────────────────────────────────────────────

    def show_sources_for_citations(self, numbers: list):
        for row in range(self.source_table.rowCount()):
            item = self.source_table.item(row, 0)
            if item and int(item.text()) in numbers:
                self.source_table.selectRow(row)
                self.source_table.scrollToItem(item)
                break

    def navigate_to_source_excerpt(self, source_number: int, excerpt: str,
                                   detail_tab_index: int):
        """Select a source, switch to the appropriate detail tab, and
        find + highlight the excerpt text (same technique as Ctrl+F)."""
        from PyQt6.QtGui import QTextCursor

        # 1. Select the source row in the table
        for row in range(self.source_table.rowCount()):
            item = self.source_table.item(row, 0)
            if item and int(item.text()) == source_number:
                self.source_table.selectRow(row)
                self.source_table.scrollToItem(item)
                break

        # 2. Switch to the correct detail tab (Summary=2, Full Text=1)
        self.detail_tabs.setCurrentIndex(detail_tab_index)

        # 3. Determine which QTextEdit to search in
        if detail_tab_index == 2:
            target_edit = self.summary_edit
        elif detail_tab_index == 1:
            target_edit = self.fulltext_edit
        else:
            return

        if not excerpt or not excerpt.strip():
            return

        search_text = excerpt.strip()

        # 4. Use QTextEdit.find() — same as FindDialog / Ctrl+F.
        #    This natively selects the match and scrolls it into view.
        cursor = target_edit.textCursor()
        cursor.movePosition(QTextCursor.MoveOperation.Start)
        target_edit.setTextCursor(cursor)

        found = target_edit.find(search_text)

        # If full excerpt not found, try progressively shorter prefixes
        if not found and len(search_text) > 80:
            for length in [200, 150, 100, 60, 40]:
                if len(search_text) > length:
                    cursor = target_edit.textCursor()
                    cursor.movePosition(QTextCursor.MoveOperation.Start)
                    target_edit.setTextCursor(cursor)
                    found = target_edit.find(search_text[:length])
                    if found:
                        break

        if found:
            target_edit.ensureCursorVisible()
            target_edit.setFocus()

    def refresh_from_session(self):
        self._load_from_session()


def _strip_code_fences(text: str) -> str:
    """Remove markdown code fences (```python ... ```) from LLM output."""
    import re
    raw = text.strip()
    # Strip opening fence like ```python or ```json or just ```
    raw = re.sub(r'^```\w*\s*\n?', '', raw)
    # Strip closing fence
    raw = re.sub(r'\n?```\s*$', '', raw)
    return raw.strip()


def aggro_parse_reason(result):
    raw = _strip_code_fences(result)

    if raw.startswith("[True,"):
        flag = True
        reason = raw[len("[True,"):].strip().rstrip("]").strip().strip("'\"")
        parsed = [flag, reason]
    elif raw.startswith("[False,"):
        flag = False
        reason = raw[len("[False,"):].strip().rstrip("]").strip().strip("'\"")
        parsed = [flag, reason]
    else:
        raise ValueError(f"Unexpected AI response format: {raw}")
    return parsed


def aggro_parse_bias(result):
    """Parse a risk of bias response in the format [judgement, 'explanation'].

    Expected judgements: 'Low', 'Some concerns', 'High'.
    Handles markdown code fences, varied quoting, and long explanations
    that contain commas and special characters.
    """
    import ast
    import re

    raw = _strip_code_fences(result)

    # Try ast.literal_eval first (works when the LLM produces clean Python)
    try:
        parsed = ast.literal_eval(raw)
        if isinstance(parsed, (list, tuple)) and len(parsed) >= 2:
            return [str(parsed[0]), str(parsed[1])]
    except (ValueError, SyntaxError):
        pass

    # Manual parsing: find the judgement at the start of a list, then
    # take everything after the first comma as the explanation.
    # This handles explanations that themselves contain commas / quotes.
    for judgement in ('Low', 'Some concerns', 'High'):
        for pattern in (
            f'["{judgement}"',
            f"['{judgement}'",
            f"[{judgement}",
        ):
            if raw.startswith(pattern):
                rest = raw[len(pattern):].lstrip()
                if rest.startswith(','):
                    rest = rest[1:].strip()
                # Strip outer brackets and quotes from explanation
                if rest.endswith(']'):
                    rest = rest[:-1].strip()
                rest = rest.strip().strip("'\"")
                return [judgement, rest]

    # Last resort: regex to find the list anywhere in the string
    m = re.search(
        r'\[\s*["\']?(Low|Some concerns|High)["\']?\s*,\s*["\']?(.*?)["\']?\s*\]\s*$',
        raw, re.DOTALL)
    if m:
        return [m.group(1), m.group(2).strip()]

    raise ValueError(f"Unexpected AI bias response format: {raw[:200]}")
````
