## Supplementary material for "In Search of Ethical Procedures for LLM-Assisted Systematic Review Production: A Proof-of-Concept Evaluation of Selected Review Components": source_data: topics_stats_tab.py

```
"""
Topics Tab — for research-question-driven evidence gathering.
Statistics Tab (Beta) — for numerical data extraction.

Both tabs support:
  - Bulk mode: one prompt with all context sources at once (default).
  - Iterative mode: loops through each source individually, assembling
    a growing response. Citations are automatic since each prompt sees
    only one source.
  - Distill: condenses a verbose topic/stat into concise text while
    keeping all citations intact.
"""
import uuid
from PyQt6.QtWidgets import (
    QWidget, QVBoxLayout, QHBoxLayout, QSplitter, QListWidget, QListWidgetItem,
    QTextEdit, QLabel, QPushButton, QGroupBox, QLineEdit, QMessageBox,
    QInputDialog, QCheckBox, QComboBox, QSpinBox, QProgressBar
)
from PyQt6.QtCore import Qt, pyqtSignal
from PyQt6.QtGui import QFont

from dialogs import (
    PromptApprovalDialog, OutputApprovalDialog, ContextConfigDialog,
    LinkToSectionsDialog,
)


# ─── helpers ─────────────────────────────────────────────────────────

def _top_n_sources_for_topic(session, topic_id: str, n: int = 10):
    """
    Return a dict {source_number: summary} of the top-n rated sources
    for a given topic_id.  Falls back to included/unscreened summaries
    if no ratings exist.  Never includes explicitly rejected sources
    in the fallback.
    """
    rated, unrated = [], []
    for s in session.sources:
        src = s if isinstance(s, dict) else s
        summary = src.get('summary')
        if not summary:
            continue
        # Never include rejected sources in fallback
        is_excluded = src.get('rating') is False
        num = src.get('number', 0)
        tr = src.get('topic_ratings') or {}
        rating = tr.get(topic_id, 0)
        if rating > 0:
            rated.append((num, summary, rating))
        elif not is_excluded:
            unrated.append((num, summary))

    if rated:
        rated.sort(key=lambda x: x[2], reverse=True)
        return {num: summ for num, summ, _ in rated[:n]}
    else:
        return {num: summ for num, summ in unrated[:n]}


def _top_n_source_list(session, topic_id: str, n: int = 10):
    """
    Like _top_n_sources_for_topic but returns a list of (num, summary)
    preserving order for iteration.  Never includes explicitly rejected
    sources in the fallback.
    """
    rated, unrated = [], []
    for s in session.sources:
        src = s if isinstance(s, dict) else s
        summary = src.get('summary')
        if not summary:
            summary = src.get('full_text')
        is_excluded = src.get('rating') is False
        num = src.get('number', 0)
        tr = src.get('topic_ratings') or {}
        rating = tr.get(topic_id, 0)
        if rating > 0:
            rated.append((num, summary, rating))
        elif not is_excluded:
            unrated.append((num, summary))

    if rated:
        rated.sort(key=lambda x: x[2], reverse=True)
        return [(num, summ) for num, summ, _ in rated[:n]]
    else:
        return unrated[:n]


def _top_n_sources_for_stat(session, stat_id: str, n: int = 10):
    """
    Return a dict {source_number: summary} of the top-n rated sources
    for a given stat_id. Falls back to included/unscreened summaries
    if no ratings exist.  Never includes explicitly rejected sources
    in the fallback.
    """
    rated, unrated = [], []
    for s in session.sources:
        src = s if isinstance(s, dict) else s
        summary = src.get('summary')
        if not summary:
            continue
        is_excluded = src.get('rating') is False
        num = src.get('number', 0)
        sr = src.get('stat_ratings') or {}
        rating = sr.get(stat_id, 0)
        if rating > 0:
            rated.append((num, summary, rating))
        elif not is_excluded:
            unrated.append((num, summary))

    if rated:
        rated.sort(key=lambda x: x[2], reverse=True)
        return {num: summ for num, summ, _ in rated[:n]}
    else:
        return {num: summ for num, summ in unrated[:n]}


def _stat_source_list(session, stat_id: str = '', n: int = 10):
    """List of (num, summary) for stats iteration, sorted by stat_ratings.
    Never includes explicitly rejected sources in the fallback."""
    rated, unrated = [], []
    for s in session.sources:
        src = s if isinstance(s, dict) else s
        summary = src.get('summary')
        if not summary:
            continue
        is_excluded = src.get('rating') is False
        num = src.get('number', 0)
        sr = src.get('stat_ratings') or {}
        rating = sr.get(stat_id, 0)
        if rating > 0:
            rated.append((num, summary, rating))
        elif not is_excluded:
            unrated.append((num, summary))

    if rated:
        rated.sort(key=lambda x: x[2], reverse=True)
        return [(num, summ) for num, summ, _ in rated[:n]]
    else:
        return unrated[:n]


# ═══════════════════════════════════════════════════════════════════
#  Topics Tab
# ═══════════════════════════════════════════════════════════════════

class TopicsTab(QWidget):
    """Tab for managing research topics with evidence from sources."""

    topics_changed = pyqtSignal()

    def __init__(self, config_manager, api_manager, session, parent=None):
        super().__init__(parent)
        self.config = config_manager
        self.api = api_manager
        self.session = session
        self._build_ui()
        self._load_from_session()

    def _build_ui(self):
        layout = QVBoxLayout(self)

        # Toolbar
        toolbar = QHBoxLayout()
        add_btn = QPushButton("+ Add Topic")
        add_btn.clicked.connect(self._add_topic)
        toolbar.addWidget(add_btn)

        auto_btn = QPushButton("✦ Auto-Generate Topics Based on Accepted Summaries")
        auto_btn.clicked.connect(self._auto_generate)
        toolbar.addWidget(auto_btn)

        remove_btn = QPushButton("- Remove Topic")
        remove_btn.clicked.connect(self._remove_topic)
        toolbar.addWidget(remove_btn)

        rate_btn = QPushButton("✦ Rate Sources for Topics")
        rate_btn.clicked.connect(self._rate_sources_for_topics)
        toolbar.addWidget(rate_btn)

        gen_q_btn = QPushButton("✦ AI-Generate Questions Based on Paper Contents")
        gen_q_btn.setToolTip(
            "Ask the AI to generate topic research questions based on "
            "the paper topic, thesis, and results subsections")
        gen_q_btn.clicked.connect(self._ai_generate_questions)
        toolbar.addWidget(gen_q_btn)

        toolbar.addStretch()
        layout.addLayout(toolbar)

        # Main area
        splitter = QSplitter(Qt.Orientation.Horizontal)

        # Left: topic list
        self.topic_list = QListWidget()
        self.topic_list.currentRowChanged.connect(self._on_topic_selected)
        splitter.addWidget(self.topic_list)

        # Right: topic detail
        right = QWidget()
        right_layout = QVBoxLayout(right)

        self.topic_title_edit = QLineEdit()
        self.topic_title_edit.setPlaceholderText(
            "Topic / Research Question")
        self.topic_title_edit.textChanged.connect(self._on_title_changed)
        right_layout.addWidget(self.topic_title_edit)

        # ── Action row 1: write + context + link ──
        btn_row = QHBoxLayout()
        write_btn = QPushButton("✦ AI Write Topic")
        write_btn.clicked.connect(self._ai_write_topic)
        btn_row.addWidget(write_btn)

        distill_btn = QPushButton("✦ Distill")
        distill_btn.setToolTip(
            "Ask the AI to condense this topic text while keeping "
            "all citations and key data intact")
        distill_btn.clicked.connect(self._distill_topic)
        btn_row.addWidget(distill_btn)

        context_btn = QPushButton("📋 Context")
        context_btn.clicked.connect(self._configure_context)
        btn_row.addWidget(context_btn)

        link_btn = QPushButton("🔗 Link to Sections")
        link_btn.clicked.connect(self._link_to_sections)
        btn_row.addWidget(link_btn)
        btn_row.addStretch()
        right_layout.addLayout(btn_row)

        # ── Action row 2: mode toggle + top-N ──
        mode_row = QHBoxLayout()

        self.iterative_cb = QCheckBox("Iterative mode")
        self.iterative_cb.setToolTip(
            "When checked, the AI examines each source individually and "
            "builds up the response source-by-source. Citations are "
            "automatic. When unchecked (bulk), all sources go in one prompt.")
        mode_row.addWidget(self.iterative_cb)

        mode_row.addWidget(QLabel("Top N sources:"))
        self.topic_top_n = QSpinBox()
        self.topic_top_n.setRange(1, 999)
        self.topic_top_n.setValue(10)
        self.topic_top_n.setToolTip(
            "How many top-rated sources to include when generating "
            "this topic (if ratings exist)")
        mode_row.addWidget(self.topic_top_n)

        mode_row.addStretch()
        right_layout.addLayout(mode_row)

        # ── Progress bar (for iterative) ──
        self.topic_progress = QProgressBar()
        self.topic_progress.setVisible(False)
        self.topic_progress.setTextVisible(True)
        right_layout.addWidget(self.topic_progress)

        # ── Text editor ──
        self.topic_text_edit = QTextEdit()
        self.topic_text_edit.setFont(QFont("Georgia", 11))
        self.topic_text_edit.textChanged.connect(self._on_text_changed)
        right_layout.addWidget(self.topic_text_edit)

        splitter.addWidget(right)
        splitter.setSizes([250, 550])
        layout.addWidget(splitter)

    # ── Session I/O ──────────────────────────────────────────────────

    def _load_from_session(self):
        self.topic_list.clear()
        for t in self.session.topics:
            top = t if isinstance(t, dict) else t
            self.topic_list.addItem(
                f"{top.get('topic_id', '?')}: "
                f"{top.get('title', 'Untitled')}")

    def _on_topic_selected(self, row):
        if row < 0 or row >= len(self.session.topics):
            return
        t = self.session.topics[row]
        self.topic_title_edit.blockSignals(True)
        self.topic_title_edit.setText(t.get('title', ''))
        self.topic_title_edit.blockSignals(False)
        self.topic_text_edit.blockSignals(True)
        self.topic_text_edit.setPlainText(t.get('text') or '')
        self.topic_text_edit.blockSignals(False)

    def _on_title_changed(self, text):
        row = self.topic_list.currentRow()
        if 0 <= row < len(self.session.topics):
            self.session.topics[row]['title'] = text
            self.topic_list.item(row).setText(
                f"{self.session.topics[row].get('topic_id', '?')}: {text}")
            self.topics_changed.emit()

    def _on_text_changed(self):
        row = self.topic_list.currentRow()
        if 0 <= row < len(self.session.topics):
            self.session.topics[row]['text'] = \
                self.topic_text_edit.toPlainText()
            self.topics_changed.emit()

    def _add_topic(self):
        title, ok = QInputDialog.getText(
            self, "New Topic", "Research question / topic:")
        if ok and title:
            tid = f"T{len(self.session.topics) + 1}"
            self.session.topics.append({
                'topic_id': tid, 'title': title, 'text': None,
                'linked_sections': [], 'context_config': None
            })
            self.topic_list.addItem(f"{tid}: {title}")
            self.topics_changed.emit()

    def _remove_topic(self):
        row = self.topic_list.currentRow()
        if row >= 0:
            topic_id = self.session.topics[row].get('topic_id', '')
            if topic_id:
                # Clean up topic_ratings on every source
                for s in self.session.sources:
                    src = s if isinstance(s, dict) else s
                    tr = src.get('topic_ratings')
                    if tr and topic_id in tr:
                        del tr[topic_id]
                # Clean up section_contexts referencing this topic
                for cfg in self.session.section_contexts.values():
                    topic_list = cfg.get('topics', [])
                    if topic_id in topic_list:
                        topic_list.remove(topic_id)
            self.topic_list.takeItem(row)
            del self.session.topics[row]
            self.topics_changed.emit()

    def _auto_generate(self):
        if not self.session.paper_topic:
            QMessageBox.warning(self, "No Topic",
                                "Set a paper topic first.")
            return
        source_dict = {}
        for s in self.session.sources:
            src = s if isinstance(s, dict) else s
            if src.get('rating') is False:
                continue
            if src.get('summary'):
                source_dict[src.get('number', 0)] = src['summary']
            if len(source_dict) > 100: # Limit to stop AI from losing context
                break

        thesis_clause = ""
        if self.session.thesis:
            thesis_clause = f"The thesis is: {self.session.thesis}. "

        variables = {
            'topic': self.session.paper_topic,
            'thesis_clause': thesis_clause,
            'context': str(source_dict),
        }
        prompt_template = self.config.get_prompt('auto_generate_topics')
        dlg = PromptApprovalDialog(
            'auto_generate_topics', prompt_template,
            variables, self.config, self)
        if dlg.exec():
            formatted = dlg.get_formatted_prompt()
            try:
                result = self.api.query(formatted, max_tokens=1000)
                topics = [t.strip() for t in result.split(';') if t.strip()]
                for title in topics:
                    tid = f"T{len(self.session.topics) + 1}"
                    self.session.topics.append({
                        'topic_id': tid, 'title': title, 'text': None,
                        'linked_sections': [], 'context_config': None
                    })
                self._load_from_session()
                self.topics_changed.emit()
            except Exception as e:
                QMessageBox.critical(self, "Error", str(e))

    # ── AI-generate research questions ──────────────────────────────

    def _ai_generate_questions(self):
        """Use AI to generate topic research questions from paper context."""
        if not self.session.paper_topic:
            QMessageBox.warning(self, "No Topic",
                                "Set a paper topic first.")
            return

        # Build context from paper topic, thesis, and results headers
        context_parts = [f"Paper topic: {self.session.paper_topic}"]
        if self.session.thesis:
            context_parts.append(f"Thesis: {self.session.thesis}")
        if self.session.results:
            headers = [rs.get('section', 'Untitled')
                       for rs in self.session.results]
            context_parts.append(
                f"Results subsection headers: {'; '.join(headers)}")

        context = "\n".join(context_parts)

        prompt_template = self.config.get_prompt(
            'auto_generate_topic_questions')
        variables = {
            'topic': self.session.paper_topic,
            'context': context,
        }
        dlg = PromptApprovalDialog(
            'auto_generate_topic_questions', prompt_template,
            variables, self.config, self)
        if dlg.exec():
            formatted = dlg.get_formatted_prompt()
            try:
                result = self.api.query(formatted, max_tokens=1500)
                questions = [q.strip()
                             for q in result.split(';') if q.strip()]
                reply = QMessageBox.question(
                    self, "Generated Topic Questions",
                    f"Generated {len(questions)} questions:\n\n" +
                    "\n".join(f"  {i+1}. {q}"
                              for i, q in enumerate(questions)) +
                    "\n\nAdd these as new topics?",
                    QMessageBox.StandardButton.Yes |
                    QMessageBox.StandardButton.No)
                if reply == QMessageBox.StandardButton.Yes:
                    for q in questions:
                        tid = f"T{len(self.session.topics) + 1}"
                        self.session.topics.append({
                            'topic_id': tid, 'title': q,
                            'text': None, 'linked_sections': [],
                            'context_config': None,
                        })
                    self._load_from_session()
                    self.topics_changed.emit()
            except Exception as e:
                QMessageBox.critical(self, "Error", str(e))

    # ── AI Write (bulk vs iterative) ─────────────────────────────────

    def _ai_write_topic(self):
        row = self.topic_list.currentRow()
        if row < 0:
            return
        if self.iterative_cb.isChecked():
            self._ai_write_topic_iterative(row)
        else:
            self._ai_write_topic_bulk(row)

    def _get_context_sources_dict(self, topic):
        """
        Resolve sources for a topic using the context dialog config.
        Priority: 1) user-configured context_config summaries,
                  2) top-N rated sources (falling back to
                     included/unscreened if no ratings).
        Returns a dict {source_number: summary}.
        """
        config = topic.get('context_config') or {}
        topic_id = topic.get('topic_id', '')
        n = self.topic_top_n.value()

        if config and config.get('summaries'):
            # User explicitly configured context — honour it exactly
            selected_nums = set(config['summaries'])
            source_dict = {}
            for s in self.session.sources:
                src = s if isinstance(s, dict) else s
                num = src.get('number')
                if num in selected_nums:
                    summary = src.get('summary')
                    if summary:
                        source_dict[num] = summary
            limit = config.get('summary_limit', 0)
            if limit > 0:
                source_dict = dict(list(source_dict.items())[:limit])
            return source_dict
        else:
            # No context config — fall back to top-N
            return _top_n_sources_for_topic(self.session, topic_id, n)

    def _get_context_sources_list(self, topic):
        """
        Like _get_context_sources_dict but returns a list of
        (num, summary) for iterative mode.
        Priority: 1) user-configured context_config summaries,
                  2) top-N rated sources (falling back to
                     included/unscreened if no ratings).
        """
        config = topic.get('context_config') or {}
        topic_id = topic.get('topic_id', '')
        n = self.topic_top_n.value()

        if config and config.get('summaries'):
            selected_nums = list(config['summaries'])
            source_list = []
            # Build lookup
            src_lookup = {}
            for s in self.session.sources:
                src = s if isinstance(s, dict) else s
                num = src.get('number')
                summary = src.get('summary') or src.get('full_text')
                if summary:
                    src_lookup[num] = summary
            for num in selected_nums:
                if num in src_lookup:
                    source_list.append((num, src_lookup[num]))
            limit = config.get('summary_limit', 0)
            if limit > 0:
                source_list = source_list[:limit]
            return source_list
        else:
            return _top_n_source_list(self.session, topic_id, n)

    def _ai_write_topic_bulk(self, row):
        """Bulk mode: all context sources in one prompt."""
        topic = self.session.topics[row]
        source_dict = self._get_context_sources_dict(topic)

        variables = {
            'topic': self.session.paper_topic,
            'topic_title': topic.get('title', ''),
            'context': str(source_dict),
        }
        prompt_template = self.config.get_prompt('generate_topic')
        dlg = PromptApprovalDialog(
            'generate_topic', prompt_template,
            variables, self.config, self)
        if dlg.exec():
            formatted = dlg.get_formatted_prompt()
            try:
                result = self.api.query(formatted, max_tokens=2000)
                out_dlg = OutputApprovalDialog(
                    topic.get('title', 'Topic'), result, self)
                if out_dlg.exec() and out_dlg.approved:
                    self.session.topics[row]['text'] = result
                    self.topic_text_edit.setPlainText(result)
                    self.topics_changed.emit()
            except Exception as e:
                QMessageBox.critical(self, "Error", str(e))

    def _ai_write_topic_iterative(self, row):
        """Iterative mode: one source per prompt, building up text."""
        topic = self.session.topics[row]
        sources = self._get_context_sources_list(topic)

        if not sources:
            QMessageBox.warning(self, "No Sources",
                                "No source summaries available.")
            return

        # Show prompt template for approval once at the start
        prompt_template = self.config.get_prompt(
            'generate_topic_iterative')
        sample_vars = {
            'topic': self.session.paper_topic or '',
            'topic_title': topic.get('title', ''),
            'source_number': '(N)',
            'source_text': '(source summary — one at a time)',
            'existing_text_clause': '(accumulated text so far)',
        }
        dlg = PromptApprovalDialog(
            'generate_topic_iterative', prompt_template,
            sample_vars, self.config, self)
        if not dlg.exec():
            return
        template = dlg.original_template

        # Iterate through each source
        accumulated = ""
        self.topic_progress.setVisible(True)
        self.topic_progress.setRange(0, len(sources))
        self.topic_progress.setValue(0)

        for i, (num, summary) in enumerate(sources):
            self.topic_progress.setValue(i)
            self.topic_progress.setFormat(
                f"Source [{num}] — {i+1}/{len(sources)}")

            existing_clause = ""
            if accumulated.strip():
                existing_clause = (
                    f"Here is the text written so far from previous "
                    f"sources:\n{accumulated}\n\n"
                    f"Add new, non-redundant information from this "
                    f"source below.\n")

            variables = {
                'topic': self.session.paper_topic or '',
                'topic_title': topic.get('title', ''),
                'source_number': num,
                'source_text': summary[:4000],
                'existing_text_clause': existing_clause,
            }
            formatted = template.format(**variables)
            try:
                result = self.api.query(
                    formatted, max_tokens=1500).strip()
                if result.upper() != 'SKIP' and len(result) > 10:
                    accumulated = (accumulated + "\n\n" + result).strip()
                    # Live preview
                    self.topic_text_edit.blockSignals(True)
                    self.topic_text_edit.setPlainText(accumulated)
                    self.topic_text_edit.blockSignals(False)
            except Exception as e:
                self.topic_progress.setFormat(
                    f"Error on [{num}]: {str(e)[:40]}")

        self.topic_progress.setValue(len(sources))
        self.topic_progress.setFormat("Done")
        self.topic_progress.setVisible(False)

        if not accumulated.strip():
            QMessageBox.information(
                self, "No Results",
                "No relevant information found in any source.")
            return

        # Final approval
        out_dlg = OutputApprovalDialog(
            topic.get('title', 'Topic'), accumulated, self)
        if out_dlg.exec() and out_dlg.approved:
            self.session.topics[row]['text'] = accumulated
            self.topic_text_edit.setPlainText(accumulated)
            self.topics_changed.emit()
        else:
            # Revert the live preview
            self.topic_text_edit.blockSignals(True)
            self.topic_text_edit.setPlainText(
                self.session.topics[row].get('text') or '')
            self.topic_text_edit.blockSignals(False)

    # ── Distill ──────────────────────────────────────────────────────

    def _distill_topic(self):
        row = self.topic_list.currentRow()
        if row < 0:
            return
        topic = self.session.topics[row]
        current_text = topic.get('text') or ''
        if not current_text.strip():
            QMessageBox.warning(self, "No Text",
                                "This topic has no text to distill.")
            return

        prompt_template = self.config.get_prompt('distill_topic')
        variables = {'text': current_text}
        dlg = PromptApprovalDialog(
            'distill_topic', prompt_template,
            variables, self.config, self)
        if dlg.exec():
            formatted = dlg.get_formatted_prompt()
            try:
                result = self.api.query(formatted, max_tokens=2000)
                out_dlg = OutputApprovalDialog(
                    f"Distill: {topic.get('title', 'Topic')}", result, self)
                if out_dlg.exec() and out_dlg.approved:
                    self.session.topics[row]['text'] = result
                    self.topic_text_edit.setPlainText(result)
                    self.topics_changed.emit()
            except Exception as e:
                QMessageBox.critical(self, "Error", str(e))

    # ── Context config ───────────────────────────────────────────────

    def _configure_context(self):
        row = self.topic_list.currentRow()
        if row < 0:
            return
        topic = self.session.topics[row]
        topic_id = topic.get('topic_id', '')
        current = topic.get('context_config', {}) or {}
        dlg = ContextConfigDialog(
            f"Topic: {topic.get('title', '?')}",
            self.session, current, self,
            topic_id=topic_id)
        if dlg.exec():
            self.session.topics[row]['context_config'] = dlg.get_config()
            self.topics_changed.emit()

    def _link_to_sections(self):
        row = self.topic_list.currentRow()
        if row < 0:
            return
        topic = self.session.topics[row]
        topic_id = topic.get('topic_id', '')
        title = topic.get('title', 'Untitled')
        current_linked = topic.get('linked_sections', [])

        dlg = LinkToSectionsDialog(
            f"{topic_id}: {title}", self.session,
            current_linked, self, topic_id=topic_id)
        if dlg.exec():
            linked = dlg.get_selected_keys()
            self.session.topics[row]['linked_sections'] = linked
            dlg.apply_to_session()
            self.topics_changed.emit()

    # ── Source-topic rating (batched to ≤10 topics per call) ────────

    def _rate_sources_for_topics(self):
        """
        Rate all sources for relevance to each topic.
        Batches topics into groups of 10 to prevent hallucination.
        """
        if not self.session.topics:
            QMessageBox.warning(self, "No Topics", "Add topics first.")
            return

        all_topic_titles = []
        all_topic_ids = []
        for t in self.session.topics:
            all_topic_titles.append(t.get('title', 'Untitled'))
            all_topic_ids.append(t.get('topic_id', ''))

        prompt_template = self.config.get_prompt(
            'rate_source_topics_batch')
        sample_batch = all_topic_titles[:10]
        sample_vars = {
            'topics': str(sample_batch),
            'journal': '(journal)',
            'summary': '(source summary)',
        }
        dlg = PromptApprovalDialog(
            'rate_source_topics_batch', prompt_template,
            sample_vars, self.config, self)
        if not dlg.exec():
            return
        template = dlg.original_template

        BATCH_SIZE = 10
        batches = []
        for start in range(0, len(all_topic_titles), BATCH_SIZE):
            batch_titles = all_topic_titles[start:start + BATCH_SIZE]
            batch_ids = all_topic_ids[start:start + BATCH_SIZE]
            batches.append((start, batch_titles, batch_ids))

        rated_count = 0
        for i, s in enumerate(self.session.sources):
            src = s if isinstance(s, dict) else s
            # Skip explicitly excluded sources
            if src.get('rating') is False:
                print(f"Source {i} is excluded, skipping...")
                continue

            summary = src.get('summary')
            if not summary:
                summary = src.get('full_text') # Use full text if summary missing
                print(f"Rating source {i} using its full text since no summary available...")
            else:
                print(f"Rating source {i} using its summary...")

            if 'topic_ratings' not in self.session.sources[i] \
               or self.session.sources[i]['topic_ratings'] is None:
                self.session.sources[i]['topic_ratings'] = {}

            for batch_start, batch_titles, batch_ids in batches:
                variables = {
                    'topics': str(batch_titles),
                    'journal': src.get('journal', 'Unknown'),
                    'summary': summary,
                }
                formatted = template.format(**variables)
                try:
                    import ast
                    result = self.api.query(
                        formatted, max_tokens=200).strip()
                    ratings = ast.literal_eval(result)
                    batch_ratings = [
                        min(10, max(1, int(r))) for r in ratings]
                    while len(batch_ratings) < len(batch_ids):
                        batch_ratings.append(5)
                    batch_ratings = batch_ratings[:len(batch_ids)]
                    for tid, rating in zip(batch_ids, batch_ratings):
                        self.session.sources[i][
                            'topic_ratings'][tid] = rating
                        rated_count += 1
                except Exception:
                    pass

        self.topics_changed.emit()
        try:
            main_window = self.window()
            if hasattr(main_window, 'sources_tab'):
                main_window.sources_tab.sources_changed.emit()
                main_window.sources_tab.refresh_from_session()
        except Exception:
            pass

        QMessageBox.information(
            self, "Done",
            f"Source-topic ratings complete. "
            f"Rated {rated_count} source-topic pairs "
            f"({len(batches)} batch(es) of ≤10 per source).")

    def refresh_from_session(self):
        self._load_from_session()


# ═══════════════════════════════════════════════════════════════════
#  Statistics Tab (Beta)
# ═══════════════════════════════════════════════════════════════════

class StatsTab(QWidget):
    """Statistics Tab (Beta) — for extracting numerical data from sources."""

    stats_changed = pyqtSignal()

    def __init__(self, config_manager, api_manager, session, parent=None):
        super().__init__(parent)
        self.config = config_manager
        self.api = api_manager
        self.session = session
        self._build_ui()
        self._load_from_session()

    def _build_ui(self):
        layout = QVBoxLayout(self)

        # Beta banner
        banner = QLabel(
            "⚠ BETA — This tab is a framework for future "
            "meta-analysis features.")
        banner.setStyleSheet(
            "background-color: #f39c12; color: white; padding: 8px; "
            "font-weight: bold; border-radius: 4px;")
        layout.addWidget(banner)

        # Toolbar
        toolbar = QHBoxLayout()
        add_btn = QPushButton("+ Add Statistical Question")
        add_btn.clicked.connect(self._add_stat)
        toolbar.addWidget(add_btn)
        remove_btn = QPushButton("- Remove")
        remove_btn.clicked.connect(self._remove_stat)
        toolbar.addWidget(remove_btn)

        gen_q_btn = QPushButton("✦ AI-Generate Questions Based on Paper Contents")
        gen_q_btn.setToolTip(
            "Ask the AI to generate statistical questions based on "
            "the paper topic, thesis, and results subsections")
        gen_q_btn.clicked.connect(self._ai_generate_stat_questions)
        toolbar.addWidget(gen_q_btn)

        rate_btn = QPushButton("✦ Rate Sources for Stats")
        rate_btn.setToolTip(
            "Rate all sources for relevance to each stat question")
        rate_btn.clicked.connect(self._rate_sources_for_stats)
        toolbar.addWidget(rate_btn)

        toolbar.addStretch()

        toolbar.addWidget(QLabel("Top N sources:"))
        self.stat_top_n = QSpinBox()
        self.stat_top_n.setRange(1, 999)
        self.stat_top_n.setValue(10)
        self.stat_top_n.setToolTip(
            "How many sources to include when querying stats")
        toolbar.addWidget(self.stat_top_n)
        layout.addLayout(toolbar)

        # Main area
        splitter = QSplitter(Qt.Orientation.Horizontal)

        self.stat_list = QListWidget()
        self.stat_list.currentRowChanged.connect(self._on_stat_selected)
        splitter.addWidget(self.stat_list)

        right = QWidget()
        right_layout = QVBoxLayout(right)

        self.stat_question_edit = QLineEdit()
        self.stat_question_edit.setPlaceholderText(
            "Statistical / mathematical question")
        self.stat_question_edit.textChanged.connect(
            self._on_question_changed)
        right_layout.addWidget(self.stat_question_edit)

        # ── Action row 1 ──
        btn_row = QHBoxLayout()
        text_btn = QPushButton("✦ Query (Text Response)")
        text_btn.clicked.connect(self._query_text)
        btn_row.addWidget(text_btn)

        python_btn = QPushButton("✦ Query (Python-parseable)")
        python_btn.clicked.connect(self._query_python)
        btn_row.addWidget(python_btn)

        distill_btn = QPushButton("✦ Distill")
        distill_btn.setToolTip(
            "Condense the text response while keeping all "
            "citations and numbers intact")
        distill_btn.clicked.connect(self._distill_stat)
        btn_row.addWidget(distill_btn)

        context_btn = QPushButton("📋 Context")
        context_btn.clicked.connect(self._configure_context)
        btn_row.addWidget(context_btn)

        link_btn = QPushButton("🔗 Link to Sections")
        link_btn.clicked.connect(self._link_to_sections)
        btn_row.addWidget(link_btn)
        btn_row.addStretch()
        right_layout.addLayout(btn_row)

        # ── Action row 2: mode toggle ──
        mode_row = QHBoxLayout()
        self.stat_iterative_cb = QCheckBox("Iterative mode")
        self.stat_iterative_cb.setToolTip(
            "When checked, the AI examines each source individually "
            "and builds up the response source-by-source.")
        mode_row.addWidget(self.stat_iterative_cb)
        mode_row.addStretch()
        right_layout.addLayout(mode_row)

        # ── Progress bar ──
        self.stat_progress = QProgressBar()
        self.stat_progress.setVisible(False)
        self.stat_progress.setTextVisible(True)
        right_layout.addWidget(self.stat_progress)

        right_layout.addWidget(QLabel("Text Response:"))
        self.text_response_edit = QTextEdit()
        self.text_response_edit.textChanged.connect(self._on_text_changed)
        right_layout.addWidget(self.text_response_edit)

        right_layout.addWidget(QLabel("Python-parseable Response:"))
        self.python_response_edit = QTextEdit()
        self.python_response_edit.setFont(QFont("Consolas", 10))
        self.python_response_edit.setMaximumHeight(120)
        self.python_response_edit.textChanged.connect(
            self._on_python_changed)
        right_layout.addWidget(self.python_response_edit)

        splitter.addWidget(right)
        splitter.setSizes([250, 550])
        layout.addWidget(splitter)

    # ── Session I/O ──────────────────────────────────────────────────

    def _load_from_session(self):
        self.stat_list.clear()
        for st in self.session.statistics:
            stat = st if isinstance(st, dict) else st
            self.stat_list.addItem(
                f"{stat.get('stat_id', '?')}: "
                f"{stat.get('question', 'Untitled')[:50]}")

    def _on_stat_selected(self, row):
        if row < 0 or row >= len(self.session.statistics):
            return
        st = self.session.statistics[row]
        self.stat_question_edit.blockSignals(True)
        self.stat_question_edit.setText(st.get('question', ''))
        self.stat_question_edit.blockSignals(False)
        self.text_response_edit.blockSignals(True)
        self.text_response_edit.setPlainText(
            st.get('text_response') or '')
        self.text_response_edit.blockSignals(False)
        self.python_response_edit.blockSignals(True)
        self.python_response_edit.setPlainText(
            st.get('python_response') or '')
        self.python_response_edit.blockSignals(False)

    def _on_question_changed(self, text):
        row = self.stat_list.currentRow()
        if row >= 0:
            self.session.statistics[row]['question'] = text
            self.stat_list.item(row).setText(
                f"{self.session.statistics[row].get('stat_id', '?')}: "
                f"{text[:50]}")
            self.stats_changed.emit()

    def _on_text_changed(self):
        row = self.stat_list.currentRow()
        if row >= 0:
            self.session.statistics[row]['text_response'] = \
                self.text_response_edit.toPlainText()
            self.stats_changed.emit()

    def _on_python_changed(self):
        row = self.stat_list.currentRow()
        if row >= 0:
            self.session.statistics[row]['python_response'] = \
                self.python_response_edit.toPlainText()
            self.stats_changed.emit()

    def _add_stat(self):
        question, ok = QInputDialog.getText(
            self, "New Statistic",
            "Statistical / mathematical question:")
        if ok and question:
            sid = f"S{len(self.session.statistics) + 1}"
            self.session.statistics.append({
                'stat_id': sid, 'question': question,
                'text_response': None, 'python_response': None,
                'linked_sections': [], 'context_config': None
            })
            self.stat_list.addItem(f"{sid}: {question[:50]}")
            self.stats_changed.emit()

    def _remove_stat(self):
        row = self.stat_list.currentRow()
        if row >= 0:
            stat_id = self.session.statistics[row].get('stat_id', '')
            if stat_id:
                # Clean up stat_ratings on every source
                for s in self.session.sources:
                    src = s if isinstance(s, dict) else s
                    sr = src.get('stat_ratings')
                    if sr and stat_id in sr:
                        del sr[stat_id]
                # Clean up section_contexts referencing this stat
                for cfg in self.session.section_contexts.values():
                    stat_list = cfg.get('statistics', [])
                    if stat_id in stat_list:
                        stat_list.remove(stat_id)
            self.stat_list.takeItem(row)
            del self.session.statistics[row]
            self.stats_changed.emit()

    # ── Context ──────────────────────────────────────────────────────

    def _get_context_sources_dict(self, stat):
        """
        Resolve sources for a stat using the context dialog config.
        Priority: 1) user-configured context_config summaries,
                  2) top-N rated sources (falling back to
                     included/unscreened if no ratings).
        Returns a dict {source_number: summary}.
        """
        config = stat.get('context_config') or {}
        stat_id = stat.get('stat_id', '')
        n = self.stat_top_n.value()

        if config and config.get('summaries'):
            selected_nums = set(config['summaries'])
            source_dict = {}
            for s in self.session.sources:
                src = s if isinstance(s, dict) else s
                num = src.get('number')
                if num in selected_nums:
                    summary = src.get('summary')
                    if summary:
                        source_dict[num] = summary
            limit = config.get('summary_limit', 0)
            if limit > 0:
                source_dict = dict(list(source_dict.items())[:limit])
            return source_dict
        else:
            return _top_n_sources_for_stat(self.session, stat_id, n)

    def _get_context_sources_list(self, stat):
        """
        Like _get_context_sources_dict but returns a list of
        (num, summary) for iterative mode.
        """
        config = stat.get('context_config') or {}
        stat_id = stat.get('stat_id', '')
        n = self.stat_top_n.value()

        if config and config.get('summaries'):
            selected_nums = list(config['summaries'])
            source_list = []
            src_lookup = {}
            for s in self.session.sources:
                src = s if isinstance(s, dict) else s
                num = src.get('number')
                summary = src.get('summary')
                if summary:
                    src_lookup[num] = summary
            for num in selected_nums:
                if num in src_lookup:
                    source_list.append((num, src_lookup[num]))
            limit = config.get('summary_limit', 0)
            if limit > 0:
                source_list = source_list[:limit]
            return source_list
        else:
            return _stat_source_list(self.session, stat_id, n)

    def _get_context_string(self, row):
        stat = self.session.statistics[row]
        source_dict = self._get_context_sources_dict(stat)
        return str(source_dict)

    # ── Query text (bulk vs iterative) ───────────────────────────────

    def _query_text(self):
        row = self.stat_list.currentRow()
        if row < 0:
            return
        if self.stat_iterative_cb.isChecked():
            self._query_text_iterative(row)
        else:
            self._query_text_bulk(row)

    def _query_text_bulk(self, row):
        stat = self.session.statistics[row]
        variables = {
            'question': stat.get('question', ''),
            'context': self._get_context_string(row),
        }
        prompt_template = self.config.get_prompt('stat_query_text')
        dlg = PromptApprovalDialog(
            'stat_query_text', prompt_template,
            variables, self.config, self)
        if dlg.exec():
            try:
                result = self.api.query(
                    dlg.get_formatted_prompt(), max_tokens=2000)
                out_dlg = OutputApprovalDialog(
                    stat.get('question', 'Stat'), result, self)
                if out_dlg.exec() and out_dlg.approved:
                    self.session.statistics[row]['text_response'] = result
                    self.text_response_edit.setPlainText(result)
                    self.stats_changed.emit()
            except Exception as e:
                QMessageBox.critical(self, "Error", str(e))

    def _query_text_iterative(self, row):
        """Iterative mode for stat text queries."""
        stat = self.session.statistics[row]
        sources = self._get_context_sources_list(stat)

        if not sources:
            QMessageBox.warning(self, "No Sources",
                                "No source summaries available.")
            return

        prompt_template = self.config.get_prompt(
            'generate_stat_iterative')
        sample_vars = {
            'question': stat.get('question', ''),
            'source_number': '(N)',
            'source_text': '(source summary — one at a time)',
            'existing_text_clause': '(accumulated text so far)',
        }
        dlg = PromptApprovalDialog(
            'generate_stat_iterative', prompt_template,
            sample_vars, self.config, self)
        if not dlg.exec():
            return
        template = dlg.original_template

        accumulated = ""
        self.stat_progress.setVisible(True)
        self.stat_progress.setRange(0, len(sources))
        self.stat_progress.setValue(0)

        for i, (num, summary) in enumerate(sources):
            self.stat_progress.setValue(i)
            self.stat_progress.setFormat(
                f"Source [{num}] — {i+1}/{len(sources)}")

            existing_clause = ""
            if accumulated.strip():
                existing_clause = (
                    f"Here is the data extracted so far from previous "
                    f"sources:\n{accumulated}\n\n"
                    f"Add new, non-redundant data from this source.\n")

            variables = {
                'question': stat.get('question', ''),
                'source_number': num,
                'source_text': summary[:4000],
                'existing_text_clause': existing_clause,
            }
            formatted = template.format(**variables)
            try:
                result = self.api.query(
                    formatted, max_tokens=1500).strip()
                if result.upper() != 'SKIP' and len(result) > 10:
                    accumulated = (accumulated + "\n\n" + result).strip()
                    self.text_response_edit.blockSignals(True)
                    self.text_response_edit.setPlainText(accumulated)
                    self.text_response_edit.blockSignals(False)
            except Exception as e:
                self.stat_progress.setFormat(
                    f"Error on [{num}]: {str(e)[:40]}")

        self.stat_progress.setValue(len(sources))
        self.stat_progress.setFormat("Done")
        self.stat_progress.setVisible(False)

        if not accumulated.strip():
            QMessageBox.information(
                self, "No Results",
                "No relevant data found in any source.")
            return

        out_dlg = OutputApprovalDialog(
            stat.get('question', 'Stat'), accumulated, self)
        if out_dlg.exec() and out_dlg.approved:
            self.session.statistics[row]['text_response'] = accumulated
            self.text_response_edit.setPlainText(accumulated)
            self.stats_changed.emit()
        else:
            self.text_response_edit.blockSignals(True)
            self.text_response_edit.setPlainText(
                self.session.statistics[row].get('text_response') or '')
            self.text_response_edit.blockSignals(False)

    # ── Query python (always bulk — iterative doesn't suit dicts) ────

    def _query_python(self):
        row = self.stat_list.currentRow()
        if row < 0:
            return
        stat = self.session.statistics[row]
        variables = {
            'question': stat.get('question', ''),
            'context': self._get_context_string(row),
        }
        prompt_template = self.config.get_prompt('stat_query_python')
        dlg = PromptApprovalDialog(
            'stat_query_python', prompt_template,
            variables, self.config, self)
        if dlg.exec():
            try:
                result = self.api.query(
                    dlg.get_formatted_prompt(), max_tokens=1000)
                self.session.statistics[row]['python_response'] = result
                self.python_response_edit.setPlainText(result)
                self.stats_changed.emit()
            except Exception as e:
                QMessageBox.critical(self, "Error", str(e))

    # ── Distill ──────────────────────────────────────────────────────

    def _distill_stat(self):
        row = self.stat_list.currentRow()
        if row < 0:
            return
        stat = self.session.statistics[row]
        current_text = stat.get('text_response') or ''
        if not current_text.strip():
            QMessageBox.warning(self, "No Text",
                                "This stat has no text response to distill.")
            return

        prompt_template = self.config.get_prompt('distill_stat')
        variables = {'text': current_text}
        dlg = PromptApprovalDialog(
            'distill_stat', prompt_template,
            variables, self.config, self)
        if dlg.exec():
            formatted = dlg.get_formatted_prompt()
            try:
                result = self.api.query(formatted, max_tokens=2000)
                out_dlg = OutputApprovalDialog(
                    f"Distill: {stat.get('question', 'Stat')}", result,
                    self)
                if out_dlg.exec() and out_dlg.approved:
                    self.session.statistics[row]['text_response'] = result
                    self.text_response_edit.setPlainText(result)
                    self.stats_changed.emit()
            except Exception as e:
                QMessageBox.critical(self, "Error", str(e))

    # ── Other ────────────────────────────────────────────────────────

    def _configure_context(self):
        row = self.stat_list.currentRow()
        if row < 0:
            return
        stat = self.session.statistics[row]
        stat_id = stat.get('stat_id', '')
        current = stat.get('context_config', {}) or {}
        dlg = ContextConfigDialog(
            f"Stat: {stat.get('question', '?')}",
            self.session, current, self,
            stat_id=stat_id)
        if dlg.exec():
            self.session.statistics[row]['context_config'] = \
                dlg.get_config()
            self.stats_changed.emit()

    def _link_to_sections(self):
        row = self.stat_list.currentRow()
        if row < 0:
            return
        stat = self.session.statistics[row]
        stat_id = stat.get('stat_id', '')
        question = stat.get('question', 'Untitled')
        current_linked = stat.get('linked_sections', [])

        dlg = LinkToSectionsDialog(
            f"{stat_id}: {question[:50]}", self.session,
            current_linked, self, stat_id=stat_id)
        if dlg.exec():
            linked = dlg.get_selected_keys()
            self.session.statistics[row]['linked_sections'] = linked
            dlg.apply_to_session()
            self.stats_changed.emit()

    # ── AI-generate statistical questions ────────────────────────────

    def _ai_generate_stat_questions(self):
        """Use AI to generate statistical questions from paper context."""
        if not self.session.paper_topic:
            QMessageBox.warning(self, "No Topic",
                                "Set a paper topic first.")
            return

        context_parts = [f"Paper topic: {self.session.paper_topic}"]
        if self.session.thesis:
            context_parts.append(f"Thesis: {self.session.thesis}")
        if self.session.results:
            headers = [rs.get('section', 'Untitled')
                       for rs in self.session.results]
            context_parts.append(
                f"Results subsection headers: {'; '.join(headers)}")

        context = "\n".join(context_parts)

        prompt_template = self.config.get_prompt(
            'auto_generate_stat_questions')
        variables = {
            'topic': self.session.paper_topic,
            'context': context,
        }
        dlg = PromptApprovalDialog(
            'auto_generate_stat_questions', prompt_template,
            variables, self.config, self)
        if dlg.exec():
            formatted = dlg.get_formatted_prompt()
            try:
                result = self.api.query(formatted, max_tokens=1500)
                questions = [q.strip()
                             for q in result.split(';') if q.strip()]
                reply = QMessageBox.question(
                    self, "Generated Stat Questions",
                    f"Generated {len(questions)} questions:\n\n" +
                    "\n".join(f"  {i+1}. {q}"
                              for i, q in enumerate(questions)) +
                    "\n\nAdd these as new statistical questions?",
                    QMessageBox.StandardButton.Yes |
                    QMessageBox.StandardButton.No)
                if reply == QMessageBox.StandardButton.Yes:
                    for q in questions:
                        sid = f"S{len(self.session.statistics) + 1}"
                        self.session.statistics.append({
                            'stat_id': sid, 'question': q,
                            'text_response': None,
                            'python_response': None,
                            'linked_sections': [],
                            'context_config': None,
                        })
                    self._load_from_session()
                    self.stats_changed.emit()
            except Exception as e:
                QMessageBox.critical(self, "Error", str(e))

    # ── Source-stat rating (batched to ≤10 per call) ─────────────────

    def _rate_sources_for_stats(self):
        """Rate all sources for relevance to each stat question."""
        if not self.session.statistics:
            QMessageBox.warning(self, "No Stats",
                                "Add statistical questions first.")
            return

        all_stat_questions = []
        all_stat_ids = []
        for st in self.session.statistics:
            all_stat_questions.append(st.get('question', 'Untitled'))
            all_stat_ids.append(st.get('stat_id', ''))

        prompt_template = self.config.get_prompt(
            'rate_source_stats_batch')
        sample_batch = all_stat_questions[:10]
        sample_vars = {
            'topics': str(sample_batch),
            'journal': '(journal)',
            'summary': '(source summary)',
        }
        dlg = PromptApprovalDialog(
            'rate_source_stats_batch', prompt_template,
            sample_vars, self.config, self)
        if not dlg.exec():
            return
        template = dlg.original_template

        BATCH_SIZE = 10
        batches = []
        for start in range(0, len(all_stat_questions), BATCH_SIZE):
            batch_qs = all_stat_questions[start:start + BATCH_SIZE]
            batch_ids = all_stat_ids[start:start + BATCH_SIZE]
            batches.append((start, batch_qs, batch_ids))

        rated_count = 0
        for i, s in enumerate(self.session.sources):
            src = s if isinstance(s, dict) else s
            # Skip explicitly excluded sources
            if src.get('rating') is False:
                continue
            summary = src.get('summary')
            if not summary:
                continue

            if 'stat_ratings' not in self.session.sources[i] \
               or self.session.sources[i]['stat_ratings'] is None:
                self.session.sources[i]['stat_ratings'] = {}

            for batch_start, batch_qs, batch_ids in batches:
                variables = {
                    'topics': str(batch_qs),
                    'journal': src.get('journal', 'Unknown'),
                    'summary': summary,
                }
                formatted = template.format(**variables)
                try:
                    import ast
                    result = self.api.query(
                        formatted, max_tokens=200).strip()
                    print(result)
                    ratings = ast.literal_eval(result)
                    batch_ratings = [
                        min(10, max(1, int(r))) for r in ratings]
                    while len(batch_ratings) < len(batch_ids):
                        batch_ratings.append(5)
                    batch_ratings = batch_ratings[:len(batch_ids)]
                    for sid, rating in zip(batch_ids, batch_ratings):
                        self.session.sources[i][
                            'stat_ratings'][sid] = rating
                        rated_count += 1
                except Exception:
                    pass

        self.stats_changed.emit()
        try:
            main_window = self.window()
            if hasattr(main_window, 'sources_tab'):
                main_window.sources_tab.sources_changed.emit()
                main_window.sources_tab.refresh_from_session()
        except Exception:
            pass

        QMessageBox.information(
            self, "Done",
            f"Source-stat ratings complete. "
            f"Rated {rated_count} source-stat pairs "
            f"({len(batches)} batch(es) of ≤10 per source).")

    def refresh_from_session(self):
        self._load_from_session()
```
