## Supplementary material for "In Search of Ethical Procedures for LLM-Assisted Systematic Review Production: A Proof-of-Concept Evaluation of Selected Review Components": source_data: semi_auto_lymphoma.pdf

### The Need for Standardization in Next-Generation Sequencing Studies for Classic Hodgkin Lymphoma: A Systematic Review

#### 1 Abstract

#### 2 Introduction

Classical Hodgkin lymphoma (cHL) represents a unique B-cell malignancy characterized by rare Hodgkin and Reed-Sternberg (HRS) cells dispersed within an extensive inflammatory microenvironment [1]. While cHL has achieved high cure rates with conventional therapies, approximately 15-20% of patients experience treatment failure or relapse, highlighting the need for improved risk stratification and personalized treatment approaches [2,3]. The advent of next-generation sequencing (NGS) technologies has provided unprecedented opportunities to characterize the genomic landscape of cHL and identify potential therapeutic targets.

The application of NGS to cHL research has revealed recurrent mutations in critical pathways including JAK/STAT signaling, NF-$\kappa$B regulation, and immune evasion mechanisms [4,5]. Studies have identified frequently mutated genes such as SOCS1 (28-60% of cases), STAT6 (21-32%), B2M (19-33%), and TNFAIP3 (14-44%), providing insights into cHL pathogenesis [4,5,6,7,8,9]. These findings have been complemented by liquid biopsy approaches using circulating tumor DNA (ctDNA), which have demonstrated the feasibility of non-invasive genetic profiling and treatment monitoring [6,7,8,10].

However, significant methodological heterogeneity exists across cHL NGS studies. The scarcity of HRS cells, comprising <1-2% of the tumor cellularity, has necessitated diverse enrichment strategies including laser capture microdissection, flow cytometry sorting, and CD30+ cell selection [5,11,12]. Technical approaches have varied from targeted gene panels covering 9-522 genes to whole-exome sequencing, with different platforms, coverage depths, and variant calling algorithms employed [4,5,6,7,10,13]. Sample types have ranged from fresh-frozen tissue to formalin-fixed paraffin-embedded specimens and liquid biopsies, each presenting unique analytical challenges [6,7,13].

This methodological diversity has resulted in inconsistent mutation frequencies and pathway involvement across studies. For instance, SOCS1 mutation rates have varied from 28% to 83% of ctDNA-positive cases, while TP53 alterations have ranged from 9% to 25% depending on the cohort and methodology [4,5,6,7,10,13]. Such variability complicates cross-study comparisons and hinders the development of standardized diagnostic and prognostic biomarkers.

Furthermore, recent consensus initiatives in mature lymphoid malignancies have emphasized the critical importance of technical standardization for reliable NGS implementation

[2]. The establishment of minimal gene panels and quality control measures has proven essential for ensuring reproducible results and facilitating multicenter collaborations.

Given these challenges, there is an urgent need for standardized NGS approaches in cHL research to enable meaningful comparison of results, accelerate biomarker development, and ultimately improve patient outcomes through precision medicine approaches.

### 5 Conclusion

This systematic review demonstrates that methodological heterogeneity in NGS studies of classic Hodgkin lymphoma has reached a critical threshold that fundamentally compromises the reliability and clinical utility of genomic findings. The pervasive variability in tumor enrichment strategies, technical approaches, analytical pipelines, and sample handling creates systematic biases that preclude meaningful cross-study comparisons and impede biomarker development. The establishment of standardized methodological approaches, guided by successful consensus initiatives in related malignancies [2], represents an urgent priority for advancing cHL genomic research toward clinically actionable precision medicine applications. Only through such harmonization efforts can the field realize the promise of NGS technologies for improving outcomes in the 15-20% of cHL patients who experience treatment failure with conventional therapies [2,3].
