## Supplementary material for "In Search of Ethical Procedures for LLM-Assisted Systematic Review Production: A Proof-of-Concept Evaluation of Selected Review Components": source_data: semi_auto_triple_negative.pdf

### Triple-Negative Breast Cancer and Predictive Markers of Response to Neoadjuvant Chemotherapy: A Systematic Review

#### 1 Abstract

Background: Triple-negative breast cancer (TNBC), characterized by the absence of estrogen receptor, progesterone receptor, and HER2 expression, represents 15-20% of breast cancers worldwide. While neoadjuvant chemotherapy (NAC) remains the standard of care, only 30-50% of patients achieve pathological complete response (pCR), highlighting the critical need for reliable predictive biomarkers.Results: Multiple biomarker classes demonstrate predictive value for NAC response. High levels of tumor-infiltrating lymphocytes (TILs) consistently correlate with improved pCR rates, particularly in lymphocyte-predominant breast cancer. CD8+ and CD4+ T cell subsets show specific predictive significance. DNA repair deficiency emerges as another crucial determinant, with BRCA1 mutation carriers achieving significantly higher pCR rates (53.8% vs 29.7%) compared to non-carriers. The TNBCtype-4 molecular classifier reveals differential response rates across subtypes: BL1 (60.6%), BL2 (52.1%), M (33.3%), and LAR (32.5%). Multiple microRNA signatures show promise, including a 4-miRNA signature (miR-4448, miR-2392, miR-2467-3p, miR-4800-3p) with an AUC of 0.7652. Blood-based markers, particularly the neutrophil-to-lymphocyte ratio, demonstrate predictive utility. Recent computational approaches, including deep learning-based histological biomarkers, achieve superior predictive performance (AUC 0.847) compared to conventional markers.Conclusions: Optimal prediction of NAC response in TNBC requires an integrated approach combining multiple biomarker classes, particularly immune parameters, DNA repair proficiency, and molecular subtyping. While individual markers show promise, their predictive power is maximized when combined in comprehensive models. Standardization of biomarker assessment methods and establishment of clinically validated cutoff values remain critical challenges for implementation in real-world settings.

#### 2 Introduction

Triple-negative breast cancer (TNBC), defined by the absence of estrogen receptor, progesterone receptor, and HER2 expression, represents 15-20% of all breast cancers diagnosed worldwide and accounts for nearly 200,000 cases annually [1]. This aggressive subtype is characterized by higher rates of recurrence and distant metastasis compared to other breast cancer subtypes [2]. In the absence of targeted therapies, neoadjuvant chemotherapy (NAC) remains the standard of care for early-stage and locally advanced TNBC [3].The achievement of pathological complete response (pCR) after NAC is strongly associated with improved survival outcomes in TNBC [2,4]. However, only approximately 30-50% of TNBC patients achieve pCR with standard NAC regimens [1,5]. This heterogeneity in treatment response highlights the critical need for reliable biomarkers to predict NAC efficacy and guide treatment decisions [6].Recent advances in molecular profiling have revealed that TNBC can be further classified into distinct subtypes with different clinical behaviors and treatment sensitivities [7]. Additionally, the tumor immune microenvironment has emerged as a crucial determinant of chemotherapy response, with tumor-infiltrating lymphocytes (TILs) showing prognostic and predictive value in TNBC [8,9]. High TIL levels are associated with improved pCR rates and better survival outcomes, particularly in TNBC compared to other breast cancer subtypes [10].Multiple potential predictive biomarkers have been investigated, including genetic alterations [11], immune signatures [12], and imaging-based markers [13]. The integration of these various biomarkers may provide more accurate prediction of NAC response than single markers alone [14]. Furthermore, understanding the biological mechanisms underlying chemotherapy resistance could lead to the development of novel therapeutic strategies to improve outcomes in TNBC [6].Given the expanding landscape of potential predictive biomarkers and their critical importance in treatment decision-making, there is a pressing need to systematically evaluate and compare these markers to identify the most reliable predictors of NAC response in TNBC [15]. This systematic review aims to comprehensively assess the current evidence regarding predictive markers of NAC response in TNBC, with the goal of identifying the most promising biomarkers for clinical implementation.

### 3 Results

#### 3.1 Clinical and Pathological Predictive Markers of Neoadjuvant Response

Several clinical and pathological features have demonstrated predictive value for neoadjuvant chemotherapy response in triple-negative breast cancer (TNBC). In terms of clinical characteristics, younger age and earlier clinical stage have been associated with higher pathological complete response (pCR) rates [16]. Studies have found that node-negative status is predictive of better response to neoadjuvant chemotherapy [16,17]. High tumor grade has also been identified as a predictor of increased pCR likelihood [2].Ki-67 proliferation index has emerged as an important predictive marker, with multiple studies demonstrating that high Ki-67 correlates with improved response to neoadjuvant chemotherapy [18,19,20]. One analysis found that tumors with Ki-67 $\geq$50% had significantly higher pCR rates compared to those with lower Ki-67 indices [20]. However, some studies have reported conflicting results regarding Ki-67's predictive value [21].The presence and extent of tumor-infiltrating lymphocytes (TILs) has been consistently associated with improved response to neoadjuvant chemotherapy across multiple studies [8,9,10]. Higher levels of stromal TILs correlate with increased pCR rates [22,23]. One study found that lymphocyte-predominant breast cancer (LPBC), defined as >40% lymphocytic infiltration, was associated with significantly higher pCR rates compared to non-LPBC tumors [22].P53 status has also been investigated as a predictive marker. Some studies have found that p53 overexpression correlates with higher pCR rates [20,24], though this association has not been uniformly observed across all analyses. The predictive value of androgen receptor (AR) expression has been examined, with AR-negative TNBC demonstrating higher pCR rates in some studies [25,26].Importantly, the integration of multiple clinical and pathological markers may provide more accurate prediction of neoadjuvant chemotherapy response compared to individual factors alone [14]. Recent research has focused on developing comprehensive prediction models that incorporate various clinicopathological features to better identify patients likely to achieve pCR [14,15,16].Further investigation is still needed to validate these markers and establish standardized cutoff values for clinical use. Additionally, the predictive value of some markers may vary based on the specific chemotherapy regimen used [5,6]. Understanding these clinical and pathological predictors of response remains crucial for optimal patient selection and treatment planning in the neoadjuvant setting.

##### 3.2 Tumor-Infiltrating Lymphocytes (TILs) and Immune-Related Biomarkers

Tumor-infiltrating lymphocytes (TILs) and immune-related markers have emerged as important predictors of response to neoadjuvant chemotherapy in TNBC. Multiple studies have demonstrated that higher levels of stromal TILs are significantly associated with increased pathological complete response (pCR) rates [8,9,10]. One analysis found that lymphocyte-predominant breast cancer (LPBC), defined as >40% lymphocytic infiltration, was associated with significantly higher pCR rates compared to non-LPBC tumors (87% vs 9.4%, p=0.001) [22]. The presence of TILs has also shown prognostic value, with higher TIL levels correlating with improved disease-free and overall survival [8,10,27].Specific immune cell subsets appear to influence treatment response. Higher levels of CD8+ cytotoxic T cells and CD4+ T cells in pre-treatment samples have been associated with improved pCR rates [23,28]. One study found that both intratumoral and stromal CD4+ T cell density independently predicted pCR, particularly in patients receiving platinum-containing chemotherapy regimens [29]. Early changes in immune infiltrate during treatment may also have predictive value, with transitions from "cold" to "hot" immune status suggesting higher pCR rates [30].Several studies have examined immune checkpoint molecules and other immune-related proteins. High PD-L1 expression has been associated with increased pCR rates in some analyses [31,32]. CD73 expression appears to play an opposing role, with high CD73 levels correlating with lower pCR rates and worse survival outcomes [33,34]. The combination of immune markers may provide additional predictive value, as demonstrated by one study showing that both PD-L1 status and TIL levels together better predicted response than either marker alone [32].Gene expression analyses have identified immune-related signatures associated with chemotherapy response. Higher expression of immune-related genes including IDO1, CXCL9, CXCL10, and STAT1 has been associated with improved survival after chemotherapy in TNBC patients [12]. The presence of these immune signatures correlated with increased CD8+ T cell infiltration in responder patients [12]. Integration of multiple immune parameters through approaches like multiplexed immunofluorescence may provide more comprehensive prediction of treatment response [30].These findings highlight the central role of the immune microenvironment in determining response to neoadjuvant chemotherapy in TNBC. The evidence suggests that pre-existing immune activation, particularly involving cytotoxic T cells, may enhance sensitivity to chemotherapy through both direct and indirect mechanisms [35,36]. This has led to growing interest in combining standard chemotherapy with immunotherapeutic approaches to potentially improve response rates in TNBC [36].

#### 3.3 Genetic and Molecular Signatures Associated with Treatment Response

Several studies have investigated genetic and molecular signatures predictive of neoadjuvant chemotherapy response in TNBC. The TNBCtype-4 molecular classifier has demonstrated significant predictive value, with differential pathological complete response (pCR) rates across subtypes: BL1 (60.6%), BL2 (52.1%), M (33.3%), LAR (32.5%) [37]. This classification remained independently associated with pCR in multivariate analysis including clinicopathological factors [37].Research has identified multiple gene expression patterns associated with chemotherapy response. A comprehensive analysis of over 700 TNBC patients found 49 genes consistently affected by neoadjuvant chemotherapy, involved in wound response, chemokine release, cell division, and programmed cell death pathways [5]. High expression of immune-related genes including IDO1, CXCL9, CXCL10, and STAT1 was associated with improved survival after chemotherapy, with STAT1 showing the strongest effect [12]. This immune signature correlated with increased CD8+ T cell infiltration in responder patients [12].Several studies have examined genetic alterations and DNA repair deficiency. Homologous recombination (HR) deficiency status significantly predicted response to standard neoadjuvant chemotherapy [38]. Analysis of chemo-response-related genes found that approximately 50% of TNBCs harbored at least one somatic mutation, with TP53 (21.4%), BRCA1 (11.6%), and RET (5.4%) being most common [11]. While individual mutation frequencies did not differ between pCR and non-pCR groups, mutations in 10 DNA repair genes involved in homologous recombination discriminated between responders and non-responders [11].BRCA1/2 status has also shown predictive value. In Chinese TNBC patients, BRCA1 mutation carriers had significantly higher pCR rates compared to non-carriers (53.8% vs 29.7%, $p<0.001$) with anthracycline-based regimens [39]. PIK3CA mutations were associated with reduced pCR rates [6,40]. Additionally, pathway analysis revealed PIK3CA mutations may confer resistance through inhibition of apoptosis and activation of PI3K/AKT/mTOR signaling [6].MicroRNA signatures have also demonstrated predictive potential. A 4-miRNA signature (miR-4448, miR-2392, miR-2467-3p, miR-4800-3p) could discriminate pCR versus non-pCR with an AUC of 0.7652 [41]. Other studies identified miR-200b-3p, miR-190a, and miR-512-5p as potentially associated with chemotherapy response [42], while miR-145-5p showed promise as a predictor of cisplatin response [43].Recent advances in artificial intelligence have enabled development of novel histological biomarkers. A deep learning-based pCR-score derived from H&E-stained tissue images achieved an AUC of 0.847 in predicting pCR, outperforming conventional biomarkers [13]. Integration of this score with clinical and pathological factors further improved predictive performance [13].

#### 3.4 BRCA1/2 Status and DNA Repair Deficiency as Response Predictors

BRCA1/2 Status and DNA Repair Deficiency (HRD) have emerged as important predictive markers of neoadjuvant chemotherapy response in TNBC. Multiple studies have demonstrated that BRCA1 mutation carriers achieve significantly higher pathological complete response (pCR) rates compared to non-carriers. In a large Chinese cohort study, BRCA1 mutation carriers had a pCR rate of 53.8% versus 29.7% in non-carriers ($p<0.001$) when treated with anthracycline-based neoadjuvant chemotherapy regimens [39]. Similarly, another study found BRCA1-associated TNBCs had higher pCR rates of 68% compared to 37% in non-carriers (p=0.01), although this did not translate to superior relapse-free survival among carriers [44].Beyond germline mutations, somatic BRCA1 alterations have also been investigated. One study identified BRCA1 somatic mutations in 3.9% of TNBC patients without germline mutations, with these carriers showing a trend toward higher pCR rates of 60% versus 30.4% in non-carriers, though this did not reach statistical significance [45]. The presence of homologous recombination deficiency (HRD), which can result from various genetic alterations beyond BRCA1/2, has also demonstrated predictive value. HR deficient tumors showed significantly higher pCR rates to standard neoadjuvant chemotherapy compared to HR proficient tumors [38]. A comprehensive genetic analysis revealed that approximately 50% of TNBCs harbored at least one somatic mutation in DNA repair genes, with TP53 (21.4%), BRCA1 (11.6%), and RET (5.4%) being the most common alterations [11].Molecular analyses have shown that mutations in 10 DNA repair genes involved in homologous recombination could discriminate between responders and non-responders to paclitaxel/carboplatin neoadjuvant chemotherapy [11]. This suggests that a broader assessment of DNA repair pathway alterations, rather than just BRCA1/2 status alone, may better predict chemotherapy response. Indeed, investigation of twelve genes involved in DNA repair demonstrated that HR deficiency status significantly predicted response to standard neoadjuvant chemotherapy [38].The relationship between HRD and immune activation has also been explored. One study found that HR deficient TNBCs showed higher levels of tumor-infiltrating lymphocytes and elevated immune gene expression signatures [46]. Moreover, the combination of HRD assessment with immune activation markers enhanced the identification of patients likely to respond to anthracycline-taxane chemotherapy [46].These findings suggest that both germline and somatic alterations affecting DNA repair pathways may serve as predictive biomarkers for neoadjuvant chemotherapy response in TNBC. The integration of HRD assessment with other molecular and immune markers may provide more comprehensive prediction of treatment outcomes.

#### 3.5 MicroRNA Expression Patterns Predicting Chemotherapy Response

Several studies have investigated the role of microRNAs (miRNAs) as potential biomarkers for predicting response to neoadjuvant chemotherapy in TNBC. In a study analyzing miRNA profiles from pre-treatment exosomes, researchers identified 16 differentially expressed exosomal miRNAs between pathological complete response (pCR) and non-pCR patients. A 4-miRNA signature comprising miR-4448, miR-2392, miR-2467-3p, and miR-4800-3p demonstrated predictive value with an AUC of 0.7652 [41]. This signature could potentially help identify chemotherapy-resistant patients at higher risk of recurrence.Further investigation of specific miRNAs revealed that miR-145-5p expression levels were significantly lower in patients achieving pCR compared to non-responders. Low miR-145-5p expression was associated with increased disease-free survival, and ROC analysis suggested its potential as a predictor of pCR (AUC=0.7899, p<0.003). Functional studies demonstrated that restoring miR-145-5p expression in MDA-MB-231 cells led to increased chemosensitivity to cisplatin therapy [43].Additional research identified three miRNAs (miR-200b-3p, miR-190a, and miR-512-5p) that showed differential expression between good and poor responders to chemotherapy, though these differences approached but did not reach statistical significance (p=0.06). Higher expression of miR-200b-3p and miR-190a, combined with lower miR-512-5p expression in diagnostic core biopsies, was associated with improved pathologic response to chemotherapy [42].A comprehensive analysis of over 700 TNBC patients identified 49 genes consistently affected by neoadjuvant chemotherapy, suggesting that early chemotherapy-induced gene expression changes may predict treatment response [5]. The integration of multiple microRNA markers, along with clinical and pathological factors, may provide more accurate prediction of neoadjuvant chemotherapy response compared to individual factors alone [14,15].

#### 3.6 Blood-Based Biomarkers and Systemic Inflammatory Markers

Multiple studies have investigated blood-based biomarkers and systemic inflammatory markers as potential predictors of response to neoadjuvant chemotherapy in TNBC. The neutrophil-to-lymphocyte ratio (NLR) has emerged as one of the most extensively studied blood-based markers. In a study of 87 TNBC patients, low baseline NLR ($\leq$1.7) was independently associated with higher pCR rates (42.1% vs 18.4%, p=0.018) and improved recurrence-free survival (5-year RFS 83.7% vs 66.9%, p=0.016) [47]. These findings were supported by another study of 177 breast cancer patients, where low NLR (<3.0) correlated with higher pCR rates, particularly in TNBC patients [48].The derived neutrophil-to-lymphocyte ratio (dNLR) has also been evaluated. A pooled analysis of two randomized studies found that high baseline and end-of-treatment dNLR were associated with lower benefit from neoadjuvant chemotherapy in TNBC [49]. The platelet-to-lymphocyte ratio (PLR) has shown promise as well, with one study finding that PLR <133.25 predicted achieving either pCR or minimal residual disease after neoadjuvant chemotherapy [50].Interestingly, studies examining the relationship between systemic inflammatory markers and local immune response have yielded mixed results. One analysis of 395 TNBC patients found no correlation between baseline NLR and stromal tumor-infiltrating lymphocytes (sTILs), though both were independent prognostic indicators for disease-free survival [51]. Another study evaluating both relative eosinophil count and lymphocyte levels found that combining these parameters into an eosinophil x lymphocyte product was significantly associated with pCR (p=0.002), relapse (p=0.028), and disease-free survival (p=0.012) [52].Several studies have also investigated the hemoglobin-albumin-lymphocyte-platelet (HALP) score. In an analysis of 92 TNBC patients, low HALP was identified as an independent risk factor for poor neoadjuvant chemotherapy efficacy, along with high NLR and high PLR [53]. This suggests that composite scores incorporating multiple blood-based parameters may provide additional predictive value.These studies highlight the potential utility of readily available blood-based markers for predicting neoadjuvant chemotherapy response in TNBC. However, standardization of cut-off values and prospective validation in larger cohorts are needed before clinical implementation.

#### 3.7 Integrative Multi-Marker Approaches for Response Prediction

Recent studies have highlighted the value of combining multiple biomarkers to improve prediction of neoadjuvant chemotherapy response in TNBC. Integration of clinical, pathological, and molecular markers has shown superior predictive performance compared to single markers alone [13]. One study demonstrated that combining PD-L1 status with tumor-infiltrating lymphocytes (TILs) enhanced the identification of patients likely to respond to chemotherapy compared to either marker alone [32].The development of comprehensive immune profiles has emerged as a promising approach. One study found that integrating multiple immune parameters through multiplexed immunofluorescence provided more detailed prognostic information than individual markers [30]. Similarly, analysis of immune cell subsets revealed that combining assessments of CD4+ T cells, CD8+ T cells, and PD-1/PD-L1 expression offered improved prediction of pathological complete response [29].Novel computational approaches have also shown promise. A deep learning-based histological biomarker achieved an AUC of 0.847 in predicting pCR, outperforming conventional markers. When combined with clinical and pathological factors, the integrated model further improved predictive performance (AUC 0.890) [13]. Gene expression-based approaches have also proven valuable, with one study identifying a signature combining IDO1, LAG3, STAT1, and GZMB that effectively predicted favorable relapse-free survival [12].Some studies have specifically examined the integration of blood-based and tissue-based markers. The combination of TILs assessment with systemic inflammatory markers like neutrophil-to-lymphocyte ratio (NLR) has been investigated, though correlation between these markers was not consistently observed [51]. Another study found that combining eosinophil and lymphocyte counts into an integrated score showed significant associations with pCR (p=0.002) and disease-free survival (p=0.012) [52].Emerging research has also explored combining traditional clinicopathological markers with novel molecular features. One study demonstrated that integrating HRD status assessment with immune activation markers enhanced the identification of patients likely to respond to anthracycline-taxane chemotherapy [46]. The evaluation of multiple biomarkers may provide more comprehensive prediction of treatment outcomes compared to individual factors alone [14,15].These integrative approaches highlight the complex interplay between different biological features in determining chemotherapy response, suggesting that multi-marker strategies may be necessary for optimal patient stratification in TNBC.

#### 3.8 Analysis of Treatment Response Definitions and their Prognostic Value

The assessment and definition of pathological complete response (pCR) has emerged as a critical consideration in evaluating neoadjuvant chemotherapy efficacy in triple-negative breast cancer (TNBC). A large pooled analysis of 6,377 patients across seven randomized trials demonstrated that disease-free survival was significantly better in patients achieving pCR defined as no invasive or in situ residuals in both breast and nodes (ypT0 ypN0) compared to other less stringent definitions [4]. This most stringent definition of pCR showed the strongest prognostic value compared to definitions that included residual in situ disease or isolated tumor cells [4].The prognostic impact of pCR appears to vary by breast cancer subtype. Multiple studies have shown that achieving pCR is particularly prognostic in TNBC compared to other subtypes [1,2]. In TNBC specifically, pCR rates with standard neoadjuvant chemotherapy regimens range from 30-50% [1,5]. Those achieving pCR consistently demonstrate significantly improved survival outcomes compared to those with residual disease [2,4].Recent research has focused on more sophisticated methods of evaluating residual disease beyond binary pCR definitions. The residual cancer burden (RCB) index has been validated as a robust method for quantifying residual disease [54]. Analysis of RCB distributions between treatment arms in clinical trials may provide additional insights beyond simple pCR rates [54]. Furthermore, studies suggest that even minimal residual disease (RCB-I) may carry different prognostic implications compared to moderate (RCB-II) or extensive (RCB-III) residual disease [54].Importantly, the prognostic value of achieving pCR may be impacted by the presence of certain biomarkers. For instance, while BRCA1 mutation carriers showed higher pCR rates with anthracycline-based chemotherapy, this did not necessarily translate to superior relapse-free survival [44]. Similarly, studies examining immune markers found that the presence of tumor-infiltrating lymphocytes retained prognostic value even after accounting for pCR status [8,9].The standardization of response assessment and reporting has been identified as crucial for comparing results across studies and determining optimal predictive biomarkers [14,15]. This includes both the technical aspects of evaluating residual disease as well as the timing of response assessment, as some studies suggest that early response dynamics may provide additional prognostic information [30].

### 4 Discussion

Triple-negative breast cancer (TNBC), defined by the absence of estrogen receptor, progesterone receptor, and HER2 expression, represents 15-20% of all breast cancers diagnosed worldwide and accounts for nearly 200,000 cases annually [1]. This aggressive subtype is characterized by higher rates of recurrence and distant metastasis compared to other breast cancer subtypes [2]. In the absence of targeted therapies, neoadjuvant chemotherapy (NAC) remains the standard of care for early-stage and locally advanced TNBC [3].The achievement of pathological complete response (pCR) after NAC is strongly associated with improved survival outcomes in TNBC [2,4]. However, only approximately 30-50% of TNBC patients achieve pCR with standard NAC regimens [1,5]. This heterogeneity in treatment response highlights the critical need for reliable biomarkers to predict NAC efficacy and guide treatment decisions [6].Recent advances in molecular profiling have revealed that TNBC can be further classified into distinct subtypes with different clinical behaviors and treatment sensitivities [7]. Additionally, the tumor immune microenvironment has emerged as a crucial determinant of chemotherapy response, with tumor-infiltrating lymphocytes (TILs) showing prognostic and predictive value in TNBC [8,9]. High TIL levels are associated with improved pCR rates and better survival outcomes, particularly in TNBC compared to other breast cancer subtypes [10].Multiple potential predictive biomarkers have been investigated, including genetic alterations [11], immune signatures [12], and imaging-based markers [13]. The integration of these various biomarkers may provide more accurate prediction of NAC response than single markers alone [14]. Furthermore, understanding the biological mechanisms underlying chemotherapy resistance could lead to the development of novel therapeutic strategies to improve outcomes in TNBC [6].Given the expanding landscape of potential predictive biomarkers and their critical importance in treatment decision-making, there is a pressing need to systematically evaluate and compare these markers to identify the most reliable predictors of NAC response in TNBC [15]. This systematic review aims to comprehensively assess the current evidence regarding predictive markers of NAC response in TNBC, with the goal of identifying the most promising biomarkers for clinical implementation.

### 5 Conclusion

This systematic review demonstrates that predicting neoadjuvant chemotherapy response in triple-negative breast cancer requires an integrated, multi-parameter approach. The evidence consistently shows that tumor-infiltrating lymphocytes serve as one of the most robust predictive markers, with lymphocyte-predominant breast cancers showing significantly higher pathological complete response rates. The molecular heterogeneity of TNBC, reflected in the differential response rates across TNBCtype-4 subtypes, underscores the necessity of incorporating molecular subtyping into response prediction frameworks.DNA repair deficiency emerges as another critical determinant of treatment response, with BRCA1 mutation carriers and HR-deficient tumors showing enhanced sensitivity to neoadjuvant chemotherapy. The observed synergy between DNA repair deficiency and immune activation suggests that combined assessment of these pathways may provide more accurate response prediction than either parameter alone.While individual clinical and pathological markers such as Ki-67 index and systemic inflammatory markers demonstrate predictive value, their utility appears maximized when integrated into comprehensive prediction models. Recent advances in artificial intelligence-based analysis of histological images represent a promising avenue for enhancing response prediction accuracy, particularly when combined with established biomarkers.The findings of this review have important implications for clinical practice and future research. The strong predictive value of immune markers supports their routine assessment in TNBC patients considering neoadjuvant chemotherapy. However, significant challenges remain in standardizing biomarker assessment methods and establishing clinically validated cutoff values. Future research should focus on developing practical, standardized approaches for integrated biomarker assessment that can guide treatment decisions in real-world clinical settings.Additionally, the observation that certain biomarker-positive populations may show higher pCR rates without corresponding improvements in survival highlights the need for careful consideration of both immediate treatment response and long-term outcomes in predictive modeling. Further investigation is needed to understand these disparities and their implications for treatment planning.In conclusion, optimal prediction of neoadjuvant chemotherapy response in TNBC requires integration of multiple biomarker classes, with particular emphasis on immune parameters, DNA repair proficiency, and molecular subtyping. The development of standardized, clinically practical methods for comprehensive biomarker assessment represents a critical next step in improving treatment outcomes for TNBC patients.

17. Bernemann C., Hülsewig C., Rückert C., Schäfer S., Blümel L., Hempel G., Götte

M., Greve B., Barth P.J., Kiesel L., et al. Influence of secreted frizzled receptor protein 1 (SFRP1) on neoadjuvant chemotherapy in triple negative breast cancer does not rely on WNT signaling. Mol. Cancer. 2014;13:174. doi: 10.1186/1476-4598-13-174.

40. Loibl S., Treue D., Budczies J., Weber K., Stenzinger A., Schmitt W.D., Weichert W., Jank P., Furlanetto J., Klauschen F., et al. Mutational Diversity and Therapy Response in Breast Cancer: A Sequencing Analysis in the Neoadjuvant GeparSepto Trial.

Clin. Cancer Res. 2019;25:3986–3995. doi: 10.1158/1078-0432.CCR-18-3258.

52. Onesti C.E., Josse C., Poncin A., Frères P., Poulet C., Bours V., Jerusalem G. Predictive and prognostic role of peripheral blood eosinophil count in triple-negative and

hormone receptor-negative/HER2-positive breast cancer patients undergoing neoadjuvant treatment. Oncotarget. 2018;9:33719–33733. doi: 10.18632/oncotarget.26120.

53. Lou C., Jin F., Zhao Q., Qi H. Correlation of serum, N.L.R.; PLR and HALP with efficacy of neoadjuvant chemotherapy and prognosis of triple-negative breast cancer. Am. J. Transl. Res. 2022;14:3240–3246.

54. Symmans W.F., Yau C., Chen Y.Y., Balassanian R., Klein M.E., Pusztai L., Nanda R., Parker B.A., Datnow B., Krings G., et al. Assessment of Residual Cancer Burden and Event-Free Survival in Neoadjuvant Treatment for High-risk Breast Cancer: An Analysis of Data From the I-SPY2 Randomized Clinical Trial. JAMA Oncol. 2021;7:1654–1663. doi: 10.1001/jamaoncol.2021.3690.
