## Supplementary material for "In Search of Ethical Procedures for LLM-Assisted Systematic Review Production: A Proof-of-Concept Evaluation of Selected Review Components": source_data: mcnemar.py

```
import pandas as pd
from scipy.stats import chi2, binomtest


def get_path():
    return input("Enter Excel file path: ").strip()


def load_table(filepath):
    df = pd.read_excel(filepath, header=0)

    # Set first column as row labels
    df = df.set_index(df.columns[0])

    # Extract values using labels (robust approach)
    a = df.loc["AI Include", "Human Include"]
    b = df.loc["AI Include", "Human Exclude"]
    c = df.loc["AI Exclude", "Human Include"]
    d = df.loc["AI Exclude", "Human Exclude"]

    return int(a), int(b), int(c), int(d)


def mcnemar_corrected(b, c):
    n = b + c
    if n == 0:
        raise ValueError("No discordant pairs.")

    chi2_stat = (abs(b - c) - 1) ** 2 / n
    p = chi2.sf(chi2_stat, df=1)

    return chi2_stat, p


def mcnemar_exact(b, c):
    n = b + c
    return binomtest(b, n=n, p=0.5, alternative="two-sided").pvalue


def main():
    path = get_path()

    a, b, c, d = load_table(path)

    print("\nContingency Table:")
    print(f"a = {a}, b = {b}, c = {c}, d = {d}")

    chi2_stat, p_chi = mcnemar_corrected(b, c)
    p_exact = mcnemar_exact(b, c)

    print("\nMcNemar (continuity corrected):")
    print(f"chi2 = {chi2_stat:.4f}, p = {p_chi:.6g}")

    print("\nMcNemar (exact binomial):")
    print(f"p = {p_exact:.6g}")


if __name__ == "__main__":
    main()
```
