## Supplementary material for "In Search of Ethical Procedures for LLM-Assisted Systematic Review Production: A Proof-of-Concept Evaluation of Selected Review Components": source_data: readme.txt

screening.py - Used to run the binomial test to assess for the ability to detect AI writing among survey respondents. Also computes a Cohen's Kappa to assess for beyond-chance agreement of raters against ground truth for survey respondents, and to assess for general agreement between AI and human screeners. Additionally runs a permutation test that shuffles the truth labels 10,00 times and coutns how often the shuffled accuracy meets or beats the observed accuracy. 

wilcoxon.py - Used to run paired Wilcoxon signed-rank test which was used to assess for significance within the survey respondents.

mcnemar.py - Used to run McNemar's test to assess for differences in leniency between the human screeners and the AI screener.
