## Supplementary material for "In Search of Ethical Procedures for LLM-Assisted Systematic Review Production: A Proof-of-Concept Evaluation of Selected Review Components": source_data: screening.py

```
import pandas as pd
import numpy as np
from scipy.stats import fisher_exact, binomtest
from sklearn.metrics import cohen_kappa_score, confusion_matrix
from itertools import product


LABELS = {0: "Full AI", 1: "Partial AI", 2: "No AI (Human)"}


def load_data(path):
    df = pd.read_excel(path, header=None)
    article_ids = df.iloc[:, 0].values
    ratings = df.iloc[:, 1:-1].astype(int)
    truth = df.iloc[:, -1].astype(int).values
    return article_ids, ratings, truth


def print_section(title):
    print(f"\n{'=' * 50}")
    print(f"  {title}")
    print('=' * 50)


def response_distribution(ratings, truth):
    print_section("DATA OVERVIEW")
    all_vals = ratings.values.flatten().astype(int)
    n_articles = ratings.shape[0]
    n_raters = ratings.shape[1]
    print(f"Articles: {n_articles}  |  Raters: {n_raters}  |  Total ratings: {len(all_vals)}")

    print("\nResponse distribution:")
    for val in sorted(LABELS.keys()):
        count = np.sum(all_vals == val)
        print(f"  {val} ({LABELS[val]}): {count}")

    print("\nTruth distribution:")
    for val in sorted(LABELS.keys()):
        count = np.sum(truth == val)
        print(f"  {val} ({LABELS[val]}): {count}")


def individual_accuracy_test(ratings, truth):
    """Treat each rating as independent observation. Test if accuracy > chance (1/3)."""
    print_section("INDIVIDUAL RATING ACCURACY")

    all_responses = ratings.values.flatten().astype(int)
    expanded_truth = np.repeat(truth, ratings.shape[1])
    n_correct = np.sum(all_responses == expanded_truth)
    n_total = len(all_responses)
    accuracy = n_correct / n_total
    chance = 1.0 / 3.0

    # Exact binomial test: are raters correct more than 1/3 of the time?
    p_val = binomtest(n_correct, n_total, chance, alternative='greater').pvalue

    print(f"Correct: {n_correct}/{n_total} ({accuracy:.1%})")
    print(f"Chance level: {chance:.1%}")
    print(f"Binomial test (one-sided, H1: accuracy > chance): p = {p_val:.4f}")

    if p_val < 0.05:
        print("-> Raters perform significantly above chance.")
    else:
        print("-> No significant evidence of above-chance performance.")


def per_rater_breakdown(ratings, truth):
    """Show accuracy and Cohen's kappa for each rater."""
    print_section("PER-RATER BREAKDOWN")

    print(f"{'Rater':<8} {'Correct':<10} {'Accuracy':<10} {'Kappa':<10}")
    print("-" * 38)

    kappas = []
    for i, col in enumerate(ratings.columns):
        rater = ratings[col].values.astype(int)
        correct = np.sum(rater == truth)
        acc = correct / len(truth)
        try:
            k = cohen_kappa_score(truth, rater)
        except Exception:
            k = float('nan')
        kappas.append(k)
        print(f"R{i+1:<7} {correct}/{len(truth):<8} {acc:<10.1%} {k:<10.3f}")

    mean_k = np.nanmean(kappas)
    print(f"\nMean kappa: {mean_k:.3f}")
    if mean_k < 0:
        print("-> Worse than chance agreement.")
    elif mean_k < 0.20:
        print("-> Slight agreement.")
    elif mean_k < 0.40:
        print("-> Fair agreement.")
    elif mean_k < 0.60:
        print("-> Moderate agreement.")
    else:
        print("-> Substantial agreement.")


def confusion_analysis(ratings, truth):
    """Build aggregate confusion matrix across all raters."""
    print_section("CONFUSION MATRIX (all ratings pooled)")

    all_responses = ratings.values.flatten().astype(int)
    expanded_truth = np.repeat(truth, ratings.shape[1])

    labels = sorted(LABELS.keys())
    cm = confusion_matrix(expanded_truth, all_responses, labels=labels)

    # Print header
    col_w = 14
    row_label_w = 22
    print(f"\n{'':>{row_label_w}}  RATED AS")
    print(f"{'':>{row_label_w}}", end="")
    for l in labels:
        print(f"{LABELS[l]:>{col_w}}", end="")
    print()
    print(" " * row_label_w + "-" * (col_w * len(labels)))

    for i, l in enumerate(labels):
        print(f"  TRUE {LABELS[l]:>{row_label_w - 9}} |", end="")
        for j in range(len(labels)):
            print(f"{cm[i][j]:>{col_w}}", end="")
        print()

    print("\nRead: each row is the true category; columns show what raters guessed.")


def bias_analysis(ratings, truth):
    """Directional bias — do raters lean toward AI or human?"""
    print_section("BIAS ANALYSIS")

    all_responses = ratings.values.flatten().astype(int)
    expanded_truth = np.repeat(truth, ratings.shape[1])
    diff = all_responses - expanded_truth

    mean_bias = np.mean(diff)
    print(f"Mean bias (response - truth): {mean_bias:+.3f}")

    if mean_bias > 0.1:
        print("-> Raters tend to rate HIGHER than truth (toward 'No AI').")
        print("   i.e., they UNDERestimate AI involvement.")
    elif mean_bias < -0.1:
        print("-> Raters tend to rate LOWER than truth (toward 'Full AI').")
        print("   i.e., they OVERestimate AI involvement.")
    else:
        print("-> No strong directional bias.")


def per_article_summary(article_ids, ratings, truth):
    """Show how each article was rated vs its truth."""
    print_section("PER-ARTICLE DETAIL")

    for i, art in enumerate(article_ids):
        row = ratings.iloc[i].values.astype(int)
        t = truth[i]
        correct = np.sum(row == t)
        print(f"\n  {art}  (truth: {t} = {LABELS[t]})")
        print(f"    Ratings: {[int(x) for x in row]}")
        print(f"    Correct: {correct}/{len(row)}")


def permutation_test(ratings, truth, n_iter=10000):
    """Permutation test — shuffles truth labels, computes overall accuracy."""
    print_section("PERMUTATION TEST")

    all_responses = ratings.values.flatten().astype(int)
    expanded_truth = np.repeat(truth, ratings.shape[1])
    observed = np.mean(all_responses == expanded_truth)

    count_ge = 0
    for _ in range(n_iter):
        shuffled = np.random.permutation(truth)
        exp_shuf = np.repeat(shuffled, ratings.shape[1])
        acc = np.mean(all_responses == exp_shuf)
        if acc >= observed:
            count_ge += 1

    p_val = count_ge / n_iter
    print(f"Observed accuracy: {observed:.3f}")
    print(f"Permutation p-value ({n_iter} iterations): {p_val:.4f}")

    if p_val < 0.05:
        print("-> Significant: raters are classifying above chance.")
    else:
        print("-> Not significant at p < 0.05.")


def main():
    path = input("Enter Excel file path: ").strip()
    articles, ratings, truth = load_data(path)

    response_distribution(ratings, truth)
    per_article_summary(articles, ratings, truth)
    confusion_analysis(ratings, truth)
    individual_accuracy_test(ratings, truth)
    per_rater_breakdown(ratings, truth)
    bias_analysis(ratings, truth)
    permutation_test(ratings, truth)

    print_section("NOTES")
    print("- Binomial test treats each rating independently (may slightly")
    print("  overstate significance due to clustering within articles).")
    print("- Cohen's kappa adjusts for chance agreement in ordinal data.")
    print("- With 3 articles, statistical power is inherently very limited.")
    print("- Consider this exploratory / descriptive rather than confirmatory.")


if __name__ == "__main__":
    main()
```
