## Supplementary material for "In Search of Ethical Procedures for LLM-Assisted Systematic Review Production: A Proof-of-Concept Evaluation of Selected Review Components": source_data: wilcoxon.py

```
import pandas as pd
from scipy.stats import wilcoxon


def get_file_path():
    return input("Enter full path to Excel file: ").strip()


def load_data(filepath):
    # Assumes:
    # Column A = AI ratings
    # Column B = Human ratings
    df = pd.read_excel(filepath)

    ai = df.iloc[:, 0].dropna()
    human = df.iloc[:, 1].dropna()

    # Ensure paired structure (same length)
    if len(ai) != len(human):
        raise ValueError(
            f"AI and Human columns have different lengths: "
            f"{len(ai)} vs {len(human)}. Check missing values."
        )

    return ai.values, human.values


def run_wilcoxon(ai, human):
    # Paired Wilcoxon signed-rank test
    # alternative='two-sided' is default but made explicit for clarity
    stat, p = wilcoxon(ai, human, alternative="two-sided", zero_method="wilcox")

    return stat, p


def main():
    filepath = get_file_path()

    ai, human = load_data(filepath)

    stat, p = run_wilcoxon(ai, human)

    print("\nPaired Wilcoxon signed-rank test results:")
    print(f"Test statistic (W): {stat:.4f}")
    print(f"p-value: {p:.6g}")

    print("\nInterpretation:")
    if p < 0.05:
        print("→ Significant difference between AI and Human ratings.")
    else:
        print("→ No statistically significant difference detected.")


if __name__ == "__main__":
    main()
```
