## Supplementary material for "In Search of Ethical Procedures for LLM-Assisted Systematic Review Production: A Proof-of-Concept Evaluation of Selected Review Components": source_data: survey_copy.pdf

UNIVERSITY OF MINNESOTA
Driven to Discover®

#### Reviewer Information

### The Need for Standardization in Next-Generation Sequencing Studies for Classic Hodgkin Lymphoma: A Systematic Review

### Online Evaluation Form

#### Reviewer Information

The topic covered in these reviews is The Need for Standardization in Next-Generation Sequencing Studies for Classic Hodgkin Lymphoma: A Systematic Review

Given the topic, what is your level of expertise in the subject matter of these reviews?

- ○ I have no familiarity with this subject.
- ○ I am familiar with this topic.

- ○ I am an expert on this topic.

Please include any relevant credentials for your level of expertise. (MD, DO, PhD, Specialty/Sub-Specialty)

Have you reviewed the consent form?
(Click here for consent form)

- ○ I have reviewed the attached consent form and consent to participating in this study [this is your electronic signature]
- ○ No, I have not reviewed the consent form and/or I do not wish to participate in this study

#### Survey

Please open the three discussion sections to the systematic reviews linked here.
These three discussion sections cover very similar topics, after reviewing the text, please answer the below questions:

- Maroon and Gold
- North Star
- Paul Bunyan

Provide an overall quality of this review taking into consideration the overall content, quality of writing, and scientific merit. Scale: 1 (poor) to 5 (Excellent)

| | 1 | 2 | 3 | 4 | 5 | |
|---|---|---|---|---|---|---|
| Maroon and Gold Paper | ○ | | | | | ☐ |
| North Star Paper | ○ | | | | | ☐ |
| Paul Bunyan Paper | ○ | | | | | ☐ |

In your opinion are the results and discussion sections appropriate for publication as part of a systematic review

| | Maroon and Gold Paper | North Star Paper | Paul Bunyan Paper |
|---|---|---|---|
| Yes | ☐ | ☐ | ☐ |

| | Maroon and Gold Paper | North Star Paper | Paul Bunyan Paper |
|---|---|---|---|
| No | ☐ | ☐ | ☐ |
| Other ☐ | ☐ | ☐ | ☐ |

To what degree do you think this paper was written with the help of Artificial Intelligence / Large Language Model (AI/LLM)

| | Maroon and Gold Paper | North Star Paper | Paul Bunyan Paper |
|---|---|---|---|
| This paper was entirely human generated | ☐ | ☐ | ☐ |
| This paper was written with the assistance of AI/LLM along with human guidance | ☐ | ☐ | ☐ |

| | Maroon and Gold Paper | North Star Paper | Paul Bunyan Paper |
|---|---|---|---|
| This paper was entirely generated by AI/LLM with minimal human intervention | ☐ | ☐ | ☐ |

What is your experience using Chatbots/Artificial Intelligence/Large Language Models
Scale:
0 - Never Used
1 - rarely use (less than once per month)
2 - sometimes use (monthly to weekly)
3 - use frequently (more than once per week)

| | 0 | 1 | 2 | 3 | |
|---|---|---|---|---|---|
| Frequency in use of AI | ○ | | | | ☐ |

Additional comments?
After submitting this part of the survey you will not be able to

go back and change your answers.

#### Debrief

Have you read a paper with this title:
The Need for Standardization in Next-Generation Sequencing Studies for Classic Hodgkin Lymphoma: A Systematic Review (The Maroon and Gold Paper)
https://pmc.ncbi.nlm.nih.gov/articles/PMC9027849/

- Yes
- No
- Other

Optional for research purposes only:

Age

Gender (M, F, non-binary)

Debrief below:

**Debriefing Script: Evaluating the Quality of Systematic Reviews**

Dear Study Participant,

Thank you for your valuable contribution to our research study. We would like to take this opportunity to provide you with complete information about the nature of the study you just participated in.

**Study Purpose and Design**

The primary aim of this study was to evaluate the quality of systematic reviews created using different methodological approaches. The three systematic reviews you evaluated were created using distinct methods and tools, which we intentionally did not disclose to prevent any potential bias in your assessment. This element of incomplete disclosure was essential to obtain unbiased evaluations of the quality, readability, and scientific merit of each review.

**Your Rights as a Participant**

Now that you are aware of the incomplete disclosure in the study design, you have the following rights:

1. You may withdraw your participation and request that your evaluations be removed from the study data.
2. You may request additional information about the specific methodologies used in creating each review.
3. You may provide additional feedback now that you understand the context of the study.

**Importance of This Research**
Your participation helps us understand the effectiveness of different approaches to systematic review creation. This knowledge is crucial as research methodologies continue to evolve. Your expert evaluation will help establish guidelines for best practices in systematic review creation and inform the medical research community.
**Questions or Concerns**
If you have any questions about the study design, your participation, or would like to exercise any of the rights mentioned above, please contact:

**Confidentiality**
We want to reassure you that your identity will remain confidential in any publications or presentations resulting from this study. Only your qualifications and expertise level will be mentioned, without any identifying information.

Powered by Qualtrics
